# Genome-wide characterization of 54 urinary metabolites reveals molecular impact of kidney function

**DOI:** 10.1101/2023.12.20.23300206

**Authors:** Erkka Valo, Anne Richmond, Stefan Mutter, Archie Campbell, David Porteous, James F Wilson, FinnDiane Study Group, Per-Henrik Groop, Caroline Hayward, Niina Sandholm

## Abstract

Dissecting the genetic mechanisms underlying urinary metabolite concentrations can provide molecular insights into kidney function and open possibilities for causal assessment of urinary metabolites with risk factors and disease outcomes. Proton nuclear magnetic resonance metabolomics provides a high-throughput means for urinary metabolite profiling, as widely applied for blood biomarker studies. Here we report a genome-wide association study meta-analysed for 3 European cohorts comprising 8,026 individuals, covering both people with type 1 diabetes and general population settings. We identified 52 associations (*p*<9.3×10^-10^) for 19 of 54 studied metabolite concentrations. Out of these, 32 were not reported previously for relevant urinary or blood metabolite traits. Subsequent two-sample Mendelian randomization analysis suggests that estimated glomerular filtration rate (eGFR) causally affects 13 urinary metabolite concentrations whereas urinary ethanolamine, an initial precursor for phosphatidylcholine and phosphatidylethanolamine, was associated with higher eGFR lending support for a potential protective role.

## INTRODUCTION

Urinary metabolite concentrations are read-outs of biological processes and can inform on the molecular basis of diseases. Automation of metabolomics technologies for urine analyses, such as nuclear magnetic resonance (NMR), has lacked behind blood profiling but now allows for accurate quantification at an entire cohort scale. This may pave way for wide-spread epidemiological and translational applications analogous to plasma NMR profiling (e.g. in the UK Biobank^1^). Such recent studies have, for example, highlighted 10 urinary metabolites being predictive of diabetic kidney disease (DKD) progression in individuals with type 1 diabetes (T1D)^2^ and multiple associations between 49 clinical measures and 12 urinary metabolites in a general population setting^3^.

Studying the genetic regulation of urinary metabolites can reveal novel biological pathways behind the identified biomarkers. Specific to the urinary biomarkers, is that they can either reflect the systemic (blood) biomarker levels but provide a less tightly regulated and more accessible source of biomarker material compared with blood; or they can reflect changes in the kidney function, related either to changes in the glomerular filtration rate, increased leakage of molecules into the urine, changes in tubular reabsorption into the blood, or originating from the kidney or the urinary system tissue.

Previous research on the genetics of urinary metabolites has identified several hundred loci associated with urinary metabolites^4–7^. Schlosser et al. (2023) identified 622 genomic intervals associated with urinary metabolite concentrations across 1,399 metabolites measured in 4,912 individuals^7^. Moreover, a study in the UK Biobank identified multiple loci associated with four clinical urinary laboratory measurements in 363,228 individuals^8^. Here, balancing between large sample size and extensive molecular coverage, we study 54 urinary metabolites in 8,026 individuals to further characterise genetics of urinary metabolites.

If a given urinary metabolite is found to be associated with a specific disease, the information on the genetic variants associated with urinary metabolites can be applied to infer potential causal relationships between urinary metabolites and the disease in question using a Mendelian randomization (MR) approach. Causal analysis benefits from multiple robust genetic instruments: although our study includes fewer metabolites than Schlosser et al. (2023), the larger sample size gives us power to potentially identify more associations with urinary metabolites.

This study investigated single nucleotide variants (SNVs) associated with 54 urinary metabolites measured by NMR in one Finnish cohort of individuals with type 1 diabetes (T1D) and two Scottish cohorts from a general population setting including in total 8,026 individuals. Furthermore, we characterized the identified associations and their molecular basis by analysing the variants’ effect on gene expression harnessing relevant expression quantitative trait loci (eQTL) data. Moreover, causal relationships were identified between the metabolites and relevant phenotypes using MR analysis.

## RESULTS

### Genome-wide association study identified 52 associations with urinary metabolites

We performed genome-wide association studies (GWAS) of 54 urinary metabolites in 3 cohorts followed by a meta-analysis (Methods). The analysis included in total 8,026 individuals from the Finnish Diabetic Nephropathy Study (FinnDiane, n= 3,244)^9,10^, Generation Scotland (GS, n= 2,743)^11^, and the VIKING study (VIKING, n= 2,027)^12^ (Figure 1, Supplementary Table 1). We measured 54 urinary metabolites with the Nightingale Health urine NMR platform (Supplementary Table 2 and 3). The metabolites were quantified in absolute concentrations and normalized with urinary creatinine concentration prior to GWAS analysis. Here we report associations for variants found in at least 2 out of 3 cohorts and with a minor allele frequency (MAF) ≥ 1%.

**Figure 1.**
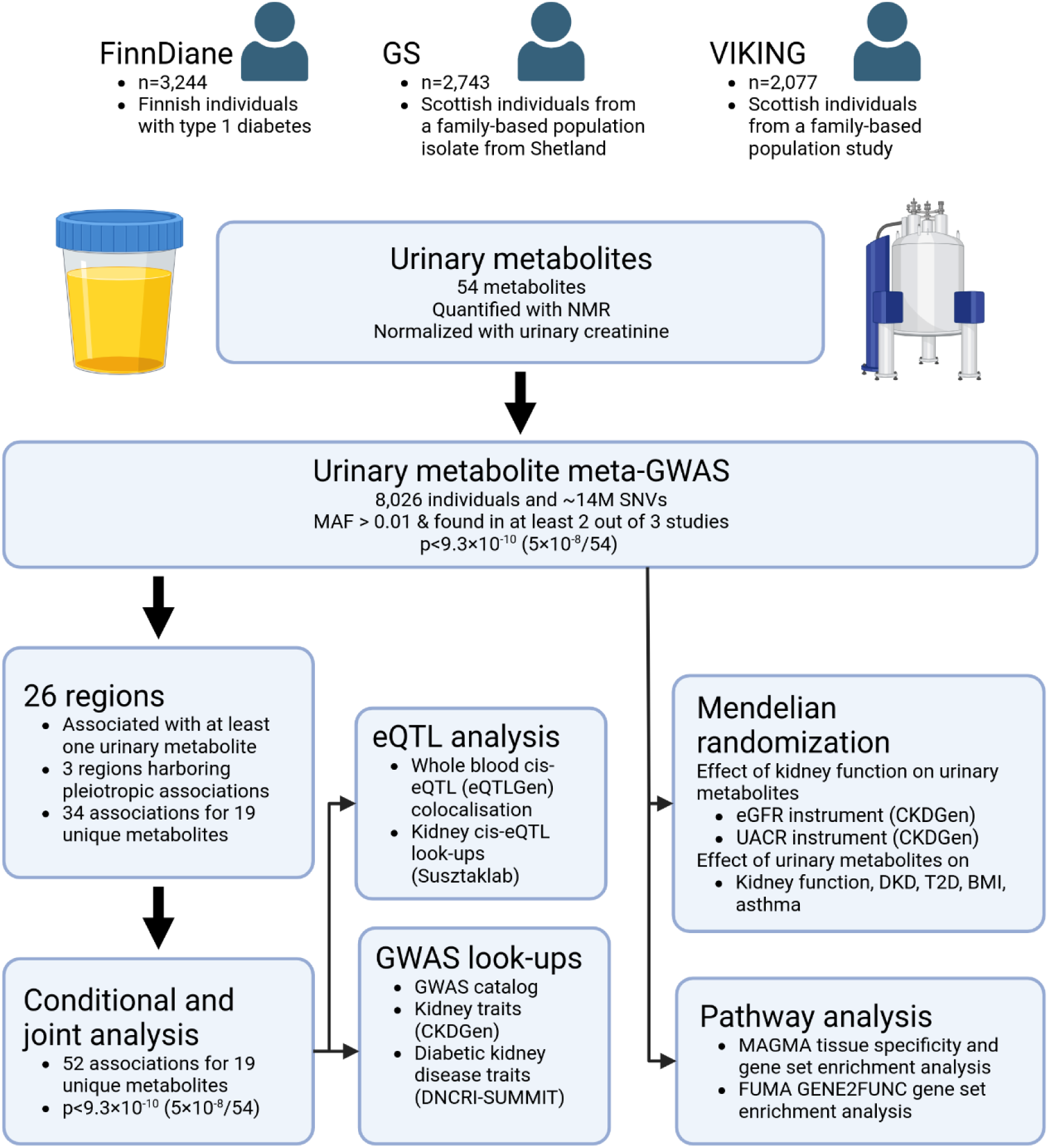
Study design overview. An overview of the genome-wide characterization of the urinary metabolites. Created with BioRender.com.

We identified 26 chromosomal regions harbouring associations with at least one metabolite amongst the 54 metabolites meta-analysed across the three cohorts (p-value < 9.3×10^-10^; Figure 2, Methods). In total, the regions contained 34 associations with 19 unique urinary metabolites and three of the 26 regions showed evidence of pleiotropy: the loci on chromosomes 5p15.33 and 17q12 associated with 5 and 4 amino acids respectively, and a locus on chromosome 7p21.1 associated with quinic acid and trigonelline.

**Figure 2.**
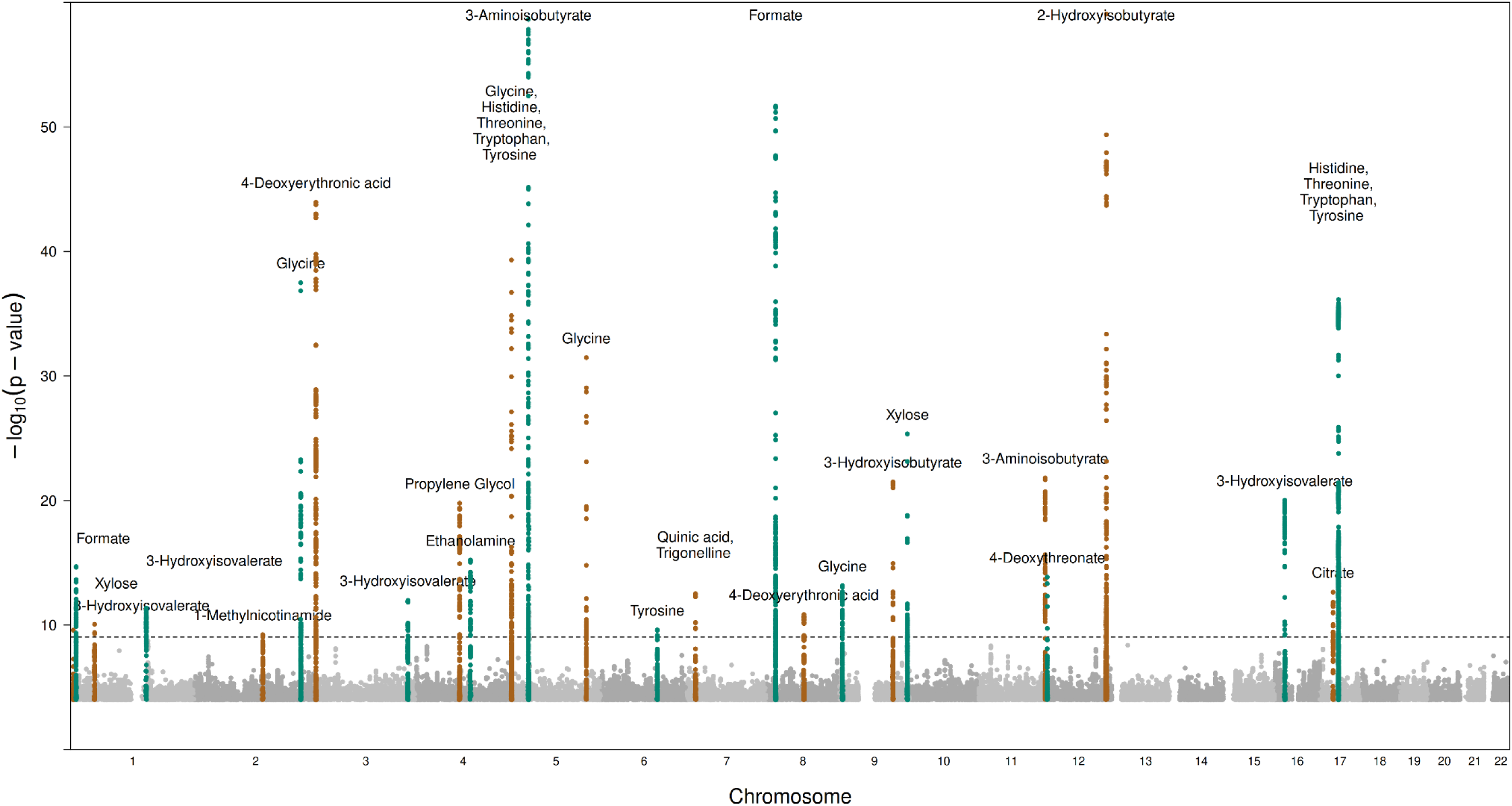
Manhattan plot of signals with p < 5.0×10^-5^ for the metabolites. Signals from different metabolites are clumped together if they are within 50kb from another signal. Pruned genome-wide significant signals with p < 5×10^-8^/54=9.3×10^-10^ and variants 1Mb around them are highlighted. Note: y-axis clipped at 60.

The urinary metabolite heritability meta-analysis estimates ranged from 0% to 36%, being the highest for urinary citrate (36%), 3-aminoisobutyrate (33%), and tyrosine (29%) concentrations (Supplementary Table 4, Supplementary Figure 1, Methods). Altogether 27 metabolites showed evidence of heritability ranging from 6% to 36% (*p* < 0.05). Only 2 out of the 27 metabolites, glycine and 2-hydroxyisobutyrate, showed evidence of between study heterogeneity in heritability estimates (*p* < 0.05).

We performed conditional and joint multiple-SNV analysis^13^ to pinpoint independent signals within these loci and found 52 study-wide significant associations (p < 9.3×10^-10^) (Supplementary Figure 2 and Supplementary Table 5, Methods). In total, 6 metabolites had multiple signals in the same locus, notably, a region on chromosome 5p13.2 had 13 associations with 3-aminoisobutyrate. Of the 52 associations, 31 signals were novel for urinary or blood metabolite traits (Table 1), whereas 21 associations were previously reported for relevant urinary and/or blood metabolite traits (Supplementary Table 6). The novel signals included 9 associations with 3-aminoisobutyrate, 4 associations with glycine, 2 associations with 3-hydroxyisovalerate, 4-deoxyeryhronic acid, threonine, and xylose, and finally, single associations with 3-hydroxyisobutyrate, 4-deoxythreonate, citrate, ethanolamine, formate, propylene glycol, quinic acid, trigonelline, tryptophan, and tyrosine (Table 1).

**Table 1.**
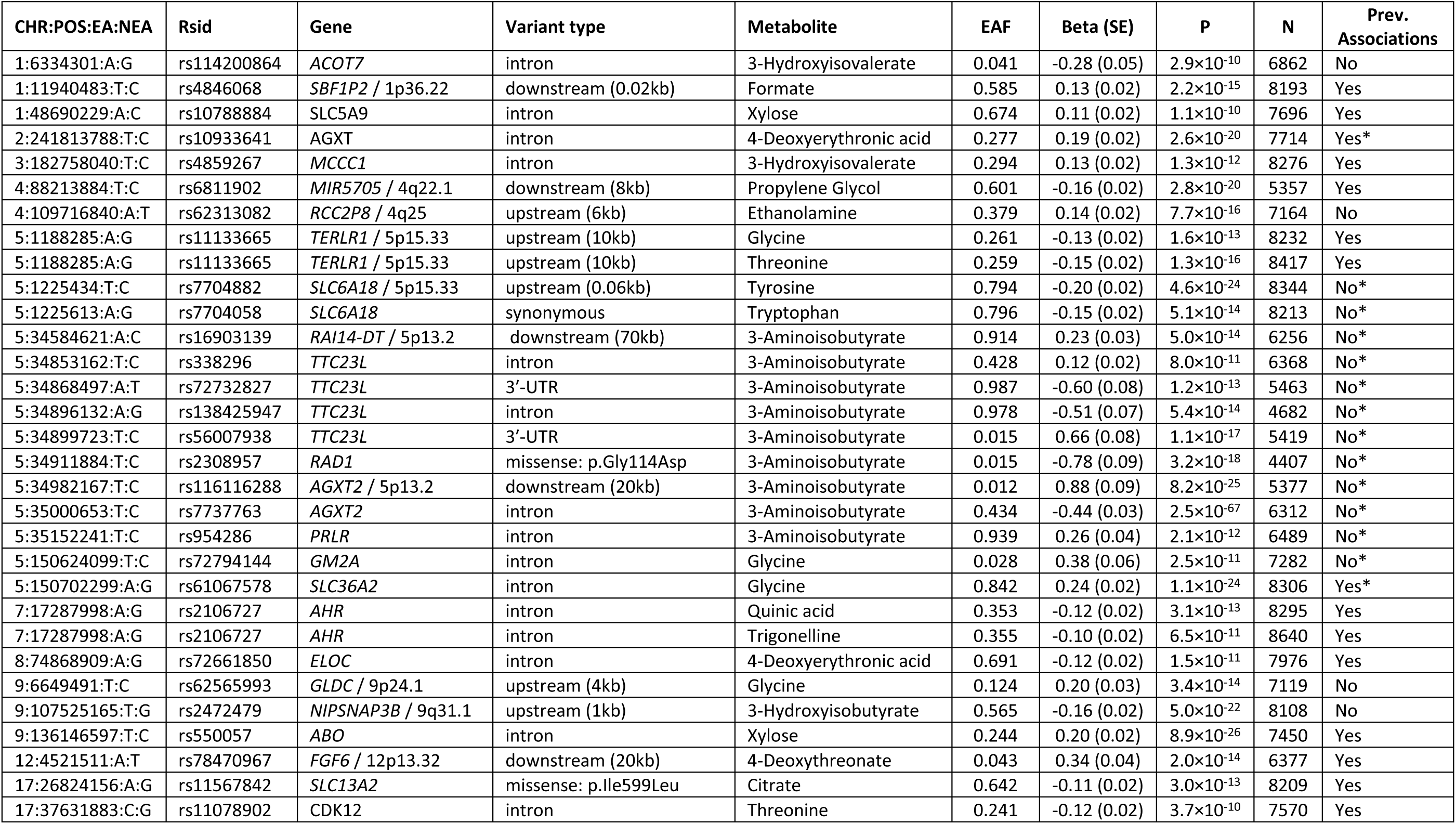
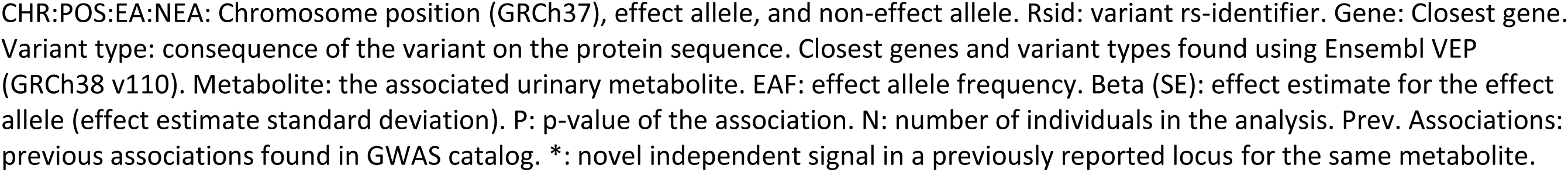
Variants associated with metabolites (*p* < 9.3×10^-10^) with no previously reported associations in the GWAS catalogue with the same metabolite in blood or urine (window size=-/+500kb, r^2^ > 0.8, and *p* < 5×10^-8^).

| CHR:POS:EA:NEA | Rsid | Gene | Variant type | Metabolite | EAF | Beta (SE) | P | N | Prev. Associations |
| --- | --- | --- | --- | --- | --- | --- | --- | --- | --- |
| 1:6334301:A:G | rs114200864 | ACOT7 | intron | 3-Hydroxyisovalerate | 0.041 | -0.28 (0.05) | $2.9 \times 10^{-10}$ | 6862 | No |
| 1:11940483:T:C | rs4846068 | SBF1P2 / 1p36.22 | downstream (0.02kb) | Formate | 0.585 | 0.13 (0.02) | $2.2 \times 10^{-15}$ | 8193 | Yes |
| 1:48690229:A:C | rs10788884 | SLC5A9 | intron | Xylose | 0.674 | 0.11 (0.02) | $1.1 \times 10^{-10}$ | 7696 | Yes |
| 2:241813788:T:C | rs10933641 | AGXT | intron | 4-Deoxyerythronic acid | 0.277 | 0.19 (0.02) | $2.6 \times 10^{-20}$ | 7714 | Yes* |
| 3:182758040:T:C | rs4859267 | MCCC1 | intron | 3-Hydroxyisovalerate | 0.294 | 0.13 (0.02) | $1.3 \times 10^{-12}$ | 8276 | Yes |
| 4:88213884:T:C | rs6811902 | MIR5705 / 4q22.1 | downstream (8kb) | Propylene Glycol | 0.601 | -0.16 (0.02) | $2.8 \times 10^{-20}$ | 5357 | Yes |
| 4:109716840:A:T | rs62313082 | RCC2P8 / 4q25 | upstream (6kb) | Ethanolamine | 0.379 | 0.14 (0.02) | $7.7 \times 10^{-16}$ | 7164 | No |
| 5:1188285:A:G | rs11133665 | TERLR1 / 5p15.33 | upstream (10kb) | Glycine | 0.261 | -0.13 (0.02) | $1.6 \times 10^{-13}$ | 8232 | Yes |
| 5:1188285:A:G | rs11133665 | TERLR1 / 5p15.33 | upstream (10kb) | Threonine | 0.259 | -0.15 (0.02) | $1.3 \times 10^{-16}$ | 8417 | Yes |
| 5:1225434:T:C | rs7704882 | SLC6A18 / 5p15.33 | upstream (0.06kb) | Tyrosine | 0.794 | -0.20 (0.02) | $4.6 \times 10^{-24}$ | 8344 | No* |
| 5:1225613:A:G | rs7704058 | SLC6A18 | synonymous | Tryptophan | 0.796 | -0.15 (0.02) | $5.1 \times 10^{-14}$ | 8213 | No* |
| 5:34584621:A:C | rs16903139 | RAI14-DT / 5p13.2 | downstream (70kb) | 3-Aminoisobutyrate | 0.914 | 0.23 (0.03) | $5.0 \times 10^{-14}$ | 6256 | No* |
| 5:34853162:T:C | rs338296 | TTC23L | intron | 3-Aminoisobutyrate | 0.428 | 0.12 (0.02) | $8.0 \times 10^{-11}$ | 6368 | No* |
| 5:34868497:A:T | rs72732827 | TTC23L | 3'-UTR | 3-Aminoisobutyrate | 0.987 | -0.60 (0.08) | $1.2 \times 10^{-13}$ | 5463 | No* |
| 5:34896132:A:G | rs138425947 | TTC23L | intron | 3-Aminoisobutyrate | 0.978 | -0.51 (0.07) | $5.4 \times 10^{-14}$ | 4682 | No* |
| 5:34899723:T:C | rs56007938 | TTC23L | 3'-UTR | 3-Aminoisobutyrate | 0.015 | 0.66 (0.08) | $1.1 \times 10^{-17}$ | 5419 | No* |
| 5:34911884:T:C | rs2308957 | RAD1 | missense: p.Gly114Asp | 3-Aminoisobutyrate | 0.015 | -0.78 (0.09) | $3.2 \times 10^{-18}$ | 4407 | No* |
| 5:34982167:T:C | rs116116288 | AGXT2 / 5p13.2 | downstream (20kb) | 3-Aminoisobutyrate | 0.012 | 0.88 (0.09) | $8.2 \times 10^{-25}$ | 5377 | No* |
| 5:35000653:T:C | rs7737763 | AGXT2 | intron | 3-Aminoisobutyrate | 0.434 | -0.44 (0.03) | $2.5 \times 10^{-67}$ | 6312 | No* |
| 5:35152241:T:C | rs954286 | PRLR | intron | 3-Aminoisobutyrate | 0.939 | 0.26 (0.04) | $2.1 \times 10^{-12}$ | 6489 | No* |
| 5:150624099:T:C | rs72794144 | GM2A | intron | Glycine | 0.028 | 0.38 (0.06) | $2.5 \times 10^{-11}$ | 7282 | No* |
| 5:150702299:A:G | rs61067578 | SLC36A2 | intron | Glycine | 0.842 | 0.24 (0.02) | $1.1 \times 10^{-24}$ | 8306 | Yes* |
| 7:17287998:A:G | rs2106727 | AHR | intron | Quinic acid | 0.353 | -0.12 (0.02) | $3.1 \times 10^{-13}$ | 8295 | Yes |
| 7:17287998:A:G | rs2106727 | AHR | intron | Trigonelline | 0.355 | -0.10 (0.02) | $6.5 \times 10^{-11}$ | 8640 | Yes |
| 8:74868909:A:G | rs72661850 | ELOC | intron | 4-Deoxyerythronic acid | 0.691 | -0.12 (0.02) | $1.5 \times 10^{-11}$ | 7976 | Yes |
| 9:6649491:T:C | rs62565993 | GLDC / 9p24.1 | upstream (4kb) | Glycine | 0.124 | 0.20 (0.03) | $3.4 \times 10^{-14}$ | 7119 | No |
| 9:107525165:T:G | rs2472479 | NIPSNAP3B / 9q31.1 | upstream (1kb) | 3-Hydroxyisobutyrate | 0.565 | -0.16 (0.02) | $5.0 \times 10^{-22}$ | 8108 | No |
| 9:136146597:T:C | rs550057 | ABO | intron | Xylose | 0.244 | 0.20 (0.02) | $8.9 \times 10^{-26}$ | 7450 | Yes |
| 12:4521511:A:T | rs78470967 | FGF6 / 12p13.32 | downstream (20kb) | 4-Deoxythreonate | 0.043 | 0.34 (0.04) | $2.0 \times 10^{-14}$ | 6377 | Yes |
| 17:26824156:A:G | rs11567842 | SLC13A2 | missense: p.Ile599Leu | Citrate | 0.642 | -0.11 (0.02) | $3.0 \times 10^{-13}$ | 8209 | Yes |
| 17:37631883:C:G | rs11078902 | CDK12 | intron | Threonine | 0.241 | -0.12 (0.02) | $3.7 \times 10^{-10}$ | 7570 | Yes |

CHR:POS:EA:NEA: Chromosome position (GRCh37), effect allele, and non-effect allele. Rsid: variant rs-identifier. Gene: Closest gene. Variant type: consequence of the variant on the protein sequence. Closest genes and variant types found using Ensembl VEP (GRCh38 v110). Metabolite: the associated urinary metabolite. EAF: effect allele frequency. Beta (SE): effect estimate for the effect allele (effect estimate standard deviation). P: p-value of the association. N: number of individuals in the analysis. Prev. Associations: previous associations found in GWAS catalog. \*: novel independent signal in a previously reported locus for the same metabolite.

Four of the 52 lead variants were missense variants, two representing previously unknown metabolite associations. rs11567842 (*SLC13A2* p.Ile599Leu) was associated with urinary citrate concentration and has previously been associated with blood urea nitrogen concentration^14^. *SLC13A2* encodes Solute Carrier Family 13 Member 2, which is a kidney sodium-coupled citrate transporter^15^. We have previously shown that urinary citrate concentration is associated with progression of DKD^2^. The citrate-associated rs11567842 was nominally associated with multiple DKD phenotypes (*p*=0.03-0.001)^16^ and with eGFR in the general population (p=0.012)^17^, although these genetic associations did not remain after correction for multiple testing. At a locus on chromosome 5p13.2, including 13 independent signals associated with 3-aminoisobutyrate, two of the lead variants were missense variants but in two different genes: rs37369 (*AGXT2* p.Val140Ile) and rs2308957 *RAD1* p.Gly114Asp. The rs37369 (*AGXT2* p.Val140Ile) variant has previously been associated with urinary and plasma 3-aminoisobutyrate levels^18,19^ whereas rs2308957 (*RAD1* p.Gly114Asp) is novel. Other lead variants in the region were eQTLs for *AGXT2* or both *AGXT2* and *RAD1* in the kidney. *AGXT2* encodes alanine–glyoxylate aminotransferase 2, expressed in kidney and liver in the human protein atlas, and is the biologically more plausible gene underlying the association signal. In single-cell RNAseq of human kidneys, the gene is expressed specifically in the proximal convoluted tubules (Supplementary Figure 3)^20^. *RAD1* encodes a ubiquitously expressed component of the 9-1-1 cell-cycle checkpoint response complex that plays a major role in DNA repair. Finally, rs1047891 (*CPS1* p.Thr1406Asn) has previously been associated with 266 traits including the currently observed glycine association in plasma^21^, as well as with eGFR^22^.

Of note, all the identified missense variants were predicted to be tolerated or benign by SIFT and PolyPhen.2 algorithms, but they may be sufficient to cause subtle changes in the protein function seen as altered urinary metabolite concentrations.

### eQTL data in kidney and whole blood highlights membrane transport proteins

As most of the identified lead variants were non-coding, they most likely represent regulatory variants affecting gene expression. We utilized expression quantitative trait loci (eQTLs) data from kidney (tubular and glomerular tissues) and whole blood to assess whether the identified variants influence the expression of nearby genes (Methods).

Among the identified variants, 26 were *cis-*eQTLs, i.e., associated with gene expression of a nearby gene, in either kidney tubules, glomeruli, and/or whole kidney (*p*<5.3×10^-4^). The associated genes included five solute carrier genes that transport solutes across cell membranes: *SLC5A9* (Sodium/Glucose Cotransporter 4 [SGLT4]), *SLC6A19* (Sodium-Dependent Neutral Amino Acid Transporter B(0)AT1), *SLC6A18* (Sodium-And Chloride-Dependent Transporter XTRP2), *SLC16A10*, and *SLC6A13* (Table 2). An intronic variant in the *SLC5A9* gene, rs10788884, was associated with urinary xylose concentrations and represents a new metabolite association. Rs10788884 is a strong eQTL for *SLC5A9* in the kidneys (*p*=3.4×10^-52^) as well as separately for the kidney tubules (*p*=3.4×10^-33^) and the glomeruli (*p*=4.3×10^-26^). The variant has previously been associated with serum uric acid^14^ and urate^23^, and urinary mannose^7^. *SLC5A9* encodes a sodium-dependent glucose transporter (SGLT4) that is expressed in the intestine and the kidneys and is an essential transporter for mannose, 1,5-anhydro-D-glucitol, and fructose^24^.

**Table 2.**
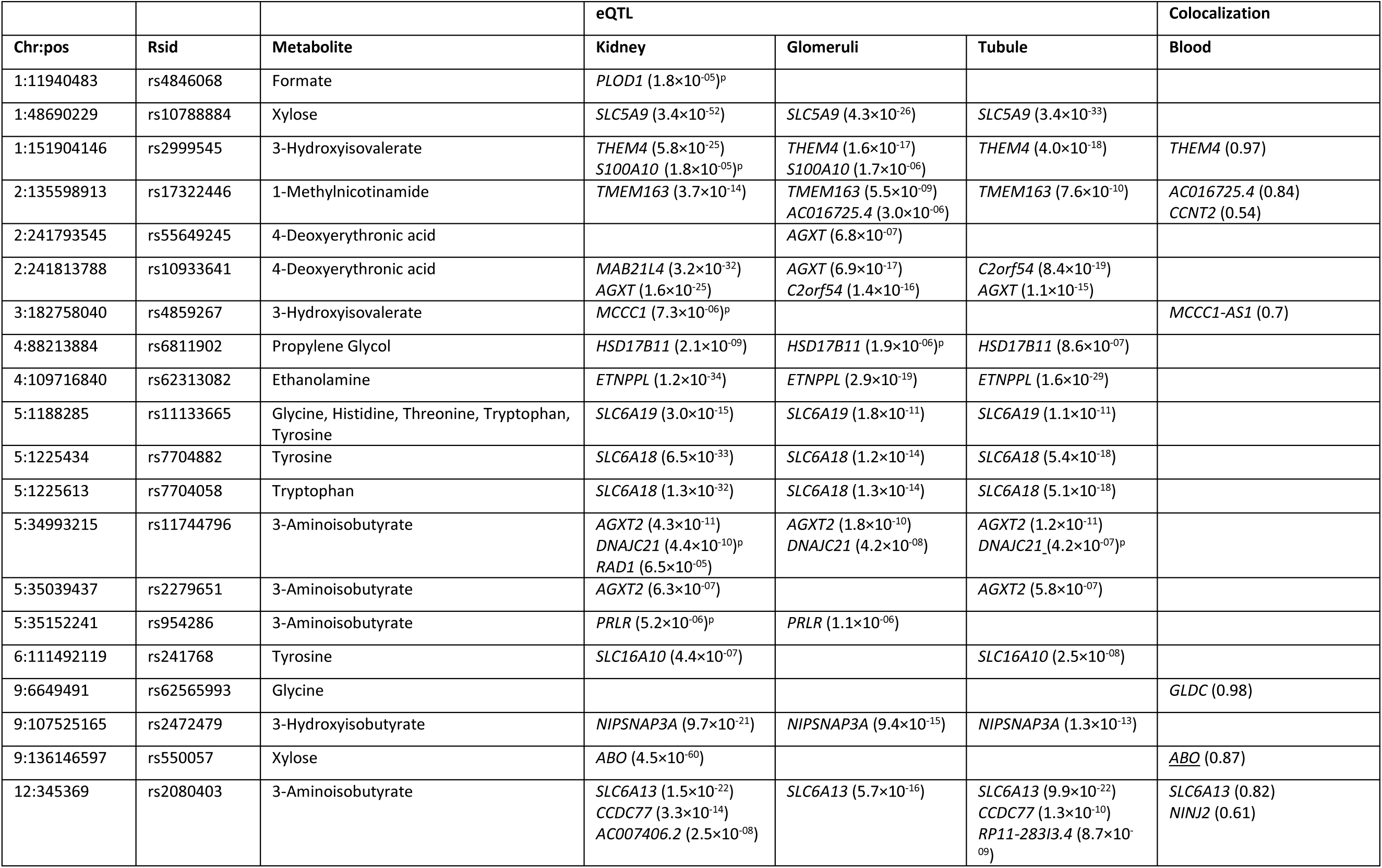
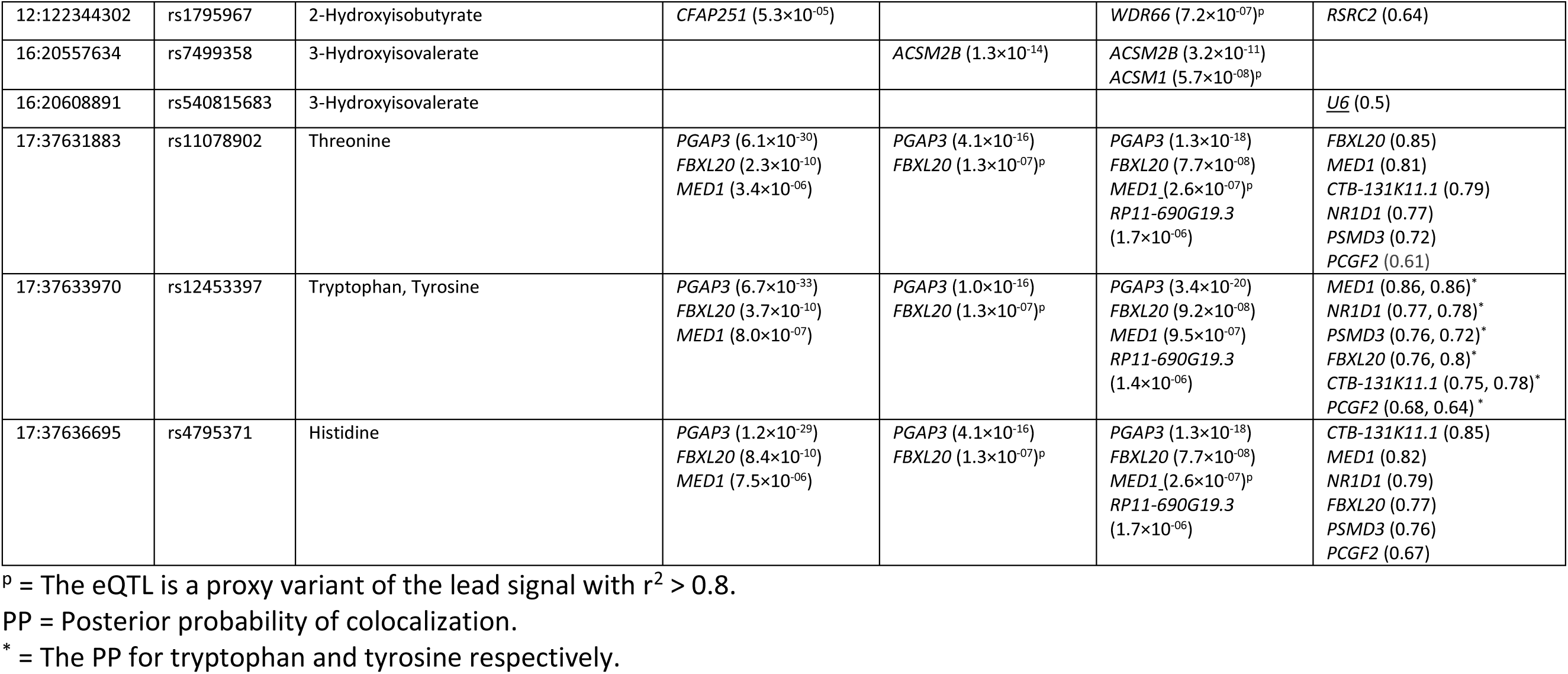
Expression quantitative trait loci (eQTL) target genes in kidney, glomeruli and tubule (p < 5.3× 10^-4^) for the COJO lead signals, and target genes of whole blood eQTL signals colocalizing (PP > 0.5) with COJO lead signals.

In the *SLC6A18 – SLC6A19* locus, the tyrosine and tryptophan-associated rs7704882 and rs7704058 (in full LD in the European population: *r*^2^=1), represent a novel metabolite association independent from a previously known intergenic metabolite locus 37 kbp away, rs11133665. Also rs11133665 was associated with tryptophan and tyrosine in our data, in addition to glycine, histidine, and threonine. The rs11133665 variant has previously been associated with urinary 6-bromotryptophan, kynurenine, tryptophan, phenylalanine, tyrosine, 3-hydroxykynurenine, and histidine/τ-methylhistidine^4,25,26^, as well as eGFR in the CKDGen data^17^ (Table 3). The rs11133665 variant was also associated with *SLC6A19* gene expression in the kidneys (Table 2). On the contrary, rs7704882 and rs7704058 are strong *SLC6A18* eQTLs for pooled kidney (6.5×10^-33^), as well as in the kidney tubules and glomeruli. However, the association signal for tyrosine around rs7704882 shows evidence of colocalization with the secondary *SLC6A19* eQTL signal (Figure 3 and Supplementary Figure 4); thus, it remains unclear whether this novel association affects *SLC6A18*, *SLC6A19*, or both. In the human protein atlas, *SLC6A18* is specifically expressed in the kidneys. *SLC6A18* encodes a sodium cotransporter for neurotransmitters, amino acids, and osmolytes like betaine, taurine, and creatine. *SLC6A19* is expressed especially in the kidney proximal tubules in the scRNAseq data^20^, and it encodes a sodium-dependent neutral amino acid transporter that mediates resorption of neutral amino acids across the apical membrane of kidney and intestinal epithelial cells. Of note, tyrosine was one of the amino acids associated with progression of DKD to kidney failure in our previous observational study^2^.

**Figure 3.**
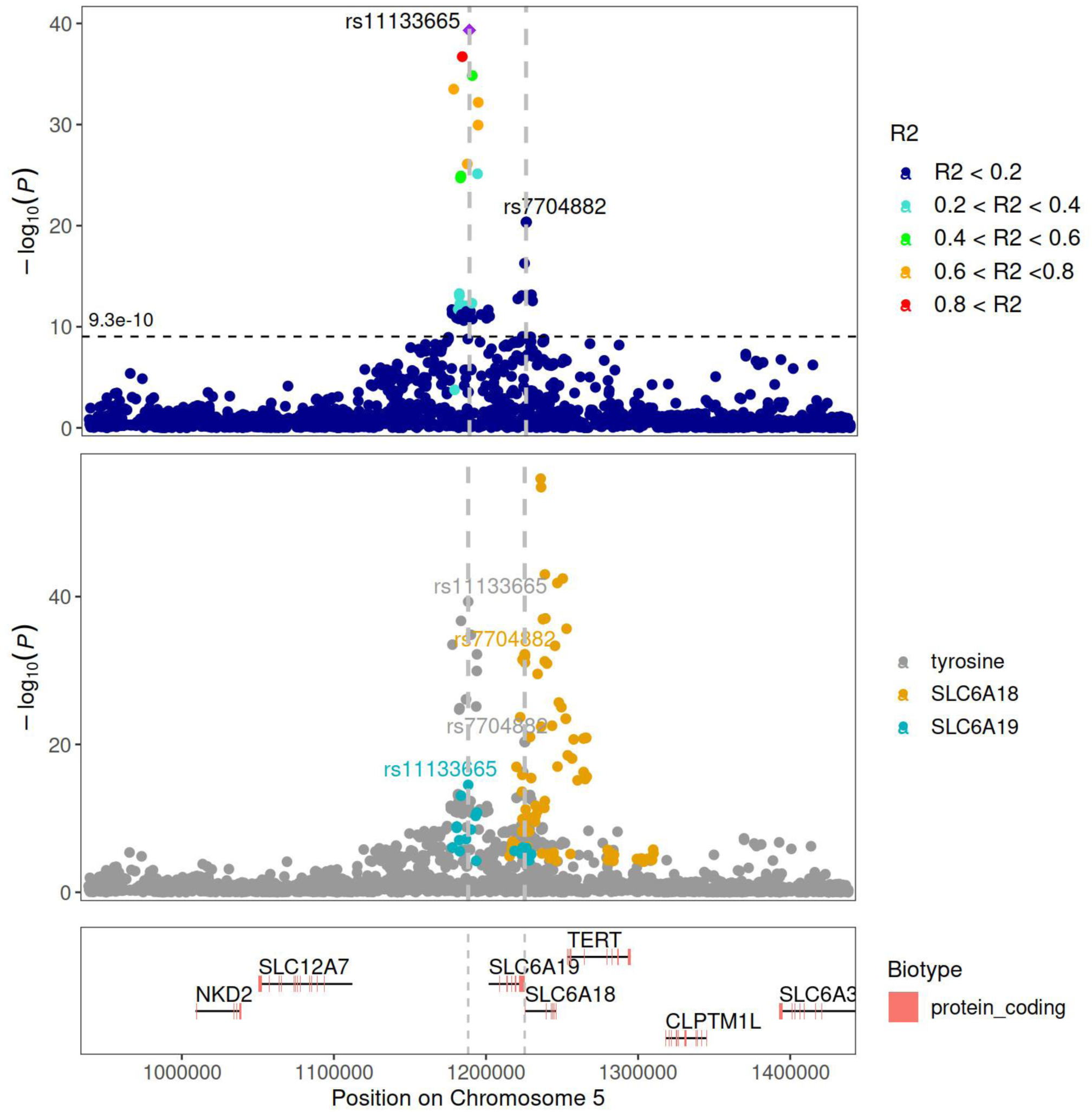
Regional association with tyrosine for lead variants rs11133665 and rs7704882 on chromosome 5. Upper panel shows LocusZoom plot centred around the previously known rs11133665 variant, and the novel signal at rs7704882 independently associated with tyrosine. The middle panel shows kidney eQTL associations for *SLC6A18* and *SLC6A19* overlaid on top of the tyrosine association signals, highlighting lead variants rs11133665 (eQTL for *SLC6A19* in kidney) and rs7704882 (eQTL for *SLC6A18* in kidney)^42^. The R2 values with rs11133665 are calculated based on 1000 Genomes phase 3 European population. Variants with no R2 information are not shown.

**Table 3:** Metabolite lead variant associations with eGFR and CKD in the CKDGen meta-analysis^17^, and DKD phenotypes in DNCRI-SUMMIT^16^ meta-analysis. All nominally significant associations (p<0.05) are shown for variants with at least one significant association (p < 3.6×10^-04^, i.e., p < 0.05 / 3 phenotypes / 46 SNPs).

| Urinary metabolite |  |  |  | eGFR |  | CKD |  | DKD |  | Phenotype |
| --- | --- | --- | --- | --- | --- | --- | --- | --- | --- | --- |
| Chr:Pos:EA:NEA | Rsid | Metabolite | Effect | Effect | P-value | Effect | P-value | Effect | P-value |  |
| 1:151904146:A:T | rs2999545 | 3-Hydroxyisovalerate | -0.11 | -0.00089 | $1.3 \times 10^{-02}$ | 0.035 | $3.5 \times 10^{-04}$ | 0.046 | $2.2 \times 10^{-02}$ | Any DKD |
| 2:211540507:A:C | rs1047891 | Glycine | 0.23 | -0.0065 | $3.6 \times 10^{-64}$ | 0.055 | $2.3 \times 10^{-07}$ | | | |
| 4:109716840:A:T | rs62313082 | Ethanolamine | 0.14 | 0.0019 | $6.1 \times 10^{-08}$ | | | | | |
| 5:1188285:A:G | rs11133665 | Glycine | -0.13 | -0.0016 | $8.6 \times 10^{-05}$ | | | | | |
| 5:1188285:A:G | rs11133665 | Histidine | -0.31 | -0.0016 | $8.6 \times 10^{-05}$ | | | | | |
| 5:1188285:A:G | rs11133665 | Threonine | -0.15 | -0.0016 | $8.6 \times 10^{-05}$ | | | | | |
| 5:1188285:A:G | rs11133665 | Tryptophan | -0.17 | -0.0016 | $8.6 \times 10^{-05}$ | | | | | |
| 5:1188285:A:G | rs11133665 | Tyrosine | -0.3 | -0.0016 | $8.6 \times 10^{-05}$ | | | | | |
| 5:150708711:C:G | rs147000073 | Glycine | -1.1 | -0.0071 | $1.1 \times 10^{-04}$ | 0.12 | $1.2 \times 10^{-02}$ | | | |
| 7:17287998:A:G | rs2106727 | Quinic acid | -0.12 | -0.0022 | $3.5 \times 10^{-10}$ | 0.032 | $9.1 \times 10^{-04}$ | 0.096 | $3.4 \times 10^{-02}$ | ESRD vs. macroalb. |
| 7:17287998:A:G | rs2106727 | Trigonelline | -0.1 | -0.0022 | $3.5 \times 10^{-10}$ | 0.032 | $9.1 \times 10^{-04}$ | 0.096 | $3.4 \times 10^{-02}$ | ESRD vs. macroalb. |
| 9:136146597:T:C | rs550057 | Xylose | 0.2 | 0.0019 | $9.2 \times 10^{-07}$ | | | 0.19 | $1.1 \times 10^{-04}$ | ESRD vs. macroalb. |
| 12:345369:C:G | rs2080403 | 3-Aminoisobutyrate | -0.16 | 0.004 | $8.5 \times 10^{-30}$ | | | 0.058 | $5.1 \times 10^{-03}$ | Any DKD |
| 12:4521511:A:T | rs78470967 | 4-Deoxythreonate | 0.34 | 0.0036 | $1.8 \times 10^{-04}$ | | | | | |
| 12:122344302:A:G | rs1795967 | 2-Hydroxyisobutyrate | -0.51 | 0.0019 | $1.9 \times 10^{-04}$ | | | | | |
| 17:37631883:C:G | rs11078902 | Threonine | -0.12 | 0.0058 | $2.1 \times 10^{-47}$ | -0.041 | $1.3 \times 10^{-04}$ | | | |
| 17:37633970:A:C | rs12453397 | Tryptophan | 0.16 | -0.0057 | $4.9 \times 10^{-46}$ | 0.041 | $1.3 \times 10^{-04}$ | | | |
| 17:37633970:A:C | rs12453397 | Tyrosine | 0.25 | -0.0057 | $4.9 \times 10^{-46}$ | 0.041 | $1.3 \times 10^{-04}$ | | | |
| 17:37636695:T:G | rs4795371 | Histidine | -0.22 | 0.0057 | $1.8 \times 10^{-46}$ | -0.041 | $1.3 \times 10^{-04}$ | | | |

Chr:Pos:EA:NEA: Chromosome position, effect allele, and non-effect allele. Rsid: variant rs-identifier. Metabolite: The associated
urinary metabolite. Effect: effect estimate. P-value: p-value of the association. Phenotype: the DKD phenotype. eGFR: estimated glomerular filtration rate. CKD: chronic kidney disease. DKD: diabetic kidney disease.

The other kidney eQTLs for solute carrier family proteins represent previously known metabolite associations: rs241768 associated with tyrosine and a kidney eQTL for *SLC16A10* (*p*=4.4×10^-07^); and rs2080403 associated with 3-aminoisobutyrate and a kidney eQTL for *SLC6A13* (*p*=1.5×10^-22^). The 3-aminoisobutyrate signal also colocalized with the *SLC6A13* blood eQTL signal (posterior probability (PP)=0.82; Table 2; Supplementary Figure 5) suggesting 3-aminoisobutyrate as an unknown substrate of *SLC6A13* as hypothesized before^4,27^.

In addition, the three variants that associated with glycine on chromosome 5q33.1 included two intronic variants, rs61067578 and rs147000073, in the *SLC36A2* gene encoding a proton-coupled amino acid transporter involved in the reabsorption of small amino acids such as glycine, proline, and alanine in the proximal tubules of the kidneys^28^. The third independent variant rs72794144 is an intronic variant in the neighbouring *GM2A* (Ganglioside GM2 Activator) gene encoding a small glycolipid transport protein. However, we did not detect any significant kidney eQTL associations for these variants.

Finally, rs62313082 upstream of *RCC2P8* gene associated with urinary ethanolamine. This variant has no previously reported associations in the GWAS catalog, however, it is a kidney eQTL for *ETNPPL* (*p*=1.2×10^-34^, Table 2), which catalyses breakdown of phosphoethanolamine^29^, and thus, represents a plausible gene underlying the metabolite association. Rs2472479 on chromosome 9 was associated with 3-hydroxyisobutyrate. The variant is located upstream of *NIPSNAP3B* (Nipsnap Homolog 3B), but was a kidney eQTL (*p*=9.7×10^-21^) for *NIPSNAP3A* (Nipsnap Homolog 3A); the genes belong to a family of proteins with putative roles in vesicular transport^30^.

We further extended the eQTL look-ups to eQTLGen whole blood data^31^, where we were able to investigate also eQTL signal colocalization with the metabolite association signals (Methods). Even though the eQTLGen data may not detect eQTLs for genes only expressed in specific target tissues such as kidneys, with 31,684 samples it has higher power to detect also weaker signals for general eQTL associations. The 52 lead variants that associated with the urinary metabolites had altogether 106 eQTLs in whole blood (*p*< 5.5×10^-5^; Supplementary Table 7), but only 34 eQTL and metabolite signal pairs showed evidence of colocalization (PP>0.5) and 24 of the 34 signals resulted from eQTL signals for 6 genes colocalizing with 4 amino acid signals in a single region on chromosome 17q12 (Table 2). Altogether 9 colocalized eQTL target genes were not detected as eQTLs in the kidney eQTL datasets. Novel but plausible findings include, e.g., a glycine-associated variant rs62565993 on chromosome 9p24.1, 4kb upstream of *GLDC (*Glycine Decarboxylase), which was a strong eQTL for *GLDC* in whole blood (*p*=2.4×10^-79^) and the glycine signal colocalizes with the eQTL signal for *GLDC* (PP=0.98). *GLDC* encodes a component of the glycine cleavage system catalysing the degradation of glycine^32^, and is a potential causal gene for the urinary glycine association.

### Gene set, pathway and tissue enrichment analyses

To gain insight into the relevant tissues and molecular pathways underlying the urinary metabolite concentrations, we performed two different types of gene set enrichment analyses (Methods). As a first approach, we used MAGMA gene set analysis that first annotates all variants, without any *p*-value threshold, to the underlying or flanking genes and evaluates the gene-level significance. MAGMA tissue expression analysis identified a positive relationship between the highly expressed genes in adipose tissue and cis-Aconitate genetic associations (*p*=6.9×10^-5^); as well as between kidney and glycine (p=1.5×10^-4^) and pituitary gland and pyroglutamate (Supplementary Table 10); confirmatory with prior studies.

MAGMA gene set enrichment analysis identified nine significantly enriched gene sets (*p*-value < 3×10^-6^, Supplementary Table 11). After the strongest enrichment between tyrosine and the positional chr5p15 breast cancer locus, the second strongest enrichment was obtained between threonine and tachykinin receptors bind tachykinins pathway (*p*=1.1×10^-7^). Of note, the five tachykinin and their receptor genes are all located in different chromosomes, thus representing a true genome-wide enrichment. Tachykinins are neuropeptides derived from alternate processing of the three tachykinin genes. They are expressed throughout the nervous and immunological system, participate in a variety of physiological processes, and contribute to multiple disease processes, including acute and chronic inflammation and pain, fibrosis, affective and addictive disorders, functional disorders of the intestine and urinary bladder, infection, and cancer^33^. Other significant gene sets included enrichment between 4-deoxyerythronic acid and pyruvate family amino acid metabolic process genes, 3-hydroxyisovalerate and uronic acid metabolic process genes, glycolic acid and eukaryotic translation initiation factor 3 complex proteins, tryptophan and genes involved in APC/C:Cdc20 mediated degradation of Cyclin B, and 4-deoxythreonate and genes involved in neuron intrinsic apoptotic signaling pathway in response to oxidative stress.

Since the gene potentially underlying the observed association in GWAS is not always the affected or closest gene, we also utilized the FUMA gene set enrichment analysis as a complementary approach. We included only variants reaching a *p*-value < 1×10^-5^, but utilised eQTL associations in addition to the positional mapping of variants to genes. Altogether 26 of the 54 metabolites were found to have significant (*p*<0.05) associations with gene set pathways following FUMA analysis (Supplementary Table 12). The metabolites 3-aminoisobutyrate, histidine, threonine, tryptophan, tyrosine and valine were associated with eGFR, and 4-deoxyerythronic acid was associated with urate concentrations, suggesting a role in kidney health.

Breast cancer was found to be most abundant with related pathways significantly associated with 12 of the metabolites, followed by asthma-related pathways which were significantly associated with 11 metabolites. Of note, the cancer gene sets typically represent single chromosomal loci with a gene cluster, rather than genome-wide enrichment. The amino acid biomarkers histidine, threonine, tryptophan, tyrosine, and valine showed very similar results being significantly associated with the same pathways. These include inflammatory bowel disease, atrial fibrillation, rheumatoid arthritis, menopause, systemic lupus erythematosus and polycystic ovary syndrome. This is due to a GWAS hit on chromosome 17q12, which is present in all 5 amino acids and thus driving most of the associations with the gene set pathways.

### Kidney health causally affects urinary metabolites

As urinary metabolites may reflect kidney health, we investigated whether the identified variants are also associated with kidney disease traits in the general population (CKDGen meta-analysis)^17^ and in individuals with diabetes (DNCRI-SUMMIT meta-analysis)^16^ (Methods). Indeed, seven of the variants were genome-wide significantly (*p*<5×10^-8^) associated with estimated glomerular filtration rate (eGFR), a main measure to monitor kidney health, and 4 variants with chronic kidney disease (CKD) (*p*<3.6×10^-4^) in the general population. Four variants were also nominally (*p*<0.05) associated with DKD or kidney failure in diabetes (Table 3).

As multiple urinary metabolite associated variants were also associated with kidney disease traits we tested whether kidney health causally affects urinary metabolite concentrations. We performed two sample Mendelian randomization analysis using two kidney function markers, eGFR and urinary albumin-creatinine ratio (UACR), as the exposures, and metabolite concentrations as the outcomes (Supplementary Table 8, Methods). A genetic instrument for eGFR, composed of 150 independent genome-wide significant SNVs, identified in the CKDGen GWAS meta-analysis^17^ was associated (*p*<4.7×10^-4^) with the urinary metabolite concentrations of 4 amino acids (alanine, glutamine, leucine, and valine), as well as 9 other metabolites: 2-hydroxyisobutyrate, 3-hydroxyisovalerate, ethanolamine, formate, glycine, glycolic acid, pseudouridine, pyroglutamate, and uracil (Supplementary Table 9, Supplementary Figure 6). This suggests a causal association of glomerular filtration rate on these urinary metabolites. For all the metabolites, higher eGFR was associated with higher metabolite concentration in the urine. The causal effects were directionally consistent across different MR analysis methods for all outcomes except for pyroglutamate and glycine. Furthermore, eGFR remained associated (*p*<0.05) with glycolic acid, 3-hydroxyisovalerate, and pseudouridine even with the MR Egger method more robust against pleiotropic effects. As eGFR causally affected multiple metabolites we tested if adjusting the GWAS analysis with eGFR affected the metabolite associations in FinnDiane. However, adjustment with eGFR had little effect on the effect estimate for the 41 COJO lead variants included in the FinnDiane GWAS data set (Supplementary Figure 7).

### Urinary metabolites potentially causally linked to kidney function and body mass index

We also performed two sample Mendelian Randomization to test whether urinary metabolites are causal risk factors or reflect causal biological processes leading to CKD and other chronic diseases (Supplementary table 8, Methods). The analysis suggested that higher urinary 3-hydroxyhippurate, quinic acid and trigonelline concentrations are causally associated with higher body mass index (BMI), lower urinary creatinine concentration and higher UACR (i.e., reflecting worse kidney health), and contradictorily, with higher eGFR (i.e., reflecting better kidney health; estimated from serum creatinine; Table 4); all three metabolites are found in coffee, and the rs2106727 and rs6968554 variants in the *AHR* locus associated with the three urinary metabolites, are in strong LD with rs4410790, that was associated with caffeine intake (*p*=2.0×10^-249^)^34^. Indeed, a previous MR study suggested that coffee consumption has a beneficial effect on kidney function and albuminuria^35^. Why the three urinary metabolites were associated with higher UACR in our data remains unclear. For BMI, previous MR studies have found contradictory evidence regarding the causality between coffee consumption and BMI or obesity^36,37^. In general, the urinary metabolites may provide a more exact estimate of the coffee intake than self-reported data on coffee consumption. However, we note that the *AHR* variants rs2106727 and rs6968554 are associated also with other traits such as blood lipid concentrations in the GWAS catalog (*p*<5×10^-8^), indicating potential pleiotropic effects.

**Table 4.** Two sample Mendelian randomization analysis results with p < 0.05 / 496 = 1.0×10^-4^ using urinary metabolites as the exposures for outcomes from IEU GWAS database, CKDGen meta-GWAS, DNCRI meta-GWAS, and DIAMANTE meta-GWAS.

| Outcome | Exposure | Rsid | Beta (SE) | P |
| --- | --- | --- | --- | --- |
| Body mass index (BMI) | 1-Methylnicotinamide | rs17322446 | 0.08 (0.02) | $2.3 \times 10^{-5}$ |
| | 3-hydroxyhippurate | rs6968554 | 0.10 (0.02) | $2.5 \times 10^{-6}$ |
| | 4-Deoxythreonate | rs181558, rs78470967 | 0.05 (0.01) | $7.6 \times 10^{-5}$ |
| | Quinic acid | rs2106727 | 0.08 (0.02) | $2.5 \times 10^{-6}$ |
| | Trigonelline | rs2106727 | 0.10 (0.02) | $2.5 \times 10^{-6}$ |
| Creatinine (enzymatic) in urine | 3-hydroxyhippurate | rs6968554 | -0.22 (0.03) | $3.4 \times 10^{-18}$ |
| | Quinic acid | rs2106727 | -0.17 (0.02) | $5.1 \times 10^{-18}$ |
| | Trigonelline | rs2106727 | -0.20 (0.02) | $5.1 \times 10^{-18}$ |
| eGFR | 3-hydroxyhippurate | rs6968554 | 0.02 (0.00) | $3.7 \times 10^{-10}$ |
| | Ethanolamine | rs62313082 | 0.01 (0.00) | $6.1 \times 10^{-8}$ |
| | Quinic acid | rs2106727 | 0.02 (0.00) | $3.5 \times 10^{-10}$ |
| | Trigonelline | rs2106727 | 0.02 (0.00) | $3.5 \times 10^{-10}$ |
| UACR | 3-hydroxyhippurate | rs6968554 | 0.23 (0.02) | $3.5 \times 10^{-25}$ |
| | Quinic acid | rs2106727 | 0.18 (0.02) | $7.6 \times 10^{-25}$ |
| | Trigonelline | rs2106727 | 0.21 (0.02) | $7.6 \times 10^{-25}$ |

Results involving one variant were obtained using Wald ratio and two variants with inverse variance weighting. UACR = urinary albumin to creatinine ratio. eGFR = estimated glomerular filtration rate.

In addition, the genetic instrument for urinary ethanolamine, was associated with higher eGFR (*p*=6.1×10^-8^). The genetic instrument was based on the rs62313082 variant which is also a kidney eQTL for *ETNPPL* gene (Table 2 and 4). Moreover, MR analysis suggested that genetic instruments for urinary 1-methylnicotinamide (*p*=2.3×10^-5^) and 4-deoxythreonate (*p*=7.6×10^-5^) were associated with higher body mass index (BMI; Table 4). However, all significant MR analysis findings were based on only one or two significant variants available for each metabolite and need to be interpreted with caution; the largest number - 8 genetic variants – were available for MR for 3-hydroxyisovalerate, which was not associated with any of the studied outcomes (*p*>0.01).

## DISCUSSION

To our knowledge this meta-analysis of three large cohorts represents the largest GWAS on urinary NMR metabolomics to date, enabling us to detect previously unidentified associations with urinary metabolites. We identified 52 genetic associations with urinary metabolites, of which 31 were novel. In line with the notion that GWAS findings for complex diseases are enriched for regulatory variants^38^, many of the metabolite associations were outside genes, but were strongly associated with gene expression in the whole kidney, tubules and glomeruli, for example, solute carriers *SLC6A18* and *SLC6A19*. While it is overall not surprising to find kidney associations for urinary metabolites, our findings may help to further describe how the kidneys regulate systemic metabolism by filtration and reabsorption.

Additionally, we observed 4 amino acids and 9 additional metabolites whose urinary concentrations were causally influenced by the glomerular filtration rate in the kidneys. This finding suggests that the glomerular filtration rate needs to be considered when investigating these metabolites as potential biomarkers of disease risk. On the other hand, MR analysis suggested that urinary ethanolamine was associated with higher eGFR lending support for a potential causal protective role. The association was based on the rs62313082 variant which is associated with higher urinary ethanolamine concentration, higher eGFR, and lower *ETNPPL* gene expression in the kidneys. The *ETNPPL* gene encodes for Ethanolamine-Phosphate Phospho-Lyase that catalyses the breakdown of phosphoethanolamine. Ethanolamine is an initial precursor for phosphoethanolamine and for the biosynthesis of two primary phospholipid classes, phosphatidylcholine (PC) and phosphatidylethanolamine (PE), as well as sphingophospholipid and a variety of N-acylethanolamines. The *ETNPPL* gene was recently implicated also in hyperinsulinemia-induced insulin resistance^39^.

Furthermore, MR analysis suggested 1-methylnicotinamide as a causal risk factor for BMI. Indeed, serum levels of 1-methylnicotinamide were positively correlated with BMI in observational setting^40^, and a caloric restriction and exercise intervention suggested that 1-methylnicotinamide enhances the utilization of energy stores in response to low muscle energy availability^41^. Thus, our findings support the previous suggestion of 1-methylnicotinamide as an early marker for metabolic disease^41^. Altogether, our findings, and the genome-wide metabolite results could be utilized to test and support biological hypotheses originating from observational studies.

Our study included individuals with reduced glomerular filtration rate potentially enhancing our power to detect associations as previous studies have shown that genetic studies on urinary metabolites in individuals with CKD can detect signals that would be harder to detect in the general population alone^25^. It is however important to note, that adding eGFR as a kidney filtration covariate in the GWAS did not have a significant impact on our genetic association results. As a limitation, the study participants were mostly of European origin and further studies are required to investigate generalizability of our findings to other populations.

Altogether, we provide a catalogue of genetic associations for 53 metabolites, which can be utilized, for example, to investigate how urinary metabolites are linked to human health and disease risk.

## ACKNOWLEDGEMENTS

We acknowledge the skilled technical assistance of Heli Krigsman, Hanna Olanne, Maikki Parkkonen, Mira Korolainen, Anna Sandelin, Jaana Tuomikangas, and Kirsi Uljala (Folkhälsan Research Center, Finland), and all the physicians and nurses at each FinnDiane study center taking part in the enrolment and clinical characterization of the participants (Supplementary table 13 for a list of study centers and investigators involved in the FinnDiane study).

We are grateful to all the families who took part, the general practitioners and the Scottish School of Primary Care for their help in recruiting them, and the whole Generation Scotland team, which includes interviewers, computer and laboratory technicians, clerical workers, research scientists, volunteers, managers, receptionists, healthcare assistants and nurses.

The Viking Health Study DNA extractions and genotyping were performed at the Edinburgh Clinical Research Facility, University of Edinburgh. We would like to acknowledge the invaluable contributions of the research nurses in Shetland, the administrative team in Edinburgh and the people of Shetland.

For the purpose of open access, the author has applied a Creative Commons Attribution (CC BY) licence to any Author Accepted Manuscript version arising from this submission.

## AUTHOR CONTRIBUTIONS

EV contributed to design of the study, data analysis, interpretation of the results, and drafted the manuscript. AR contributed to data analysis, manuscript writing, and interpretation of the results. SM contributed to metabolite data quantification, manuscript writing and interpretation of the results. DP and AC contributed to data acquisition. JFW contributed to data acquisition. PHG contributed to design of the study. CH and NS contributed to design of the study, data analysis, interpretation of the results, and manuscript writing. All authors critically read and approved the final version to be submitted and published, and agree to be accountable for all aspects of the work in ensuring that questions related to the accuracy or integrity of any part of the work are appropriately investigated and resolved.

## DATA AVAILABILITY STATEMENT

The GWAS meta-analysis summary statistics for the 54 studied metabolites except glucose will be publicly available at GWAS Catalog (https://ebi.ac.uk/gwas).

The individual-level FinnDiane, VIKING and Generation Scotland metabolomics and GWAS datasets analysed during the current study are not publicly available as the patients’ written consent does not allow data sharing. The Readers may propose collaboration to research the individual level data with correspondence with the lead investigators.

## ETHICS APPROVAL AND CONSENT TO PARTICIPATE

The FinnDiane study protocol was approved by the Ethical Committee of the Helsinki and Uusimaa Hospital District (491/E5/2006, 238/13/03/00/2015, and HUS-3313-2018, July 3rd, 2019) and the participants gave their informed consent before recruitment. The FinnDiane study was performed following the Declaration of Helsinki.

All the Viking Health Study – Shetland (VIKING) participants gave informed consent and the study was approved by the South East Scotland Research Ethics Committee, NHS Lothian (reference: 12/SS/0151). The VIKING study was performed in accordance with the Declaration of Helsinki.

All the GS:SFHS participant gave informed consent and the ethical approval for the study was obtained from the Tayside Committee on Medical Research Ethics (on behalf of the National Health Service, reference 05/S1401/89). The GS:SFHS study was performed in accordance with the Declaration of Helsinki.

## METHODS

### Study cohorts

Generation Scotland (GS) is a family-based population study of around 20,000 individuals from across Scotland^11^. Individuals aged between 35 and 65 years were recruited at random from 2006 to 2010 from collaborating medical practices. These participants then identified ≥first degree relatives who would also be able to participate, resulting in a final age range of 18 to 98 years. Participants attended a staffed research clinic where they completed a health questionnaire, had physical and clinical characteristics measured and fasting blood and urine samples collected according to standard operating procedures. Serum and urine aliquots were stored at -80°C for future analyses.

VIKING is a family-based population study of over 2,000 individuals from the population isolate of the Shetland Isles in northern Scotland^12^. Recruitment ran from 2013 to 2015 with the selection criteria requiring individuals to be ≥18 years and have two or more grandparents born in the Shetland Isles. Over 90% of resulting participants had three or four grandparents from Shetland and most were related individuals from large kindreds. Participants attended clinics where physical characteristics were measured and fasting blood and urine samples were collected according to standard operating procedures. Plasma, serum, whole blood and urine aliquots were stored at -80°C for future analyses.

Finnish diabetic nephropathy study (FinnDiane) is an ongoing (1997->) nation-wide multicentre study focusing on diabetic complications and currently comprises of over 5,000 adults with type 1 diabetes^9,10^. In this study we included individuals with genotype data and urinary metabolite data measured from 24h urine collection, except one overnight urine collection, stored at -20C°. In addition, to assure correct diagnosis of T1D, we required age at onset of diabetes < 40 years and insulin treatment initiated within one year from diabetes diagnosis. Moreover, since kidney function may affect the urinary metabolite levels, we excluded Individuals with prevalent end-stage kidney disease (ESKD), defined as kidney transplantation or dialysis treatment, and individuals with eGFR < 10 mL/min/1.73m^2^, at urine collection day. Finally, 3,244 individuals were included in the analysis.

### Metabolite quantification by NMR

The urinary metabolite quantification has been described previously^2^. Briefly, metabolite quantification of the urine samples was performed using a proprietary NMR metabolite profiling service (Nightingale Health, Helsinki, Finland). The NMR-based measurements were conducted from 500 μl of stored samples using a 600 MHz Bruker AVANCE III HD NMR spectrometer (Bruker BioSpin, Switzerland) with automated sample changer and cryoprobe. The spectral data were acquired using standard water-suppressed acquisition settings. The sample preparation and NMR acquisition parameters were designed for high-throughput initially selecting metabolites based on feasibility for automated quantification. This approach emphasises metabolites at high abundance in urine, and those which generate minimal signal overlap in the proton NMR spectrum. As such, the metabolite selection was not based on prior biological relevance of the selected metabolites or emphasis of certain metabolic pathways. The urinary metabolite concentrations were divided by urinary creatinine concentration to normalise for urine volume.

### Genotyping and imputation

GS and VIKING samples were genotyped using the Illumina HumanOmniExpressExome-8v1-2 chip (Illumina, San Diego, CA) and individuals with a call rate of ≤ 98% and SNPs with a call rate of ≤ 98%, HWE of ≤ 1×10^-06^ and a MAF of ≤ 1% were excluded during quality control. Phasing was carried out using SHAPEIT (v2 r837) and imputation was performed using the Haplotype Reference Consortium reference panel (HRC.r1-1) on the Sanger Imputation Server with the PBWT software. Post imputation quality control excluded duplicate and monomorphic variants and SNPs with an imputation quality score of < 0.4.

FinnDiane samples was genotyped using the HumanCoreExome-12 v1.0, -12 v1.1, and -24 v1.0 BeadChips (Illumina, San Diego, CA). The quality control and data processing has been described in more detail before^43,44^. In short, SNVs with call rate of ≤ 95% or excessive deviation from Hardy-Weinberg equilibrium were excluded, Haplotypes were phased with SHAPEIT (v2 r837) and genotypes imputed with Minimac3 (v1.0.14) using 1000 Genomes phase 3 version 5 as the reference panel.

### GWAS analysis, meta-analysis with Metal, and GCTA-COJO

Study-level GWAS analysis was conducted separately for each cohort and the results were first quality controlled and harmonized before meta-analysis, and finally, a conditional joint analysis was performed to identify SNVs independently associated with urinary metabolites. Before GWAS analysis urinary metabolite to creatinine ratios and creatinine values were regressed on the covariates and the residuals were inverse normal transformed.

GS and VIKING GWAS were performed using RegScan accounting for relatedness within each cohort^45^. The analysis model included age and sex as covariates. FinnDiane GWAS was executed with SNPTest (v2.5.2). Before the analysis, first-degree relatives were removed preferring individuals with most complete metabolite data until no first-degree relative pairs were left in the data set. Two models were fitted: minimal model included age, sex, genotyping batch and two first genetic principal components as covariates; full model included minimal model and eGFR. The association of genetic variants with urinary metabolites was tested using a frequentist test and an additive model applying the score method to account for genotype uncertainty.

Before the meta-analysis study-level quality control was performed with EasyQC R-package (v9.2, www.genepi-regensburg.de/easyqc)^46^. First, any association results with missing or implausible data, monomorphic variants, and variants with imputation quality < 0.4 were removed, second, allele coding and marker names were harmonized and possible duplicates were removed, finally, variants were checked against the appropriate reference data and any variants with mismatching alleles or allele frequency difference >0.2 compared to the reference were removed.

Meta-analysis of the individual GWAS was performed using METAL software (version 2011-03-25) applying inverse variance weighted method and genomic control correction^47^.The results were filtered to include variants with MAF ≥ 0.01 and found at least in 2 out of 3 studies. Signals for the same metabolite were considered distinct if they were at least 3Mbp apart.

Approximate conditional and joint GWAS analysis was performed to identify SNVs independently associated with urinary metabolites applying the GTCA-COJO software (v1.93.2beta)^13,48^. The filtered METAL results were used as the input and whole FinnDiane cohort (n=6019) was used as the reference population to estimate LD. Default options were used to perform stepwise model selection to select independently associated SNVs. Association results for glucose were spurious and are not reported.

The regional association signal around the COJO lead variants was visualized using LocusZoom stand-alone software (v1.4, http://genome.sph.umich.edu/wiki/LocusZoom_Standalone). The LD information was calculated using the 1000 Genomes phase 3 European population.

### Annotation of the COJO lead variants

The lead SNPs from COJO analysis were annotated with genes and variant effects with Ensembl Variant Effect Predictor (VEP) web tool (Assembly GRCh38.14 version 110). The SNPs were queried for all consequences. The most severe consequence per SNP and gene was selected based on the severity as estimated by Ensembl (www.ensembl.org/info/genome/variation/prediction/predicted_data.html accessed 2023-11-14). If the variant was not located in a transcript the closest gene was selected.

### Heritability analysis

The heritability of the urinary metabolites was analysed in each cohort. In FinnDiane GTCA tool (v1.93.2beta) was utilized to estimate the genetic relationship matrix which was filtered to not include any individuals with relatedness greater than 0.025. The variance explained by all the SNVs was estimated by restricted maximum likelihood (REML) analysis (GTCA-GREML) using the default options and adjusting for age, sex, eGFR, genotyping batch, and two first genetic principal components^48,49^. In GS and VIKING heritability was estimated using a variance component model available within the RegScan GWAS pipeline. The heritability estimates were meta-analysed with random-effects model utilizing the inverse variance method and between study variance τ^2^ was estimated with restricted maximum-likelihood estimator. The between study heterogeneity was tested with Q-test. Analysis was performed with R-package meta (v.6.5-0).

### Additional phenotypic data

Kidney function was quantified by eGFR calculated with the CKD-EPI formula^50^ from serum and plasma creatinine values, and, in addition, by urinary albumin excretion rate (AER) in the FinnDiane cohort: normal AER (AER ≤ 30mg/24h), moderate albuminuria (30mg/24h < AER ≤ 300mg/24h), and severe albuminuria (AER > 300mg/24h). Albuminuria category was determined as the highest category in at least 2 out of 3 consecutive determinations.

### eQTL analysis in kidney and whole blood

Expression quantitative trait locus (eQTL) associations in cis were queried for the variants associated with urinary metabolites. We utilized kidney eQTL data from microdissected human kidney tubule (N=356) and glomeruli (N=303) samples^51^, and meta-analysis of 686 kidney samples^42^ downloaded from https://susztaklab.com/Kidney_eQTL/download.php. Cis-eQTL associations in whole blood were queried from eQTLGen data set^31^ (https://eqtlgen.org/cis-eqtls.html and IEU GWAS database).

In the kidney data sets we identified eQTLs and their target genes at the urinary metabolite lead variants and any additional eQTLs at proxies of the lead variants (R^2^ > 0.8) using R-package LDlinkR (v.1.2.3). In total, we found 95 candidate eQTL target genes and selected eQTLs with p < 0.05 / 95 = 5.35×10^-4^. However, we only had access to kidney eQTL data sets pre-filtered to include signals with FDR < 0.05 (tubule and glomeruli) and FDR < 0.01 (kidney meta-analysis) and consequently all candidate eQTLs were significant.

In the whole blood cis-eQTL data we identified 911 eQTLs at the lead variants and selected eQTLs with p < 0.05 / 911 = 5.5×10^-5^ as significant. Furthermore, we tested for colocalization of cis-eQTL signal for gene expression in blood with urinary metabolite signals. First, we identified genes with cis-eQTLs at the lead loci, if no genes were found we selected all genes with cis-eQTLs within 100kbp from the lead locus. Second, we tested colocalization between the genes eQTL signal and the urinary metabolite signal in a region extending 250kbp from the lead locus with R-package coloc (v.5.1.0.1). More specifically, we used the Bayesian colocalization analysis assuming one causal variant for each trait implemented in *coloc.abf* function, and calculated a posterior probability (PP) for one common causal variant^52^. Signals were considered colocalized if PP > 0.5.

### GWAS look-ups: GWAS catalog, CKDGen, and DNCRI-SUMMIT

Lead variant associations were queried (2023-09-21) for previously reported associations from the GWAS catalog using R packages LDlinkR (v.1.2.3) and gwasrapidd (v.0.99.14). We included previous associations with r^2^ > 0.8 within -/+500,000 bp from the lead variant and with p < 5×10^-8^ using the 1000 Genomes European population as the reference. The previously reported traits were classified as urinary metabolite, blood metabolite or other trait by searching for key words in the phenotype description and p-value annotation. Urinary metabolite traits were matched with regular expression “*urinary metabolite*”, and blood metabolites with “*serum metabolite|blood metabolite|serum uric acid levels|blood urea nitrogen levels*”. Furthermore, targeted lookups were performed for kidney related traits from the CKDGen consortium meta-analyses on CKD and eGFR in individuals with European ancestry from the general population^17^ available from https://ckdgen.imbi.uni-freiburg.de; and for DKD phenotypes from the DNCRI-SUMMIT meta-analysis^16^ available from https://t2d.hugeamp.org/downloads.html.

## FUMA

FUMA v.1.3.7^54^ web interface was used to perform MAGMA v1.08 tissue specificity (GTEx v8) and gene set enrichment analysis. SNPs were mapped to the protein coding genes within 10kb windows (with unique Ensembl ID). For the gene set enrichment analysis, 15,496 gene sets (5,500 curated gene sets (9 data resources including KEGG, Reactome and BioCarta), 9996 GO terms (biological processes (bp), cellular components (cc) and molecular functions (mf))) from MsigDB v7.0 were included and run with default parameters. Gene-sets with p-value < 3×10^-6^ were defined as significant by MAGMA Bonferroni correction. For tissue expression analysis, gene expression data sets were obtained from GTEx v8. MAGMA gene-property test was performed for average gene-expression per category (e.g. tissue type) conditioning on average expression across all categories (one-side) to test the positive relationship between gene expression in a specific tissue and genetic associations.

In addition, gene set enrichment was performed using the GENE2FUNC tool in FUMA v.1.3.7^54^. Genes annotated to SNPs with a p-value of < 1×10^-5^ were used as input. The list of genes was compared to a set of 19,283 background genes using hypergeometric tests to determine overrepresentation of biological functions. A minimum number of two genes per gene set and an FDR Benjamini-Hochberg adjusted p-value of < 0.05 were required for gene sets to be reported.

### Mendelian randomization analysis

We performed two sample Mendelian randomization analysis to test if kidney function, measured by estimated glomerular filtration rate (eGFR) or urinary albumin creatine ratio (UACR), causally affects urinary metabolite concentrations, and conversely, if urinary metabolite levels causally affect kidney function or other traits including type 2 diabetes (T2D), BMI, and kidney disease (Supplementary table 8).

In the first analysis we used two kidney function markers, eGFR and UACR, as the exposures and urinary metabolites as the outcomes. As the instrumental variables (IV) for eGFR we used 225 variants associated with eGFR (150 independent variants after clumping) in a European ancestry sub-analysis with 567,460 individuals (*p* < 5×10^-8^) from the CKDGen consortium^17^, and as the IV for UACR we used 61 variants associated with UACR (51 independent variants after clumping) in the European ancestry analysis with 547,361 individuals (*p* < 5×10^-8^) from the CKDGen consortium^53^. We utilized only genome-wide significantly associated variants to ensure that the IVs are strongly associated with the exposures.

In the second analysis, we employed urinary metabolites as the exposures. We selected IVs to be the variants associated with urinary metabolites in the COJO analysis with *p* < 5×10^-8^ (Supplementary Table 14). As outcomes we used 10 DKD traits from the JDRF DNCRI GWAS^43^ (downloaded from https://t2d.hugeamp.org/downloads.html), 3 traits from CKDGen consortium related to kidney function (downloaded from https://ckdgen.imbi.uni-freiburg.de), T2D from the DIAMANTE consortium (https://t2d.hugeamp.org/downloads.html), and 6 traits from the IEU GWAS database (Supplementary Table 8).

Both analyses were performed with TwoSampleMR R package (v.0.5.6). Shortly, the genetic variants for the exposures were first clumped using a 10,000kb window, a clumping R-square cut-off of 0.001, and 1000 Genomes European samples to estimate LD. Second, the effect alleles were harmonized between the exposure and outcome GWASes, and finally MR analysis was performed with default MR methods in the TwoSampleMR package: We used inverse variance-weighted (IVW) regression if at least 2 variants remained as valid IVs for the exposure, or Wald’s ratio test if only one variant was available. For exposures with 3 or more IVs, causality was further assessed using methods less sensitive to pleiotropy/heterogeneity (weighted median, simple and weighted mode, and MR-Egger regression). All the steps were performed using the default options.

