## Supplementary Information for "Genome-wide characterization of 54 urinary metabolites reveals molecular impact of kidney function"

**Supplementary Table 1.** Characteristics of the study cohorts.

**Supplementary Table 2.** Urinary metabolite characteristics in FinnDiane, GS and VIKING.

**Supplementary Table 3.** Urinary metabolite names and database ids.

**Supplementary Table 4.** Heritability estimates for the urinary metabolites.

**Supplementary Table 5.** Genome-wide significant associations with urinary metabolites.

**Supplementary Table 6.** Previously reported traits for the lead variants from the GWAS catalog.

**Supplementary Table 7.** Whole blood eQTLs at the COJO lead signals.

**Supplementary Table 8.** Mendelian randomization analysis outcomes and exposures.

**Supplementary Table 9.** Mendelian randomization analysis of kidney trait exposure on urinary metabolites.

**Supplementary Table 10.** MAGMA tissue enrichment analysis, tissues with p<0.01.

**Supplementary Table 11.** MAGMA v1.6 gene set analysis for “Curated gene sets” and “GO terms”.

**Supplementary Table 12.** FUMA GENE2FUNC gene set analysis results.

**Supplementary Table 13.** FinnDiane physicians and nurses participating in the collection of the FinnDiane study subjects.

**Supplementary Table 14.** Instruments for the urinary metabolites in the two sample MR analysis.

**Supplementary Figure 1.** Heritability estimates for the urinary metabolites.

**Supplementary Figure 2.** LocusZoom plots of the COJO lead signals.

**Supplementary Figure 3.** *AGTX2* expression in the kidneys.

**Supplementary Figure 4.** Tyrosine GWAS signal vs. *SLC6A18* and *SLC6A19* kidney eQTL signal.

**Supplementary Figure 5.** Regional association with 3-aminoisobutyrate and blood and kidney eQTL target genes

**Supplementary Figure 6.** Two-sample Mendelian Randomization results scatter plots for association of eGFR with urinary metabolites

**Supplementary Figure 7.** Effect of eGFR adjustment on the metabolite associations.

### Supplementary Table 1

**Characteristics of the study cohorts.**

|  | **FinnDiane** **(N=3,244)** | **GS**  **(N=2,743)** | **VIKING**  **(N=2,027)** |
| --- | --- | --- | --- |
| **Age (years)** |  |  |  |
| Mean (SD) | 37.7 (12.2) | 55.8 (10.5) | 49.8 (15.2) |
| Median [Min, Max] | 36.7 [13.8, 78.4] | 57 [18, 93] | 49.7 [18, 92] |
| **Sex** |  |  |  |
| M | 1632 (50.3%) | 1373 (50.05%) | 812 (40.1%) |
| F | 1612 (49.7%) | 1370 (49.95%) | 1215 (59.9%) |
| **BMI (kg/m²)** |  |  |  |
| Mean (SD) | 25.1 (3.53) | 27.0 (5.0) | 27.4 (4.88) |
| Median [Min, Max] | 24.7 [15.0, 46.5] | 26.3 [15.9, 71.3] | 26.7 [17.0,50.9] |
| Missing | 63 (1.9%) | 7 (0.25%) | 4 (0.2%) |
| **Diabetes** |  |  |  |
| Type 1 diabetes | 3244 (100%) | 18 (0.065%) |  |
| Type 2 diabetes | 0 (0%) | 170 (6.2%) | 36 (1.8%) |
| No | 0 (0%) | 2500 (91%) | 1987 (98.0%) |
| **eGFR (mL/min/1.73m²)** |  |  |  |
| Mean (SD) | 98.2 (25.1) | 87.7 (16.9) | 93.1 (16.6) |
| Median [Min, Max] | 102 [10.2, 190] | 87 [17, 167] | 94.3 [30.6, 132] |
| Missing | 5 (0.2%) | 67 (2.4%) | 16 (0.8%) |
| **CKD category** |  |  |  |
| CKD1 | 2267 (69.9%) | 1151 (42%) | 1234 (60.9%) |
| CKD2 | 712 (21.9%) | 1432 (52%) | 722 (35.6%) |
| CKD3 | 185 (5.7%) | 90 (3.3%) | 55 (2.7%) |
| CKD4 | 59 (1.8%) | 3 (0.01%) | 0 (0%) |
| CKD5 | 16 (0.5%) | 0 (0%) | 0 (0%) |
| Missing | 5 (0.2%) | 67 (2.4%) | 16 (0.8%) |
| **Albuminuria status** |  |  |  |
| Normal AER | 2233 (68.8%) | NA | NA |
| Moderate albuminuria | 441 (13.6%) | NA | NA |
| Severe albuminuria | 427 (13.2%) | NA | NA |
| Missing | 143 (4.4%) | 2743 (100%) | 2027 (100%) |

### Supplementary Table 2

**Urinary metabolite characteristics in FinnDiane, GS and VIKING**. Metabolite values are given as metabolite to creatinine ratios for all metabolites except creatinine.

| **Metabolite** | **FinnDiane (n=3,244)** | **GS (n=2,743)** | **VIKING (n=2,028)** |
| --- | --- | --- | --- |
| **1-methylnicotinamide (μmol/mmol)** |  |  |  |
| mean (SD) | 5.776 (6.692) | 3.193 (3.72) | 3.751 (3.02) |
| median [range] | 4.81 [0.00385, 150.6] | 2.502 [0, 51.91] | 3.384 [0, 32.21] |
| missing | 0 (0%) | 0 (0%) | 0 (0%) |
| **2-furoylglycine (μmol/mmol)** |  |  |  |
| mean (SD) | 27.06 (18.56) | 4.907 (7.58) | 2.5 (3.05) |
| median [range] | 24.41 [0.5641, 176.8] | 2.858 [0.362, 162.4] | 1.714 [0.34, 49.34] |
| missing | 74 (2.3%) | 188 (6.9%) | 165 (8.1%) |
| **2-hydroxyisobutyrate (μmol/mmol)** |  |  |  |
| mean (SD) | 6.068 (2.276) | 5.779 (1.67) | 4.781 (1.53) |
| median [range] | 5.596 [1.648, 24.75] | 5.631 [1.767, 24.44] | 4.575 [1.181, 15.11] |
| missing | 19 (0.6%) | 13 (0.5%) | 4 (0.2%) |
| **3-(3-hydroxyphenyl)-3-hydroxypropionic acid (μmol/mmol)** |  |  |  |
| mean (SD) | 54.95 (47.22) | 10.31 (13.84) | 8.273 (11.35) |
| median [range] | 42.81 [0.2714, 392.4] | 5.21 [0, 170.7] | 4.097 [0, 124.4] |
| missing | 407 (12.5%) | 351 (12.8%) | 255 (12.6%) |
| **3-aminoisobutyrate (μmol/mmol)** |  |  |  |
| mean (SD) | 6.218 (8.976) | 1.834 (4.25) | 1.946 (4.09) |
| median [range] | 3.113 [0.001629, 117.1] | 0.111 [0, 41.76] | 0.511 [0, 39.32] |
| missing | 1116 (34.4%) | 364 (13.3%) | 184 (9.1%) |
| **3-hydroxyhippurate (μmol/mmol)** |  |  |  |
| mean (SD) | 65.27 (57.65) | 21.64 (15.3) | 19.19 (14.04) |
| median [range] | 48.42 [2.826, 702.2] | 17.96 [2.682, 167.7] | 15.5 [2.615, 147.4] |
| missing | 63 (1.9%) | 381 (13.9%) | 353 (17.4%) |
| **3-hydroxyisobutyrate (μmol/mmol)** |  |  |  |
| mean (SD) | 22.91 (14.46) | 7.894 (4.02) | 7.134 (3.22) |
| median [range] | 19.77 [0.9126, 165.2] | 7.066 [1.368, 55.8] | 6.447 [1.162, 36.49] |
| missing | 3 (0.1%) | 95 (3.5%) | 63 (3.1%) |
| **3-hydroxyisovalerate (μmol/mmol)** |  |  |  |
| mean (SD) | 7.934 (4.543) | 3.361 (1.87) | 3.393 (2.1) |
| median [range] | 7.145 [0.04425, 62.02] | 3.073 [0, 19.26] | 2.994 [0, 39.99] |
| missing | 3 (0.1%) | 0 (0%) | 1 (0%) |
| **3-methylhistidine (μmol/mmol)** |  |  |  |
| mean (SD) | 69.67 (64.9) | 69.63 (49.74) | 83.29 (64.05) |
| median [range] | 46.27 [10.06, 612.8] | 54.38 [8.869, 461.3] | 62.77 [9.788, 357.5] |
| missing | 1173 (36.2%) | 1246 (45.4%) | 624 (30.8%) |
| **4-deoxyerythronic acid (μmol/mmol)** |  |  |  |
| mean (SD) | 8.438 (4.354) | 7.818 (3.26) | 8.313 (3.59) |
| median [range] | 7.504 [1.82, 58.38] | 7.279 [1.124, 35.14] | 7.777 [0.914, 43.84] |
| missing | 213 (6.6%) | 120 (4.4%) | 69 (3.4%) |
| **4-deoxythreonate (μmol/mmol)** |  |  |  |
| mean (SD) | 26.32 (13.61) | 15.15 (7.54) | 13.9 (5.4) |
| median [range] | 23.14 [4.923, 161.6] | 13.61 [4.226, 153.1] | 12.82 [4.52, 70.17] |
| missing | 289 (8.9%) | 84 (3.1%) | 26 (1.3%) |
| **4-hydroxyhippurate (μmol/mmol)** |  |  |  |
| mean (SD) | 12.66 (9.786) | 9.071 (7.5) | 8.549 (7.12) |
| median [range] | 9.983 [1.823, 130.9] | 7.143 [1.092, 134] | 6.838 [2.095, 127.7] |
| missing | 27 (0.8%) | 76 (2.8%) | 68 (3.4%) |
| **acetate (μmol/mmol)** |  |  |  |
| mean (SD) | 70.58 (128.3) | 6.369 (31.98) | 4.977 (25.64) |
| median [range] | 26.08 [0.002437, 998.2] | 0.68 [0, 749.5] | 0 [0, 415.2] |
| missing | 9 (0.3%) | 4 (0.1%) | 0 (0%) |
| **alanine (μmol/mmol)** |  |  |  |
| mean (SD) | 50.64 (26.3) | 20.55 (8.54) | 20.03 (9) |
| median [range] | 45.45 [8.999, 365.3] | 18.9 [6.526, 113.9] | 18.18 [4.81, 123.1] |
| missing | 2 (0.1%) | 47 (1.7%) | 30 (1.5%) |
| **allantoin (μmol/mmol)** |  |  |  |
| mean (SD) | 25.05 (15.66) | 28.65 (15.97) | 24.66 (12.3) |
| median [range] | 22.29 [5.403, 456.9] | 24.39 [5.173, 117.4] | 22.11 [4.956, 130.5] |
| missing | 1072 (33%) | 58 (2.1%) | 17 (0.8%) |
| **arabinose (μmol/mmol)** |  |  |  |
| mean (SD) | 19.64 (9.742) | 27.16 (15.49) | 22.43 (11.6) |
| median [range] | 17.63 [4.044, 160.4] | 22.61 [6.369, 114.3] | 19.67 [4.636, 108.5] |
| missing | 160 (4.9%) | 115 (4.2%) | 89 (4.4%) |
| **cis-aconitate (μmol/mmol)** |  |  |  |
| mean (SD) | 20.9 (31.18) | 30.46 (10.88) | 27.93 (9.8) |
| median [range] | 14.67 [2.294, 1179] | 29.72 [2.197, 121.1] | 26.66 [1.77, 85.98] |
| missing | 240 (7.4%) | 283 (10.3%) | 169 (8.3%) |
| **citrate (μmol/mmol)** |  |  |  |
| mean (SD) | 315.4 (162.2) | 247.3 (110.86) | 202.6 (100.05) |
| median [range] | 300.1 [0.7294, 1170] | 234 [0, 817.8] | 189.1 [0, 744.4] |
| missing | 108 (3.3%) | 103 (3.8%) | 27 (1.3%) |
| **creatine (μmol/mmol)** |  |  |  |
| mean (SD) | 130.7 (102.7) | 82.96 (111.74) | 105.3 (137.55) |
| median [range] | 101.3 [0.01846, 897.1] | 41.85 [0, 764.7] | 70.07 [0, 1070] |
| missing | 1610 (49.6%) | 2150 (78.4%) | 1608 (79.3%) |
| **creatinine (mmol/l)** |  |  |  |
| mean (SD) | 4.958 (2.82) | 8.906 (5.79) | 10.6 (6.11) |
| median [range] | 4.337 [0.2265, 31.26] | 7.61 [1.092, 41.02] | 9.161 [1.112, 54.01] |
| missing | 0 (0%) | 0 (0%) | 0 (0%) |
| **dimethylamine (μmol/mmol)** |  |  |  |
| mean (SD) | 32.06 (11.23) | 21.68 (9.19) | 26.36 (14.45) |
| median [range] | 30.14 [0.1886, 186.4] | 21.34 [0, 141.6] | 23.19 [0, 251.6] |
| missing | 13 (0.4%) | 0 (0%) | 0 (0%) |
| **ethanol (μmol/mmol)** |  |  |  |
| mean (SD) | 216.9 (723.9) | 24.54 (113.1) | 38.37 (249.04) |
| median [range] | 34.16 [1.461, 11010] | 12.43 [1.765, 3047] | 10.78 [1.128, 5071] |
| missing | 599 (18.5%) | 0 (0%) | 0 (0%) |
| **ethanolamine (μmol/mmol)** |  |  |  |
| mean (SD) | 40.98 (9.98) | 36.65 (10.07) | 32.7 (9.14) |
| median [range] | 39.95 [15.04, 108.6] | 35.54 [11.23, 83.46] | 31.92 [9.404, 84.59] |
| missing | 874 (26.9%) | 100 (3.6%) | 152 (7.5%) |
| **formate (μmol/mmol)** |  |  |  |
| mean (SD) | 75.98 (98.71) | 26.24 (13.68) | 25.27 (17.48) |
| median [range] | 52.74 [0.2438, 1210] | 23.63 [0, 127.6] | 22.14 [0, 295.2] |
| missing | 5 (0.2%) | 7 (0.3%) | 1 (0%) |
| **glucose (μmol/mmol)** |  |  |  |
| mean (SD) | 10340 (13820) | 120.6 (1771.38) | 49.05 (1100.87) |
| median [range] | 5176 [0.1887, 223700] | 0 [0, 50850] | 0 [0, 44670] |
| missing | 191 (5.9%) | 21 (0.8%) | 90 (4.4%) |
| **glutamine (μmol/mmol)** |  |  |  |
| mean (SD) | 54.47 (21.55) | 55.28 (20.91) | 50.26 (17.92) |
| median [range] | 51.65 [11.35, 277.3] | 51.35 [12.65, 170.7] | 46.84 [16.77, 167.6] |
| missing | 90 (2.8%) | 764 (27.9%) | 567 (28%) |
| **glycine (μmol/mmol)** |  |  |  |
| mean (SD) | 187.1 (159.8) | 82.73 (66.45) | 86.45 (70.77) |
| median [range] | 141.8 [0.2982, 1794] | 66.24 [0, 672.6] | 67.81 [0, 839.2] |
| missing | 205 (6.3%) | 7 (0.3%) | 4 (0.2%) |
| **glycolic acid (μmol/mmol)** |  |  |  |
| mean (SD) | 55.78 (26.54) | 44.82 (18) | 44.48 (19.37) |
| median [range] | 47.38 [13.36, 513] | 41.84 [11.16, 504.1] | 41.28 [13.37, 556.8] |
| missing | 1198 (36.9%) | 370 (13.5%) | 185 (9.1%) |
| **hippurate (μmol/mmol)** |  |  |  |
| mean (SD) | 506.5 (320.2) | 157.9 (140.15) | 165.2 (140.48) |
| median [range] | 450.8 [3.014, 2367] | 125.5 [0, 2522] | 136.2 [0, 3292] |
| missing | 0 (0%) | 1 (0%) | 2 (0.1%) |
| **histidine (μmol/mmol)** |  |  |  |
| mean (SD) | 102.4 (49.55) | 46.31 (27.47) | 47.32 (25.27) |
| median [range] | 93.87 [15.57, 469.5] | 41.62 [1.999, 560.1] | 42.83 [0, 262.6] |
| missing | 1264 (39%) | 1116 (40.7%) | 664 (32.7%) |
| **hypoxanthine (μmol/mmol)** |  |  |  |
| mean (SD) | 6.634 (4.587) | 9.97 (5.93) | 6.744 (3.75) |
| median [range] | 5.569 [1.114, 110.8] | 8.919 [1.741, 96.43] | 5.737 [1.965, 40.27] |
| missing | 388 (12%) | 120 (4.4%) | 102 (5%) |
| **indoxyl sulfate (μmol/mmol)** |  |  |  |
| mean (SD) | 26.02 (17.03) | 22.2 (14.58) | 22.73 (13.12) |
| median [range] | 22.79 [0.01633, 203.3] | 20.07 [0, 100.8] | 20.84 [0, 186.1] |
| missing | 387 (11.9%) | 48 (1.7%) | 13 (0.6%) |
| **isoleucine (μmol/mmol)** |  |  |  |
| mean (SD) | 2.862 (1.992) | 3.382 (2.23) | 2.775 (1.82) |
| median [range] | 2.386 [0.6895, 33.16] | 2.697 [0.696, 15.39] | 2.299 [0.764, 17.22] |
| missing | 732 (22.6%) | 1449 (52.8%) | 933 (46%) |
| **lactate (μmol/mmol)** |  |  |  |
| mean (SD) | 87.59 (202.2) | 6.996 (21.7) | 5.462 (7.5) |
| median [range] | 34.53 [0.01933, 3366] | 3.508 [0, 749.1] | 3.558 [0, 94.12] |
| missing | 41 (1.3%) | 58 (2.1%) | 62 (3.1%) |
| **leucine (μmol/mmol)** |  |  |  |
| mean (SD) | 5.428 (2.945) | 4.104 (1.91) | 3.592 (1.53) |
| median [range] | 4.788 [0.8384, 33.63] | 3.614 [1.125, 18.6] | 3.269 [0.974, 13.84] |
| missing | 20 (0.6%) | 86 (3.1%) | 75 (3.7%) |
| **mannitol (μmol/mmol)** |  |  |  |
| mean (SD) | 101.9 (106.4) | 64.88 (115.13) | 56.59 (98.12) |
| median [range] | 63.37 [6.046, 668.8] | 37.72 [6.374, 3406] | 30.78 [5.191, 2255] |
| missing | 2590 (79.8%) | 217 (7.9%) | 223 (11%) |
| **proline betaine (μmol/mmol)** |  |  |  |
| mean (SD) | 78.53 (84.72) | 55.08 (43.37) | 38.86 (32.74) |
| median [range] | 48.55 [5.443, 1269] | 43.11 [5.306, 415.5] | 29.34 [5.758, 486.7] |
| missing | 625 (19.3%) | 712 (26%) | 440 (21.7%) |
| **propylene glycol (μmol/mmol)** |  |  |  |
| mean (SD) | 10.51 (8.637) | 1.987 (7.1) | 1.951 (4.69) |
| median [range] | 8.093 [0.007594, 65.69] | 0 [0, 166.6] | 0.069 [0, 55.56] |
| missing | 1321 (40.7%) | 914 (33.3%) | 739 (36.4%) |
| **pseudouridine (μmol/mmol)** |  |  |  |
| mean (SD) | 36.73 (7.793) | 30.83 (4.86) | 30.62 (4.96) |
| median [range] | 36 [8.987, 96.65] | 30.44 [15.57, 85.34] | 30.3 [9.586, 85.88] |
| missing | 6 (0.2%) | 7 (0.3%) | 5 (0.2%) |
| **pyroglutamate (μmol/mmol)** |  |  |  |
| mean (SD) | 32.64 (9.67) | 22.9 (9.03) | 19.08 (5.3) |
| median [range] | 31.56 [8.874, 120.4] | 21.04 [5.89, 115.9] | 18.25 [8.377, 69.32] |
| missing | 163 (5%) | 35 (1.3%) | 8 (0.4%) |
| **quinic acid (μmol/mmol)** |  |  |  |
| mean (SD) | 36.23 (23.32) | 15.76 (15.91) | 11.24 (12.18) |
| median [range] | 33.31 [1.969, 167.7] | 9.931 [1.146, 125.1] | 7.12 [0.666, 161.9] |
| missing | 51 (1.6%) | 186 (6.8%) | 117 (5.8%) |
| **sucrose (μmol/mmol)** |  |  |  |
| mean (SD) | 9.103 (18.5) | 6.154 (9.13) | 6.492 (11.91) |
| median [range] | 3.871 [0.3521, 344.6] | 3.731 [0.293, 184.9] | 3.868 [0.352, 282.5] |
| missing | 32 (1%) | 94 (3.4%) | 14 (0.7%) |
| **taurine (μmol/mmol)** |  |  |  |
| mean (SD) | 116.1 (126.7) | 59.17 (30.12) | 51.45 (30.27) |
| median [range] | 61.48 [10.87, 641.8] | 54.72 [6.763, 396] | 44.24 [8.434, 392.8] |
| missing | 2733 (84.2%) | 866 (31.6%) | 721 (35.6%) |
| **threonine (μmol/mmol)** |  |  |  |
| mean (SD) | 16.37 (18.83) | 7.867 (4.63) | 8.16 (5.75) |
| median [range] | 11.63 [1.179, 369.5] | 6.912 [1.364, 83.37] | 6.882 [1.734, 88.05] |
| missing | 25 (0.8%) | 22 (0.8%) | 4 (0.2%) |
| **trans-aconitate (μmol/mmol)** |  |  |  |
| mean (SD) | 24.88 (11.67) | 4.992 (3.32) | 5.494 (3.8) |
| median [range] | 23.72 [3.062, 204] | 4.161 [0.915, 54.82] | 4.627 [0.78, 46.53] |
| missing | 33 (1%) | 59 (2.2%) | 89 (4.4%) |
| **trigonelline (μmol/mmol)** |  |  |  |
| mean (SD) | 142.6 (104.7) | 13 (19.83) | 11.03 (16.99) |
| median [range] | 130 [0.05629, 732] | 4.352 [0, 153.3] | 4.713 [0, 193.9] |
| missing | 0 (0%) | 0 (0%) | 0 (0%) |
| **trimethylamine-n-oxide (μmol/mmol)** |  |  |  |
| mean (SD) | 54.54 (82.22) | 41.25 (51.27) | 86.27 (141.66) |
| median [range] | 34.82 [0.009741, 1381] | 27.95 [0, 618.1] | 35.47 [0, 1315] |
| missing | 46 (1.4%) | 30 (1.1%) | 11 (0.5%) |
| **tryptophan (μmol/mmol)** |  |  |  |
| mean (SD) | 9.508 (5.158) | 8.529 (3.48) | 7.314 (2.7) |
| median [range] | 8.481 [2.158, 136.2] | 7.832 [2.25, 57.13] | 6.781 [1.937, 29.16] |
| missing | 175 (5.4%) | 65 (2.4%) | 55 (2.7%) |
| **tyrosine (μmol/mmol)** |  |  |  |
| mean (SD) | 14.71 (6.785) | 9.785 (4.78) | 8.19 (3.6) |
| median [range] | 13.42 [2.359, 66.73] | 9.022 [2.615, 114.8] | 7.575 [2.411, 64.62] |
| missing | 15 (0.5%) | 118 (4.3%) | 45 (2.2%) |
| **uracil (μmol/mmol)** |  |  |  |
| mean (SD) | 0.9429 (0.7207) | 0.94 (0.92) | 0.772 (0.46) |
| median [range] | 0.8851 [0.211, 24.37] | 0.796 [0.171, 22.41] | 0.685 [0.157, 9.922] |
| missing | 0 (0%) | 1 (0%) | 0 (0%) |
| **urea (μmol/mmol)** |  |  |  |
| mean (SD) | 39370 (12560) | 27550 (9445.44) | 27640 (10434.96) |
| median [range] | 38010 [7733, 245200] | 26450 [2222, 84460] | 26880 [5325, 134600] |
| missing | 0 (0%) | 0 (0%) | 0 (0%) |
| **valine (μmol/mmol)** |  |  |  |
| mean (SD) | 6.874 (4.604) | 2.925 (1.08) | 2.75 (0.95) |
| median [range] | 6.003 [0.4362, 65.96] | 2.781 [0.465, 19.87] | 2.609 [0.412, 12.06] |
| missing | 52 (1.6%) | 85 (3.1%) | 72 (3.6%) |
| **xanthosine (μmol/mmol)** |  |  |  |
| mean (SD) | 11.84 (2.966) | 14.84 (4.65) | 14.66 (5.38) |
| median [range] | 11.15 [4.496, 40.27] | 14.05 [3.539, 44.4] | 13.64 [4.721, 66.46] |
| missing | 0 (0%) | 0 (0%) | 0 (0%) |
| **xylose (μmol/mmol)** |  |  |  |
| mean (SD) | 14.51 (18.18) | 12 (15.09) | 10.79 (13.16) |
| median [range] | 8.028 [1.13, 365.3] | 7.98 [1.551, 266.9] | 7.558 [1.691, 188.9] |
| missing | 548 (16.9%) | 115 (4.2%) | 78 (3.8%) |

### Supplementary Table 3

**Urinary metabolite names and database ids.**

| **Metabolite** | **Metabolite short name** | **HMDB ID** | **CAS Registry Number** |
| --- | --- | --- | --- |
| 1-methylnicotinamide | omna | HMDB0000699 | 3106-60-3 |
| 2-furoylglycine | furgly | HMDB0000439 | 5657-19-2 |
| 2-hydroxyisobutyrate | aohibut | HMDB0000729 | 594-61-6 |
| 3-(3-hydroxyphenyl)-3-hydroxypropionic acid | hphpa | HMDB0002643 | 3247-75-4 |
| 3-aminoisobutyrate | bnhibut | HMDB0002166 | 4249-19-8 |
| 3-hydroxyhippurate | mohhip | HMDB0006116 | 1637-75-8 |
| 3-hydroxyisobutyrate | bohibut | - | 2068-83-9 |
| 3-hydroxyisovalerate | bohival | HMDB0000754 | 625-08-1 |
| 3-methylhistidine | tmehis | HMDB0000479 | 368-16-1 |
| 4-deoxyerythronic acid | doeta | HMDB0000498 | 759-06-8 |
| 4-deoxythreonate | dta | HMDB0002453 | 5057-93-2 |
| 4-hydroxyhippurate | pohhip | HMDB0013678 | 2482-25-9 |
| acetate | ace | HMDB0000042 | 64-19-7 |
| alanine | ala | HMDB0000161 | 56-41-7 |
| allantoin | aln | HMDB0000462 | 97-59-6 |
| arabinose | arb | - | 147-81-9 |
| cis-aconitate | caco | HMDB0000072 | 585-84-2 |
| citrate | cit | HMDB0000094 | 77-92-9 |
| creatine | cr | HMDB0000064 | 57-00-1 |
| creatinine | crea | HMDB0000562 | 60-27-5 |
| dimethylamine | dma | HMDB0000087 | 124-40-3 |
| ethanol | etoh | HMDB0000108 | 64-17-5 |
| ethanolamine | etnh | HMDB0000149 | 141-43-5 |
| formate | form | HMDB0000142 | 64-18-6 |
| glucose | glc | HMDB0000122 | 50-99-7 |
| glutamine | gln | HMDB0000641 | 56-85-9 |
| glycine | gly | HMDB0000123 | 56-40-6 |
| glycolic acid | glya | HMDB0000115 | 79-14-1 |
| hippurate | hip | HMDB0000714 | 495-69-2 |
| histidine | his | HMDB0000177 | 71-00-1 |
| hypoxanthine | hyp | HMDB0000157 | 68-94-0 |
| indoxyl sulfate | ind | HMDB0000682 | 487-94-5 |
| isoleucine | ile | HMDB0000172 | 73-32-5 |
| lactate | lac | HMDB0000190 | 79-33-4 |
| leucine | leu | HMDB0000687 | 61-90-5 |
| mannitol | mnt | HMDB0000765 | 69-65-8 |
| proline betaine | probet | HMDB0004827 | 471-87-4 |
| propylene glycol | prgly | HMDB0001881 | 4254-14-2 |
| pseudouridine | pseur | HMDB0000767 | 1445-07-4 |
| pyroglutamate | pglu | HMDB0000267 | 98-79-3 |
| quinic acid | quina | HMDB0003072 | 77-95-2 |
| sucrose | scr | HMDB0000258 | 57-50-1 |
| taurine | tau | HMDB0000251 | 107-35-7 |
| threonine | thre | HMDB0000167 | 72-19-5 |
| trans-aconitate | taco | HMDB0000958 | 4023-65-8 |
| trigonelline | trig | HMDB0000875 | 535-83-1 |
| trimethylamine-n-oxide | tmao | HMDB0000925 | 1184-78-7 |
| tryptophan | trp | HMDB0000929 | 73-22-3 |
| tyrosine | tyr | HMDB0000158 | 60-18-4 |
| uracil | ura | HMDB0000300 | 66-22-8 |
| urea | ure | HMDB0000294 | 57-13-6 |
| valine | val | HMDB0000883 | 72-18-4 |
| xanthosine | xan | HMDB0000299 | 146-80-5 |
| xylose | xyl | HMDB0000098 | 58-86-6 |

### **Supplementary Table 4**

**Heritability estimates for the urinary metabolites.**

|  | **FinnDiane** | | **GS** | | **VIKING** | | **Meta-analysis** | | |
| --- | --- | --- | --- | --- | --- | --- | --- | --- | --- |
| **Metabolite** | **Heritability (SE)** | **P-value** | **Heritability (SE)** | **P-value** | **Heritability (SE)** | **P-value** | **Heritability (SE)** | **P-value** | **Heterogeneity p-value** |
| 1-Methylnicotinamide | 0.00 (0.10) | 5.0×10^-01^ | 0.18 (0.11) | 5.0×10^-02^ | 0.08 (0.04) | 1.9×10^-02^ | 0.08 (0.03) | 1.7×10^-02^ | 4.9×10^-01^ |
| 2-Furoylglycine | 0.00 (0.10) | 5.0×10^-01^ | 0.08 (0.07) | 1.3×10^-01^ | 0.00 (0.00) | 2.5×10^-01^ | 0.00 (0.00) | 5.0×10^-01^ | 5.4×10^-01^ |
| 2-Hydroxyisobutyrate | 0.05 (0.10) | 3.2×10^-01^ | 0.13 (0.09) | 8.6×10^-02^ | 0.32 (0.04) | 1.9×10^-14^ | 0.19 (0.09) | 3.0×10^-02^ | 1.9×10^-02^ |
| 3-(3-Hydroxyphenyl)-3-hydroxypropionic acid | 0.00 (0.11) | 5.0×10^-01^ | 0.09 (0.08) | 1.4×10^-01^ | 0.01 (0.01) | 2.3×10^-01^ | 0.01 (0.01) | 3.8×10^-01^ | 6.1×10^-01^ |
| 3-Aminoisobutyrate | 0.38 (0.15) | 4.2×10^-03^ | 0.33 (0.14) | 7.9×10^-03^ | 0.32 (0.06) | 4.4×10^-09^ | 0.33 (0.05) | 1.7×10^-11^ | 9.3×10^-01^ |
| 3-hydroxyhippurate | 0.00 (0.10) | 5.0×10^-01^ | 0.17 (0.11) | 6.7×10^-02^ | 0.14 (0.05) | 3.4×10^-03^ | 0.12 (0.04) | 5.0×10^-03^ | 4.3×10^-01^ |
| 3-Hydroxyisobutyrate | 0.13 (0.10) | 1.0×10^-01^ | 0.29 (0.13) | 1.1×10^-02^ | 0.39 (0.05) | 2.7×10^-18^ | 0.29 (0.08) | 7.4×10^-04^ | 5.2×10^-02^ |
| 3-Hydroxyisovalerate | 0.14 (0.10) | 8.7×10^-02^ | 0.30 (0.12) | 5.6×10^-03^ | 0.37 (0.05) | 9.1×10^-16^ | 0.29 (0.07) | 9.6×10^-05^ | 1.0×10^-01^ |
| 3-Methylhistidine | 0.00 (0.15) | 5.0×10^-01^ | 0.29 (0.19) | 6.9×10^-02^ | 0.07 (0.04) | 6.7×10^-02^ | 0.07 (0.04) | 8.6×10^-02^ | 4.7×10^-01^ |
| 4-Deoxyerythronic acid | 0.01 (0.11) | 4.5×10^-01^ | 0.37 (0.13) | 2.6×10^-03^ | 0.24 (0.05) | 2.5×10^-07^ | 0.21 (0.09) | 2.3×10^-02^ | 7.7×10^-02^ |
| 4-Deoxythreonate | 0.16 (0.11) | 6.6×10^-02^ | 0.15 (0.11) | 7.9×10^-02^ | 0.25 (0.05) | 5.6×10^-07^ | 0.22 (0.04) | 2.3×10^-07^ | 5.8×10^-01^ |
| 4-Hydroxyhippurate | 0.13 (0.10) | 1.1×10^-01^ | 0.21 (0.12) | 3.2×10^-02^ | 0.03 (0.03) | 1.3×10^-01^ | 0.08 (0.05) | 1.3×10^-01^ | 2.3×10^-01^ |
| Acetate | 0.00 (0.10) | 5.0×10^-01^ | 0.16 (0.10) | 5.9×10^-02^ | 0.09 (0.04) | 7.9×10^-03^ | 0.09 (0.03) | 8.0×10^-03^ | 5.4×10^-01^ |
| Alanine | 0.01 (0.10) | 4.8×10^-01^ | 0.16 (0.11) | 6.8×10^-02^ | 0.30 (0.04) | 3.6×10^-12^ | 0.17 (0.09) | 6.0×10^-02^ | 1.5×10^-02^ |
| Allantoin | 0.22 (0.14) | 4.9×10^-02^ | 0.00 (0.00) | 2.5×10^-01^ | 0.07 (0.04) | 4.1×10^-02^ | 0.04 (0.04) | 3.1×10^-01^ | 6.3×10^-02^ |
| Arabinose | 0.20 (0.11) | 3.1×10^-02^ | 0.00 (0.00) | 2.5×10^-01^ | 0.10 (0.04) | 1.3×10^-02^ | 0.07 (0.05) | 1.9×10^-01^ | 1.7×10^-02^ |
| cis-Aconitate | 0.00 (0.10) | 5.0×10^-01^ | 0.00 (0.00) | 2.5×10^-01^ | 0.19 (0.05) | 2.2×10^-05^ | 0.07 (0.07) | 3.2×10^-01^ | 2.3×10^-04^ |
| Citrate | 0.26 (0.11) | 6.0×10^-03^ | 0.34 (0.13) | 3.7×10^-03^ | 0.39 (0.04) | 4.7×10^-18^ | 0.36 (0.04) | 2.3×10^-20^ | 5.3×10^-01^ |
| Creatine | 0.00 (0.19) | 5.0×10^-01^ | 0.00 (0.00) | 2.5×10^-01^ | 0.27 (0.15) | 3.9×10^-02^ | 0.05 (0.08) | 5.2×10^-01^ | 2.1×10^-01^ |
| Creatinine | 0.01 (0.10) | 4.6×10^-01^ | 0.00 (0.00) | 2.5×10^-01^ | 0.11 (0.04) | 4.7×10^-03^ | 0.04 (0.04) | 3.5×10^-01^ | 3.4×10^-02^ |
| Dimethylamine | 0.09 (0.10) | 1.9×10^-01^ | 0.00 (0.00) | 2.5×10^-01^ | 0.03 (0.03) | 9.9×10^-02^ | 0.01 (0.01) | 5.3×10^-01^ | 3.0×10^-01^ |
| Ethanol | 0.00 (0.12) | 5.0×10^-01^ | 0.21 (0.11) | 2.9×10^-02^ | 0.00 (0.00) | 2.5×10^-01^ | 0.04 (0.06) | 4.9×10^-01^ | 1.7×10^-01^ |
| Ethanolamine | 0.00 (0.13) | 5.0×10^-01^ | 0.00 (0.00) | 2.5×10^-01^ | 0.16 (0.05) | 1.1×10^-03^ | 0.06 (0.06) | 3.4×10^-01^ | 9.0×10^-03^ |
| Formate | 0.02 (0.10) | 4.0×10^-01^ | 0.06 (0.06) | 1.6×10^-01^ | 0.20 (0.04) | 2.8×10^-06^ | 0.12 (0.06) | 4.6×10^-02^ | 9.0×10^-02^ |
| Glucose | 0.00 (0.10) | 5.0×10^-01^ | 0.00 (0.00) | 2.5×10^-01^ | 0.07 (0.05) | 7.4×10^-02^ | 0.01 (0.02) | 5.9×10^-01^ | 3.5×10^-01^ |
| Glutamine | 0.21 (0.10) | 1.4×10^-02^ | 0.16 (0.13) | 1.1×10^-01^ | 0.14 (0.06) | 6.3×10^-03^ | 0.16 (0.05) | 6.5×10^-04^ | 8.4×10^-01^ |
| Glycine | 0.22 (0.11) | 1.6×10^-02^ | 0.08 (0.07) | 1.3×10^-01^ | 0.35 (0.05) | 1.4×10^-13^ | 0.23 (0.09) | 8.6×10^-03^ | 8.7×10^-03^ |
| Glycolic acid | 0.00 (0.15) | 5.0×10^-01^ | 0.17 (0.11) | 6.0×10^-02^ | 0.23 (0.05) | 4.4×10^-07^ | 0.20 (0.04) | 7.8×10^-06^ | 3.3×10^-01^ |
| Hippurate | 0.00 (0.10) | 5.0×10^-01^ | 0.35 (0.12) | 1.8×10^-03^ | 0.20 (0.04) | 4.6×10^-07^ | 0.18 (0.09) | 4.1×10^-02^ | 6.3×10^-02^ |
| Histidine | 0.10 (0.16) | 2.8×10^-01^ | 0.00 (0.00) | 2.5×10^-01^ | 0.47 (0.07) | 1.3×10^-11^ | 0.19 (0.15) | 2.1×10^-01^ | 1.8×10^-10^ |
| Hypoxanthine | 0.01 (0.12) | 4.6×10^-01^ | 0.00 (0.00) | 2.5×10^-01^ | 0.11 (0.04) | 2.8×10^-03^ | 0.04 (0.04) | 3.4×10^-01^ | 2.2×10^-02^ |
| Indoxyl Sulfate | 0.00 (0.11) | 5.0×10^-01^ | 0.00 (0.00) | 2.5×10^-01^ | 0.19 (0.04) | 1.6×10^-05^ | 0.07 (0.07) | 3.2×10^-01^ | 1.8×10^-04^ |
| Isoleucine | 0.24 (0.13) | 3.3×10^-02^ | 0.00 (0.00) | 2.5×10^-01^ | 0.11 (0.06) | 4.6×10^-02^ | 0.07 (0.06) | 2.4×10^-01^ | 4.6×10^-02^ |
| Lactate | 0.00 (0.11) | 5.0×10^-01^ | 0.00 (0.00) | 2.5×10^-01^ | 0.20 (0.05) | 2.4×10^-05^ | 0.07 (0.07) | 3.2×10^-01^ | 2.6×10^-04^ |
| Leucine | 0.06 (0.10) | 2.7×10^-01^ | 0.06 (0.06) | 1.7×10^-01^ | 0.15 (0.04) | 3.6×10^-04^ | 0.11 (0.04) | 1.6×10^-03^ | 4.6×10^-01^ |
| Mannitol | 0.22 (0.45) | 3.2×10^-01^ | 0.02 (0.03) | 2.2×10^-01^ | 0.04 (0.03) | 1.1×10^-01^ | 0.03 (0.02) | 1.5×10^-01^ | 8.7×10^-01^ |
| Proline betaine | 0.28 (0.12) | 5.4×10^-03^ | 0.06 (0.07) | 1.9×10^-01^ | 0.09 (0.05) | 3.1×10^-02^ | 0.10 (0.04) | 6.9×10^-03^ | 2.7×10^-01^ |
| Propylene Glycol | 0.00 (0.15) | 5.0×10^-01^ | 0.00 (0.00) | 2.5×10^-01^ | 0.10 (0.06) | 3.5×10^-02^ | 0.03 (0.04) | 4.7×10^-01^ | 1.9×10^-01^ |
| Pseudouridine | 0.15 (0.10) | 6.0×10^-02^ | 0.11 (0.09) | 1.0×10^-01^ | 0.27 (0.05) | 2.0×10^-09^ | 0.20 (0.06) | 5.8×10^-04^ | 1.8×10^-01^ |
| Pyroglutamate | 0.07 (0.11) | 2.7×10^-01^ | 0.03 (0.04) | 2.1×10^-01^ | 0.10 (0.04) | 1.0×10^-02^ | 0.06 (0.03) | 3.5×10^-02^ | 4.9×10^-01^ |
| Quinic acid | 0.00 (0.10) | 5.0×10^-01^ | 0.09 (0.08) | 1.3×10^-01^ | 0.13 (0.04) | 1.9×10^-03^ | 0.10 (0.04) | 4.0×10^-03^ | 5.0×10^-01^ |
| Sucrose | 0.32 (0.11) | 1.0×10^-03^ | 0.20 (0.11) | 3.2×10^-02^ | 0.11 (0.04) | 3.3×10^-03^ | 0.18 (0.07) | 6.3×10^-03^ | 1.4×10^-01^ |
| Taurine | 0.67 (0.55) | 1.1×10^-01^ | 0.00 (0.00) | 2.5×10^-01^ | 0.08 (0.05) | 6.8×10^-02^ | 0.03 (0.04) | 4.8×10^-01^ | 1.5×10^-01^ |
| Threonine | 0.22 (0.10) | 1.6×10^-02^ | 0.15 (0.10) | 7.7×10^-02^ | 0.30 (0.05) | 2.5×10^-10^ | 0.26 (0.05) | 8.0×10^-08^ | 3.7×10^-01^ |
| trans-Aconitate | 0.08 (0.10) | 2.1×10^-01^ | 0.01 (0.01) | 2.4×10^-01^ | 0.13 (0.05) | 3.3×10^-03^ | 0.06 (0.05) | 1.9×10^-01^ | 4.1×10^-02^ |
| Trigonelline | 0.05 (0.10) | 3.1×10^-01^ | 0.11 (0.09) | 9.6×10^-02^ | 0.07 (0.04) | 2.7×10^-02^ | 0.08 (0.03) | 2.1×10^-02^ | 8.7×10^-01^ |
| Trimethylamine-N-oxide | 0.21 (0.10) | 2.0×10^-02^ | 0.00 (0.00) | 2.5×10^-01^ | 0.11 (0.04) | 1.2×10^-03^ | 0.08 (0.05) | 1.5×10^-01^ | 1.4×10^-03^ |
| Tryptophan | 0.26 (0.11) | 4.7×10^-03^ | 0.00 (0.00) | 2.5×10^-01^ | 0.18 (0.05) | 3.6×10^-05^ | 0.13 (0.08) | 1.1×10^-01^ | 1.8×10^-05^ |
| Tyrosine | 0.30 (0.10) | 1.8×10^-03^ | 0.15 (0.11) | 7.8×10^-02^ | 0.31 (0.05) | 1.6×10^-10^ | 0.29 (0.04) | 4.9×10^-12^ | 3.7×10^-01^ |
| Uracil | 0.16 (0.10) | 5.9×10^-02^ | 0.25 (0.11) | 1.5×10^-02^ | 0.14 (0.04) | 4.4×10^-04^ | 0.15 (0.04) | 3.0×10^-05^ | 6.8×10^-01^ |
| Urea | 0.17 (0.10) | 4.5×10^-02^ | 0.21 (0.12) | 3.3×10^-02^ | 0.19 (0.05) | 3.7×10^-05^ | 0.19 (0.04) | 3.1×10^-06^ | 9.7×10^-01^ |
| Valine | 0.03 (0.10) | 3.8×10^-01^ | 0.05 (0.06) | 1.8×10^-01^ | 0.34 (0.05) | 5.5×10^-13^ | 0.15 (0.10) | 1.5×10^-01^ | 1.0×10^-04^ |
| Xanthosine | 0.29 (0.11) | 3.2×10^-03^ | 0.28 (0.12) | 1.1×10^-02^ | 0.16 (0.04) | 1.6×10^-04^ | 0.20 (0.05) | 3.8×10^-05^ | 3.8×10^-01^ |
| Xylose | 0.06 (0.12) | 3.0×10^-01^ | 0.04 (0.05) | 1.9×10^-01^ | 0.19 (0.05) | 2.9×10^-05^ | 0.11 (0.06) | 6.3×10^-02^ | 7.3×10^-02^ |

### **Supplementary Table 5**

**Genome-wide significant associations with urinary metabolites.** Associations (n=52) of variants with metabolites with p < 9.3×10^-10^ from the COJO analysis. Previously reported associations fetched from the GWAS catalog (window size=-/+500kb, r^2^ > 0.8, and p < 5×10^-8^).

| **CHR:POS:EA:NEA** | **Rsid** | **Gene** | **Variant type** | **Metabolite** | **EAF** | **Beta (SE)** | **P** | **N** | **Prev. Assoc.** | **Novel Assoc.** |
| --- | --- | --- | --- | --- | --- | --- | --- | --- | --- | --- |
| 1:6334301:A:G | rs114200864 | *ACOT7* | intron | 3-Hydroxyisovalerate | 0.041 | -0.28 (0.05) | 2.9×10^-10^ | 6862 | No | Yes |
| 1:11940483:T:C | rs4846068 | *SBF1P2* / 1p36.22 | downstream (0.02kb) | Formate | 0.585 | 0.13 (0.02) | 2.2×10^-15^ | 8193 | Yes | Yes |
| 1:48690229:A:C | rs10788884 | *SLC5A9* | intron | Xylose | 0.674 | 0.11 (0.02) | 1.1×10^-10^ | 7696 | Yes | Yes |
| 1:151904146:A:T | rs2999545 | *KRT8P28* / 1q21.3 | downstream (20kb) | 3-Hydroxyisovalerate | 0.385 | -0.11 (0.02) | 4.7×10^-12^ | 8464 | Yes | No |
| 2:135598913:A:G | rs17322446 | *ACMSD, CCNT2-AS1* | intron | 1-Methylnicotinamide | 0.183 | -0.13 (0.02) | 6.6×10^-10^ | 7619 | Yes | No |
| 2:211540507:A:C | rs1047891 | *CPS1* | missense | Glycine | 0.317 | 0.23 (0.02) | 2.9×10^-37^ | 7548 | Yes | No |
| 2:241793545:A:G | rs55649245 | *AGXT* / 2q37.3 | upstream (10kb) | 4-Deoxyerythronic acid | 0.326 | -0.17 (0.02) | 3.2×10^-20^ | 8058 | Yes | No |
| 2:241813788:T:C | rs10933641 | *AGXT* | intron | 4-Deoxyerythronic acid | 0.277 | 0.19 (0.02) | 2.6×10^-20^ | 7714 | Yes | Yes* |
| 3:182758040:T:C | rs4859267 | *MCCC1* | intron | 3-Hydroxyisovalerate | 0.294 | 0.13 (0.02) | 1.3×10^-12^ | 8276 | Yes | Yes |
| 4:88213884:T:C | rs6811902 | *MIR5705* / 4q22.1 | downstream (8kb) | Propylene Glycol | 0.601 | -0.16 (0.02) | 2.8×10^-20^ | 5357 | Yes | Yes |
| 4:109716840:A:T | rs62313082 | *RCC2P8* / 4q25 | upstream (6kb) | Ethanolamine | 0.379 | 0.14 (0.02) | 7.7×10^-16^ | 7164 | No | Yes |
| 5:1188285:A:G | rs11133665 | *TERLR1* / 5p15.33 | upstream (10kb) | Glycine | 0.261 | -0.13 (0.02) | 1.6×10^-13^ | 8232 | Yes | Yes |
| 5:1188285:A:G | rs11133665 | *TERLR1* / 5p15.33 | upstream (10kb) | Histidine | 0.255 | -0.31 (0.02) | 1.9×10^-39^ | 5257 | Yes | No |
| 5:1188285:A:G | rs11133665 | *TERLR1* / 5p15.33 | upstream (10kb) | Threonine | 0.259 | -0.15 (0.02) | 1.3×10^-16^ | 8417 | Yes | Yes |
| 5:1188285:A:G | rs11133665 | *TERLR1* / 5p15.33 | upstream (10kb) | Tryptophan | 0.258 | -0.17 (0.02) | 1.4×10^-19^ | 8133 | Yes | No |
| 5:1188285:A:G | rs11133665 | *TERLR1* / 5p15.33 | upstream (10kb) | Tyrosine | 0.259 | -0.30 (0.02) | 7.5×10^-55^ | 8234 | Yes | No |
| 5:1225434:T:C | rs7704882 | *SLC6A18* / 5p15.33 | upstream (0.06kb) | Tyrosine | 0.794 | -0.20 (0.02) | 4.6×10^-24^ | 8344 | No | Yes* |
| 5:1225613:A:G | rs7704058 | *SLC6A18* | synonymous | Tryptophan | 0.796 | -0.15 (0.02) | 5.1×10^-14^ | 8213 | No | Yes* |
| 5:34584621:A:C | rs16903139 | *RAI14-DT* / 5p13.2 | downstream (70kb) | 3-Aminoisobutyrate | 0.914 | 0.23 (0.03) | 5.0×10^-14^ | 6256 | No | Yes* |
| 5:34853162:T:C | rs338296 | *TTC23L* | intron | 3-Aminoisobutyrate | 0.428 | 0.12 (0.02) | 8.0×10^-11^ | 6368 | No | Yes* |
| 5:34868497:A:T | rs72732827 | *TTC23L* | 3′-UTR | 3-Aminoisobutyrate | 0.987 | -0.60 (0.08) | 1.2×10^-13^ | 5463 | No | Yes* |
| 5:34896132:A:G | rs138425947 | *TTC23L* | intron | 3-Aminoisobutyrate | 0.978 | -0.51 (0.07) | 5.4×10^-14^ | 4682 | No | Yes* |
| 5:34899723:T:C | rs56007938 | *TTC23L* | 3′-UTR | 3-Aminoisobutyrate | 0.015 | 0.66 (0.08) | 1.1×10^-17^ | 5419 | No | Yes* |
| 5:34911884:T:C | rs2308957 | *RAD1* | missense | 3-Aminoisobutyrate | 0.015 | -0.78 (0.09) | 3.2×10^-18^ | 4407 | No | Yes* |
| 5:34982167:T:C | rs116116288 | *AGXT2* / 5p13.2 | downstream (20kb) | 3-Aminoisobutyrate | 0.012 | 0.88 (0.09) | 8.2×10^-25^ | 5377 | No | Yes* |
| 5:34993215:A:G | rs11744796 | *AGXT2* / 5p13.2 | downstream (5kb) | 3-Aminoisobutyrate | 0.433 | -0.18 (0.02) | 9.5×10^-24^ | 6496 | Yes | No |
| 5:35000653:T:C | rs7737763 | *AGXT2* | intron | 3-Aminoisobutyrate | 0.434 | -0.44 (0.03) | 2.5×10^-67^ | 6312 | No | Yes* |
| 5:35003112:A:G | rs468327 | *AGXT2* | intron | 3-Aminoisobutyrate | 0.218 | 0.36 (0.03) | 1.3×10^-33^ | 5888 | Yes | No |
| 5:35037115:T:C | rs37369 | *AGXT2* | missense | 3-Aminoisobutyrate | 0.100 | 0.84 (0.04) | 4.3×10^-99^ | 5317 | Yes | No |
| 5:35039437:A:G | rs2279651 | *AGXT2* | synonymous | 3-Aminoisobutyrate | 0.431 | 0.45 (0.02) | 7.5×10^-82^ | 6419 | Yes | No |
| 5:35152241:T:C | rs954286 | *PRLR* | intron | 3-Aminoisobutyrate | 0.939 | 0.26 (0.04) | 2.1×10^-12^ | 6489 | No | Yes* |
| 5:150624099:T:C | rs72794144 | *GM2A* | intron | Glycine | 0.028 | 0.38 (0.06) | 2.5×10^-11^ | 7282 | No | Yes* |
| 5:150702299:A:G | rs61067578 | *SLC36A2* | intron | Glycine | 0.842 | 0.24 (0.02) | 1.1×10^-24^ | 8306 | Yes | Yes* |
| 5:150708711:C:G | rs147000073 | *SLC36A2* | intron | Glycine | 0.987 | -1.08 (0.11) | 1.7×10^-24^ | 3645 | Yes | No |
| 6:111492119:T:C | rs241768 | *SLC16A10* | intron | Tyrosine | 0.294 | 0.11 (0.02) | 2.6×10^-10^ | 8179 | Yes | No |
| 7:17287998:A:G | rs2106727 | *AHR* | intron | Quinic acid | 0.353 | -0.12 (0.02) | 3.1×10^-13^ | 8295 | Yes | Yes |
| 7:17287998:A:G | rs2106727 | *AHR* | intron | Trigonelline | 0.355 | -0.10 (0.02) | 6.5×10^-11^ | 8640 | Yes | Yes |
| 8:18272377:T:C | rs4921913 | *NAT2* / 8p22 | downstream (10kb) | Formate | 0.770 | -0.32 (0.02) | 9.3×10^-64^ | 8125 | Yes | No |
| 8:74868909:A:G | rs72661850 | *ELOC* | intron | 4-Deoxyerythronic acid | 0.691 | -0.12 (0.02) | 1.5×10^-11^ | 7976 | Yes | Yes |
| 9:6649491:T:C | rs62565993 | *GLDC* / 9p24.1 | upstream (4kb) | Glycine | 0.124 | 0.20 (0.03) | 3.4×10^-14^ | 7119 | No | Yes |
| 9:107525165:T:G | rs2472479 | *NIPSNAP3B* / 9q31.1 | upstream (1kb) | 3-Hydroxyisobutyrate | 0.565 | -0.16 (0.02) | 5.0×10^-22^ | 8108 | No | Yes |
| 9:136146597:T:C | rs550057 | *ABO* | intron | Xylose | 0.244 | 0.20 (0.02) | 8.9×10^-26^ | 7450 | Yes | Yes |
| 12:345369:C:G | rs2080403 | *SLC6A13* | intron | 3-Aminoisobutyrate | 0.454 | -0.16 (0.02) | 3.5×10^-22^ | 6425 | Yes | No |
| 12:4521511:A:T | rs78470967 | *FGF6* / 12p13.32 | downstream (20kb) | 4-Deoxythreonate | 0.043 | 0.34 (0.04) | 2.0×10^-14^ | 6377 | Yes | Yes |
| 12:122344302:A:G | rs1795967 | *PSMD9* | intron | 2-Hydroxyisobutyrate | 0.169 | -0.51 (0.02) | 6.5×10^-120^ | 7822 | Yes | No |
| 16:20557634:A:T | rs7499358 | *ACSM2B* | intron | 3-Hydroxyisovalerate | 0.982 | -0.62 (0.06) | 8.7×10^-22^ | 7387 | Yes | No |
| 16:20608891:C:G | rs540815683 | *ACSM5P1* | intron | 3-Hydroxyisovalerate | 0.986 | -0.81 (0.11) | 1.9×10^-13^ | 3282 | Yes | No |
| 17:26824156:A:G | rs11567842 | *SLC13A2* | missense | Citrate | 0.642 | -0.11 (0.02) | 3.0×10^-13^ | 8209 | Yes | Yes |
| 17:37631883:C:G | rs11078902 | *CDK12* | intron | Threonine | 0.241 | -0.12 (0.02) | 3.7×10^-10^ | 7570 | Yes | Yes |
| 17:37633970:A:C | rs12453397 | *CDK12* | intron | Tryptophan | 0.756 | 0.16 (0.02) | 7.2×10^-16^ | 7358 | Yes | No |
| 17:37633970:A:C | rs12453397 | *CDK12* | intron | Tyrosine | 0.758 | 0.25 (0.02) | 5.7×10^-36^ | 7351 | Yes | No |
| 17:37636695:T:G | rs4795371 | *CDK12* | intron | Histidine | 0.243 | -0.22 (0.02) | 5.1×10^-18^ | 4642 | Yes | No |

CHR:POS:EA:NEA: Chromosome position (GRCh37), effect allele, and non-effect allele. Rsid: variant rs-identifier. Gene: Closest gene. Variant type: consequence of the variant on the protein sequence. Closest genes and variant types found using Ensembl VEP (GRCh38 v110). Metabolite: the associated urinary metabolite. EAF: effect allele frequency. Beta (SE): effect estimate for the effect allele (effect estimate standard deviation). P: p-value of the association. N: number of individuals in the analysis. Prev. Assoc.: previous associations found in GWAS catalog. Novel Assoc.: the variant has not been reported before for the same urinary metabolite. *: novel independent signal in a previously reported locus for the same metabolite.

### Supplementary Table 6

**Previously reported traits for the lead variants from the GWAS catalog.** Previously associated traits in the GWAS catalog (window size=-/+500kb, r^2^ > 0.8, and p < 5e-08) for the lead variants (n=30) associated with urinary metabolites. The ‘Previously reported urinary/blood/other traits’ columns lists previously associated variants with the reported trait (p-value annotation). The previously reported traits are classified as urinary, blood or other by searching key words.

| **Chr:Pos** | **Rsid** | **Metabolite** | **Previously associated urinary metabolite traits** | **Previously associated blood metabolite traits** | **Previously associated other traits** |
| --- | --- | --- | --- | --- | --- |
| 1:11940483 | rs4846068 | Formate |  |  | [rs11588551](https://www.ebi.ac.uk/gwas/variants/rs11588551) Blood protein levels *(KIAA2013, 6538_90_3)* Serum levels of protein KIAA2013 [rs198384](https://www.ebi.ac.uk/gwas/variants/rs198384) Sex hormone-binding globulin levels Sex hormone-binding globulin levels adjusted for BMI |
| 1:48690229 | rs10788884 | Xylose | [rs10788884](https://www.ebi.ac.uk/gwas/variants/rs10788884) Urine mannose levels in chronic kidney disease | [rs926979](https://www.ebi.ac.uk/gwas/variants/rs926979) Serum uric acid levels | [rs926979](https://www.ebi.ac.uk/gwas/variants/rs926979) Urate levels |
| 1:151904146 | rs2999545 | 3-Hydroxyisovalerate | [rs7541453](https://www.ebi.ac.uk/gwas/variants/rs7541453) Urine X-24363 levels in chronic kidney disease | [rs1060870](https://www.ebi.ac.uk/gwas/variants/rs1060870) Serum metabolite levels *(3-hydroxylaurate)* Serum metabolite levels *(3-hydroxyoctanoate)* [rs10788817](https://www.ebi.ac.uk/gwas/variants/rs10788817) Serum metabolite levels *(3-hydroxydecanoate)* [rs2279501](https://www.ebi.ac.uk/gwas/variants/rs2279501) Serum metabolite levels *(3-hydroxysebacate)* [rs7541453](https://www.ebi.ac.uk/gwas/variants/rs7541453) Serum metabolite levels *(indoleacetylglutamine)* | [rs1038747](https://www.ebi.ac.uk/gwas/variants/rs1038747) Plasma X-21607 levels in chronic kidney disease [rs10788817](https://www.ebi.ac.uk/gwas/variants/rs10788817) 3-hydroxylaurate levels Metabolite peak levels (QI2718) Tetradecadienoate (14:2) levels [rs10888445](https://www.ebi.ac.uk/gwas/variants/rs10888445) Beta-hydroxyisovalerate levels [rs11204914](https://www.ebi.ac.uk/gwas/variants/rs11204914) Carnitine C9 levels Metabolite peak levels (QI6204) Metabolite peak levels (QI6252) [rs2130525](https://www.ebi.ac.uk/gwas/variants/rs2130525) Glycine conjugate of C10H14O2 (1) levels [rs2338201](https://www.ebi.ac.uk/gwas/variants/rs2338201) Dodecadienoate (12:2) (X-21343) levels Dodecadienoate (12:2) levels Tetradecadienoate (14:2) (X-12442) levels [rs2999541](https://www.ebi.ac.uk/gwas/variants/rs2999541) Cis-3,4-methyleneheptanoylcarnitine levels [rs2999545](https://www.ebi.ac.uk/gwas/variants/rs2999545) Pimeloylcarnitine/3-methyladipoylcarnitine (C7-DC) levels [rs6685187](https://www.ebi.ac.uk/gwas/variants/rs6685187) X-12442–5,8-tetradecadienoate levels [rs6698740](https://www.ebi.ac.uk/gwas/variants/rs6698740) Beta-hydroxyisovalerate levels [rs7523082](https://www.ebi.ac.uk/gwas/variants/rs7523082) Metabolite peak levels (QI9845) [rs7542137](https://www.ebi.ac.uk/gwas/variants/rs7542137) 3-hydroxysebacate levels Beta-hydroxyisovalerate levels Plasma cis-4-decenoate (10:1n6)* levels in chronic kidney disease |
| 2:135598913 | rs17322446 | 1-Methylnicotinamide | [rs10496732](https://www.ebi.ac.uk/gwas/variants/rs10496732) Urine quinolinate levels in chronic kidney disease |  | [rs17322446](https://www.ebi.ac.uk/gwas/variants/rs17322446) 1-methylnicotinamide levels N1-methyl-2-pyridone-5-carboxamide levels [rs17698630](https://www.ebi.ac.uk/gwas/variants/rs17698630) Walking pace [rs1942052](https://www.ebi.ac.uk/gwas/variants/rs1942052) Quinolinate levels [rs41449851](https://www.ebi.ac.uk/gwas/variants/rs41449851) FEV1 [rs76328456](https://www.ebi.ac.uk/gwas/variants/rs76328456) Quinolinate levels |
| 2:211540507 | rs1047891 | Glycine | [rs1047891](https://www.ebi.ac.uk/gwas/variants/rs1047891) Urinary metabolite levels in chronic kidney disease *(2-methylbutyrylglycine)* Urinary metabolite levels in chronic kidney disease *(3-methylcrotonylglycine)* Urinary metabolite levels in chronic kidney disease *(3-methylglutaconate)* Urinary metabolite levels in chronic kidney disease *(3-methylglutarate/2-methylglutarate)* Urinary metabolite levels in chronic kidney disease *(3-methylglutarylcarnitine (2))* Urinary metabolite levels in chronic kidney disease *(X - 16567)* Urinary metabolite levels in chronic kidney disease *(X - 24402)* Urinary metabolite levels in chronic kidney disease *(hexanoylglycine)* Urinary metabolite levels in chronic kidney disease *(isobutyrylglycine)* Urinary metabolite levels in chronic kidney disease *(isocaproylglycine)* Urinary metabolite levels in chronic kidney disease *(isovalerylglycine)* Urinary metabolite levels in chronic kidney disease *(tigloylglycine)* Urinary metabolite modules (eigenmetabolites) in chronic kidney disease *(2-methylbutyrylglycine, 3-methylcrotonylglycine, indoleacetylglutamine, indoleacetylglycine, isobutyrylglycine, isovalerylglutamine, isovalerylglycine, tigloylglycine, X-17688)* Urinary metabolite modules (eigenmetabolites) in chronic kidney disease *(3-hydroxy-3-methylglutarate, 3-methylglutaconate, 3-methylglutarate, 2-methylglutarate, 3-methylglutarylcarnitine)* Urinary metabolite modules (eigenmetabolites) in chronic kidney disease *(hexanoylglycine, isocaproylglycine, X-16567, X-24402)* Urine 2-butenoylglycine levels in chronic kidney disease Urine 2-methylbutyrylglycine levels in chronic kidney disease Urine 3-hydroxyoctanoyl glycine levels in chronic kidney disease Urine 3-methylcrotonylglycine levels in chronic kidney disease Urine 3-methylglutaconate levels in chronic kidney disease Urine 3-methylglutarylcarnitine (2) levels in chronic kidney disease Urine 4-methylhexanoylglycine levels in chronic kidney disease Urine 5-hydroxymethyl-2-furoylcarnitine* levels in chronic kidney disease Urine X-17325 levels in chronic kidney disease Urine X-17367 levels in chronic kidney disease Urine X-24465 levels in chronic kidney disease Urine glycine conjugate of C6H10O2 (1)* levels in chronic kidney disease Urine glycine levels in chronic kidney disease Urine hexanoylglycine levels in chronic kidney disease Urine indolepropionylglycine levels in chronic kidney disease Urine isobutyrylglycine levels in chronic kidney disease Urine isocaproylglycine levels in chronic kidney disease Urine isovalerylglycine levels in chronic kidney disease Urine phenylpropionylglycine levels in chronic kidney disease Urine tigloylglycine levels in chronic kidney disease [rs715](https://www.ebi.ac.uk/gwas/variants/rs715) Urinary metabolites *(3.555 ppm/2.547 ppm)* Urinary metabolites *(glycine/threonine)* Urine 2-methylserine levels in chronic kidney disease Urine 3-methylglutarate/2-methylglutarate levels in chronic kidney disease Urine X-24406 levels in chronic kidney disease Urine cis-3,4-methyleneheptanoylglycine levels in chronic kidney disease Urine trans-2-hexenoylglycine levels in chronic kidney disease Urine trans-3,4-methyleneheptanoylglycine levels in chronic kidney disease | [rs1047891](https://www.ebi.ac.uk/gwas/variants/rs1047891) Blood urea nitrogen levels Serum metabolite concentrations in chronic kidney disease *(Glycine)* Serum metabolite levels *(3-methylglutaconate)* Serum metabolite levels *(3-methylglutarylcarnitine (2))* Serum metabolite levels *(3b-hydroxy-5-cholenoic acid)* Serum metabolite levels *(N-acetylglycine)* Serum metabolite levels *(N-palmitoylglycine)* Serum metabolite levels *(betaine)* Serum metabolite levels *(cinnamoylglycine)* Serum metabolite levels *(creatine)* Serum metabolite levels *(gamma-glutamylglycine)* Serum metabolite levels *(glycine)* Serum metabolite levels *(homoarginine)* Serum metabolite levels *(isobutyrylglycine)* Serum metabolite levels *(isovalerylglycine)* Serum metabolite levels *(propionylglycine)* Serum metabolite levels *(serine)* Serum uric acid levels [rs715](https://www.ebi.ac.uk/gwas/variants/rs715) Serum metabolite levels *(Creatine)* Serum metabolite levels *(Glycine)* | [rs1047891](https://www.ebi.ac.uk/gwas/variants/rs1047891) 1-ribosyl-imidazoleacetate (X-11334) levels 2-butenoylglycine levels 2R,3R-dihydroxybutyrate levels 3-hydroxybutyroylglycine levels 3-methylglutaconate levels 3-methylglutarylcarnitine (2) levels 5-methylthioadenosine (mta) levels Alanine aminotransferase levels Alanine levels Alanine transaminase levels Apolipoprotein A1 levels Appendicular lean mass Betaine levels Body surface area Butyrylglycine levels Ceramide (d17:1/24:1) levels Cholesterol levels in medium HDL Cholesterol to total lipids ratio in IDL Cholesterol to total lipids ratio in small LDL Cholesteryl ester levels in HDL Cholesteryl ester levels in medium HDL Chronic elevation of alanine aminotransferase (cALT) levels Chronic kidney disease Cinnamoylglycine levels Cis-3,4-methyleneheptanoylglycine levels Citrate levels Citrate levels (UKB data field 23473) Concentration of HDL particles Concentration of medium HDL particles Creatine levels Creatinine levels Creatinine levels (UKB data field 23478) Diastolic blood pressure Eosinophil counts Estimated glomerular filtration rate Estimated glomerular filtration rate (creatinine) Estimated glomerular filtration rate (creatinine) *(conditioned on rs72944180)* Estimated glomerular filtration rate (cystatin c) Estimated glomerular filtration rate *(EA)* Estimated glomerular filtration rate in diabetes Estimated glomerular filtration rate in non-diabetics Fat-free mass Fat-free mass *(female)* Free androgen index Free cholesterol levels in medium HDL Gamma-glutamylglycine levels Glomerular filtration rate (creatinine) Glutarylcarnitine (c5-dc) levels Glycine levels Glycine levels (UKB data field 23462) Glycine levels in chronic kidney disease Glycosyl-N-(2-hydroxynervonoyl)-sphingosine (d18:1/24:1(2OH)) levels HDL cholesterol HDL cholesterol levels HDL cholesterol levels in current drinkers HDL cholesterol levels x alcohol consumption (drinkers vs non-drinkers) interaction (2df) HDL cholesterol levels x alcohol consumption (drinkers vs non-drinkers) interaction (2df) *(EA)* HDL cholesterol levels x alcohol consumption (regular vs non-regular drinkers) interaction (2df) HDL cholesterol levels x alcohol consumption (regular vs non-regular drinkers) interaction (2df) *(EA)* Height Hexanoylglycine levels High density lipoprotein cholesterol levels Histidine levels Histidine levels (UKB data field 23463) Homoarginine levels Isobutyrylglycine levels Isovalerylglycine levels Lipoprotein (a) levels Liver enzyme levels (alkaline phosphatase) Lymphocyte count Macular telangiectasia type 2 Mean corpuscular hemoglobin Mean corpuscular volume Mean platelet volume Metabolite levels *(Glycine)* Metabolite peak levels (QI6688) Metabolite peak levels (QI6816) *(African American or Afro-Caribbean)* Metabolite peak levels (QI6823) Metabolite peak levels (QI6840) Metabolite peak levels (QI9202) Metabolite peak levels (QI9368) Metabolite peak levels (QI9376) Methylglutarylcarnitine levels N-acetylaspartic acid levels *(African American or Afro-Caribbean)* N-acetylglycine levels N-palmitoylglycine levels NMMA levels Neutrophil count Phospholipids to total lipids ratio in IDL Phospholipids to total lipids ratio in very small VLDL Plasma 2,3-dihydroxy-5-methylthio-4-pentenoate (DMTPA)* levels in chronic kidney disease Plasma 3-methylcrotonylglycine levels in chronic kidney disease Plasma 3-methylglutaconate levels in chronic kidney disease Plasma 3-methylglutarylcarnitine (2) levels in chronic kidney disease Plasma N-acetylglycine levels in chronic kidney disease Plasma N-palmitoylglycine levels in chronic kidney disease Plasma X-25982 levels in chronic kidney disease Plasma betaine levels in chronic kidney disease Plasma choline levels in chronic kidney disease Plasma gamma-glutamylglycine levels in chronic kidney disease Plasma glycine levels in chronic kidney disease Plasma hexanoylglycine levels in chronic kidney disease Plasma homoarginine levels in chronic kidney disease Plasma homocysteine levels (post-methionine load test) Plasma propionylglycine levels in chronic kidney disease Plasma tigloylglycine levels in chronic kidney disease Platelet count Propionylglycine levels Protein quantitative trait loci (liver) *(GLDC)* Pyroglutamine levels Red blood cell count Red cell distribution width Red meat related metabolomic signature Serine levels Serum 25-Hydroxyvitamin D levels Serum alkaline phosphatase levels Serum creatinine levels Serum levels of protein SHMT2 Serum total protein level Sex hormone-binding globulin levels Sex hormone-binding globulin levels adjusted for BMI Sex hormone-binding globulin levels in postmenopausal women Sex hormone-binding globulin levels in premenopausal women Systolic blood pressure Systolic blood pressure (standard GWA) Threonine levels Total bilirubin levels Total lipid levels in medium HDL Trans-2-hexenoylglycine levels Urate levels Urate levels *(women)* Urinary albumin excretion Urinary albumin excretion (no hypertensive medication) Urinary albumin-to-creatinine ratio Urinary albumin-to-creatinine ratio *(EA)* Urinary potassium to creatinine ratio Urinary sodium to creatinine ratio Weight White blood cell count X-17325 levels X-17367 levels X-21467 levels X-24757 levels X-24795 levels [rs715](https://www.ebi.ac.uk/gwas/variants/rs715) Arginine levels Aspartate levels Betaine levels Betaine levels in individuals undergoing cardiac evaluation Body mass index Body mass index (MTAG) Body mass index or knee osteoarthritis (pleiotropy) Calcium levels Cholelithiasis Coronary artery disease or fibrinogen levels (pleiotropy) Creatine levels Creatinine levels Eosinophil counts Eosinophil percentage of granulocytes Eosinophil percentage of white cells Fibrinogen *(EA)* Fibrinogen levels Fibrinogen levels or factor VII levels (pleiotropy) Fibrinogen levels or factor VII levels or factor XI levels or tissue plasminogen activator levels (pleiotropy) Fibrinogen levels or plasminogen activator inhibitor 1 levels (pleiotropy) Fibrinogen levels or tissue plasminogen activator levels (pleiotropy) Gamma-glutamylglycine levels Glomerular filtration rate Glutamine degradant levels Glutaroyl carnitine levels Glycine levels Height Histidine levels Ischemic stroke or fibrinogen levels (pleiotropy) Macular telangiectasia type 2 Mean corpuscular hemoglobin Mean corpuscular volume Metabolite levels *(glycine)* N-acetylglycine levels Neutrophil count Phenylalanine levels Plasma 3-hydroxybutyroylglycine** levels in chronic kidney disease Plasma cis-3,4-methyleneheptanoylglycine levels in chronic kidney disease Plasma free amino acid levels (adjusted for twenty other PFAAs) *(Gly)* Platelet distribution width Plateletcrit Propionylglycine levels Pyroglutamine levels Red blood cell count Red cell distribution width Serine levels Threonine levels Tyrosine levels Urate levels Venous thromboembolism or fibrinogen levels (pleiotropy) X-08988 levels X-16570 levels X-17325 levels X-25420 levels |
| 2:241793545 | rs55649245 | 4-Deoxyerythronic acid |  |  | [rs55649245](https://www.ebi.ac.uk/gwas/variants/rs55649245) 2R,3R-dihydroxybutyrate levels Plasma 2R,3R-dihydroxybutyrate levels in chronic kidney disease X-23639 levels |
| 2:241813788 | rs10933641 | 4-Deoxyerythronic acid | [rs6748734](https://www.ebi.ac.uk/gwas/variants/rs6748734) Urinary metabolites *(1.086 ppm)* |  | [rs4344931](https://www.ebi.ac.uk/gwas/variants/rs4344931) Height [rs4675869](https://www.ebi.ac.uk/gwas/variants/rs4675869) X-23639 levels [rs4675874](https://www.ebi.ac.uk/gwas/variants/rs4675874) X-12556 levels [rs6739772](https://www.ebi.ac.uk/gwas/variants/rs6739772) Hip circumference adjusted for BMI *(EA)* Hip circumference adjusted for BMI *(EA, women)* [rs6752657](https://www.ebi.ac.uk/gwas/variants/rs6752657) Lung function (FVC) |
| 3:182758040 | rs4859267 | 3-Hydroxyisovalerate |  |  | [rs11928508](https://www.ebi.ac.uk/gwas/variants/rs11928508) Lymphocyte count [rs2270968](https://www.ebi.ac.uk/gwas/variants/rs2270968) Hydroxyisovaleroyl carnitine levels Parkinson’s disease |
| 4:88213884 | rs6811902 | Propylene Glycol |  |  | [rs10026092](https://www.ebi.ac.uk/gwas/variants/rs10026092) Protein quantitative trait loci (liver) *(HSD17B13)* [rs11735092](https://www.ebi.ac.uk/gwas/variants/rs11735092) Height Phospholipids to total lipids ratio in small HDL Platelet count Total testosterone levels [rs13133311](https://www.ebi.ac.uk/gwas/variants/rs13133311) Free androgen index Protein quantitative trait loci (liver) *(HSD17B13)* [rs13141053](https://www.ebi.ac.uk/gwas/variants/rs13141053) Liver enzyme levels (gamma-glutamyl transferase) Triglyceride levels x long total sleep time interaction (2df test) Triglyceride levels x short total sleep time interaction (2df test) [rs13142655](https://www.ebi.ac.uk/gwas/variants/rs13142655) Aspartate aminotransferase levels [rs13150068](https://www.ebi.ac.uk/gwas/variants/rs13150068) Phospholipids to total lipids ratio in large HDL Platelet count Sex hormone-binding globulin levels Sex hormone-binding globulin levels adjusted for BMI Sex hormone-binding globulin levels in postmenopausal women [rs6811902](https://www.ebi.ac.uk/gwas/variants/rs6811902) Low testosterone levels Sex hormone-binding globulin levels [rs6831888](https://www.ebi.ac.uk/gwas/variants/rs6831888) Total testosterone levels [rs6849526](https://www.ebi.ac.uk/gwas/variants/rs6849526) Platelet count [rs7694379](https://www.ebi.ac.uk/gwas/variants/rs7694379) Alanine transaminase levels in high alcohol intake Aspartate aminotransferase platelet ratio index in high alcohol intake Aspartate transaminase levels in high alcohol intake Platelet count Protein quantitative trait loci (liver) *(HSD17B13)* Sex hormone-binding globulin levels Sex hormone-binding globulin levels adjusted for BMI |
| 5:1188285 | rs11133665 | Glycine, Histidine, Threonine, Tryptophan, Tyrosine | [rs11133665](https://www.ebi.ac.uk/gwas/variants/rs11133665) Urinary metabolite levels in chronic kidney disease *(3-hydroxykynurenine)* Urinary metabolite levels in chronic kidney disease *(kynurenine)* Urinary metabolite levels in chronic kidney disease *(phenylalanine)* Urinary metabolite levels in chronic kidney disease *(tryptophan)* Urinary metabolite levels in chronic kidney disease *(tyrosine)* Urinary metabolites *(histidine/τ-methylhistidine)* Urine 6-bromotryptophan levels in chronic kidney disease Urine N-acetyl-1-methylhistidine* levels in chronic kidney disease Urine asparagine levels in chronic kidney disease Urine histidine levels in chronic kidney disease Urine kynurenine levels in chronic kidney disease Urine methionine sulfoxide levels in chronic kidney disease Urine phenylalanine levels in chronic kidney disease Urine tryptophan levels in chronic kidney disease Urine tyrosine levels in chronic kidney disease [rs11750211](https://www.ebi.ac.uk/gwas/variants/rs11750211) Urine 3-hydroxykynurenine levels in chronic kidney disease |  | [rs111306756](https://www.ebi.ac.uk/gwas/variants/rs111306756) 1-methylhistidine levels [rs11133665](https://www.ebi.ac.uk/gwas/variants/rs11133665) 1-methylhistidine levels Methionine sulfone levels Plasma methionine sulfone levels in chronic kidney disease [rs11750211](https://www.ebi.ac.uk/gwas/variants/rs11750211) 3-methoxytyrosine levels Estimated glomerular filtration rate (creatinine) |
| 5:34993215 | rs11744796 | 3-Aminoisobutyrate |  |  | [rs10941225](https://www.ebi.ac.uk/gwas/variants/rs10941225) Menarche (age at onset) [rs11744796](https://www.ebi.ac.uk/gwas/variants/rs11744796) 3-aminoisobutyrate levels [rs37440](https://www.ebi.ac.uk/gwas/variants/rs37440) Protein quantitative trait loci (liver) *(AGXT2)* |
| 5:35003112 | rs468327 | 3-Aminoisobutyrate | [rs468327](https://www.ebi.ac.uk/gwas/variants/rs468327) Urinary metabolites (H-NMR features) *(3.0275, Aminoisobutyrate)* |  |  |
| 5:35037115 | rs37369 | 3-Aminoisobutyrate | [rs37369](https://www.ebi.ac.uk/gwas/variants/rs37369) Urinary metabolite levels in chronic kidney disease *(3-aminoisobutyrate)* Urinary metabolite levels in chronic kidney disease *(5-aminovalerate)* Urinary metabolite levels in chronic kidney disease *(X - 12096)* Urinary metabolite levels in chronic kidney disease *(X - 12097)* Urinary metabolite levels in chronic kidney disease *(X - 12117)* Urinary metabolite levels in chronic kidney disease *(X - 24387)* Urinary metabolite levels in chronic kidney disease *(X - 24518)* Urinary metabolite modules (eigenmetabolites) in chronic kidney disease *(3-aminoisobutyrate, 5-aminovalerate, X-12096, X-12097, X-12117, X-24518)* Urinary metabolites (H-NMR features) *(1.1975, Aminoisobutyrate)* Urinary metabolites (H-NMR features) *(1.2025, Aminoisobutyrate)* Urinary metabolites (H-NMR features) *(2.6075, Aminoisobutyrate)* Urinary metabolites (H-NMR features) *(2.6125, Aminoisobutyrate)* Urinary metabolites (H-NMR features) *(2.6275, Aminoisobutyrate)* Urinary metabolites (H-NMR features) *(3.0975, Aminoisobutyrate)* Urinary metabolites (H-NMR features) *(3.1075, Aminoisobutyrate)* Urinary metabolites *(1.171 ppm/1.973 ppm)* Urinary metabolites *(3-Aminoisobutyrate concentration)* Urinary metabolites *(3-aminoisobutyrate)* Urine 3-aminoisobutyrate levels in chronic kidney disease Urine 5-aminovalerate levels in chronic kidney disease Urine N2,N2-dimethylguanosine levels in chronic kidney disease Urine X-12096 levels in chronic kidney disease Urine X-12097 levels in chronic kidney disease Urine X-12125 levels in chronic kidney disease Urine X-24387 levels in chronic kidney disease Urine dimethylarginine (SDMA + ADMA) levels in chronic kidney disease | [rs37369](https://www.ebi.ac.uk/gwas/variants/rs37369) Serum metabolite levels *(3-aminoisobutyrate)* | [rs37369](https://www.ebi.ac.uk/gwas/variants/rs37369) 3-aminoisobutyrate levels Beta-aminoisobutyric acid levels *(Hispanic)* DMGV levels Dimethylarginine (sdma + adma) levels Dimethylguanido valerate levels in chronic kidney disease Metabolite peak levels (QI10969) *(African American or Afro-Caribbean)* Plasma 3-aminoisobutyrate levels in chronic kidney disease Plasma X-12117 levels in chronic kidney disease Plasma X-24518 levels in chronic kidney disease Plasma dimethylarginine (SDMA + ADMA) levels in chronic kidney disease Protein quantitative trait loci (liver) *(AGXT2)* Serum dimethylarginine levels (asymmetric/symetric ratio) Symmetric dimethylarginine levels Symmetric dimethylarginine levels in chronic kidney disease Symmetrical dimethylarginine levels X-12117 levels X-24518 levels |
| 5:35039437 | rs2279651 | 3-Aminoisobutyrate |  |  | [rs2279651](https://www.ebi.ac.uk/gwas/variants/rs2279651) X-24518 levels [rs6892674](https://www.ebi.ac.uk/gwas/variants/rs6892674) 3-aminoisobutyrate levels Protein quantitative trait loci (liver) *(AGXT2)* |
| 5:150702299 | rs61067578 | Glycine |  |  | [rs67910448](https://www.ebi.ac.uk/gwas/variants/rs67910448) Glutamine degradant levels |
| 5:150708711 | rs147000073 | Glycine | [rs149235996](https://www.ebi.ac.uk/gwas/variants/rs149235996) Urine glycine levels in chronic kidney disease |  | [rs146767775](https://www.ebi.ac.uk/gwas/variants/rs146767775) Glutamine degradant levels [rs147000073](https://www.ebi.ac.uk/gwas/variants/rs147000073) Estimated glomerular filtration rate (creatinine) [rs149235996](https://www.ebi.ac.uk/gwas/variants/rs149235996) Acetylcarnitine levels Carnitine levels [rs77010315](https://www.ebi.ac.uk/gwas/variants/rs77010315) Carnitine levels Hexanoylcarnitine levels (Biocrates platform) Octanoylcarnitine levels Propionylcarnitine levels X-11381 levels X-24801 levels |
| 6:111492119 | rs241768 | Tyrosine |  |  | [rs1216019](https://www.ebi.ac.uk/gwas/variants/rs1216019) Tryptophan levels [rs241768](https://www.ebi.ac.uk/gwas/variants/rs241768) Tyrosine levels [rs9400467](https://www.ebi.ac.uk/gwas/variants/rs9400467) Tyrosine levels |
| 7:17287998 | rs2106727 | Quinic acid, Trigonelline |  | [rs4410790](https://www.ebi.ac.uk/gwas/variants/rs4410790) Blood urea nitrogen levels [rs6968865](https://www.ebi.ac.uk/gwas/variants/rs6968865) Blood urea nitrogen levels | [rs2106727](https://www.ebi.ac.uk/gwas/variants/rs2106727) 1-methylxanthine levels Coffee consumption (cups per day) Serum creatinine levels Sex hormone-binding globulin levels adjusted for BMI Total bilirubin levels Triglyceride levels [rs4410790](https://www.ebi.ac.uk/gwas/variants/rs4410790) Bitter beverage consumption Bitter non-alcoholic beverage consumption Body mass index Caffeine consumption Caffeine consumption from coffee Caffeine consumption from coffee or tea Caffeine consumption from tea Caffeine levels Caffeine metabolism (plasma 1,3,7-trimethylxanthine (caffeine) level) Caffeine metabolism (plasma 1,7-dimethylxanthine (paraxanthine) to 1,3,7-trimethylxanthine (caffeine) ratio) Coffee consumption Coffee consumption (cups per day) Estimated glomerular filtration rate HDL cholesterol levels Microalbuminuria Non-HDL cholesterol levels Tea consumption Triglyceride levels Urinary albumin excretion Urinary albumin excretion (no hypertensive medication) Urinary albumin-to-creatinine ratio Urinary albumin-to-creatinine ratio *(EA)* Urinary potassium excretion *(EA)* Urinary sodium excretion *(EA)* [rs6968554](https://www.ebi.ac.uk/gwas/variants/rs6968554) Body mass index Body mass index (MTAG) Caffeine levels Caffeine metabolism (plasma 1,3,7-trimethylxanthine (caffeine) level) Caffeine metabolism (plasma 1,7-dimethylxanthine (paraxanthine) to 1,3,7-trimethylxanthine (caffeine) ratio) Coffee consumption Coffee consumption (cups per day) Estimated glomerular filtration rate Hemoglobin A1c levels High density lipoprotein cholesterol levels [rs6968865](https://www.ebi.ac.uk/gwas/variants/rs6968865) Coffee consumption Coffee consumption (cups per day) Coffee max liking Estimated glomerular filtration rate (creatinine) F-chocolate/coffee liking (derived food-liking factor) Low density lipoprotein cholesterol levels Triglyceride levels Triglyceride levels in non-type 2 diabetes |
| 8:18272377 | rs4921913 | Formate | [rs1495741](https://www.ebi.ac.uk/gwas/variants/rs1495741) Urinary metabolite levels in chronic kidney disease *(1-methylxanthine)* [rs35246381](https://www.ebi.ac.uk/gwas/variants/rs35246381) Urinary metabolite levels in chronic kidney disease *(5-acetylamino-6-amino-3-methyluracil)* Urinary metabolite levels in chronic kidney disease *(5-acetylamino-6-formylamino-3-methyluracil)* Urinary metabolite levels in chronic kidney disease *(N-acetylputrescine)* Urinary metabolite modules (eigenmetabolites) in chronic kidney disease *(4-acetamidobutanoate, allo-threonine, N-acetylputrescine)* Urinary metabolites *(2.159 ppm/3.324 ppm)* Urine 5-acetylamino-6-amino-3-methyluracil levels in chronic kidney disease Urine 5-acetylamino-6-formylamino-3-methyluracil levels in chronic kidney disease Urine N-acetylputrescine levels in chronic kidney disease [rs4921914](https://www.ebi.ac.uk/gwas/variants/rs4921914) Urinary metabolite levels in chronic kidney disease *(1-methylurate)* Urinary metabolite levels in chronic kidney disease *(X - 12410)* Urinary metabolites (H-NMR features) *(2.1875, Unknown)* Urinary metabolites *(Formate/succinate ratio)* Urine 4-acetamidobutanoate levels in chronic kidney disease Urine X-12410 levels in chronic kidney disease [rs4921915](https://www.ebi.ac.uk/gwas/variants/rs4921915) Urinary metabolite levels in chronic kidney disease *(4-acetamidobutanoate)* Urine N-acetyl-cadaverine levels in chronic kidney disease | [rs146812806](https://www.ebi.ac.uk/gwas/variants/rs146812806) Serum metabolite levels *(5-acetylamino-6-amino-3-methyluracil)* [rs35246381](https://www.ebi.ac.uk/gwas/variants/rs35246381) Serum metabolite levels *(5-acetylamino-6-formylamino-3-methyluracil)* [rs35570672](https://www.ebi.ac.uk/gwas/variants/rs35570672) Serum metabolite levels *(1-methylxanthine)* Serum metabolite levels *(N-acetylputrescine)* [rs4921913](https://www.ebi.ac.uk/gwas/variants/rs4921913) Serum metabolite levels *(4-acetamidobutanoate)* | [rs11784251](https://www.ebi.ac.uk/gwas/variants/rs11784251) N-acetylputrescine levels in chronic kidney disease Triglyceride levels [rs146812806](https://www.ebi.ac.uk/gwas/variants/rs146812806) Mean corpuscular hemoglobin concentration [rs1495741](https://www.ebi.ac.uk/gwas/variants/rs1495741) 1-methylurate levels 4-acetamidobutanoate levels in chronic kidney disease 5-acetylamino-6-amino-3-methyluracil levels 5-acetylamino-6-formylamino-3-methyluracil levels 5-acetylamino-6-formylamino-3-methyluracil levels in elite athletes Apolipoprotein B levels Bladder cancer Cholesterol in chylomicrons and extremely large VLDL (UKB data field 23484) Cholesterol levels in chylomicrons and extremely large VLDL Cholesterol to total lipids ratio in medium HDL Cholesterol to total lipids ratio in small HDL Cholesterol, total Cholesteryl ester levels in chylomicrons and extremely large VLDL Cholesteryl esters in chylomicrons and extremely large VLDL (UKB data field 23485) Cholesteryl esters to total lipids ratio in large HDL Cholesteryl esters to total lipids ratio in medium HDL Cholesteryl esters to total lipids ratio in small HDL Cholesteryl esters to total lipids ratio in small LDL Cholesteryl esters to total lipids ratio in very large HDL Concentration of chylomicrons and extremely large VLDL particles Free cholesterol in chylomicrons and extremely large VLDL (UKB data field 23486) Free cholesterol levels in chylomicrons and extremely large VLDL Free cholesterol to total lipids ratio in large LDL Free cholesterol to total lipids ratio in medium LDL Free cholesterol to total lipids ratio in small LDL LDL cholesterol levels Liver injury in anti-tuberculosis drug treatment *(recessive)* Low density lipoprotein cholesterol levels Medication use (HMG CoA reductase inhibitors) Medication use for hyperlipidemia (number of purchases) Monounsaturated fatty acid levels Monounsaturated fatty acids levels (UKB data field 23447) N-acetylputrescine levels in chronic kidney disease N-acetylputrescine levels in elite athletes Non-HDL cholesterol levels Phospholipids to total lipids ratio in large HDL Phospholipids to total lipids ratio in large VLDL Phospholipids to total lipids ratio in medium HDL Phospholipids to total lipids ratio in small HDL Phospholipids to total lipids ratio in small LDL Polyunsaturated fatty acids to monounsaturated fatty acids ratio (UKB data field 23458) Polyunsaturated fatty acids to total fatty acids percentage (UKB data field 23453) Ratio of monounsaturated fatty acids to total fatty acids Ratio of omega-6 fatty acids to total fatty acids Ratio of polyunsaturated fatty acids to monounsaturated fatty acids Ratio of polyunsaturated fatty acids to total fatty acids Saturated fatty acid levels Saturated fatty acids levels (UKB data field 23448) Total cholesterol levels Total cholesterol levels *(EA)* Total fatty acid levels Total fatty acids levels (UKB data field 23442) Total lipid levels in chylomicrons and extremely large VLDL Triglyceride levels Triglyceride levels in HDL Triglyceride levels in IDL Triglyceride levels in LDL Triglyceride levels in large HDL Triglyceride levels in large LDL Triglyceride levels in medium HDL Triglyceride levels in medium LDL Triglyceride levels in small LDL Triglyceride levels in very large HDL Triglyceride levels in very small VLDL Triglycerides Triglycerides *(EA)* Triglycerides in IDL (UKB data field 23529) Triglycerides in LDL (UKB data field 23409) Triglycerides in medium LDL (UKB data field 23543) Triglycerides in small LDL (UKB data field 23550) Triglycerides in very large HDL (UKB data field 23557) Triglycerides to total lipids ratio in large HDL Triglycerides to total lipids ratio in medium HDL Triglycerides to total lipids ratio in medium LDL Triglycerides to total lipids ratio in small HDL Triglycerides to total lipids ratio in small LDL Triglycerides to total lipids ratio in very large HDL Youthful appearance (self-reported) [rs1495745](https://www.ebi.ac.uk/gwas/variants/rs1495745) C-reactive protein levels Cholesteryl esters to total lipids ratio in very small VLDL [rs1495747](https://www.ebi.ac.uk/gwas/variants/rs1495747) C-reactive protein levels (MTAG) Calcium levels N-acetylputrescine levels [rs35246381](https://www.ebi.ac.uk/gwas/variants/rs35246381) 4-acetamidobutanoate levels Cholesterol to total lipids ratio in large HDL Cholesterol to total lipids ratio in medium VLDL Cholesterol to total lipids ratio in small VLDL Cholesterol to total lipids ratio in very small VLDL Cholesteryl esters to total lipids ratio in medium VLDL Free cholesterol to total lipids in large LDL percentage (UKB data field 23617) Free cholesterol to total lipids in small LDL percentage (UKB data field 23627) Free cholesterol to total lipids in small VLDL percentage (UKB data field 23602) Free cholesterol to total lipids ratio in IDL Free cholesterol to total lipids ratio in medium VLDL Free cholesterol to total lipids ratio in small VLDL Free cholesterol to total lipids ratio in very small VLDL Mean corpuscular hemoglobin concentration N-acetylputrescine levels Phosphatidylcholine levels Phosphoglycerides levels Phospholipid levels in small HDL Phospholipids to total lipids ratio in small VLDL Plasma 5-acetylamino-6-amino-3-methyluracil levels in chronic kidney disease Plasma N-acetylputrescine levels in chronic kidney disease Triglyceride levels Triglyceride levels (MTAG) Triglyceride levels x short total sleep time interaction (2df test) Triglycerides in very small VLDL (UKB data field 23522) Triglycerides to total lipids in small LDL percentage (UKB data field 23628) Triglycerides to total lipids in very small VLDL percentage (UKB data field 23608) Triglycerides to total lipids ratio in medium VLDL Triglycerides to total lipids ratio in small VLDL Triglycerides to total lipids ratio in very small VLDL Triglycerides x physical activity interaction (2df test) [rs35570672](https://www.ebi.ac.uk/gwas/variants/rs35570672) 1-methylurate levels 1-methylxanthine levels Mean corpuscular hemoglobin Total cholesterol levels Triglyceride levels X-12410 levels [rs4921913](https://www.ebi.ac.uk/gwas/variants/rs4921913) 1-methylxanthine levels 4-acetamidobutanoate levels 4-acetamidobutanoate-to-N1-methyladenosine ratio 5-acetylamino-6-amino-3-methyluracil levels 5-acetylamino-6-formylamino-3-methyluracil levels Average diameter for VLDL particles Cholesterol levels in large VLDL Cholesterol levels in very large VLDL Cholesteryl ester levels in large VLDL Cholesteryl ester levels in very large VLDL Concentration of chylomicrons and extremely large VLDL particles (UKB data field 23481) Concentration of large VLDL particles Concentration of small VLDL particles Concentration of very large VLDL particles Free cholesterol levels in VLDL Free cholesterol levels in large VLDL Liver enzyme levels (gamma-glutamyl transferase) Low-density lipoprotein levels (MTAG) Multi-trait sum score N-acetylputrescine levels Omega-6 fatty acid levels Phospholipid levels in VLDL Phospholipid levels in large VLDL Phospholipid levels in very large VLDL Phospholipids in chylomicrons and extremely large VLDL (UKB data field 23483) Total lipid levels in VLDL Total lipid levels in large VLDL Total lipid levels in small VLDL Total lipid levels in very large VLDL Total lipids in chylomicrons and extremely large VLDL (UKB data field 23482) Triglyceride levels Triglyceride levels in VLDL Triglyceride levels in chylomicrons and extremely large VLDL Triglyceride levels in large VLDL Triglyceride levels in medium VLDL Triglyceride levels in very large VLDL Triglycerides in chylomicrons and extremely large VLDL (UKB data field 23487) [rs4921914](https://www.ebi.ac.uk/gwas/variants/rs4921914) 1-methylxanthine levels 5-acetylamino-6-amino-3-methyluracil levels 5-acetylamino-6-formylamino-3-methyluracil levels Mean corpuscular hemoglobin concentration N-acetylputrescine levels Total cholesterol levels Triglyceride levels *(Trans-ethnic initial)* [rs4921915](https://www.ebi.ac.uk/gwas/variants/rs4921915) Albumin levels Cholesterol to total lipids ratio in very large VLDL Cholesteryl esters to total lipids ratio in very large VLDL Free cholesterol levels in very large VLDL Free cholesterol to total lipids in medium LDL percentage (UKB data field 23622) Iron status biomarkers (transferrin levels) Phospholipid levels in chylomicrons and extremely large VLDL Plasma X-12410 levels in chronic kidney disease Polyunsaturated fatty acid levels Ratio of triglycerides to phosphoglycerides Total cholesterol levels Total lipid levels in small HDL Total triglycerides levels Triglyceride levels in small HDL Triglyceride levels in small VLDL Triglycerides |
| 8:74868909 | rs72661850 | 4-Deoxyerythronic acid |  |  | [rs62509311](https://www.ebi.ac.uk/gwas/variants/rs62509311) Liver enzyme levels (alkaline phosphatase) Low density lipoprotein cholesterol levels [rs7013120](https://www.ebi.ac.uk/gwas/variants/rs7013120) Total cholesterol levels [rs72661850](https://www.ebi.ac.uk/gwas/variants/rs72661850) Gamma-glutamylthreonine levels [rs72661853](https://www.ebi.ac.uk/gwas/variants/rs72661853) Serum alkaline phosphatase levels Threonine levels |
| 9:136146597 | rs550057 | Xylose |  | [rs550057](https://www.ebi.ac.uk/gwas/variants/rs550057) Blood urea nitrogen levels | [rs550057](https://www.ebi.ac.uk/gwas/variants/rs550057) Alpha-(1,3)-fucosyltransferase 10 levels Asthma or COVID-19 (pleiotropy) Basal Cell Adhesion Molecule levels Beta-1,4-galactosyltransferase 2 levels Brain morphology (MOSTest) COVID-19 with respiratory failure requiring mechanical ventilation Calcium levels Carbohydrate sulfotransferase 12 levels Carbohydrate sulfotransferase 15 levels Carbohydrate sulfotransferase 15 levels (CHST15.4469.78.2) Cytokine network levels (multivariate analysis) Estimated glomerular filtration rate Estimated glomerular filtration rate (creatinine) Golgi membrane protein 1 levels Gut microbiota abundance (k_Bacteria.p_Actinobacteria.c_Actinobacteria.o_Coriobacteriales) Gut microbiota abundance (k_Bacteria.p_Actinobacteria.c_Actinobacteria.o_Coriobacteriales.f_Coriobacteriaceae) Gut microbiota abundance (k_Bacteria.p_Actinobacteria.c_Actinobacteria.o_Coriobacteriales.f_Coriobacteriaceae.g_Collinsella) Gut microbiota abundance (k_Bacteria.p_Actinobacteria.c_Actinobacteria.o_Coriobacteriales.f_Coriobacteriaceae.g_Collinsella.s_Collinsella_aerofaciens) Hematocrit Hemoglobin concentration Inflammatory biomarkers (multivariate analysis) Intestinal-type alkaline phosphatase levels Intestinal-type alkaline phosphatase levels (ALPI.10463.23.3) LDL cholesterol LDL cholesterol levels Low density lipoprotein cholesterol levels Low density lipoprotein cholesterol levels *(EA)* Low density lipoprotein cholesterol levels *(Hispanic)* Lung function (FVC) Pancreatic alpha-amylase levels Pancreatic triacylglycerol lipase levels Peak expiratory flow Protein CASC4 levels Protein FAM177A1 levels Protein FAM177A1 levels (FAM177A1.8039.41.3) Protein FAM3B levels Protein FAM3D levels Protein jagged-1 levels (JAG1.5092.51.3) Red blood cell count Scavenger receptor class F member 2 levels Secreted and transmembrane protein 1 levels Serum creatinine levels Serum levels of protein C1GALT1C1 Serum levels of protein CASC4 Serum levels of protein CHST15 Serum levels of protein FAM177A1 Serum levels of protein FAM3D Serum levels of protein GOLM1 Serum levels of protein ISLR2 Serum levels of protein QSOX2 Serum levels of protein TIE1 Total cholesterol levels Total cholesterol levels *(EA)* Total cholesterol levels *(Hispanic)* Tumor necrosis factor ligand superfamily member 8 levels Two-hour glucose Tyrosine-protein kinase receptor Tie-1, soluble levels X-26054 levels [rs9411378](https://www.ebi.ac.uk/gwas/variants/rs9411378) C-C motif chemokine 25 levels (CCL25.2705.5.2) COVID-19 (covid vs negative) Calcium levels Coronary artery disease Gamma glutamyl transferase levels Low density lipoprotein cholesterol levels N-acetyl neuraminic acid levels Red cell distribution width Serum levels of protein CHST15 Serum levels of protein GNS Total cholesterol levels |
| 12:345369 | rs2080403 | 3-Aminoisobutyrate | [rs11613331](https://www.ebi.ac.uk/gwas/variants/rs11613331) Urinary metabolite levels in chronic kidney disease *(3-aminoisobutyrate)* Urine 3-aminoisobutyrate levels in chronic kidney disease | [rs11613331](https://www.ebi.ac.uk/gwas/variants/rs11613331) Serum metabolite levels *(3-aminoisobutyrate)* Serum metabolite levels *(imidazole lactate)* Serum metabolite levels *(pyroglutamine*)* | [rs11062167](https://www.ebi.ac.uk/gwas/variants/rs11062167) Estimated glomerular filtration rate Glomerular filtration rate (creatinine) [rs11613331](https://www.ebi.ac.uk/gwas/variants/rs11613331) 1-ribosyl-imidazoleacetate levels 3-hydroxybutyrate + 3-aminoisobutyrate levels Beta-aminoisobutyric acid levels *(Hispanic)* Deoxycarnitine levels in elite athletes Height Imidazole propionate levels Metabolite levels *(beta aminoisobutyric acid)* Pyroglutamine levels [rs2080402](https://www.ebi.ac.uk/gwas/variants/rs2080402) 1-ribosyl-imidazoleacetate levels Photoreceptor cell layer thickness phenotypes (MTAG) X-25420 levels [rs3782860](https://www.ebi.ac.uk/gwas/variants/rs3782860) Chronotype [rs7969761](https://www.ebi.ac.uk/gwas/variants/rs7969761) 3-aminoisobutyrate levels Betaine-to-pyroglutamine ratio Estimated glomerular filtration rate in non-diabetics Guanidinoacetate levels Imidazole lactate levels Imidazole propionate levels Plasma 3-aminoisobutyrate levels in chronic kidney disease |
| 12:4521511 | rs78470967 | 4-Deoxythreonate |  |  | [rs78470967](https://www.ebi.ac.uk/gwas/variants/rs78470967) Height Type 2 diabetes |
| 12:122344302 | rs1795967 | 2-Hydroxyisobutyrate | [rs10840627](https://www.ebi.ac.uk/gwas/variants/rs10840627) Urine caffeate levels in chronic kidney disease [rs11043222](https://www.ebi.ac.uk/gwas/variants/rs11043222) Urine alpha-hydroxyisovalerate levels in chronic kidney disease [rs1678967](https://www.ebi.ac.uk/gwas/variants/rs1678967) Urine 2-hydroxybutyrate/2-hydroxyisobutyrate levels in chronic kidney disease [rs2247139](https://www.ebi.ac.uk/gwas/variants/rs2247139) Urine 2-hydroxyphenylacetate levels in chronic kidney disease [rs2688412](https://www.ebi.ac.uk/gwas/variants/rs2688412) Urinary metabolite levels in chronic kidney disease *(4-hydroxyphenylpyruvate)* Urine 4-hydroxyphenylpyruvate levels in chronic kidney disease [rs4760099](https://www.ebi.ac.uk/gwas/variants/rs4760099) Urinary metabolites *(2-hydroxyisobutyrate)* [rs7314056](https://www.ebi.ac.uk/gwas/variants/rs7314056) Urinary metabolites (H-NMR features) *(1.3625, 2-Hydroxyisobutyrate)* [rs830124](https://www.ebi.ac.uk/gwas/variants/rs830124) Urinary metabolites *(2-Hydroxyisobutyrate concentration)* | [rs11614623](https://www.ebi.ac.uk/gwas/variants/rs11614623) Serum metabolite levels *(2-hydroxyphenylacetate)* [rs1168674](https://www.ebi.ac.uk/gwas/variants/rs1168674) Serum metabolite levels *(beta-hydroxyisovalerate)* [rs1720067](https://www.ebi.ac.uk/gwas/variants/rs1720067) Serum metabolite levels *(4-hydroxyphenylpyruvate)* | [rs10840627](https://www.ebi.ac.uk/gwas/variants/rs10840627) Beta-hydroxyisovalerate levels [rs11502392](https://www.ebi.ac.uk/gwas/variants/rs11502392) 2-hydroxyphenylacetate levels [rs1154510](https://www.ebi.ac.uk/gwas/variants/rs1154510) 4-hydroxyphenylpyruvate levels Beta-hydroxyisovalerate levels [rs1154515](https://www.ebi.ac.uk/gwas/variants/rs1154515) 4-hydroxyphenylpyruvate levels [rs11614623](https://www.ebi.ac.uk/gwas/variants/rs11614623) 2-hydroxyphenylacetate levels [rs1168674](https://www.ebi.ac.uk/gwas/variants/rs1168674) Plasma beta-hydroxyisovalerate levels in chronic kidney disease [rs1169075](https://www.ebi.ac.uk/gwas/variants/rs1169075) Tyrosine levels [rs1667583](https://www.ebi.ac.uk/gwas/variants/rs1667583) Total cholesterol levels [rs1667584](https://www.ebi.ac.uk/gwas/variants/rs1667584) 4-hydroxyphenylpyruvate levels [rs2247139](https://www.ebi.ac.uk/gwas/variants/rs2247139) Low density lipoprotein cholesterol levels [rs493519](https://www.ebi.ac.uk/gwas/variants/rs493519) 2-hydroxyisobutyrate levels [rs7314056](https://www.ebi.ac.uk/gwas/variants/rs7314056) Plasma 2-hydroxyphenylacetate levels in chronic kidney disease |
| 16:20557634 | rs7499358 | 3-Hydroxyisovalerate |  | [rs7499271](https://www.ebi.ac.uk/gwas/variants/rs7499271) Serum metabolite levels *(Phenylacetate)* Serum metabolite levels *(phenylacetate)* Serum metabolite levels *(picolinate)* | [rs58395451](https://www.ebi.ac.uk/gwas/variants/rs58395451) Beta-hydroxyisovalerate levels [rs7499557](https://www.ebi.ac.uk/gwas/variants/rs7499557) Plasma picolinate levels in chronic kidney disease [rs9922704](https://www.ebi.ac.uk/gwas/variants/rs9922704) X-17676 levels |
| 16:20608891 | rs540815683 | 3-Hydroxyisovalerate |  |  | [rs185603444](https://www.ebi.ac.uk/gwas/variants/rs185603444) X-17676 levels [rs192218016](https://www.ebi.ac.uk/gwas/variants/rs192218016) Beta-hydroxyisovalerate levels [rs193030024](https://www.ebi.ac.uk/gwas/variants/rs193030024) Salicylate levels |
| 17:26824156 | rs11567842 | Citrate |  | [rs11567842](https://www.ebi.ac.uk/gwas/variants/rs11567842) Blood urea nitrogen levels |  |
| 17:37631883 | rs11078902 | Threonine | [rs7219014](https://www.ebi.ac.uk/gwas/variants/rs7219014) Urinary metabolites *(histidine/τ-methylhistidine)* [rs7220650](https://www.ebi.ac.uk/gwas/variants/rs7220650) Urine tryptophan levels in chronic kidney disease Urine tyrosine levels in chronic kidney disease [rs755500](https://www.ebi.ac.uk/gwas/variants/rs755500) Urine histidine levels in chronic kidney disease |  | [rs10432031](https://www.ebi.ac.uk/gwas/variants/rs10432031) Sulfatide (d18:1/16:0) levels [rs10491128](https://www.ebi.ac.uk/gwas/variants/rs10491128) Estimated glomerular filtration rate (creatinine) Estimated glomerular filtration rate in non-diabetics [rs10491129](https://www.ebi.ac.uk/gwas/variants/rs10491129) Mean reticulocyte volume [rs1077715](https://www.ebi.ac.uk/gwas/variants/rs1077715) Coronary artery disease [rs11078893](https://www.ebi.ac.uk/gwas/variants/rs11078893) Estimated glomerular filtration rate [rs11078901](https://www.ebi.ac.uk/gwas/variants/rs11078901) Serum creatinine levels [rs11078916](https://www.ebi.ac.uk/gwas/variants/rs11078916) Type 2 diabetes [rs113414512](https://www.ebi.ac.uk/gwas/variants/rs113414512) Creatinine levels (UKB data field 23478) [rs113825099](https://www.ebi.ac.uk/gwas/variants/rs113825099) Drinks per week [rs12451586](https://www.ebi.ac.uk/gwas/variants/rs12451586) Glomerular filtration rate (creatinine) [rs12453397](https://www.ebi.ac.uk/gwas/variants/rs12453397) Drinks per week [rs12600751](https://www.ebi.ac.uk/gwas/variants/rs12600751) Asthma (childhood onset) Educational attainment (years of education) *(conditional-joint)* [rs12942352](https://www.ebi.ac.uk/gwas/variants/rs12942352) 1-methylhistidine levels Methionine sulfone levels Sulfatide (d18:1/16:0) levels [rs12947281](https://www.ebi.ac.uk/gwas/variants/rs12947281) Mean platelet volume [rs1619021](https://www.ebi.ac.uk/gwas/variants/rs1619021) Red blood cell count [rs2100651](https://www.ebi.ac.uk/gwas/variants/rs2100651) Mean platelet volume [rs2168785](https://www.ebi.ac.uk/gwas/variants/rs2168785) Serum creatinine levels [rs34473775](https://www.ebi.ac.uk/gwas/variants/rs34473775) Brain morphology (MOSTest) [rs35497503](https://www.ebi.ac.uk/gwas/variants/rs35497503) Refractive error [rs3964723](https://www.ebi.ac.uk/gwas/variants/rs3964723) Platelet count [rs4239222](https://www.ebi.ac.uk/gwas/variants/rs4239222) Hemoglobin concentration [rs4390625](https://www.ebi.ac.uk/gwas/variants/rs4390625) Creatinine levels [rs4794813](https://www.ebi.ac.uk/gwas/variants/rs4794813) Estimated glomerular filtration rate Estimated glomerular filtration rate (creatinine) [rs4794814](https://www.ebi.ac.uk/gwas/variants/rs4794814) Estimated glomerular filtration rate [rs4795364](https://www.ebi.ac.uk/gwas/variants/rs4795364) Spherical equivalent or myopia (age of diagnosis) [rs4795384](https://www.ebi.ac.uk/gwas/variants/rs4795384) Estimated glomerular filtration rate Whole brain restricted directional diffusion (multivariate analysis) [rs4795386](https://www.ebi.ac.uk/gwas/variants/rs4795386) Free cholesterol to total lipids in very large HDL percentage (UKB data field 23632) [rs55722796](https://www.ebi.ac.uk/gwas/variants/rs55722796) Drinks per week Estimated glomerular filtration rate (creatinine) *(conditioned on rs35289683, rs533075, rs114777282, rs1031459)* [rs6503506](https://www.ebi.ac.uk/gwas/variants/rs6503506) Calcium levels [rs677888](https://www.ebi.ac.uk/gwas/variants/rs677888) Microalbuminuria Urinary albumin-to-creatinine ratio [rs7212715](https://www.ebi.ac.uk/gwas/variants/rs7212715) Creatinine levels Glomerular filtration rate [rs755500](https://www.ebi.ac.uk/gwas/variants/rs755500) Red blood cell count [rs8069451](https://www.ebi.ac.uk/gwas/variants/rs8069451) Asthma and cardiovascular disease Childhood asthma with severe exacerbations FEV1 Lung function (FVC) Smoking initiation [rs8072297](https://www.ebi.ac.uk/gwas/variants/rs8072297) Creatinine levels [rs8076546](https://www.ebi.ac.uk/gwas/variants/rs8076546) Mean corpuscular hemoglobin [rs9747342](https://www.ebi.ac.uk/gwas/variants/rs9747342) Drinks per week [rs9892055](https://www.ebi.ac.uk/gwas/variants/rs9892055) Systemic lupus erythematosus (MTAG) |
| 17:37633970 | rs12453397 | Tryptophan, Tyrosine | [rs7219014](https://www.ebi.ac.uk/gwas/variants/rs7219014) Urinary metabolites *(histidine/τ-methylhistidine)* [rs7220650](https://www.ebi.ac.uk/gwas/variants/rs7220650) Urine tryptophan levels in chronic kidney disease Urine tyrosine levels in chronic kidney disease [rs755500](https://www.ebi.ac.uk/gwas/variants/rs755500) Urine histidine levels in chronic kidney disease |  | [rs10432031](https://www.ebi.ac.uk/gwas/variants/rs10432031) Sulfatide (d18:1/16:0) levels [rs10491128](https://www.ebi.ac.uk/gwas/variants/rs10491128) Estimated glomerular filtration rate (creatinine) Estimated glomerular filtration rate in non-diabetics [rs10491129](https://www.ebi.ac.uk/gwas/variants/rs10491129) Mean reticulocyte volume [rs1077715](https://www.ebi.ac.uk/gwas/variants/rs1077715) Coronary artery disease [rs11078893](https://www.ebi.ac.uk/gwas/variants/rs11078893) Estimated glomerular filtration rate [rs11078901](https://www.ebi.ac.uk/gwas/variants/rs11078901) Serum creatinine levels [rs11078916](https://www.ebi.ac.uk/gwas/variants/rs11078916) Type 2 diabetes [rs113414512](https://www.ebi.ac.uk/gwas/variants/rs113414512) Creatinine levels (UKB data field 23478) [rs113825099](https://www.ebi.ac.uk/gwas/variants/rs113825099) Drinks per week [rs12451586](https://www.ebi.ac.uk/gwas/variants/rs12451586) Glomerular filtration rate (creatinine) [rs12453397](https://www.ebi.ac.uk/gwas/variants/rs12453397) Drinks per week [rs12600751](https://www.ebi.ac.uk/gwas/variants/rs12600751) Asthma (childhood onset) Educational attainment (years of education) *(conditional-joint)* [rs12942352](https://www.ebi.ac.uk/gwas/variants/rs12942352) 1-methylhistidine levels Methionine sulfone levels Sulfatide (d18:1/16:0) levels [rs12947281](https://www.ebi.ac.uk/gwas/variants/rs12947281) Mean platelet volume [rs1619021](https://www.ebi.ac.uk/gwas/variants/rs1619021) Red blood cell count [rs2100651](https://www.ebi.ac.uk/gwas/variants/rs2100651) Mean platelet volume [rs2168785](https://www.ebi.ac.uk/gwas/variants/rs2168785) Serum creatinine levels [rs34473775](https://www.ebi.ac.uk/gwas/variants/rs34473775) Brain morphology (MOSTest) [rs35497503](https://www.ebi.ac.uk/gwas/variants/rs35497503) Refractive error [rs3964723](https://www.ebi.ac.uk/gwas/variants/rs3964723) Platelet count [rs4239222](https://www.ebi.ac.uk/gwas/variants/rs4239222) Hemoglobin concentration [rs4390625](https://www.ebi.ac.uk/gwas/variants/rs4390625) Creatinine levels [rs4794813](https://www.ebi.ac.uk/gwas/variants/rs4794813) Estimated glomerular filtration rate Estimated glomerular filtration rate (creatinine) [rs4794814](https://www.ebi.ac.uk/gwas/variants/rs4794814) Estimated glomerular filtration rate [rs4795364](https://www.ebi.ac.uk/gwas/variants/rs4795364) Spherical equivalent or myopia (age of diagnosis) [rs4795384](https://www.ebi.ac.uk/gwas/variants/rs4795384) Estimated glomerular filtration rate Whole brain restricted directional diffusion (multivariate analysis) [rs4795386](https://www.ebi.ac.uk/gwas/variants/rs4795386) Free cholesterol to total lipids in very large HDL percentage (UKB data field 23632) [rs55722796](https://www.ebi.ac.uk/gwas/variants/rs55722796) Drinks per week Estimated glomerular filtration rate (creatinine) *(conditioned on rs35289683, rs533075, rs114777282, rs1031459)* [rs6503506](https://www.ebi.ac.uk/gwas/variants/rs6503506) Calcium levels [rs677888](https://www.ebi.ac.uk/gwas/variants/rs677888) Microalbuminuria Urinary albumin-to-creatinine ratio [rs7212715](https://www.ebi.ac.uk/gwas/variants/rs7212715) Creatinine levels Glomerular filtration rate [rs755500](https://www.ebi.ac.uk/gwas/variants/rs755500) Red blood cell count [rs8069451](https://www.ebi.ac.uk/gwas/variants/rs8069451) Asthma and cardiovascular disease Childhood asthma with severe exacerbations FEV1 Lung function (FVC) Smoking initiation [rs8072297](https://www.ebi.ac.uk/gwas/variants/rs8072297) Creatinine levels [rs8076546](https://www.ebi.ac.uk/gwas/variants/rs8076546) Mean corpuscular hemoglobin [rs9747342](https://www.ebi.ac.uk/gwas/variants/rs9747342) Drinks per week [rs9892055](https://www.ebi.ac.uk/gwas/variants/rs9892055) Systemic lupus erythematosus (MTAG) |
| 17:37636695 | rs4795371 | Histidine | [rs7219014](https://www.ebi.ac.uk/gwas/variants/rs7219014) Urinary metabolites *(histidine/τ-methylhistidine)* [rs7220650](https://www.ebi.ac.uk/gwas/variants/rs7220650) Urine tryptophan levels in chronic kidney disease Urine tyrosine levels in chronic kidney disease [rs755500](https://www.ebi.ac.uk/gwas/variants/rs755500) Urine histidine levels in chronic kidney disease |  | [rs10432031](https://www.ebi.ac.uk/gwas/variants/rs10432031) Sulfatide (d18:1/16:0) levels [rs10491128](https://www.ebi.ac.uk/gwas/variants/rs10491128) Estimated glomerular filtration rate (creatinine) Estimated glomerular filtration rate in non-diabetics [rs10491129](https://www.ebi.ac.uk/gwas/variants/rs10491129) Mean reticulocyte volume [rs1077715](https://www.ebi.ac.uk/gwas/variants/rs1077715) Coronary artery disease [rs11078893](https://www.ebi.ac.uk/gwas/variants/rs11078893) Estimated glomerular filtration rate [rs11078901](https://www.ebi.ac.uk/gwas/variants/rs11078901) Serum creatinine levels [rs11078916](https://www.ebi.ac.uk/gwas/variants/rs11078916) Type 2 diabetes [rs113414512](https://www.ebi.ac.uk/gwas/variants/rs113414512) Creatinine levels (UKB data field 23478) [rs113825099](https://www.ebi.ac.uk/gwas/variants/rs113825099) Drinks per week [rs12451586](https://www.ebi.ac.uk/gwas/variants/rs12451586) Glomerular filtration rate (creatinine) [rs12453397](https://www.ebi.ac.uk/gwas/variants/rs12453397) Drinks per week [rs12600751](https://www.ebi.ac.uk/gwas/variants/rs12600751) Asthma (childhood onset) Educational attainment (years of education) *(conditional-joint)* [rs12942352](https://www.ebi.ac.uk/gwas/variants/rs12942352) 1-methylhistidine levels Methionine sulfone levels Sulfatide (d18:1/16:0) levels [rs12947281](https://www.ebi.ac.uk/gwas/variants/rs12947281) Mean platelet volume [rs1619021](https://www.ebi.ac.uk/gwas/variants/rs1619021) Red blood cell count [rs2100651](https://www.ebi.ac.uk/gwas/variants/rs2100651) Mean platelet volume [rs2168785](https://www.ebi.ac.uk/gwas/variants/rs2168785) Serum creatinine levels [rs34473775](https://www.ebi.ac.uk/gwas/variants/rs34473775) Brain morphology (MOSTest) [rs35497503](https://www.ebi.ac.uk/gwas/variants/rs35497503) Refractive error [rs3964723](https://www.ebi.ac.uk/gwas/variants/rs3964723) Platelet count [rs4239222](https://www.ebi.ac.uk/gwas/variants/rs4239222) Hemoglobin concentration [rs4390625](https://www.ebi.ac.uk/gwas/variants/rs4390625) Creatinine levels [rs4794813](https://www.ebi.ac.uk/gwas/variants/rs4794813) Estimated glomerular filtration rate Estimated glomerular filtration rate (creatinine) [rs4794814](https://www.ebi.ac.uk/gwas/variants/rs4794814) Estimated glomerular filtration rate [rs4795364](https://www.ebi.ac.uk/gwas/variants/rs4795364) Spherical equivalent or myopia (age of diagnosis) [rs4795384](https://www.ebi.ac.uk/gwas/variants/rs4795384) Estimated glomerular filtration rate Whole brain restricted directional diffusion (multivariate analysis) [rs4795386](https://www.ebi.ac.uk/gwas/variants/rs4795386) Free cholesterol to total lipids in very large HDL percentage (UKB data field 23632) [rs55722796](https://www.ebi.ac.uk/gwas/variants/rs55722796) Drinks per week Estimated glomerular filtration rate (creatinine) *(conditioned on rs35289683, rs533075, rs114777282, rs1031459)* [rs6503506](https://www.ebi.ac.uk/gwas/variants/rs6503506) Calcium levels [rs677888](https://www.ebi.ac.uk/gwas/variants/rs677888) Microalbuminuria Urinary albumin-to-creatinine ratio [rs7212715](https://www.ebi.ac.uk/gwas/variants/rs7212715) Creatinine levels Glomerular filtration rate [rs755500](https://www.ebi.ac.uk/gwas/variants/rs755500) Red blood cell count [rs8069451](https://www.ebi.ac.uk/gwas/variants/rs8069451) Asthma and cardiovascular disease Childhood asthma with severe exacerbations FEV1 Lung function (FVC) Smoking initiation [rs8072297](https://www.ebi.ac.uk/gwas/variants/rs8072297) Creatinine levels [rs8076546](https://www.ebi.ac.uk/gwas/variants/rs8076546) Mean corpuscular hemoglobin [rs9747342](https://www.ebi.ac.uk/gwas/variants/rs9747342) Drinks per week [rs9892055](https://www.ebi.ac.uk/gwas/variants/rs9892055) Systemic lupus erythematosus (MTAG) |

### Supplementary Table 7

**Blood eQTLs at the COJO lead variants.** Expression quantitative trait loci (eQTL) target genes in whole blood (p < 5.5×10^-05^) at the COJO lead signals.

| **Chr:pos** | **Rsid** | **Metabolite** | **eQTL Blood** |
| --- | --- | --- | --- |
| 1:6334301 | rs114200864 | 3-Hydroxyisovalerate | [ACOT7](https://www.genecards.org/cgi-bin/carddisp.pl?gene=ENSG00000097021) (2.5×10^-65^), [ZBTB48](https://www.genecards.org/cgi-bin/carddisp.pl?gene=ENSG00000204859) (2.4×10^-15^), [ESPN](https://www.genecards.org/cgi-bin/carddisp.pl?gene=ENSG00000187017) (2.8×10^-15^), [RNF207](https://www.genecards.org/cgi-bin/carddisp.pl?gene=ENSG00000158286) (3.5×10^-07^), [CHD5](https://www.genecards.org/cgi-bin/carddisp.pl?gene=ENSG00000116254) (6.6×10^-06^) |
| 1:11940483 | rs4846068 | Formate | [PLOD1](https://www.genecards.org/cgi-bin/carddisp.pl?gene=ENSG00000083444) (2.3×10^-132^), [MFN2](https://www.genecards.org/cgi-bin/carddisp.pl?gene=ENSG00000116688) (3.9×10^-123^), [KIAA2013](https://www.genecards.org/cgi-bin/carddisp.pl?gene=ENSG00000116685) (2.1×10^-100^), [MTHFR](https://www.genecards.org/cgi-bin/carddisp.pl?gene=ENSG00000177000) (1.8×10^-60^), [NPPA-AS1](https://www.genecards.org/cgi-bin/carddisp.pl?gene=ENSG00000242349) (2.6×10^-31^), [NPPA](https://www.genecards.org/cgi-bin/carddisp.pl?gene=ENSG00000175206) (6.2×10^-13^), [CLCN6](https://www.genecards.org/cgi-bin/carddisp.pl?gene=ENSG00000011021) (3.0×10^-07^) |
| 1:151904146 | rs2999545 | 3-Hydroxyisovalerate | [THEM4](https://www.genecards.org/cgi-bin/carddisp.pl?gene=ENSG00000159445) (3.3×10^-302^), [S100A10](https://www.genecards.org/cgi-bin/carddisp.pl?gene=ENSG00000197747) (1.2×10^-12^), [S100A11](https://www.genecards.org/cgi-bin/carddisp.pl?gene=ENSG00000163191) (1.4×10^-07^), [LINGO4](https://www.genecards.org/cgi-bin/carddisp.pl?gene=ENSG00000213171) (6.2×10^-07^) |
| 2:135598913 | rs17322446 | 1-Methylnicotinamide | [CCNT2](https://www.genecards.org/cgi-bin/carddisp.pl?gene=ENSG00000082258) (8.6×10^-287^), [AC011893.3](https://www.genecards.org/cgi-bin/carddisp.pl?gene=ENSG00000226806) (6.9×10^-30^), [AC016725.4](https://www.genecards.org/cgi-bin/carddisp.pl?gene=ENSG00000224043) (8.1×10^-18^), [TMEM163](https://www.genecards.org/cgi-bin/carddisp.pl?gene=ENSG00000152128) (4.7×10^-17^) |
| 2:241793545 | rs55649245 | 4-Deoxyerythronic acid | [ANKMY1](https://www.genecards.org/cgi-bin/carddisp.pl?gene=ENSG00000144504) (3.7×10^-13^) |
| 2:241813788 | rs10933641 | 4-Deoxyerythronic acid | [GPR35](https://www.genecards.org/cgi-bin/carddisp.pl?gene=ENSG00000178623) (3.5×10^-07^), [AC104809.4](https://www.genecards.org/cgi-bin/carddisp.pl?gene=ENSG00000233392) (7.0×10^-07^) |
| 3:182758040 | rs4859267 | 3-Hydroxyisovalerate | [MCCC1](https://www.genecards.org/cgi-bin/carddisp.pl?gene=ENSG00000078070) (1.6×10^-95^), [ATP11B](https://www.genecards.org/cgi-bin/carddisp.pl?gene=ENSG00000058063) (3.2×10^-10^) |
| 4:88213884 | rs6811902 | Propylene Glycol | [HSD17B13](https://www.genecards.org/cgi-bin/carddisp.pl?gene=ENSG00000170509) (7.8×10^-197^), [HSD17B11](https://www.genecards.org/cgi-bin/carddisp.pl?gene=ENSG00000198189) (3.9×10^-163^), [KLHL8](https://www.genecards.org/cgi-bin/carddisp.pl?gene=ENSG00000145332) (8.5×10^-21^), [NUDT9](https://www.genecards.org/cgi-bin/carddisp.pl?gene=ENSG00000170502) (1.9×10^-05^) |
| 5:34993215 | rs11744796 | 3-Aminoisobutyrate | [DNAJC21](https://www.genecards.org/cgi-bin/carddisp.pl?gene=ENSG00000168724) (1.1×10^-86^) |
| 5:150624099 | rs72794144 | Glycine | [GM2A](https://www.genecards.org/cgi-bin/carddisp.pl?gene=ENSG00000196743) (9.1×10^-19^), [ANXA6](https://www.genecards.org/cgi-bin/carddisp.pl?gene=ENSG00000197043) (2.9×10^-09^), [SLC36A1](https://www.genecards.org/cgi-bin/carddisp.pl?gene=ENSG00000123643) (6.1×10^-08^) |
| 5:150702299 | rs61067578 | Glycine | [GM2A](https://www.genecards.org/cgi-bin/carddisp.pl?gene=ENSG00000196743) (1.6×10^-19^), [SLC36A1](https://www.genecards.org/cgi-bin/carddisp.pl?gene=ENSG00000123643) (1.4×10^-07^) |
| 5:150708711 | rs147000073 | Glycine | [GM2A](https://www.genecards.org/cgi-bin/carddisp.pl?gene=ENSG00000196743) (3.5×10^-07^) |
| 6:111492119 | rs241768 | Tyrosine | [C6orf3](https://www.genecards.org/cgi-bin/carddisp.pl?gene=ENSG00000255389) (2.4×10^-10^), [TRAF3IP2](https://www.genecards.org/cgi-bin/carddisp.pl?gene=ENSG00000056972) (3.0×10^-10^), [FYN](https://www.genecards.org/cgi-bin/carddisp.pl?gene=ENSG00000010810) (4.9×10^-05^) |
| 7:17287998 | rs2106727 | Quinic acid, Trigonelline | [AHR](https://www.genecards.org/cgi-bin/carddisp.pl?gene=ENSG00000106546) (3.9×10^-09^) |
| 8:74868909 | rs72661850 | 4-Deoxyerythronic acid | [TMEM70](https://www.genecards.org/cgi-bin/carddisp.pl?gene=ENSG00000175606) (2.5×10^-25^), [LY96](https://www.genecards.org/cgi-bin/carddisp.pl?gene=ENSG00000154589) (1.8×10^-15^) |
| 9:6649491 | rs62565993 | Glycine | [GLDC](https://www.genecards.org/cgi-bin/carddisp.pl?gene=ENSG00000178445) (2.4×10^-79^), [RP11-390F4.3](https://www.genecards.org/cgi-bin/carddisp.pl?gene=ENSG00000225489) (1.0×10^-06^) |
| 9:107525165 | rs2472479 | 3-Hydroxyisobutyrate | [NIPSNAP3A](https://www.genecards.org/cgi-bin/carddisp.pl?gene=ENSG00000136783) (3.3×10^-302^), [NIPSNAP3B](https://www.genecards.org/cgi-bin/carddisp.pl?gene=ENSG00000165028) (7.7×10^-24^) |
| 9:136146597 | rs550057 | Xylose | [ABO](https://www.genecards.org/cgi-bin/carddisp.pl?gene=ENSG00000175164) (3.3×10^-302^), [GBGT1](https://www.genecards.org/cgi-bin/carddisp.pl?gene=ENSG00000148288) (3.6×10^-22^), [SURF6](https://www.genecards.org/cgi-bin/carddisp.pl?gene=ENSG00000148296) (7.3×10^-06^), [GTF3C5](https://www.genecards.org/cgi-bin/carddisp.pl?gene=ENSG00000148308) (4.6×10^-05^) |
| 12:345369 | rs2080403 | 3-Aminoisobutyrate | [CCDC77](https://www.genecards.org/cgi-bin/carddisp.pl?gene=ENSG00000120647) (3.3×10^-302^), [KDM5A](https://www.genecards.org/cgi-bin/carddisp.pl?gene=ENSG00000073614) (2.1×10^-162^), [RP11-283I3.6](https://www.genecards.org/cgi-bin/carddisp.pl?gene=ENSG00000261799) (4.1×10^-35^), [SLC6A12](https://www.genecards.org/cgi-bin/carddisp.pl?gene=ENSG00000111181) (2.6×10^-09^), [IQSEC3](https://www.genecards.org/cgi-bin/carddisp.pl?gene=ENSG00000120645) (2.3×10^-08^) |
| 12:122344302 | rs1795967 | 2-Hydroxyisobutyrate | [AC084018.1](https://www.genecards.org/cgi-bin/carddisp.pl?gene=ENSG00000212694) (2.4×10^-14^), [PSMD9](https://www.genecards.org/cgi-bin/carddisp.pl?gene=ENSG00000110801) (3.4×10^-08^), [BCL7A](https://www.genecards.org/cgi-bin/carddisp.pl?gene=ENSG00000110987) (3.0×10^-07^) |
| 16:20557634 | rs7499358 | 3-Hydroxyisovalerate | [ACSM1](https://www.genecards.org/cgi-bin/carddisp.pl?gene=ENSG00000166743) (3.1×10^-11^) |
| 17:26824156 | rs11567842 | Citrate | [ALDOC](https://www.genecards.org/cgi-bin/carddisp.pl?gene=ENSG00000109107) (8.5×10^-12^), [RAB34](https://www.genecards.org/cgi-bin/carddisp.pl?gene=ENSG00000109113) (1.1×10^-05^) |
| 17:37631883 | rs11078902 | Threonine | [GSDMB](https://www.genecards.org/cgi-bin/carddisp.pl?gene=ENSG00000073605) (3.3×10^-302^), [ORMDL3](https://www.genecards.org/cgi-bin/carddisp.pl?gene=ENSG00000172057) (2.0×10^-254^), [PGAP3](https://www.genecards.org/cgi-bin/carddisp.pl?gene=ENSG00000161395) (1.2×10^-163^), [CDK12](https://www.genecards.org/cgi-bin/carddisp.pl?gene=ENSG00000167258) (1.4×10^-101^), [MED1](https://www.genecards.org/cgi-bin/carddisp.pl?gene=ENSG00000125686) (3.3×10^-71^), [IKZF3](https://www.genecards.org/cgi-bin/carddisp.pl?gene=ENSG00000161405) (3.5×10^-56^), [FBXL20](https://www.genecards.org/cgi-bin/carddisp.pl?gene=ENSG00000108306) (4.6×10^-35^), [PSMD3](https://www.genecards.org/cgi-bin/carddisp.pl?gene=ENSG00000108344) (5.4×10^-18^), [PPP1R1B](https://www.genecards.org/cgi-bin/carddisp.pl?gene=ENSG00000131771) (4.7×10^-16^), [CTB-131K11.1](https://www.genecards.org/cgi-bin/carddisp.pl?gene=ENSG00000266469) (1.8×10^-12^), [STARD3](https://www.genecards.org/cgi-bin/carddisp.pl?gene=ENSG00000131748) (3.8×10^-10^), [ERBB2](https://www.genecards.org/cgi-bin/carddisp.pl?gene=ENSG00000141736) (4.0×10^-07^), [GSDMA](https://www.genecards.org/cgi-bin/carddisp.pl?gene=ENSG00000167914) (4.8×10^-07^), [RP11-94L15.2](https://www.genecards.org/cgi-bin/carddisp.pl?gene=ENSG00000264198) (7.5×10^-07^), [NR1D1](https://www.genecards.org/cgi-bin/carddisp.pl?gene=ENSG00000126368) (2.0×10^-05^) |
| 17:37633970 | rs12453397 | Tryptophan, Tyrosine | [GSDMB](https://www.genecards.org/cgi-bin/carddisp.pl?gene=ENSG00000073605) (3.3×10^-302^), [ORMDL3](https://www.genecards.org/cgi-bin/carddisp.pl?gene=ENSG00000172057) (4.9×10^-204^), [PGAP3](https://www.genecards.org/cgi-bin/carddisp.pl?gene=ENSG00000161395) (7.0×10^-135^), [CDK12](https://www.genecards.org/cgi-bin/carddisp.pl?gene=ENSG00000167258) (5.4×10^-101^), [MED1](https://www.genecards.org/cgi-bin/carddisp.pl?gene=ENSG00000125686) (1.7×10^-62^), [IKZF3](https://www.genecards.org/cgi-bin/carddisp.pl?gene=ENSG00000161405) (1.4×10^-46^), [FBXL20](https://www.genecards.org/cgi-bin/carddisp.pl?gene=ENSG00000108306) (1.1×10^-37^), [PPP1R1B](https://www.genecards.org/cgi-bin/carddisp.pl?gene=ENSG00000131771) (7.2×10^-16^), [PSMD3](https://www.genecards.org/cgi-bin/carddisp.pl?gene=ENSG00000108344) (9.4×10^-16^), [CTB-131K11.1](https://www.genecards.org/cgi-bin/carddisp.pl?gene=ENSG00000266469) (5.1×10^-09^), [STARD3](https://www.genecards.org/cgi-bin/carddisp.pl?gene=ENSG00000131748) (1.6×10^-08^), [GSDMA](https://www.genecards.org/cgi-bin/carddisp.pl?gene=ENSG00000167914) (8.0×10^-07^), [RP11-94L15.2](https://www.genecards.org/cgi-bin/carddisp.pl?gene=ENSG00000264198) (8.6×10^-06^), [ERBB2](https://www.genecards.org/cgi-bin/carddisp.pl?gene=ENSG00000141736) (1.8×10^-05^), [NR1D1](https://www.genecards.org/cgi-bin/carddisp.pl?gene=ENSG00000126368) (2.4×10^-05^) |
| 17:37636695 | rs4795371 | Histidine | [GSDMB](https://www.genecards.org/cgi-bin/carddisp.pl?gene=ENSG00000073605) (3.3×10^-302^), [ORMDL3](https://www.genecards.org/cgi-bin/carddisp.pl?gene=ENSG00000172057) (4.3×10^-251^), [PGAP3](https://www.genecards.org/cgi-bin/carddisp.pl?gene=ENSG00000161395) (5.3×10^-155^), [CDK12](https://www.genecards.org/cgi-bin/carddisp.pl?gene=ENSG00000167258) (1.5×10^-99^), [MED1](https://www.genecards.org/cgi-bin/carddisp.pl?gene=ENSG00000125686) (2.4×10^-69^), [IKZF3](https://www.genecards.org/cgi-bin/carddisp.pl?gene=ENSG00000161405) (1.7×10^-48^), [FBXL20](https://www.genecards.org/cgi-bin/carddisp.pl?gene=ENSG00000108306) (3.3×10^-35^), [PSMD3](https://www.genecards.org/cgi-bin/carddisp.pl?gene=ENSG00000108344) (8.8×10^-18^), [PPP1R1B](https://www.genecards.org/cgi-bin/carddisp.pl?gene=ENSG00000131771) (2.0×10^-16^), [CTB-131K11.1](https://www.genecards.org/cgi-bin/carddisp.pl?gene=ENSG00000266469) (1.1×10^-12^), [STARD3](https://www.genecards.org/cgi-bin/carddisp.pl?gene=ENSG00000131748) (1.4×10^-09^), [GSDMA](https://www.genecards.org/cgi-bin/carddisp.pl?gene=ENSG00000167914) (1.3×10^-08^), [ERBB2](https://www.genecards.org/cgi-bin/carddisp.pl?gene=ENSG00000141736) (3.8×10^-07^), [RP11-94L15.2](https://www.genecards.org/cgi-bin/carddisp.pl?gene=ENSG00000264198) (8.9×10^-07^), [NR1D1](https://www.genecards.org/cgi-bin/carddisp.pl?gene=ENSG00000126368) (7.9×10^-06^) |

### Supplementary Table 8

**Mendelian randomization analysis outcomes and exposures.** Two sample Mendelian randomization analysis outcomes (n=18) used with urinary metabolites as the exposures, and exposures (n=2) used with urinary metabolites as the outcomes.

| **Analysis** | **Source** | **Trait** | **Population** | **PMID** | **IEU GWAS database ID** |
| --- | --- | --- | --- | --- | --- |
| Metabolite -> Outcome | DNCRI | allvcntrl | European | 31537649 | – |
|  |  | ckddn | European | 31537649 | – |
|  |  | ckdextreme | European | 31537649 | – |
|  |  | ckd | European | 31537649 | – |
|  |  | DN | European | 31537649 | – |
|  |  | esrd | European | 31537649 | – |
|  |  | esrdvall | European | 31537649 | – |
|  |  | esrdvmacro | European | 31537649 | – |
|  |  | macro | European | 31537649 | – |
|  |  | micro | European | 31537649 | – |
|  | CKDGen | eGFR | European | 31152163 | – |
|  |  | CKD | European | 31152163 | – |
|  |  | UACR | European | 31511532 | – |
|  | DIAMANTE | T2D | European | 35551307 | – |
|  | IEU GWAS database | Asthma | European | – | finn-b-J10_ASTHMA |
|  |  | Body mass index (BMI) | European | – | ukb-b-19953 |
|  |  | Coronary heart disease | Mixed | 26343387 | ieu-a-7 |
|  |  | Creatinine (enzymatic) in urine | European | – | ukb-a-333 |
| Exposure -> Metabolite | CKDGen | eGFR | European | 31152163 | – |
|  |  | UACR | European | 31511532 | – |

### Supplementary Table 9

**Mendelian randomization analysis of kidney trait exposure on urinary metabolites.**

Two sample Mendelian randomization analysis results using eGFR and UACR as exposures and urinary metabolites as outcomes. Exposure-metabolite pairs with p < 4.7×10^-04^ (0.05 / number of unique exposure-outcome pairs = 0.05 / 106) in at least one analysis are reported. For each analysed exposure-outcome pair all the performed analysis results are shown.

| **Outcome** | **Exposure** | **Method** | **Beta (SE)** | **P** | **N SNV** | **P_het_** |
| --- | --- | --- | --- | --- | --- | --- |
| Glycolic acid | eGFR | Inverse variance weighted | 3.71 (0.50) | **1.1×10^-13^** | 142 | 2.0×10^-01^ |
| Glycolic acid | eGFR | Weighted median | 3.15 (0.74) | **2.4×10^-05^** | 142 |  |
| Glycolic acid | eGFR | Simple mode | 6.01 (1.89) | **1.8×10^-03^** | 142 |  |
| Glycolic acid | eGFR | MR Egger | 2.76 (1.24) | **2.7×10^-02^** | 142 | 2.0×10^-01^ |
| Glycolic acid | eGFR | Weighted mode | 2.07 (1.28) | 1.1×10^-01^ | 142 |  |
| Ethanolamine | eGFR | Inverse variance weighted | 2.23 (0.44) | **3.1×10^-07^** | 142 | 4.9×10^-01^ |
| Ethanolamine | eGFR | Weighted median | 2.70 (0.70) | **1.2×10^-04^** | 142 |  |
| Ethanolamine | eGFR | Weighted mode | 2.97 (1.12) | **8.9×10^-03^** | 142 |  |
| Ethanolamine | eGFR | Simple mode | 2.97 (1.61) | 6.7×10^-02^ | 142 |  |
| Ethanolamine | eGFR | MR Egger | 0.69 (1.08) | 5.2×10^-01^ | 142 | 5.2×10^-01^ |
| Valine | eGFR | Inverse variance weighted | 2.59 (0.52) | **6.2×10^-07^** | 143 | **6.1×10^-04^** |
| Valine | eGFR | Weighted median | 2.56 (0.71) | **3.2×10^-04^** | 143 |  |
| Valine | eGFR | Weighted mode | 2.33 (1.27) | 6.9×10^-02^ | 143 |  |
| Valine | eGFR | Simple mode | 2.52 (1.88) | 1.8×10^-01^ | 143 |  |
| Valine | eGFR | MR Egger | 0.93 (1.28) | 4.7×10^-01^ | 143 | **8.0×10^-04^** |
| Uracil | eGFR | Inverse variance weighted | 2.09 (0.43) | **9.0×10^-07^** | 143 | 4.1×10^-01^ |
| Uracil | eGFR | Weighted median | 2.46 (0.68) | **3.0×10^-04^** | 143 |  |
| Uracil | eGFR | Weighted mode | 3.08 (1.41) | **3.1×10^-02^** | 143 |  |
| Uracil | eGFR | MR Egger | 2.06 (1.05) | 5.2×10^-02^ | 143 | 3.9×10^-01^ |
| Uracil | eGFR | Simple mode | 2.88 (1.94) | 1.4×10^-01^ | 143 |  |
| Formate | eGFR | Inverse variance weighted | 2.12 (0.45) | **2.9×10^-06^** | 143 | 1.5×10^-01^ |
| Formate | eGFR | Weighted median | 2.31 (0.64) | **3.4×10^-04^** | 143 |  |
| Formate | eGFR | Weighted mode | 3.69 (1.75) | **3.7×10^-02^** | 143 |  |
| Formate | eGFR | Simple mode | 3.04 (1.81) | 9.6×10^-02^ | 143 |  |
| Formate | eGFR | MR Egger | 1.84 (1.12) | 1.0×10^-01^ | 143 | 1.4×10^-01^ |
| Leucine | eGFR | Inverse variance weighted | 2.09 (0.45) | **3.4×10^-06^** | 143 | 1.2×10^-01^ |
| Leucine | eGFR | Weighted median | 2.50 (0.68) | **2.5×10^-04^** | 143 |  |
| Leucine | eGFR | Weighted mode | 2.81 (1.19) | **1.9×10^-02^** | 143 |  |
| Leucine | eGFR | Simple mode | 3.37 (1.60) | **3.7×10^-02^** | 143 |  |
| Leucine | eGFR | MR Egger | 0.50 (1.11) | 6.5×10^-01^ | 143 | 1.4×10^-01^ |
| Glutamine | eGFR | Inverse variance weighted | 1.99 (0.49) | **4.3×10^-05^** | 143 | 1.4×10^-01^ |
| Glutamine | eGFR | Weighted median | 1.93 (0.75) | **1.0×10^-02^** | 143 |  |
| Glutamine | eGFR | MR Egger | 1.70 (1.21) | 1.6×10^-01^ | 143 | 1.3×10^-01^ |
| Glutamine | eGFR | Simple mode | 1.17 (2.20) | 5.9×10^-01^ | 143 |  |
| Glutamine | eGFR | Weighted mode | 0.14 (1.66) | 9.3×10^-01^ | 143 |  |
| 2-Hydroxyisobutyrate | eGFR | Inverse variance weighted | 1.72 (0.43) | **5.0×10^-05^** | 143 | 4.4×10^-01^ |
| 2-Hydroxyisobutyrate | eGFR | Weighted median | 1.84 (0.68) | **7.2×10^-03^** | 143 |  |
| 2-Hydroxyisobutyrate | eGFR | Weighted mode | 2.65 (1.47) | 7.5×10^-02^ | 143 |  |
| 2-Hydroxyisobutyrate | eGFR | Simple mode | 2.75 (1.74) | 1.2×10^-01^ | 143 |  |
| 2-Hydroxyisobutyrate | eGFR | MR Egger | 1.58 (1.05) | 1.4×10^-01^ | 143 | 4.2×10^-01^ |
| Alanine | eGFR | Inverse variance weighted | 2.10 (0.52) | **5.1×10^-05^** | 143 | **2.4×10^-04^** |
| Alanine | eGFR | Weighted median | 2.49 (0.69) | **3.1×10^-04^** | 143 |  |
| Alanine | eGFR | Simple mode | 3.43 (1.96) | 8.3×10^-02^ | 143 |  |
| Alanine | eGFR | Weighted mode | 2.36 (1.59) | 1.4×10^-01^ | 143 |  |
| Alanine | eGFR | MR Egger | 0.81 (1.28) | 5.3×10^-01^ | 143 | **2.6×10^-04^** |
| 3-Hydroxyisovalerate | eGFR | Weighted median | 2.79 (0.72) | **1.1×10^-04^** | 143 |  |
| 3-Hydroxyisovalerate | eGFR | Inverse variance weighted | 1.77 (0.50) | **4.3×10^-04^** | 143 | **2.6×10^-03^** |
| 3-Hydroxyisovalerate | eGFR | MR Egger | 3.40 (1.24) | **6.7×10^-03^** | 143 | **3.3×10^-03^** |
| 3-Hydroxyisovalerate | eGFR | Weighted mode | 3.50 (1.37) | **1.2×10^-02^** | 143 |  |
| 3-Hydroxyisovalerate | eGFR | Simple mode | 3.71 (1.82) | **4.3×10^-02^** | 143 |  |
| Pseudouridine | eGFR | Inverse variance weighted | 1.64 (0.44) | **2.1×10^-04^** | 143 | **2.4×10^-02^** |
| Pseudouridine | eGFR | Weighted median | 1.56 (0.61) | **1.0×10^-02^** | 143 |  |
| Pseudouridine | eGFR | MR Egger | 2.20 (1.10) | **4.6×10^-02^** | 143 | **2.2×10^-02^** |
| Pseudouridine | eGFR | Weighted mode | 2.17 (1.30) | 9.9×10^-02^ | 143 |  |
| Pseudouridine | eGFR | Simple mode | 1.29 (1.62) | 4.3×10^-01^ | 143 |  |
| Pyroglutamate | eGFR | Inverse variance weighted | 1.53 (0.43) | **3.6×10^-04^** | 143 | 7.0×10^-01^ |
| Pyroglutamate | eGFR | Weighted median | 1.19 (0.68) | 8.0×10^-02^ | 143 |  |
| Pyroglutamate | eGFR | Weighted mode | 1.21 (1.28) | 3.4×10^-01^ | 143 |  |
| Pyroglutamate | eGFR | MR Egger | 0.98 (1.06) | 3.6×10^-01^ | 143 | 6.8×10^-01^ |
| Pyroglutamate | eGFR | Simple mode | -0.69 (1.62) | 6.7×10^-01^ | 143 |  |
| Glycine | eGFR | Weighted median | 2.60 (0.73) | **3.7×10^-04^** | 143 |  |
| Glycine | eGFR | Inverse variance weighted | 1.52 (0.69) | **2.9×10^-02^** | 143 | **5.6×10^-22^** |
| Glycine | eGFR | Weighted mode | 3.19 (1.48) | **3.3×10^-02^** | 143 |  |
| Glycine | eGFR | Simple mode | 4.13 (2.23) | 6.6×10^-02^ | 143 |  |
| Glycine | eGFR | MR Egger | -0.67 (1.71) | 7.0×10^-01^ | 143 | **1.6×10^-21^** |

Outcome: the outcome trait in the MR analysis. Exposure: the exposure trait in the MR analysis. Method: the MR analysis method. Beta (SE): the effect estimate (standard-error) of the exposure on the outcome. P: p-value for the effect estimate. N SNV: the number of SNVs in the analysis. P_het_: p-value for the heterogeneity test. eGFR: estimated glomerular filtration rate.

### Supplementary Table 10

**MAGMA tissue enrichment analysis, tissues with p<0.01.**

| **Metabolite** | **Tissue** | **N genes** | **Beta** | **SE** | **P** |
| --- | --- | --- | --- | --- | --- |
| cis-Aconitate | Adipose Tissue | 17391 | 0.049 | 0.013 | 6.9E-05 |
| glycine | Kidney | 17393 | 0.033 | 0.009 | 1.5E-04 |
| Pyroglutamate | Pituitary | 17392 | 0.025 | 0.008 | 6.8E-04 |
| Allantoin | Cervix Uteri | 17390 | 0.042 | 0.013 | 0.0010 |
| 2-Furoylglycine | Testis | 17391 | 0.016 | 0.005 | 0.0014 |
| Mannitol | Small Intestine | 17387 | 0.030 | 0.010 | 0.0016 |
| Acetate | Esophagus | 17393 | 0.048 | 0.016 | 0.0018 |
| Pseudouridine | Blood Vessel | 17393 | 0.031 | 0.011 | 0.0030 |
| 3-(3-Hydroxyphenyl)-3-hydroxypropionic acid | Adipose Tissue | 17390 | 0.034 | 0.013 | 0.0042 |
| 3-(3-Hydroxyphenyl)-3-hydroxypropionic acid | Breast | 17390 | 0.039 | 0.015 | 0.0049 |
| cis-Aconitate | Blood Vessel | 17391 | 0.028 | 0.011 | 0.0057 |
| Quinic acid | Ovary | 17392 | 0.024 | 0.010 | 0.0067 |
| Allantoin | Thyroid | 17390 | 0.025 | 0.010 | 0.0071 |
| 2-Hydroxyisobutyrate | Muscle | 17393 | 0.016 | 0.007 | 0.0094 |
| Proline betaine | Blood | 17390 | 0.014 | 0.006 | 0.0094 |

### Supplementary Table 11

**MAGMA v1.6 gene set analysis for “Curated gene sets” and “GO terms”**. Implemented with FUMA. Gene sets that remain significant after Bonferroni correction for the metabolite-wise analysis are shown.

| **Metabolite** | **N  genes** | **Beta** | **SE** | **P** | **FULL_NAME** |  |
| --- | --- | --- | --- | --- | --- | --- |
| Tyrosine | 26 | 1.377 | 0.264 | 9.0E-08 | nikolsky_breast_cancer_5p15_amplicon | 26 genes on chromosome 5p15 |
| Threonine | 5 | 1.744 | 0.336 | 1.1E-07 | Reactome: tachykinin receptors bind tachykinins | TACR1 (2:75263590-75436826:-1),  TACR3 (4:104497188-104650973:-1),  TAC1 (7:97351220-97379784:1), TACR2 (10:71153659-71186623:-1),  TAC3 (12:57393784-57432667:-1) |
| 4-deoxyerythronic acid | 5 | 1.842 | 0.376 | 4.9E-07 | GO: pyruvate family amino acid metabolic process | ***AGXT (2:241797896-241829919:1)*** AGXT2 (5:34988206-35058198:-1)  GPT (8:145718356-145742557:1) DAO (12:109242708-109304819:1)  GPT2 (16: 46908290-46975209:1) |
| Tyrosine | 39 | 1.308 | 0.270 | 6.4E-07 | nikolsky breast cancer 1q21 amplicon | 39 genes on chromosome 1q21 |
| 3-hydroxyisovalerate | 20 | 1.382 | 0.287 | 7.4E-07 | GO: uronic acid metabolic process | ***PRKCE*** ***(2: 45868484-46425129:1)***  UGT1A8 (2:234526291-234681956:1) UGT1A10 (2:234545100-234681951:1) UGT1A9 (2:234570499-234691946:1)  UGT1A7 (2:234580584-234691945:1)  UGT1A6 (2: 234590253-234691946:1) UGT1A5 (2:234611638-234691945:1)  UGT1A4 (2:234617424-234691945:1) UGT1A3 (2:234627754-234691945:1) UGT1A1 (2:234668894-234681945:1)  ABHD10 (3:111697857-111712210:1) UGT2B17 (4:69402902-69434245:-1) UGT2B15 (4:69512348-69536346:-1) UGT2A3 (4:69794181-69817509:-1) ***UGT2B7 (4:69917081-69978705:1)*** UGT2B11 (4:70065669-70080449:-1) ***UGT2B28 (4:70146217-70160768:1) UGT2B4 (4:70345883-70391732:-1) UGT2A1 (4:70454135-70518965:-1) UGT2A1 (4:70454912-70518967:-1)*** |
| Glycolic acid | 15 | 0.742 | 0.162 | 2.4E-06 | GO: eukaryotic translation initiation factor 3 complex | EIF3I (1:32687529-32697205:1) EIF3B (7:2393721-2420380:1) COPS5 (8:67955314-67996018:-1) EIF3E (8:109213445-109447562:-1) EIF3H (8:117654369-117779164:-1) EIF3A (10:120794356-120840316:-1) EIF33F (11:7991798-8023409:1) EIF3M (11:32605344-32627808:1) EIF3J (15:44829255-44855227:1) EIF3CL (16:28390900-28415200:-1) EIF3C (16:28699879-28747051:1) EIF3G (19:10225693-10230596:-1) EIF3K (19:39109735-39127595:1)  EIF3D (22:36906897-36925483:-1) EIF3L (22:38244875-38285414:1) |
| Tryptophan | 23 | 0.768 | 0.169 | 2.7E-06 | Reactome: apc c:cdc20 mediated degradation of cyclin b | CDC20 (1:43824626-43828874:1) ***RPS27A*** (2:55459039-55462989:1) ANAPC1 (2:112523848-112642267:-1) UBE2E1 (3:23847394-23932807:1) ANAPC4 (4:25378835-25420120:1) ANAPC10 (4:145888264-146019693:-1) CCNB1 (5:68462837-68474072:1) CDC23 (5:137523339-137549032:-1) CDC26 (9:116018115-116037869:-1) ANAPC2 (9:140069236-140082989:-1) UBE2D1 (10:60094735-60130513:1) CDK1 (10:62538089-62554610:1) ANAPC16 (10:73975787-73995618:1) ANAPC15 (11:71817424-71823826:-1) ANAPC7 (12:110810705-110841535:-1) ANAPC5 (12:121746048-121837699:-1) UBC (12:125396150-125401914:-1) CDC16 (13:115000362-115038198:1) UBB (17:16284112-16286059:1) CDC27 (17:45195069-45266788:-1) ***ANAPC11*** (17:79848666-79858867:1) UBA52 (19:18682540-18688360:1) UBE2C (20:44441215-44445596:1) |
| 3-hydroxyisovalerate | 22 | 1.330 | 0.292 | 2.7E-06 | okawa neuroblastoma 1p36 31 deletion | 22 genes on chromosome 1p36.31 |
| 4-deoxythreonate | 7 | 1.458 | 0.321 | 2.8E-06 | GO: neuron intrinsic apoptotic signaling pathway in response to oxidative stress | PARK7 (1:8014351-8045565:1) PINK1 (1:20959948-20978004:1) MCL1 (1:150547032-150552066:-1) PARP1 (1:226548392-226595780:-1) FBXW7 (4:153242410-153457253:-1) ***PARK2 (6:161768452-163148803:-1) HIF1A (14:62162231-62214976:1)*** |

Genes in ***bold cursive*** were nominally associated with the metabolite (*p*<0.05) in the MAGMA gene level analysis.

### Supplementary Table 12

**FUMA GENE2FUNC gene set analysis results.** Gene sets considered signficant if adjP<0.05

| trait | Category | GeneSet | N_genes | N_overlap | p | adjP | genes |
| --- | --- | --- | --- | --- | --- | --- | --- |
| ace | Chemical_and_Genetic_pertubation | NIKOLSKY_BREAST_CANCER_20P13_AMPLICON | 8 | 2 | 1.38E-07 | 0.000456 | SPEF1:CENPB |
| ace | Positional_gene_sets | chr20p13 | 108 | 3 | 2.88E-08 | 8.62E-06 | SPEF1:CENPB:CDC25B |
| ace | Curated_gene_sets | NIKOLSKY_BREAST_CANCER_20P13_AMPLICON | 8 | 2 | 1.38E-07 | 0.000759 | SPEF1:CENPB |
| ace | Curated_gene_sets | PID_FOXM1_PATHWAY | 40 | 2 | 3.84E-06 | 0.010565 | CENPB:CDC25B |
| ace | Canonical_Pathways | PID_FOXM1_PATHWAY | 40 | 2 | 3.84E-06 | 0.008446 | CENPB:CDC25B |
| ala | Positional_gene_sets | chr12q21 | 102 | 2 | 2.53E-05 | 0.007575 | METTL25:TMTC2 |
| aohibut | GO_bp | GO_REGULATION_OF_PROSTAGLANDIN_SECRETION | 11 | 2 | 9.47E-06 | 0.043173 | P2RX7:P2RX4 |
| aohibut | GO_bp | GO_PROSTAGLANDIN_TRANSPORT | 16 | 2 | 2.06E-05 | 0.043173 | P2RX7:P2RX4 |
| aohibut | GO_bp | GO_CELLULAR_RESPONSE_TO_ATP | 18 | 2 | 2.63E-05 | 0.043173 | P2RX7:P2RX4 |
| aohibut | GO_bp | GO_POSITIVE_REGULATION_OF_FATTY_ACID_TRANSPORT | 18 | 2 | 2.63E-05 | 0.043173 | P2RX7:P2RX4 |
| aohibut | GO_bp | GO_REGULATION_OF_ICOSANOID_SECRETION | 19 | 2 | 2.94E-05 | 0.043173 | P2RX7:P2RX4 |
| aohibut | GO_bp | GO_PURINERGIC_NUCLEOTIDE_RECEPTOR_SIGNALING_PATHWAY | 22 | 2 | 3.96E-05 | 0.048565 | P2RX7:P2RX4 |
| aohibut | GWAScatalog | Mean platelet volume | 245 | 6 | 5.35E-10 | 9.71E-07 | TMEM120B:RHOF:SETD1B:PSMD9:WDR66:BCL7A |
| aohibut | GWAScatalog | Urinary metabolites | 20 | 2 | 3.26E-05 | 0.029607 | HPD:WDR66 |
| aohibut | GO_cc | GO_NUCLEAR_INNER_MEMBRANE | 57 | 3 | 1.85E-06 | 0.001856 | P2RX7:P2RX4:TMEM120B |
| aohibut | GO_cc | GO_NUCLEAR_MEMBRANE_PART | 16 | 2 | 2.06E-05 | 0.010323 | P2RX7:P2RX4 |
| aohibut | Cancer_modules | MODULE_267 | 15 | 2 | 1.81E-05 | 0.00778 | P2RX7:P2RX4 |
| aohibut | Positional_gene_sets | chr12q24 | 345 | 14 | 9.73E-28 | 2.91E-25 | P2RX7:P2RX4:RNF34:KDM2B:TMEM120B:RHOF:AC084018.1:SETD1B:HPD:PSMD9:RNU7-170P:WDR66:BCL7A:B3GNT4 |
| aohibut | GO_mf | GO_ATP_GATED_ION_CHANNEL_ACTIVITY | 8 | 2 | 4.82E-06 | 0.007932 | P2RX7:P2RX4 |
| aohibut | GO_mf | GO_NUCLEOTIDE_RECEPTOR_ACTIVITY | 21 | 2 | 3.60E-05 | 0.02596 | P2RX7:P2RX4 |
| aohibut | GO_mf | GO_PURINERGIC_RECEPTOR_ACTIVITY | 24 | 2 | 4.73E-05 | 0.02596 | P2RX7:P2RX4 |
| aohibut | GO_mf | GO_EXCITATORY_EXTRACELLULAR_LIGAND_GATED_ION_CHANNEL_ACTIVITY | 31 | 2 | 7.96E-05 | 0.032746 | P2RX7:P2RX4 |
| aohibut | Computational_gene_sets | MODULE_267 | 15 | 2 | 1.81E-05 | 0.015488 | P2RX7:P2RX4 |
| aohibut | Reactome | REACTOME_ELEVATION_OF_CYTOSOLIC_CA2PLUS_LEVELS | 16 | 2 | 2.06E-05 | 0.030917 | P2RX7:P2RX4 |
| aohibut | Reactome | REACTOME_PLATELET_CALCIUM_HOMEOSTASIS | 28 | 2 | 6.48E-05 | 0.04855 | P2RX7:P2RX4 |
| aohibut | Canonical_Pathways | REACTOME_ELEVATION_OF_CYTOSOLIC_CA2PLUS_LEVELS | 16 | 2 | 2.06E-05 | 0.045355 | P2RX7:P2RX4 |
| arb | GWAScatalog | Atopic march | 16 | 3 | 7.90E-10 | 1.43E-06 | PAQR8:EFHC1:TRAM2 |
| arb | Positional_gene_sets | chr6p12 | 107 | 4 | 4.17E-10 | 1.25E-07 | PAQR8:EFHC1:TRAM2:TRAM2-AS1 |
| bnhibut | GWAScatalog | Bipolar disorder | 653 | 8 | 5.52E-09 | 8.46E-06 | FAM83E:SPHK2:SEC1P:NTN5:FUT2:MAMSTR:RASIP1:IZUMO1 |
| bnhibut | GWAScatalog | Alcohol use disorder (total score) | 39 | 4 | 1.16E-08 | 8.46E-06 | FUT2:MAMSTR:RASIP1:IZUMO1 |
| bnhibut | GWAScatalog | Dietary macronutrient intake | 8 | 3 | 1.40E-08 | 8.46E-06 | FUT2:RASIP1:IZUMO1 |
| bnhibut | GWAScatalog | Retinal vascular caliber | 18 | 3 | 2.03E-07 | 9.20E-05 | FUT2:RASIP1:IZUMO1 |
| bnhibut | GWAScatalog | Crohn's disease | 600 | 6 | 1.98E-06 | 0.00072 | SPHK2:NTN5:FUT2:MAMSTR:RASIP1:IZUMO1 |
| bnhibut | GWAScatalog | Vitamin B levels in ischemic stroke | 8 | 2 | 1.16E-05 | 0.003512 | FUT2:RASIP1 |
| bnhibut | GWAScatalog | Estimated glomerular filtration rate | 527 | 5 | 2.07E-05 | 0.005372 | SLC6A13:FUT2:MAMSTR:RASIP1:RAI14 |
| bnhibut | GWAScatalog | Elevated serum carcinoembryonic antigen levels | 14 | 2 | 3.76E-05 | 0.008539 | FAM83E:FUT2 |
| bnhibut | GWAScatalog | Response to angiotensin II receptor blocker therapy | 16 | 2 | 4.96E-05 | 0.00949 | IQSEC3:DNAJC21 |
| bnhibut | GWAScatalog | Inflammatory bowel disease | 640 | 5 | 5.23E-05 | 0.00949 | NTN5:FUT2:MAMSTR:RASIP1:IZUMO1 |
| bnhibut | GWAScatalog | Blood urea nitrogen levels | 145 | 3 | 0.000117 | 0.019322 | MAMSTR:IZUMO1:RAI14 |
| bnhibut | GO_cc | GO_INHIBITORY_SYNAPSE | 16 | 2 | 4.96E-05 | 0.049641 | IQSEC3:GAD1 |
| bnhibut | Positional_gene_sets | chr5p13 | 112 | 5 | 1.00E-08 | 2.99E-06 | RAI14:TTC23L:RAD1:DNAJC21:NIPBL |
| bnhibut | Positional_gene_sets | chr19q13 | 1073 | 8 | 2.53E-07 | 3.79E-05 | FAM83E:SPHK2:SEC1P:NTN5:FUT2:MAMSTR:RASIP1:IZUMO1 |
| bnhibut | Positional_gene_sets | chr12p13 | 292 | 4 | 3.75E-05 | 0.00374 | IQSEC3:SLC6A13:KDM5A:CCDC77 |
| bnhibut | Positional_gene_sets | chr2q31 | 131 | 3 | 8.67E-05 | 0.006479 | GAD1:GORASP2:TLK1 |
| bnhibut | microRNA_targets | TTTGTAG_MIR520D | 336 | 4 | 6.47E-05 | 0.0143 | GAD1:RAI14:NIPBL:ATXN1 |
| bohibut | GWAScatalog | vWF levels | 27 | 2 | 1.21E-05 | 0.021923 | NIPSNAP3A:NIPSNAP3B |
| bohibut | Positional_gene_sets | chr2q14 | 122 | 3 | 1.44E-06 | 0.000432 | PSD4:PAX8-AS1:PAX8 |
| bohibut | Positional_gene_sets | chr9q31 | 118 | 2 | 0.000235 | 0.035207 | NIPSNAP3A:NIPSNAP3B |
| bohibut | Cancer_gene_neighborhoods | MORF_PTPN9 | 61 | 2 | 6.28E-05 | 0.017167 | PSD4:PAX8 |
| bohibut | Cancer_gene_neighborhoods | MORF_ORC1L | 69 | 2 | 8.04E-05 | 0.017167 | PSD4:PAX8 |
| bohibut | Cancer_gene_neighborhoods | MORF_CASP2 | 102 | 2 | 0.000176 | 0.021062 | PSD4:PAX8 |
| bohibut | Cancer_gene_neighborhoods | MORF_CNTN1 | 108 | 2 | 0.000197 | 0.021062 | PSD4:PAX8 |
| bohibut | Cancer_gene_neighborhoods | MORF_PHB | 121 | 2 | 0.000248 | 0.021145 | PSD4:PAX8 |
| bohibut | Cancer_gene_neighborhoods | MORF_IKBKG | 136 | 2 | 0.000313 | 0.022249 | PSD4:PAX8 |
| bohibut | Computational_gene_sets | MORF_PTPN9 | 61 | 2 | 6.28E-05 | 0.034496 | PSD4:PAX8 |
| bohibut | Computational_gene_sets | MORF_ORC1L | 69 | 2 | 8.04E-05 | 0.034496 | PSD4:PAX8 |
| bohibut | Computational_gene_sets | MORF_CASP2 | 102 | 2 | 0.000176 | 0.042322 | PSD4:PAX8 |
| bohibut | Computational_gene_sets | MORF_CNTN1 | 108 | 2 | 0.000197 | 0.042322 | PSD4:PAX8 |
| bohibut | Computational_gene_sets | MORF_PHB | 121 | 2 | 0.000248 | 0.042489 | PSD4:PAX8 |
| bohibut | Computational_gene_sets | MORF_IKBKG | 136 | 2 | 0.000313 | 0.044707 | PSD4:PAX8 |
| bohival | GWAScatalog | Subjective response to lithium treatment | 16 | 4 | 4.01E-11 | 7.28E-08 | DCUN1D1:MCCC1:LAMP3:MCF2L2 |
| bohival | GWAScatalog | Asthma or allergic disease (pleiotropy) | 161 | 5 | 5.68E-09 | 5.16E-06 | NBPF18P:S100A11:FLG-AS1:HRNR:FLG |
| bohival | GWAScatalog | Allergic disease (asthma, hay fever or eczema) | 283 | 5 | 9.52E-08 | 5.76E-05 | NBPF18P:S100A11:FLG-AS1:HRNR:FLG |
| bohival | GWAScatalog | Parkinson's disease | 166 | 3 | 4.61E-05 | 0.02093 | DCUN1D1:MCCC1:LAMP3 |
| bohival | Positional_gene_sets | chr1q21 | 317 | 9 | 1.80E-15 | 5.37E-13 | LINGO4:THEM4:KRT8P28:S100A10:NBPF18P:S100A11:FLG-AS1:HRNR:FLG |
| bohival | Positional_gene_sets | chr3q27 | 99 | 4 | 8.12E-08 | 1.21E-05 | MCCC1:MCCC1-AS1:LAMP3:MCF2L2 |
| bohival | GO_mf | GO_STRUCTURAL_CONSTITUENT_OF_EPIDERMIS | 16 | 2 | 2.06E-05 | 0.033929 | HRNR:FLG |
| bohival | Canonical_Pathways | NABA_SECRETED_FACTORS | 339 | 4 | 1.10E-05 | 0.02414 | S100A10:S100A11:HRNR:FLG |
| cit | Chemical_and_Genetic_pertubation | NIKOLSKY_BREAST_CANCER_17Q11_Q21_AMPLICON | 132 | 6 | 7.27E-14 | 2.40E-10 | FOXN1:SUPT6H:PHF12:PIPOX:SEZ6:ACACA |
| cit | Positional_gene_sets | chr17q11 | 143 | 5 | 5.97E-11 | 1.79E-08 | FOXN1:SUPT6H:PHF12:PIPOX:SEZ6 |
| cit | Curated_gene_sets | NIKOLSKY_BREAST_CANCER_17Q11_Q21_AMPLICON | 132 | 6 | 7.27E-14 | 4.00E-10 | FOXN1:SUPT6H:PHF12:PIPOX:SEZ6:ACACA |
| cit | microRNA_targets | GGCAGTG_MIR3243P | 96 | 2 | 0.000207 | 0.045854 | PHF12:ACACA |
| cit | TF_targets | TST1_01 | 264 | 3 | 2.33E-05 | 0.014222 | SUPT6H:PIPOX:ACACA |
| cit | TF_targets | MYOGNF1_01 | 48 | 2 | 5.16E-05 | 0.01574 | SEZ6:ACACA |
| doeta | GWAScatalog | Urate levels in obese individuals | 50 | 2 | 3.01E-05 | 0.035537 | STAU2:UBE2W |
| doeta | GWAScatalog | Manganese levels | 57 | 2 | 3.92E-05 | 0.035537 | UBE2W:TCEB1 |
| doeta | Chemical_and_Genetic_pertubation | NIKOLSKY_BREAST_CANCER_8Q12_Q22_AMPLICON | 132 | 5 | 4.29E-12 | 1.42E-08 | STAU2:UBE2W:TCEB1:TMEM70:LY96 |
| doeta | Chemical_and_Genetic_pertubation | ONKEN_UVEAL_MELANOMA_UP | 774 | 4 | 3.48E-06 | 0.005742 | UBE2W:TCEB1:TMEM70:LY96 |
| doeta | Immunologic_signatures | GSE17721_LPS_VS_CPG_6H_BMDC_UP | 197 | 3 | 3.50E-06 | 0.008658 | TCEB1:TMEM70:LY96 |
| doeta | Immunologic_signatures | GSE20198_UNTREATED_VS_IL12_TREATED_ACT_CD4_TCELL_UP | 198 | 3 | 3.55E-06 | 0.008658 | STAU2:TCEB1:TMEM70 |
| doeta | Positional_gene_sets | chr8q21 | 150 | 6 | 5.71E-15 | 1.71E-12 | STAU2:UBE2W:TCEB1:TMEM70:RPS20P21:LY96 |
| doeta | Curated_gene_sets | NIKOLSKY_BREAST_CANCER_8Q12_Q22_AMPLICON | 132 | 5 | 4.29E-12 | 2.36E-08 | STAU2:UBE2W:TCEB1:TMEM70:LY96 |
| doeta | Curated_gene_sets | ONKEN_UVEAL_MELANOMA_UP | 774 | 4 | 3.48E-06 | 0.009565 | UBE2W:TCEB1:TMEM70:LY96 |
| doeta | Curated_gene_sets | REACTOME_CLASS_I_MHC_MEDIATED_ANTIGEN_PROCESSING_PRESENTATION | 359 | 3 | 2.11E-05 | 0.038706 | UBE2W:TCEB1:LY96 |
| doeta | Oncogenic_signatures | HOXA9_DN.V1_UP | 189 | 2 | 0.000432 | 0.047397 | UBE2W:LY96 |
| doeta | Oncogenic_signatures | STK33_SKM_UP | 271 | 2 | 0.000883 | 0.047397 | UBE2W:LY96 |
| doeta | Oncogenic_signatures | STK33_UP | 283 | 2 | 0.000962 | 0.047397 | UBE2W:LY96 |
| doeta | Oncogenic_signatures | STK33_NOMO_UP | 289 | 2 | 0.001003 | 0.047397 | UBE2W:LY96 |
| doeta | KEGG | KEGG_UBIQUITIN_MEDIATED_PROTEOLYSIS | 134 | 2 | 0.000217 | 0.040428 | UBE2W:TCEB1 |
| doeta | microRNA_targets | CAGTATT_MIR200B_MIR200C_MIR429 | 461 | 3 | 4.45E-05 | 0.009831 | UBE2W:TCEB1:TMEM70 |
| doeta | Reactome | REACTOME_CLASS_I_MHC_MEDIATED_ANTIGEN_PROCESSING_PRESENTATION | 359 | 3 | 2.11E-05 | 0.031642 | UBE2W:TCEB1:LY96 |
| doeta | Canonical_Pathways | REACTOME_CLASS_I_MHC_MEDIATED_ANTIGEN_PROCESSING_PRESENTATION | 359 | 3 | 2.11E-05 | 0.046418 | UBE2W:TCEB1:LY96 |
| dta | GO_bp | GO_ANTIBIOTIC_BIOSYNTHETIC_PROCESS | 16 | 4 | 2.94E-11 | 1.08E-07 | DUOX2:DUOXA2:DUOXA1:DUOX1 |
| dta | GO_bp | GO_THYROID_HORMONE_GENERATION | 16 | 4 | 2.94E-11 | 1.08E-07 | DUOX2:DUOXA2:DUOXA1:DUOX1 |
| dta | GO_bp | GO_THYROID_HORMONE_METABOLIC_PROCESS | 20 | 4 | 7.82E-11 | 1.92E-07 | DUOX2:DUOXA2:DUOXA1:DUOX1 |
| dta | GO_bp | GO_CELLULAR_MODIFIED_AMINO_ACID_METABOLIC_PROCESS | 37 | 4 | 1.06E-09 | 1.95E-06 | DUOX2:DUOXA2:DUOXA1:DUOX1 |
| dta | GO_bp | GO_DRUG_METABOLIC_PROCESS | 338 | 6 | 2.22E-09 | 3.27E-06 | DUOX2:DUOXA2:DUOXA1:DUOX1:GATM:SQRDL |
| dta | GO_bp | GO_PHENOL_CONTAINING_COMPOUND_METABOLIC_PROCESS | 60 | 4 | 7.80E-09 | 9.56E-06 | DUOX2:DUOXA2:DUOXA1:DUOX1 |
| dta | GO_bp | GO_ANTIBIOTIC_METABOLIC_PROCESS | 92 | 4 | 4.44E-08 | 4.66E-05 | DUOX2:DUOXA2:DUOXA1:DUOX1 |
| dta | GO_bp | GO_REACTIVE_OXYGEN_SPECIES_BIOSYNTHETIC_PROCESS | 109 | 4 | 8.80E-08 | 8.09E-05 | DUOX2:DUOXA2:DUOXA1:DUOX1 |
| dta | GO_bp | GO_REACTIVE_OXYGEN_SPECIES_METABOLIC_PROCESS | 163 | 4 | 4.43E-07 | 0.00033 | DUOX2:DUOXA2:DUOXA1:DUOX1 |
| dta | GO_bp | GO_POSITIVE_REGULATION_OF_HYDROGEN_PEROXIDE_BIOSYNTHETIC_PROCESS | 3 | 2 | 4.48E-07 | 0.00033 | DUOXA2:DUOXA1 |
| dta | GO_bp | GO_ORGANIC_HYDROXY_COMPOUND_METABOLIC_PROCESS | 484 | 5 | 9.08E-07 | 0.000607 | SORD:DUOX2:DUOXA2:DUOXA1:DUOX1 |
| dta | GO_bp | GO_COFACTOR_BIOSYNTHETIC_PROCESS | 207 | 4 | 1.15E-06 | 0.000704 | DUOX2:DUOXA2:DUOXA1:DUOX1 |
| dta | GO_bp | GO_HORMONE_METABOLIC_PROCESS | 217 | 4 | 1.39E-06 | 0.000784 | DUOX2:DUOXA2:DUOXA1:DUOX1 |
| dta | GO_bp | GO_REGULATION_OF_THYROID_HORMONE_GENERATION | 5 | 2 | 1.49E-06 | 0.000784 | DUOXA2:DUOXA1 |
| dta | GO_bp | GO_POSITIVE_REGULATION_OF_COFACTOR_METABOLIC_PROCESS | 6 | 2 | 2.24E-06 | 0.001098 | DUOXA2:DUOXA1 |
| dta | GO_bp | GO_REGULATION_OF_HYDROGEN_PEROXIDE_BIOSYNTHETIC_PROCESS | 7 | 2 | 3.14E-06 | 0.00144 | DUOXA2:DUOXA1 |
| dta | GO_bp | GO_HYDROGEN_PEROXIDE_METABOLIC_PROCESS | 13 | 2 | 1.16E-05 | 0.005028 | DUOXA2:DUOXA1 |
| dta | GO_bp | GO_COFACTOR_METABOLIC_PROCESS | 441 | 4 | 2.28E-05 | 0.009305 | DUOX2:DUOXA2:DUOXA1:DUOX1 |
| dta | GO_bp | GO_OXIDATION_REDUCTION_PROCESS | 952 | 5 | 2.44E-05 | 0.009436 | SORD:DUOX2:DUOX1:C15orf48:SQRDL |
| dta | GO_bp | GO_REGULATION_OF_HORMONE_LEVELS | 510 | 4 | 4.02E-05 | 0.014769 | DUOX2:DUOXA2:DUOXA1:DUOX1 |
| dta | GO_bp | GO_HYDROGEN_PEROXIDE_CATABOLIC_PROCESS | 32 | 2 | 7.36E-05 | 0.025772 | DUOX2:DUOX1 |
| dta | GO_bp | GO_SUPEROXIDE_ANION_GENERATION | 33 | 2 | 7.84E-05 | 0.026181 | DUOX2:DUOX1 |
| dta | GO_bp | GO_REGULATION_OF_COFACTOR_METABOLIC_PROCESS | 35 | 2 | 8.83E-05 | 0.028208 | DUOXA2:DUOXA1 |
| dta | GO_bp | GO_REGULATION_OF_HORMONE_METABOLIC_PROCESS | 37 | 2 | 9.88E-05 | 0.030245 | DUOXA2:DUOXA1 |
| dta | Chemical_and_Genetic_pertubation | BENPORATH_SUZ12_TARGETS | 1031 | 7 | 5.51E-08 | 0.000182 | DUOX2:DUOXA2:DUOXA1:DUOX1:GATM:SLC30A4:SQRDL |
| dta | Chemical_and_Genetic_pertubation | BENPORATH_PRC2_TARGETS | 648 | 5 | 3.79E-06 | 0.006257 | DUOX2:DUOXA2:DUOXA1:DUOX1:SLC30A4 |
| dta | Chemical_and_Genetic_pertubation | BENPORATH_EED_TARGETS | 1045 | 5 | 3.81E-05 | 0.04038 | DUOX2:DUOXA2:DUOXA1:DUOX1:SLC30A4 |
| dta | Chemical_and_Genetic_pertubation | BENPORATH_ES_WITH_H3K27ME3 | 1101 | 5 | 4.89E-05 | 0.04038 | DUOX2:DUOXA2:DUOXA1:DUOX1:SLC30A4 |
| dta | GO_cc | GO_NADPH_OXIDASE_COMPLEX | 15 | 2 | 1.56E-05 | 0.015664 | DUOX2:DUOX1 |
| dta | Positional_gene_sets | chr15q21 | 138 | 13 | 4.52E-31 | 1.35E-28 | CTD-2008A1.2:SORD:DUOX2:DUOXA2:DUOXA1:DUOX1:SHF:SLC28A2:GATM:SPATA5L1:C15orf48:SLC30A4:SQRDL |
| dta | GO_mf | GO_NAD_P_H_OXIDASE_ACTIVITY | 8 | 2 | 4.18E-06 | 0.005397 | DUOX2:DUOX1 |
| dta | GO_mf | GO_OXIDOREDUCTASE_ACTIVITY | 730 | 5 | 6.77E-06 | 0.005397 | SORD:DUOX2:DUOX1:C15orf48:SQRDL |
| dta | GO_mf | GO_SUPEROXIDE_GENERATING_NADPH_OXIDASE_ACTIVITY | 12 | 2 | 9.84E-06 | 0.005397 | DUOX2:DUOX1 |
| dta | GO_mf | GO_OXIDOREDUCTASE_ACTIVITY_ACTING_ON_NAD_P_H_OXYGEN_AS_ACCEPTOR | 17 | 2 | 2.03E-05 | 0.008332 | DUOX2:DUOX1 |
| dta | GO_mf | GO_COFACTOR_BINDING | 493 | 4 | 3.52E-05 | 0.011586 | SORD:DUOX2:DUOX1:SQRDL |
| dta | GO_mf | GO_COENZYME_BINDING | 284 | 3 | 0.000182 | 0.049816 | SORD:DUOX1:SQRDL |
| dta | Curated_gene_sets | BENPORATH_SUZ12_TARGETS | 1031 | 7 | 5.51E-08 | 0.000303 | DUOX2:DUOXA2:DUOXA1:DUOX1:GATM:SLC30A4:SQRDL |
| dta | Curated_gene_sets | BENPORATH_PRC2_TARGETS | 648 | 5 | 3.79E-06 | 0.010423 | DUOX2:DUOXA2:DUOXA1:DUOX1:SLC30A4 |
| dta | Curated_gene_sets | REACTOME_THYROXINE_BIOSYNTHESIS | 10 | 2 | 6.71E-06 | 0.012312 | DUOX2:DUOX1 |
| dta | Curated_gene_sets | REACTOME_METABOLISM_OF_AMINO_ACIDS_AND_DERIVATIVES | 367 | 4 | 1.11E-05 | 0.015247 | DUOX2:DUOX1:GATM:SQRDL |
| dta | Curated_gene_sets | REACTOME_METABOLISM_OF_AMINE_DERIVED_HORMONES | 18 | 2 | 2.28E-05 | 0.02507 | DUOX2:DUOX1 |
| dta | Curated_gene_sets | BENPORATH_EED_TARGETS | 1045 | 5 | 3.81E-05 | 0.034955 | DUOX2:DUOXA2:DUOXA1:DUOX1:SLC30A4 |
| dta | Curated_gene_sets | BENPORATH_ES_WITH_H3K27ME3 | 1101 | 5 | 4.89E-05 | 0.038441 | DUOX2:DUOXA2:DUOXA1:DUOX1:SLC30A4 |
| dta | Reactome | REACTOME_THYROXINE_BIOSYNTHESIS | 10 | 2 | 6.71E-06 | 0.008309 | DUOX2:DUOX1 |
| dta | Reactome | REACTOME_METABOLISM_OF_AMINO_ACIDS_AND_DERIVATIVES | 367 | 4 | 1.11E-05 | 0.008309 | DUOX2:DUOX1:GATM:SQRDL |
| dta | Reactome | REACTOME_METABOLISM_OF_AMINE_DERIVED_HORMONES | 18 | 2 | 2.28E-05 | 0.011386 | DUOX2:DUOX1 |
| dta | Canonical_Pathways | REACTOME_THYROXINE_BIOSYNTHESIS | 10 | 2 | 6.71E-06 | 0.01219 | DUOX2:DUOX1 |
| dta | Canonical_Pathways | REACTOME_METABOLISM_OF_AMINO_ACIDS_AND_DERIVATIVES | 367 | 4 | 1.11E-05 | 0.01219 | DUOX2:DUOX1:GATM:SQRDL |
| dta | Canonical_Pathways | REACTOME_METABOLISM_OF_AMINE_DERIVED_HORMONES | 18 | 2 | 2.28E-05 | 0.016703 | DUOX2:DUOX1 |
| etnh | GWAScatalog | Retinal arteriolar caliber | 6 | 2 | 1.48E-07 | 0.000268 | MXRA7:MFSD11 |
| etnh | Chemical_and_Genetic_pertubation | CREIGHTON_ENDOCRINE_THERAPY_RESISTANCE_3 | 715 | 4 | 1.75E-07 | 0.000577 | ACOX1:ST6GALNAC1:MXRA7:MFSD11 |
| etnh | Positional_gene_sets | chr17q25 | 263 | 4 | 3.15E-09 | 9.43E-07 | ACOX1:ST6GALNAC1:MXRA7:MFSD11 |
| etnh | Curated_gene_sets | CREIGHTON_ENDOCRINE_THERAPY_RESISTANCE_3 | 715 | 4 | 1.75E-07 | 0.000962 | ACOX1:ST6GALNAC1:MXRA7:MFSD11 |
| form | GWAScatalog | Tuberculosis | 58 | 7 | 1.19E-15 | 2.17E-12 | MTHFR:CLCN6:NPPA:KIAA2013:PLOD1:MFN2:MIIP |
| form | GWAScatalog | Plasma homocysteine levels | 6 | 3 | 2.73E-09 | 2.48E-06 | MTHFR:RNU5E-1:MFN2 |
| form | GWAScatalog | Serum folate levels | 15 | 3 | 6.20E-08 | 3.17E-05 | MTHFR:RNU5E-1:KIAA2013 |
| form | GWAScatalog | Atopic march | 16 | 3 | 7.63E-08 | 3.17E-05 | PAQR8:EFHC1:TRAM2 |
| form | GWAScatalog | Diastolic blood pressure x alcohol consumption interaction (2df test) | 78 | 4 | 8.73E-08 | 3.17E-05 | C1orf167:CLCN6:KIAA2013:PLOD1 |
| form | GWAScatalog | Diastolic blood pressure x alcohol consumption (light vs heavy) interaction (2df test) | 36 | 3 | 9.66E-07 | 0.000292 | MTHFR:CLCN6:KIAA2013 |
| form | GWAScatalog | Diastolic blood pressure x smoking status (ever vs never) interaction (2df test) | 88 | 3 | 1.46E-05 | 0.00354 | MTHFR:CLCN6:NPPA |
| form | GWAScatalog | Systolic blood pressure x smoking status (ever vs never) interaction (2df test) | 90 | 3 | 1.56E-05 | 0.00354 | MTHFR:CLCN6:NPPA |
| form | GWAScatalog | Diastolic blood pressure x smoking status (current vs non-current) interaction (2df test) | 101 | 3 | 2.21E-05 | 0.004447 | MTHFR:CLCN6:NPPA |
| form | GWAScatalog | Systolic blood pressure x smoking status (current vs non-current) interaction (2df test) | 106 | 3 | 2.55E-05 | 0.004625 | MTHFR:CLCN6:NPPA |
| form | GWAScatalog | Moyamoya disease | 16 | 2 | 3.36E-05 | 0.005536 | MTHFR:PLOD1 |
| form | GWAScatalog | Pulse pressure x alcohol consumption interaction (2df test) | 46 | 2 | 0.000287 | 0.043348 | C1orf167:CLCN6 |
| form | Positional_gene_sets | chr1p36 | 545 | 10 | 6.48E-14 | 1.94E-11 | C1orf167:MTHFR:CLCN6:NPPA:RNU5E-1:RNU5E-4P:KIAA2013:PLOD1:MFN2:MIIP |
| form | Positional_gene_sets | chr6p12 | 107 | 4 | 3.13E-07 | 4.67E-05 | PAQR8:EFHC1:TRAM2:TRAM2-AS1 |
| form | Positional_gene_sets | chr3q22 | 103 | 3 | 2.34E-05 | 0.002331 | DZIP1L:A4GNT:DBR1 |
| glc | GO_bp | GO_REGULATION_OF_RESPONSE_TO_DNA_DAMAGE_STIMULUS | 206 | 6 | 1.37E-06 | 0.010044 | EYA3:FOXM1:ZNF385A:WDR76:PPP4R2:DEK |
| glc | Chemical_and_Genetic_pertubation | NIKOLSKY_BREAST_CANCER_11Q12_Q14_AMPLICON | 158 | 5 | 7.34E-06 | 0.02424 | SPCS2P4:TMEM126B:TMEM126A:SYTL2:PICALM |
| glc | Positional_gene_sets | chr11q14 | 116 | 4 | 4.58E-05 | 0.0137 | TMEM126B:TMEM126A:SYTL2:PICALM |
| glc | Positional_gene_sets | chr4q25 | 71 | 3 | 0.000242 | 0.036152 | CFI:GAR1:RRH |
| glc | Positional_gene_sets | chr12q13 | 364 | 5 | 0.000379 | 0.037779 | ESPL1:PFDN5:C12orf10:AAAS:ZNF385A |
| glc | Wikipathways | Nuclear Receptors | 39 | 3 | 4.02E-05 | 0.021819 | ROR1:ESR2:NR2C2 |
| glc | Curated_gene_sets | NIKOLSKY_BREAST_CANCER_11Q12_Q14_AMPLICON | 158 | 5 | 7.34E-06 | 0.040383 | SPCS2P4:TMEM126B:TMEM126A:SYTL2:PICALM |
| glc | Oncogenic_signatures | PRC2_EZH2_UP.V1_DN | 187 | 4 | 0.000289 | 0.030761 | WDR76:SOGA1:SLC7A2:PTPRD |
| glc | Oncogenic_signatures | RAF_UP.V1_DN | 193 | 4 | 0.000326 | 0.030761 | SYTL2:PALLD:SLC7A2:PDGFRL |
| glc | microRNA_targets | GCAAAAA_MIR129 | 185 | 5 | 1.58E-05 | 0.003481 | ZNF385A:ZNF609:NR2C2:KDM1B:MAN1A1 |
| glya | Chemical_and_Genetic_pertubation | WAKABAYASHI_ADIPOGENESIS_PPARG_RXRA_BOUND_8D | 824 | 6 | 6.82E-06 | 0.022529 | AGBL5:OST4:EMILIN1:ABHD1:PREB:OXSR1 |
| glya | Chemical_and_Genetic_pertubation | MARSON_BOUND_BY_FOXP3_STIMULATED | 970 | 6 | 1.72E-05 | 0.02846 | SLC35F6:CENPA:CGREF1:ABHD1:CAD:DLEC1 |
| glya | Positional_gene_sets | chr2p23 | 128 | 17 | 7.70E-39 | 2.30E-36 | SLC35F6:CENPA:DPYSL5:MAPRE3:TMEM214:AGBL5:AGBL5-IT1:OST4:EMILIN1:KHK:CGREF1:ABHD1:PREB:CAD:MPV17:SNX17:SLC4A1AP |
| glya | Positional_gene_sets | chr3p22 | 153 | 4 | 2.01E-06 | 0.0003 | DLEC1:OXSR1:SLC22A14:XYLB |
| glya | Curated_gene_sets | WAKABAYASHI_ADIPOGENESIS_PPARG_RXRA_BOUND_8D | 824 | 6 | 6.82E-06 | 0.037532 | AGBL5:OST4:EMILIN1:ABHD1:PREB:OXSR1 |
| glya | Curated_gene_sets | MARSON_BOUND_BY_FOXP3_STIMULATED | 970 | 6 | 1.72E-05 | 0.047414 | SLC35F6:CENPA:CGREF1:ABHD1:CAD:DLEC1 |
| glya | microRNA_targets | TTCCGTT_MIR191 | 29 | 2 | 0.000139 | 0.030649 | MAPRE3:OXSR1 |
| gly | Chemical_and_Genetic_pertubation | BOHN_PRIMARY_IMMUNODEFICIENCY_SYNDROM_UP | 46 | 2 | 1.70E-06 | 0.005614 | CENPE:GLDC |
| gly | Curated_gene_sets | BOHN_PRIMARY_IMMUNODEFICIENCY_SYNDROM_UP | 46 | 2 | 1.70E-06 | 0.009352 | CENPE:GLDC |
| his | GWAScatalog | Ulcerative colitis | 366 | 16 | 1.05E-26 | 1.91E-23 | STAC2:FBXL20:MED1:CDK12:PPP1R1B:STARD3:TCAP:PNMT:PGAP3:ERBB2:MIEN1:IKZF3:ZPBP2:GSDMB:ORMDL3:GSDMA |
| his | GWAScatalog | Asthma or allergic disease (pleiotropy) | 161 | 12 | 1.58E-22 | 1.34E-19 | STARD3:TCAP:PNMT:PGAP3:ERBB2:MIR4728:MIEN1:IKZF3:ZPBP2:GSDMB:ORMDL3:GSDMA |
| his | GWAScatalog | Bronchial hyperresponsiveness in asthma | 8 | 7 | 2.22E-22 | 1.34E-19 | CDK12:PNMT:PGAP3:IKZF3:GSDMB:ORMDL3:GSDMA |
| his | GWAScatalog | Pancreatic ductal adenocarcinoma | 22 | 6 | 3.98E-15 | 1.81E-12 | STARD3:TCAP:PNMT:PGAP3:ERBB2:MED24 |
| his | GWAScatalog | Asthma | 315 | 8 | 2.62E-11 | 9.50E-09 | PGAP3:ERBB2:MIEN1:IKZF3:ZPBP2:GSDMB:ORMDL3:GSDMA |
| his | GWAScatalog | Adult asthma | 27 | 4 | 2.99E-09 | 9.04E-07 | ERBB2:ZPBP2:GSDMB:ORMDL3 |
| his | GWAScatalog | White blood cell count | 206 | 6 | 4.85E-09 | 1.26E-06 | GSDMB:ORMDL3:GSDMA:PSMD3:CSF3:MED24 |
| his | GWAScatalog | Primary biliary cirrhosis | 44 | 4 | 2.29E-08 | 5.20E-06 | IKZF3:ZPBP2:GSDMB:ORMDL3 |
| his | GWAScatalog | Rheumatoid arthritis | 208 | 5 | 2.78E-07 | 5.61E-05 | MED1:IKZF3:GSDMB:ORMDL3:CSF3 |
| his | GWAScatalog | Primary biliary cholangitis | 98 | 4 | 5.95E-07 | 0.000108 | IKZF3:ZPBP2:GSDMB:ORMDL3 |
| his | GWAScatalog | Mitochondrial DNA copy number | 31 | 3 | 1.27E-06 | 0.000209 | PSMD3:CSF3:MED24 |
| his | GWAScatalog | Systemic lupus erythematosus | 330 | 5 | 2.69E-06 | 0.000407 | MED1:ERBB2:MIEN1:IKZF3:GSDMB |
| his | GWAScatalog | Asthma (age of onset) | 48 | 3 | 4.84E-06 | 0.000676 | ZPBP2:GSDMB:ORMDL3 |
| his | GWAScatalog | Neutrophil count | 169 | 4 | 5.23E-06 | 0.000678 | PSMD3:CSF3:MED24:SLC12A7 |
| his | GWAScatalog | Systemic sclerosis | 55 | 3 | 7.33E-06 | 0.000886 | GSDMB:ORMDL3:GSDMA |
| his | GWAScatalog | Rheumatoid arthritis (ACPA-positive) | 72 | 3 | 1.65E-05 | 0.001875 | IKZF3:GSDMB:ORMDL3 |
| his | GWAScatalog | Asthma (childhood onset) | 255 | 4 | 2.64E-05 | 0.002817 | ZPBP2:GSDMB:ORMDL3:GSDMA |
| his | GWAScatalog | Type 1 diabetes | 89 | 3 | 3.12E-05 | 0.003149 | IKZF3:GSDMB:ORMDL3 |
| his | GWAScatalog | Menopause (age at onset) | 93 | 3 | 3.56E-05 | 0.003402 | CDK12:STARD3:PGAP3 |
| his | GWAScatalog | Hematological parameters | 14 | 2 | 4.10E-05 | 0.003725 | ORMDL3:GSDMA |
| his | GWAScatalog | Crohn's disease | 600 | 5 | 4.79E-05 | 0.004143 | IKZF3:ZPBP2:GSDMB:ORMDL3:GSDMA |
| his | GWAScatalog | Inflammatory bowel disease | 640 | 5 | 6.51E-05 | 0.005367 | IKZF3:ZPBP2:GSDMB:ORMDL3:GSDMA |
| his | GWAScatalog | Bipolar disorder | 653 | 5 | 7.15E-05 | 0.005588 | ERBB2:GSDMA:PSMD3:CSF3:MED24 |
| his | GWAScatalog | White blood cell types | 19 | 2 | 7.70E-05 | 0.005588 | PSMD3:CSF3 |
| his | GWAScatalog | Fractional exhaled nitric oxide (childhood) | 19 | 2 | 7.70E-05 | 0.005588 | ZPBP2:GSDMB |
| his | GWAScatalog | Urinary metabolites | 20 | 2 | 8.55E-05 | 0.005968 | PNMT:SLC6A19 |
| his | GWAScatalog | Asthma and hay fever | 22 | 2 | 0.000104 | 0.006981 | IKZF3:GSDMA |
| his | GWAScatalog | Triglyceride levels | 144 | 3 | 0.000131 | 0.008474 | CDK12:STARD3:PGAP3 |
| his | GWAScatalog | Self-reported allergy | 34 | 2 | 0.000251 | 0.015705 | IKZF3:GSDMB |
| his | GWAScatalog | Lung function (FVC) | 182 | 3 | 0.000261 | 0.015765 | FBXL20:MED1:CDK12 |
| his | GWAScatalog | Selective IgA deficiency | 39 | 2 | 0.000331 | 0.019366 | IKZF3:ORMDL3 |
| his | GWAScatalog | Polycystic ovary syndrome | 41 | 2 | 0.000366 | 0.020743 | ERBB2:IKZF3 |
| his | GWAScatalog | Estimated glomerular filtration rate | 527 | 4 | 0.00043 | 0.023648 | MED1:CDK12:PPP1R1B:MED24 |
| his | GWAScatalog | Creatine kinase levels | 50 | 2 | 0.000544 | 0.029055 | PSMD3:CSF3 |
| his | GWAScatalog | Atrial fibrillation | 240 | 3 | 0.000584 | 0.030308 | ZPBP2:GSDMB:ORMDL3 |
| his | GWAScatalog | Subcutaneous adipose tissue | 62 | 2 | 0.000836 | 0.042147 | ZPBP2:GSDMB |
| his | Chemical_and_Genetic_pertubation | NIKOLSKY_BREAST_CANCER_17Q11_Q21_AMPLICON | 132 | 19 | 1.03E-42 | 3.39E-39 | STAC2:FBXL20:MED1:CDK12:PPP1R1B:STARD3:TCAP:PNMT:PGAP3:ERBB2:MIEN1:IKZF3:ZPBP2:GSDMB:ORMDL3:GSDMA:PSMD3:CSF3:MED24 |
| his | Chemical_and_Genetic_pertubation | FARMER_BREAST_CANCER_CLUSTER_8 | 6 | 5 | 5.91E-16 | 9.77E-13 | MED1:PNMT:PGAP3:ERBB2:GSDMB |
| his | Chemical_and_Genetic_pertubation | SMID_BREAST_CANCER_ERBB2_UP | 148 | 8 | 6.01E-14 | 6.61E-11 | MED1:TCAP:PNMT:PGAP3:ERBB2:GSDMB:PSMD3:MED24 |
| his | Chemical_and_Genetic_pertubation | LU_EZH2_TARGETS_UP | 281 | 4 | 3.85E-05 | 0.031802 | PPP1R1B:PGAP3:MIEN1:GSDMA |
| his | Positional_gene_sets | chr17q12 | 125 | 13 | 2.03E-26 | 6.07E-24 | STAC2:FBXL20:MED1:CDK12:RNU6-233P:PPP1R1B:STARD3:TCAP:PNMT:PGAP3:ERBB2:MIR4728:MIEN1 |
| his | Positional_gene_sets | chr17q21 | 399 | 8 | 1.71E-10 | 2.55E-08 | IKZF3:ZPBP2:GSDMB:ORMDL3:GSDMA:PSMD3:CSF3:MED24 |
| his | Curated_gene_sets | NIKOLSKY_BREAST_CANCER_17Q11_Q21_AMPLICON | 132 | 19 | 1.03E-42 | 5.65E-39 | STAC2:FBXL20:MED1:CDK12:PPP1R1B:STARD3:TCAP:PNMT:PGAP3:ERBB2:MIEN1:IKZF3:ZPBP2:GSDMB:ORMDL3:GSDMA:PSMD3:CSF3:MED24 |
| his | Curated_gene_sets | FARMER_BREAST_CANCER_CLUSTER_8 | 6 | 5 | 5.91E-16 | 1.63E-12 | MED1:PNMT:PGAP3:ERBB2:GSDMB |
| his | Curated_gene_sets | SMID_BREAST_CANCER_ERBB2_UP | 148 | 8 | 6.01E-14 | 1.10E-10 | MED1:TCAP:PNMT:PGAP3:ERBB2:GSDMB:PSMD3:MED24 |
| hyp | GWAScatalog | L-arginine levels | 7 | 2 | 7.24E-07 | 0.001314 | CYP4V2:KLKB1 |
| hyp | GWAScatalog | Cardiac troponin-I levels | 19 | 2 | 5.89E-06 | 0.005344 | CYP4V2:KLKB1 |
| hyp | GWAScatalog | Loneliness (linear analysis) | 24 | 2 | 9.50E-06 | 0.005748 | SLC25A51:SHB |
| hyp | GWAScatalog | Type 2 diabetes (age of onset) | 47 | 2 | 3.71E-05 | 0.016847 | SLC25A51:SHB |
| hyp | Immunologic_signatures | GSE5589_UNSTIM_VS_45MIN_LPS_STIM_MACROPHAGE_UP | 196 | 3 | 6.01E-06 | 0.015088 | FAM149A:KLKB1:SHB |
| hyp | Immunologic_signatures | GSE36078_UNTREATED_VS_AD5_INF_MOUSE_LUNG_DC_UP | 198 | 3 | 6.19E-06 | 0.015088 | DNA2:TET1:FAM149A |
| hyp | Positional_gene_sets | chr4q35 | 91 | 3 | 5.96E-07 | 0.000178 | FAM149A:CYP4V2:KLKB1 |
| hyp | Positional_gene_sets | chr10q21 | 95 | 2 | 0.000153 | 0.022822 | DNA2:TET1 |
| hyp | Positional_gene_sets | chr9p13 | 148 | 2 | 0.00037 | 0.036881 | SLC25A51:SHB |
| leu | TF_targets | AGCYRWTTC_UNKNOWN | 120 | 2 | 1.17E-05 | 0.006916 | KIRREL3:PPARG |
| leu | TF_targets | TCF11MAFG_01 | 204 | 2 | 3.40E-05 | 0.006916 | KIRREL3:PPARG |
| leu | TF_targets | HFH4_01 | 204 | 2 | 3.40E-05 | 0.006916 | KIRREL3:PPARG |
| omna | GWAScatalog | Body mass index | 1358 | 5 | 1.74E-06 | 0.003164 | TMEM163:ACMSD:CCNT2:UBXN4:MCM6 |
| omna | GWAScatalog | Parkinson's disease | 166 | 3 | 3.65E-06 | 0.003259 | TMEM163:ACMSD:CCNT2 |
| omna | GWAScatalog | Diisocyanate-induced asthma | 189 | 3 | 5.39E-06 | 0.003259 | TMEM163:ACMSD:CCNT2 |
| omna | Positional_gene_sets | chr2q21 | 114 | 7 | 3.29E-18 | 9.85E-16 | TMEM163:CCNT2-AS1:ACMSD:MIR5590:CCNT2:UBXN4:MCM6 |
| pohhip | GWAScatalog | Alzheimer's disease or HDL levels (pleiotropy) | 59 | 3 | 9.15E-08 | 0.000166 | ZYX:EPHA1:EPHA1-AS1 |
| pohhip | Positional_gene_sets | chr7q35 | 57 | 2 | 3.92E-05 | 0.011709 | EPHA1:EPHA1-AS1 |
| prgly | GWAScatalog | Plasma renin activity levels | 7 | 2 | 5.17E-07 | 0.000939 | SNN:TXNDC11 |
| probet | Positional_gene_sets | chr10q22 | 173 | 2 | 2.44E-05 | 0.007307 | MCU:OIT3 |
| quina | Positional_gene_sets | chr1p31 | 187 | 2 | 2.86E-05 | 0.008542 | MIR3671:MIR101-1 |
| thre | GWAScatalog | Ulcerative colitis | 366 | 14 | 1.01E-25 | 1.82E-22 | STAC2:FBXL20:MED1:CDK12:PPP1R1B:STARD3:TCAP:PNMT:PGAP3:ERBB2:ZPBP2:GSDMB:ORMDL3:GSDMA |
| thre | GWAScatalog | Bronchial hyperresponsiveness in asthma | 8 | 6 | 1.38E-19 | 1.25E-16 | CDK12:PNMT:PGAP3:GSDMB:ORMDL3:GSDMA |
| thre | GWAScatalog | Asthma or allergic disease (pleiotropy) | 161 | 9 | 1.78E-17 | 1.08E-14 | STARD3:TCAP:PNMT:PGAP3:ERBB2:ZPBP2:GSDMB:ORMDL3:GSDMA |
| thre | GWAScatalog | Pancreatic ductal adenocarcinoma | 22 | 6 | 3.67E-16 | 1.66E-13 | STARD3:TCAP:PNMT:PGAP3:ERBB2:MED24 |
| thre | GWAScatalog | Adult asthma | 27 | 4 | 6.72E-10 | 2.44E-07 | ERBB2:ZPBP2:GSDMB:ORMDL3 |
| thre | GWAScatalog | Asthma | 315 | 6 | 5.87E-09 | 1.78E-06 | PGAP3:ERBB2:ZPBP2:GSDMB:ORMDL3:GSDMA |
| thre | GWAScatalog | White blood cell count | 206 | 5 | 3.99E-08 | 1.03E-05 | GSDMB:ORMDL3:GSDMA:CSF3:MED24 |
| thre | GWAScatalog | Primary biliary cirrhosis | 44 | 3 | 1.26E-06 | 0.000285 | ZPBP2:GSDMB:ORMDL3 |
| thre | GWAScatalog | Asthma (age of onset) | 48 | 3 | 1.64E-06 | 0.00033 | ZPBP2:GSDMB:ORMDL3 |
| thre | GWAScatalog | Systemic sclerosis | 55 | 3 | 2.48E-06 | 0.00045 | GSDMB:ORMDL3:GSDMA |
| thre | GWAScatalog | Rheumatoid arthritis | 208 | 4 | 2.75E-06 | 0.000453 | MED1:GSDMB:ORMDL3:CSF3 |
| thre | GWAScatalog | Asthma (childhood onset) | 255 | 4 | 6.15E-06 | 0.00093 | ZPBP2:GSDMB:ORMDL3:GSDMA |
| thre | GWAScatalog | Menopause (age at onset) | 93 | 3 | 1.21E-05 | 0.001693 | CDK12:STARD3:PGAP3 |
| thre | GWAScatalog | Primary biliary cholangitis | 98 | 3 | 1.42E-05 | 0.00184 | ZPBP2:GSDMB:ORMDL3 |
| thre | GWAScatalog | Hematological parameters | 14 | 2 | 2.03E-05 | 0.002451 | ORMDL3:GSDMA |
| thre | GWAScatalog | Fractional exhaled nitric oxide (childhood) | 19 | 2 | 3.80E-05 | 0.004312 | ZPBP2:GSDMB |
| thre | GWAScatalog | Triglyceride levels | 144 | 3 | 4.49E-05 | 0.004789 | CDK12:STARD3:PGAP3 |
| thre | GWAScatalog | Lung function (FVC) | 182 | 3 | 8.99E-05 | 0.009068 | FBXL20:MED1:CDK12 |
| thre | GWAScatalog | Mitochondrial DNA copy number | 31 | 2 | 0.000103 | 0.009503 | CSF3:MED24 |
| thre | GWAScatalog | Estimated glomerular filtration rate | 527 | 4 | 0.000105 | 0.009503 | MED1:CDK12:PPP1R1B:MED24 |
| thre | GWAScatalog | Crohn's disease | 600 | 4 | 0.000172 | 0.014898 | ZPBP2:GSDMB:ORMDL3:GSDMA |
| thre | GWAScatalog | Atrial fibrillation | 240 | 3 | 0.000203 | 0.016786 | ZPBP2:GSDMB:ORMDL3 |
| thre | GWAScatalog | Inflammatory bowel disease | 640 | 4 | 0.000221 | 0.01741 | ZPBP2:GSDMB:ORMDL3:GSDMA |
| thre | GWAScatalog | Bipolar disorder | 653 | 4 | 0.000238 | 0.018015 | ERBB2:GSDMA:CSF3:MED24 |
| thre | GWAScatalog | Subcutaneous adipose tissue | 62 | 2 | 0.000415 | 0.030147 | ZPBP2:GSDMB |
| thre | GWAScatalog | Systemic lupus erythematosus | 330 | 3 | 0.000517 | 0.036059 | MED1:ERBB2:GSDMB |
| thre | GWAScatalog | Rheumatoid arthritis (ACPA-positive) | 72 | 2 | 0.00056 | 0.037622 | GSDMB:ORMDL3 |
| thre | GWAScatalog | Glomerular filtration rate (creatinine) | 82 | 2 | 0.000725 | 0.047002 | FBXL20:CDK12 |
| thre | Chemical_and_Genetic_pertubation | NIKOLSKY_BREAST_CANCER_17Q11_Q21_AMPLICON | 132 | 16 | 1.16E-38 | 3.83E-35 | STAC2:FBXL20:MED1:CDK12:PPP1R1B:STARD3:TCAP:PNMT:PGAP3:ERBB2:ZPBP2:GSDMB:ORMDL3:GSDMA:CSF3:MED24 |
| thre | Chemical_and_Genetic_pertubation | FARMER_BREAST_CANCER_CLUSTER_8 | 6 | 5 | 8.61E-17 | 1.42E-13 | MED1:PNMT:PGAP3:ERBB2:GSDMB |
| thre | Chemical_and_Genetic_pertubation | SMID_BREAST_CANCER_ERBB2_UP | 148 | 7 | 4.02E-13 | 4.42E-10 | MED1:TCAP:PNMT:PGAP3:ERBB2:GSDMB:MED24 |
| thre | Chemical_and_Genetic_pertubation | SMID_BREAST_CANCER_BASAL_DN | 691 | 5 | 1.52E-05 | 0.012585 | TCAP:PNMT:PGAP3:ERBB2:MED24 |
| thre | Positional_gene_sets | chr17q12 | 125 | 11 | 9.65E-24 | 2.89E-21 | STAC2:FBXL20:MED1:CDK12:RNU6-233P:PPP1R1B:STARD3:TCAP:PNMT:PGAP3:ERBB2 |
| thre | Positional_gene_sets | chr17q21 | 399 | 6 | 2.40E-08 | 3.58E-06 | ZPBP2:GSDMB:ORMDL3:GSDMA:CSF3:MED24 |
| thre | GO_mf | GO_VITAMIN_D_RECEPTOR_BINDING | 15 | 2 | 2.34E-05 | 0.038443 | MED1:MED24 |
| thre | GO_mf | GO_THYROID_HORMONE_RECEPTOR_BINDING | 28 | 2 | 8.38E-05 | 0.04596 | MED1:MED24 |
| thre | GO_mf | GO_PHOSPHATIDYLINOSITOL_4_PHOSPHATE_BINDING | 28 | 2 | 8.38E-05 | 0.04596 | GSDMB:GSDMA |
| thre | Curated_gene_sets | NIKOLSKY_BREAST_CANCER_17Q11_Q21_AMPLICON | 132 | 16 | 1.16E-38 | 6.38E-35 | STAC2:FBXL20:MED1:CDK12:PPP1R1B:STARD3:TCAP:PNMT:PGAP3:ERBB2:ZPBP2:GSDMB:ORMDL3:GSDMA:CSF3:MED24 |
| thre | Curated_gene_sets | FARMER_BREAST_CANCER_CLUSTER_8 | 6 | 5 | 8.61E-17 | 2.37E-13 | MED1:PNMT:PGAP3:ERBB2:GSDMB |
| thre | Curated_gene_sets | SMID_BREAST_CANCER_ERBB2_UP | 148 | 7 | 4.02E-13 | 7.36E-10 | MED1:TCAP:PNMT:PGAP3:ERBB2:GSDMB:MED24 |
| thre | Curated_gene_sets | SMID_BREAST_CANCER_BASAL_DN | 691 | 5 | 1.52E-05 | 0.020966 | TCAP:PNMT:PGAP3:ERBB2:MED24 |
| trp | GWAScatalog | Ulcerative colitis | 366 | 16 | 1.09E-27 | 1.98E-24 | STAC2:FBXL20:MED1:CDK12:PPP1R1B:STARD3:TCAP:PNMT:PGAP3:ERBB2:MIEN1:IKZF3:ZPBP2:GSDMB:ORMDL3:GSDMA |
| trp | GWAScatalog | Asthma or allergic disease (pleiotropy) | 161 | 12 | 3.81E-23 | 3.46E-20 | STARD3:TCAP:PNMT:PGAP3:ERBB2:MIR4728:MIEN1:IKZF3:ZPBP2:GSDMB:ORMDL3:GSDMA |
| trp | GWAScatalog | Bronchial hyperresponsiveness in asthma | 8 | 7 | 1.09E-22 | 6.61E-20 | CDK12:PNMT:PGAP3:IKZF3:GSDMB:ORMDL3:GSDMA |
| trp | GWAScatalog | Pancreatic ductal adenocarcinoma | 22 | 6 | 2.21E-15 | 1.00E-12 | STARD3:TCAP:PNMT:PGAP3:ERBB2:MED24 |
| trp | GWAScatalog | Asthma | 315 | 8 | 1.16E-11 | 4.20E-09 | PGAP3:ERBB2:MIEN1:IKZF3:ZPBP2:GSDMB:ORMDL3:GSDMA |
| trp | GWAScatalog | Adult asthma | 27 | 4 | 2.06E-09 | 6.23E-07 | ERBB2:ZPBP2:GSDMB:ORMDL3 |
| trp | GWAScatalog | White blood cell count | 206 | 6 | 2.71E-09 | 7.04E-07 | GSDMB:ORMDL3:GSDMA:PSMD3:CSF3:MED24 |
| trp | GWAScatalog | Primary biliary cirrhosis | 44 | 4 | 1.58E-08 | 3.59E-06 | IKZF3:ZPBP2:GSDMB:ORMDL3 |
| trp | GWAScatalog | Rheumatoid arthritis | 208 | 5 | 1.74E-07 | 3.51E-05 | MED1:IKZF3:GSDMB:ORMDL3:CSF3 |
| trp | GWAScatalog | Primary biliary cholangitis | 98 | 4 | 4.12E-07 | 7.47E-05 | IKZF3:ZPBP2:GSDMB:ORMDL3 |
| trp | GWAScatalog | Mitochondrial DNA copy number | 31 | 3 | 9.67E-07 | 0.000159 | PSMD3:CSF3:MED24 |
| trp | GWAScatalog | Systemic lupus erythematosus | 330 | 5 | 1.69E-06 | 0.000256 | MED1:ERBB2:MIEN1:IKZF3:GSDMB |
| trp | GWAScatalog | Asthma (age of onset) | 48 | 3 | 3.69E-06 | 0.000516 | ZPBP2:GSDMB:ORMDL3 |
| trp | GWAScatalog | Systemic sclerosis | 55 | 3 | 5.59E-06 | 0.000724 | GSDMB:ORMDL3:GSDMA |
| trp | GWAScatalog | Rheumatoid arthritis (ACPA-positive) | 72 | 3 | 1.26E-05 | 0.001526 | IKZF3:GSDMB:ORMDL3 |
| trp | GWAScatalog | Asthma (childhood onset) | 255 | 4 | 1.84E-05 | 0.002084 | ZPBP2:GSDMB:ORMDL3:GSDMA |
| trp | GWAScatalog | Type 1 diabetes | 89 | 3 | 2.38E-05 | 0.002546 | IKZF3:GSDMB:ORMDL3 |
| trp | GWAScatalog | Menopause (age at onset) | 93 | 3 | 2.72E-05 | 0.002743 | CDK12:STARD3:PGAP3 |
| trp | GWAScatalog | Crohn's disease | 600 | 5 | 3.06E-05 | 0.002919 | IKZF3:ZPBP2:GSDMB:ORMDL3:GSDMA |
| trp | GWAScatalog | Hematological parameters | 14 | 2 | 3.44E-05 | 0.003119 | ORMDL3:GSDMA |
| trp | GWAScatalog | Inflammatory bowel disease | 640 | 5 | 4.15E-05 | 0.003591 | IKZF3:ZPBP2:GSDMB:ORMDL3:GSDMA |
| trp | GWAScatalog | Bipolar disorder | 653 | 5 | 4.57E-05 | 0.003771 | ERBB2:GSDMA:PSMD3:CSF3:MED24 |
| trp | GWAScatalog | Fractional exhaled nitric oxide (childhood) | 19 | 2 | 6.45E-05 | 0.004875 | ZPBP2:GSDMB |
| trp | GWAScatalog | White blood cell types | 19 | 2 | 6.45E-05 | 0.004875 | PSMD3:CSF3 |
| trp | GWAScatalog | Asthma and hay fever | 22 | 2 | 8.70E-05 | 0.006315 | IKZF3:GSDMA |
| trp | GWAScatalog | Triglyceride levels | 144 | 3 | 0.0001 | 0.006985 | CDK12:STARD3:PGAP3 |
| trp | GWAScatalog | Neutrophil count | 169 | 3 | 0.000161 | 0.010797 | PSMD3:CSF3:MED24 |
| trp | GWAScatalog | Lung function (FVC) | 182 | 3 | 0.0002 | 0.012951 | FBXL20:MED1:CDK12 |
| trp | GWAScatalog | Self-reported allergy | 34 | 2 | 0.00021 | 0.013161 | IKZF3:GSDMB |
| trp | GWAScatalog | Selective IgA deficiency | 39 | 2 | 0.000277 | 0.016772 | IKZF3:ORMDL3 |
| trp | GWAScatalog | Estimated glomerular filtration rate | 527 | 4 | 0.000303 | 0.017387 | MED1:CDK12:PPP1R1B:MED24 |
| trp | GWAScatalog | Polycystic ovary syndrome | 41 | 2 | 0.000307 | 0.017387 | ERBB2:IKZF3 |
| trp | GWAScatalog | Atrial fibrillation | 240 | 3 | 0.000449 | 0.024363 | ZPBP2:GSDMB:ORMDL3 |
| trp | GWAScatalog | Creatine kinase levels | 50 | 2 | 0.000456 | 0.024363 | PSMD3:CSF3 |
| trp | GWAScatalog | Subcutaneous adipose tissue | 62 | 2 | 0.000701 | 0.036366 | ZPBP2:GSDMB |
| trp | Chemical_and_Genetic_pertubation | NIKOLSKY_BREAST_CANCER_17Q11_Q21_AMPLICON | 132 | 20 | 1.83E-47 | 6.03E-44 | CACNB1:STAC2:FBXL20:MED1:CDK12:PPP1R1B:STARD3:TCAP:PNMT:PGAP3:ERBB2:MIEN1:IKZF3:ZPBP2:GSDMB:ORMDL3:GSDMA:PSMD3:CSF3:MED24 |
| trp | Chemical_and_Genetic_pertubation | FARMER_BREAST_CANCER_CLUSTER_8 | 6 | 5 | 3.66E-16 | 6.05E-13 | MED1:PNMT:PGAP3:ERBB2:GSDMB |
| trp | Chemical_and_Genetic_pertubation | SMID_BREAST_CANCER_ERBB2_UP | 148 | 8 | 2.63E-14 | 2.89E-11 | MED1:TCAP:PNMT:PGAP3:ERBB2:GSDMB:PSMD3:MED24 |
| trp | Chemical_and_Genetic_pertubation | LU_EZH2_TARGETS_UP | 281 | 4 | 2.69E-05 | 0.022172 | PPP1R1B:PGAP3:MIEN1:GSDMA |
| trp | Chemical_and_Genetic_pertubation | SMID_BREAST_CANCER_BASAL_DN | 691 | 5 | 5.98E-05 | 0.03947 | TCAP:PNMT:PGAP3:ERBB2:MED24 |
| trp | Positional_gene_sets | chr17q12 | 125 | 14 | 8.43E-30 | 2.52E-27 | CACNB1:STAC2:FBXL20:MED1:CDK12:RNU6-233P:PPP1R1B:STARD3:TCAP:PNMT:PGAP3:ERBB2:MIR4728:MIEN1 |
| trp | Positional_gene_sets | chr17q21 | 399 | 8 | 7.58E-11 | 1.13E-08 | IKZF3:ZPBP2:GSDMB:ORMDL3:GSDMA:PSMD3:CSF3:MED24 |
| trp | Curated_gene_sets | NIKOLSKY_BREAST_CANCER_17Q11_Q21_AMPLICON | 132 | 20 | 1.83E-47 | 1.00E-43 | CACNB1:STAC2:FBXL20:MED1:CDK12:PPP1R1B:STARD3:TCAP:PNMT:PGAP3:ERBB2:MIEN1:IKZF3:ZPBP2:GSDMB:ORMDL3:GSDMA:PSMD3:CSF3:MED24 |
| trp | Curated_gene_sets | FARMER_BREAST_CANCER_CLUSTER_8 | 6 | 5 | 3.66E-16 | 1.01E-12 | MED1:PNMT:PGAP3:ERBB2:GSDMB |
| trp | Curated_gene_sets | SMID_BREAST_CANCER_ERBB2_UP | 148 | 8 | 2.63E-14 | 4.82E-11 | MED1:TCAP:PNMT:PGAP3:ERBB2:GSDMB:PSMD3:MED24 |
| trp | Curated_gene_sets | LU_EZH2_TARGETS_UP | 281 | 4 | 2.69E-05 | 0.036938 | PPP1R1B:PGAP3:MIEN1:GSDMA |
| trp | TF_targets | NF1_Q6_01 | 275 | 4 | 2.47E-05 | 0.015059 | CACNB1:FBXL20:STARD3:ORMDL3 |
| trp | TF_targets | CTTTGA_LEF1_Q2 | 1235 | 6 | 8.90E-05 | 0.02567 | STAC2:PPP1R1B:IKZF3:ZPBP2:ORMDL3:GSDMA |
| trp | TF_targets | AACTTT_UNKNOWN | 1926 | 7 | 0.000126 | 0.02567 | CACNB1:STAC2:CDK12:PPP1R1B:ERBB2:IKZF3:CSF3 |
| trp | TF_targets | RREB1_01 | 210 | 3 | 0.000304 | 0.046377 | FBXL20:CDK12:PPP1R1B |
| tyr | GWAScatalog | Ulcerative colitis | 366 | 19 | 1.13E-28 | 2.05E-25 | STAC2:FBXL20:MED1:CDK12:PPP1R1B:STARD3:TCAP:PNMT:PGAP3:ERBB2:MIEN1:IKZF3:ZPBP2:GSDMB:ORMDL3:GSDMA:KIAA1919:REV3L:TRAF3IP2 |
| tyr | GWAScatalog | Asthma or allergic disease (pleiotropy) | 161 | 13 | 5.52E-22 | 5.01E-19 | EVI5:STARD3:TCAP:PNMT:PGAP3:ERBB2:MIR4728:MIEN1:IKZF3:ZPBP2:GSDMB:ORMDL3:GSDMA |
| tyr | GWAScatalog | Bronchial hyperresponsiveness in asthma | 8 | 7 | 5.34E-21 | 3.23E-18 | CDK12:PNMT:PGAP3:IKZF3:GSDMB:ORMDL3:GSDMA |
| tyr | GWAScatalog | Pancreatic ductal adenocarcinoma | 22 | 6 | 5.73E-14 | 2.60E-11 | STARD3:TCAP:PNMT:PGAP3:ERBB2:MED24 |
| tyr | GWAScatalog | Asthma | 315 | 8 | 9.80E-10 | 3.56E-07 | PGAP3:ERBB2:MIEN1:IKZF3:ZPBP2:GSDMB:ORMDL3:GSDMA |
| tyr | GWAScatalog | Inflammatory bowel disease | 640 | 9 | 1.34E-08 | 4.07E-06 | KIAA1107:IKZF3:ZPBP2:GSDMB:ORMDL3:GSDMA:KIAA1919:REV3L:TRAF3IP2 |
| tyr | GWAScatalog | Adult asthma | 27 | 4 | 1.65E-08 | 4.27E-06 | ERBB2:ZPBP2:GSDMB:ORMDL3 |
| tyr | GWAScatalog | Systemic lupus erythematosus | 330 | 7 | 4.19E-08 | 9.50E-06 | KIAA1107:EVI5:MED1:ERBB2:MIEN1:IKZF3:GSDMB |
| tyr | GWAScatalog | White blood cell count | 206 | 6 | 6.61E-08 | 1.33E-05 | GSDMB:ORMDL3:GSDMA:PSMD3:CSF3:MED24 |
| tyr | GWAScatalog | Primary biliary cirrhosis | 44 | 4 | 1.26E-07 | 2.28E-05 | IKZF3:ZPBP2:GSDMB:ORMDL3 |
| tyr | GWAScatalog | Rheumatoid arthritis | 208 | 5 | 2.33E-06 | 0.000384 | MED1:IKZF3:GSDMB:ORMDL3:CSF3 |
| tyr | GWAScatalog | Menopause (age at onset) | 93 | 4 | 2.61E-06 | 0.000395 | CDK12:STARD3:PGAP3:REV3L |
| tyr | GWAScatalog | Primary biliary cholangitis | 98 | 4 | 3.22E-06 | 0.000449 | IKZF3:ZPBP2:GSDMB:ORMDL3 |
| tyr | GWAScatalog | Mitochondrial DNA copy number | 31 | 3 | 4.44E-06 | 0.000576 | PSMD3:CSF3:MED24 |
| tyr | GWAScatalog | Asthma (age of onset) | 48 | 3 | 1.69E-05 | 0.002044 | ZPBP2:GSDMB:ORMDL3 |
| tyr | GWAScatalog | Systemic sclerosis | 55 | 3 | 2.55E-05 | 0.002892 | GSDMB:ORMDL3:GSDMA |
| tyr | GWAScatalog | Crohn's disease | 600 | 6 | 3.17E-05 | 0.00338 | KIAA1107:IKZF3:ZPBP2:GSDMB:ORMDL3:GSDMA |
| tyr | GWAScatalog | Rheumatoid arthritis (ACPA-positive) | 72 | 3 | 5.73E-05 | 0.005775 | IKZF3:GSDMB:ORMDL3 |
| tyr | GWAScatalog | Hematological parameters | 14 | 2 | 9.34E-05 | 0.008528 | ORMDL3:GSDMA |
| tyr | GWAScatalog | Inflammatory skin disease | 85 | 3 | 9.40E-05 | 0.008528 | KIAA1919:REV3L:TRAF3IP2 |
| tyr | GWAScatalog | Type 1 diabetes | 89 | 3 | 0.000108 | 0.009312 | IKZF3:GSDMB:ORMDL3 |
| tyr | GWAScatalog | Asthma (childhood onset) | 255 | 4 | 0.000137 | 0.011262 | ZPBP2:GSDMB:ORMDL3:GSDMA |
| tyr | GWAScatalog | White blood cell types | 19 | 2 | 0.000175 | 0.013236 | PSMD3:CSF3 |
| tyr | GWAScatalog | Fractional exhaled nitric oxide (childhood) | 19 | 2 | 0.000175 | 0.013236 | ZPBP2:GSDMB |
| tyr | GWAScatalog | Estimated glomerular filtration rate | 527 | 5 | 0.000197 | 0.014196 | RP11-345M22.1:MED1:CDK12:PPP1R1B:MED24 |
| tyr | GWAScatalog | Allergic disease (asthma, hay fever or eczema) | 283 | 4 | 0.000203 | 0.014196 | EVI5:FAM69A:GSDMB:PSMD3 |
| tyr | GWAScatalog | Asthma and hay fever | 22 | 2 | 0.000236 | 0.015862 | IKZF3:GSDMA |
| tyr | GWAScatalog | Triglyceride levels | 144 | 3 | 0.000445 | 0.028828 | CDK12:STARD3:PGAP3 |
| tyr | GWAScatalog | Bipolar disorder | 653 | 5 | 0.000527 | 0.032997 | ERBB2:GSDMA:PSMD3:CSF3:MED24 |
| tyr | GWAScatalog | Self-reported allergy | 34 | 2 | 0.000569 | 0.034401 | IKZF3:GSDMB |
| tyr | GWAScatalog | Neutrophil count | 169 | 3 | 0.000709 | 0.041482 | PSMD3:CSF3:MED24 |
| tyr | GWAScatalog | Selective IgA deficiency | 39 | 2 | 0.000749 | 0.042462 | IKZF3:ORMDL3 |
| tyr | GWAScatalog | Polycystic ovary syndrome | 41 | 2 | 0.000827 | 0.045506 | ERBB2:IKZF3 |
| tyr | GWAScatalog | Lung function (FVC) | 182 | 3 | 0.000878 | 0.046867 | FBXL20:MED1:CDK12 |
| tyr | Chemical_and_Genetic_pertubation | NIKOLSKY_BREAST_CANCER_17Q11_Q21_AMPLICON | 132 | 21 | 1.36E-42 | 4.49E-39 | CACNB1:RPL19:STAC2:FBXL20:MED1:CDK12:PPP1R1B:STARD3:TCAP:PNMT:PGAP3:ERBB2:MIEN1:IKZF3:ZPBP2:GSDMB:ORMDL3:GSDMA:PSMD3:CSF3:MED24 |
| tyr | Chemical_and_Genetic_pertubation | FARMER_BREAST_CANCER_CLUSTER_8 | 6 | 5 | 5.24E-15 | 8.66E-12 | MED1:PNMT:PGAP3:ERBB2:GSDMB |
| tyr | Chemical_and_Genetic_pertubation | SMID_BREAST_CANCER_ERBB2_UP | 148 | 8 | 2.37E-12 | 2.61E-09 | MED1:TCAP:PNMT:PGAP3:ERBB2:GSDMB:PSMD3:MED24 |
| tyr | Chemical_and_Genetic_pertubation | MIDORIKAWA_AMPLIFIED_IN_LIVER_CANCER | 55 | 3 | 2.55E-05 | 0.021048 | CACNB1:RPL19:FBXL20 |
| tyr | Chemical_and_Genetic_pertubation | SMID_BREAST_CANCER_BASAL_DN | 691 | 6 | 6.93E-05 | 0.045753 | AGL:TCAP:PNMT:PGAP3:ERBB2:MED24 |
| tyr | Positional_gene_sets | chr17q12 | 125 | 15 | 4.49E-28 | 1.34E-25 | CACNB1:RPL19:STAC2:FBXL20:MED1:CDK12:RNU6-233P:PPP1R1B:STARD3:TCAP:PNMT:PGAP3:ERBB2:MIR4728:MIEN1 |
| tyr | Positional_gene_sets | chr17q21 | 399 | 8 | 6.23E-09 | 9.32E-07 | IKZF3:ZPBP2:GSDMB:ORMDL3:GSDMA:PSMD3:CSF3:MED24 |
| tyr | Positional_gene_sets | chr6q21 | 105 | 4 | 4.23E-06 | 0.000395 | KIAA1919:REV3L:TRAF3IP2-AS1:TRAF3IP2:C6orf3 |
| tyr | Positional_gene_sets | chr1p22 | 111 | 4 | 5.28E-06 | 0.000395 | EPHX4:RPAP2:EVI5:FAM69A |
| tyr | Curated_gene_sets | NIKOLSKY_BREAST_CANCER_17Q11_Q21_AMPLICON | 132 | 21 | 1.36E-42 | 7.47E-39 | CACNB1:RPL19:STAC2:FBXL20:MED1:CDK12:PPP1R1B:STARD3:TCAP:PNMT:PGAP3:ERBB2:MIEN1:IKZF3:ZPBP2:GSDMB:ORMDL3:GSDMA:PSMD3:CSF3:MED24 |
| tyr | Curated_gene_sets | FARMER_BREAST_CANCER_CLUSTER_8 | 6 | 5 | 5.24E-15 | 1.44E-11 | MED1:PNMT:PGAP3:ERBB2:GSDMB |
| tyr | Curated_gene_sets | SMID_BREAST_CANCER_ERBB2_UP | 148 | 8 | 2.37E-12 | 4.34E-09 | MED1:TCAP:PNMT:PGAP3:ERBB2:GSDMB:PSMD3:MED24 |
| tyr | Curated_gene_sets | MIDORIKAWA_AMPLIFIED_IN_LIVER_CANCER | 55 | 3 | 2.55E-05 | 0.035066 | CACNB1:RPL19:FBXL20 |
| val | GWAScatalog | Ulcerative colitis | 366 | 13 | 1.38E-19 | 2.51E-16 | STAC2:FBXL20:MED1:CDK12:PPP1R1B:STARD3:TCAP:PNMT:PGAP3:ERBB2:GSDMB:ORMDL3:GSDMA |
| val | GWAScatalog | Bronchial hyperresponsiveness in asthma | 8 | 6 | 2.57E-18 | 2.33E-15 | CDK12:PNMT:PGAP3:GSDMB:ORMDL3:GSDMA |
| val | GWAScatalog | Pancreatic ductal adenocarcinoma | 22 | 6 | 6.80E-15 | 4.11E-12 | STARD3:TCAP:PNMT:PGAP3:ERBB2:MED24 |
| val | GWAScatalog | Asthma or allergic disease (pleiotropy) | 161 | 8 | 2.51E-13 | 1.14E-10 | STARD3:TCAP:PNMT:PGAP3:ERBB2:GSDMB:ORMDL3:GSDMA |
| val | GWAScatalog | White blood cell count | 206 | 5 | 4.06E-07 | 0.000147 | GSDMB:ORMDL3:GSDMA:CSF3:MED24 |
| val | GWAScatalog | Adult asthma | 27 | 3 | 1.06E-06 | 0.000321 | ERBB2:GSDMB:ORMDL3 |
| val | GWAScatalog | Estimated glomerular filtration rate | 527 | 6 | 2.05E-06 | 0.000533 | UBE2Q2:NRG4:MED1:CDK12:PPP1R1B:MED24 |
| val | GWAScatalog | Asthma | 315 | 5 | 3.27E-06 | 0.000742 | PGAP3:ERBB2:GSDMB:ORMDL3:GSDMA |
| val | GWAScatalog | Urate levels in obese individuals | 50 | 3 | 7.03E-06 | 0.001418 | UBE2Q2:FBXO22:NRG4 |
| val | GWAScatalog | Systemic sclerosis | 55 | 3 | 9.39E-06 | 0.001704 | GSDMB:ORMDL3:GSDMA |
| val | GWAScatalog | Rheumatoid arthritis | 208 | 4 | 1.65E-05 | 0.00273 | MED1:GSDMB:ORMDL3:CSF3 |
| val | GWAScatalog | Glomerular filtration rate (creatinine) | 82 | 3 | 3.13E-05 | 0.00473 | UBE2Q2:FBXL20:CDK12 |
| val | GWAScatalog | Menopause (age at onset) | 93 | 3 | 4.56E-05 | 0.006264 | CDK12:STARD3:PGAP3 |
| val | GWAScatalog | Hematological parameters | 14 | 2 | 4.83E-05 | 0.006264 | ORMDL3:GSDMA |
| val | GWAScatalog | Glomerular filtration rate | 106 | 3 | 6.73E-05 | 0.008146 | UBE2Q2:MED1:CDK12 |
| val | GWAScatalog | Creatinine levels | 128 | 3 | 0.000118 | 0.013368 | FBXO22:FBXL20:MED1 |
| val | GWAScatalog | Triglyceride levels | 144 | 3 | 0.000167 | 0.01782 | CDK12:STARD3:PGAP3 |
| val | GWAScatalog | Mitochondrial DNA copy number | 31 | 2 | 0.000245 | 0.024701 | CSF3:MED24 |
| val | GWAScatalog | Lung function (FVC) | 182 | 3 | 0.000332 | 0.031732 | FBXL20:MED1:CDK12 |
| val | GWAScatalog | Glomerular filtration rate in non diabetics (creatinine) | 38 | 2 | 0.000369 | 0.033501 | UBE2Q2:CDK12 |
| val | GWAScatalog | Primary biliary cirrhosis | 44 | 2 | 0.000495 | 0.042817 | GSDMB:ORMDL3 |
| val | GWAScatalog | Asthma (age of onset) | 48 | 2 | 0.00059 | 0.048644 | GSDMB:ORMDL3 |
| val | Chemical_and_Genetic_pertubation | NIKOLSKY_BREAST_CANCER_17Q11_Q21_AMPLICON | 132 | 15 | 1.52E-30 | 5.03E-27 | STAC2:FBXL20:MED1:CDK12:PPP1R1B:STARD3:TCAP:PNMT:PGAP3:ERBB2:GSDMB:ORMDL3:GSDMA:CSF3:MED24 |
| val | Chemical_and_Genetic_pertubation | FARMER_BREAST_CANCER_CLUSTER_8 | 6 | 5 | 9.15E-16 | 1.51E-12 | MED1:PNMT:PGAP3:ERBB2:GSDMB |
| val | Chemical_and_Genetic_pertubation | SMID_BREAST_CANCER_ERBB2_UP | 148 | 7 | 1.32E-11 | 1.45E-08 | MED1:TCAP:PNMT:PGAP3:ERBB2:GSDMB:MED24 |
| val | Chemical_and_Genetic_pertubation | NIKOLSKY_BREAST_CANCER_7P15_AMPLICON | 11 | 2 | 2.92E-05 | 0.024139 | HOXA10:HOXA11 |
| val | Positional_gene_sets | chr17q12 | 125 | 11 | 5.86E-21 | 1.75E-18 | STAC2:FBXL20:MED1:CDK12:RNU6-233P:PPP1R1B:STARD3:TCAP:PNMT:PGAP3:ERBB2 |
| val | Positional_gene_sets | chr15q24 | 103 | 6 | 1.25E-10 | 1.87E-08 | MAN2C1:SNUPN:MIR4313:UBE2Q2:FBXO22:NRG4 |
| val | Positional_gene_sets | chr7p15 | 86 | 4 | 4.93E-07 | 4.92E-05 | HOXA10-AS:HOXA10:HOXA11:HOXA11-AS |
| val | Positional_gene_sets | chr17q21 | 399 | 5 | 1.03E-05 | 0.000769 | GSDMB:ORMDL3:GSDMA:CSF3:MED24 |
| val | Curated_gene_sets | NIKOLSKY_BREAST_CANCER_17Q11_Q21_AMPLICON | 132 | 15 | 1.52E-30 | 8.38E-27 | STAC2:FBXL20:MED1:CDK12:PPP1R1B:STARD3:TCAP:PNMT:PGAP3:ERBB2:GSDMB:ORMDL3:GSDMA:CSF3:MED24 |
| val | Curated_gene_sets | FARMER_BREAST_CANCER_CLUSTER_8 | 6 | 5 | 9.15E-16 | 2.52E-12 | MED1:PNMT:PGAP3:ERBB2:GSDMB |
| val | Curated_gene_sets | SMID_BREAST_CANCER_ERBB2_UP | 148 | 7 | 1.32E-11 | 2.41E-08 | MED1:TCAP:PNMT:PGAP3:ERBB2:GSDMB:MED24 |
| val | Curated_gene_sets | NIKOLSKY_BREAST_CANCER_7P15_AMPLICON | 11 | 2 | 2.92E-05 | 0.038906 | HOXA10:HOXA11 |
| val | Curated_gene_sets | REACTOME_ERBB2_ACTIVATES_PTK6_SIGNALING | 13 | 2 | 4.14E-05 | 0.038906 | NRG4:ERBB2 |
| val | Curated_gene_sets | PID_ERBB_NETWORK_PATHWAY | 15 | 2 | 5.57E-05 | 0.038906 | NRG4:ERBB2 |
| val | Curated_gene_sets | REACTOME_ERBB2_REGULATES_CELL_MOTILITY | 15 | 2 | 5.57E-05 | 0.038906 | NRG4:ERBB2 |
| val | Curated_gene_sets | REACTOME_GRB2_EVENTS_IN_ERBB2_SIGNALING | 16 | 2 | 6.37E-05 | 0.038906 | NRG4:ERBB2 |
| val | Curated_gene_sets | REACTOME_PI3K_EVENTS_IN_ERBB2_SIGNALING | 16 | 2 | 6.37E-05 | 0.038906 | NRG4:ERBB2 |
| val | Curated_gene_sets | YAO_HOXA10_TARGETS_VIA_PROGESTERONE_DN | 19 | 2 | 9.06E-05 | 0.049828 | HOXA10:HOXA11 |
| val | Reactome | REACTOME_ERBB2_ACTIVATES_PTK6_SIGNALING | 13 | 2 | 4.14E-05 | 0.023854 | NRG4:ERBB2 |
| val | Reactome | REACTOME_ERBB2_REGULATES_CELL_MOTILITY | 15 | 2 | 5.57E-05 | 0.023854 | NRG4:ERBB2 |
| val | Reactome | REACTOME_GRB2_EVENTS_IN_ERBB2_SIGNALING | 16 | 2 | 6.37E-05 | 0.023854 | NRG4:ERBB2 |
| val | Reactome | REACTOME_PI3K_EVENTS_IN_ERBB2_SIGNALING | 16 | 2 | 6.37E-05 | 0.023854 | NRG4:ERBB2 |
| val | Reactome | REACTOME_SHC1_EVENTS_IN_ERBB2_SIGNALING | 22 | 2 | 0.000122 | 0.036634 | NRG4:ERBB2 |
| val | Reactome | REACTOME_DOWNREGULATION_OF_ERBB2_SIGNALING | 28 | 2 | 0.000199 | 0.049818 | NRG4:ERBB2 |
| val | Canonical_Pathways | REACTOME_ERBB2_ACTIVATES_PTK6_SIGNALING | 13 | 2 | 4.14E-05 | 0.027994 | NRG4:ERBB2 |
| val | Canonical_Pathways | PID_ERBB_NETWORK_PATHWAY | 15 | 2 | 5.57E-05 | 0.027994 | NRG4:ERBB2 |
| val | Canonical_Pathways | REACTOME_ERBB2_REGULATES_CELL_MOTILITY | 15 | 2 | 5.57E-05 | 0.027994 | NRG4:ERBB2 |
| val | Canonical_Pathways | REACTOME_GRB2_EVENTS_IN_ERBB2_SIGNALING | 16 | 2 | 6.37E-05 | 0.027994 | NRG4:ERBB2 |
| val | Canonical_Pathways | REACTOME_PI3K_EVENTS_IN_ERBB2_SIGNALING | 16 | 2 | 6.37E-05 | 0.027994 | NRG4:ERBB2 |
| val | Canonical_Pathways | REACTOME_SHC1_EVENTS_IN_ERBB2_SIGNALING | 22 | 2 | 0.000122 | 0.044784 | NRG4:ERBB2 |
| xan | GWAScatalog | Atopic march | 16 | 3 | 3.16E-10 | 5.74E-07 | PAQR8:EFHC1:TRAM2 |
| xan | Positional_gene_sets | chr6p12 | 107 | 4 | 8.36E-11 | 2.50E-08 | PAQR8:EFHC1:TRAM2:TRAM2-AS1 |
| xyl | GO_bp | GO_LIPID_GLYCOSYLATION | 12 | 2 | 3.03E-06 | 0.022286 | GBGT1:ABO |
| xyl | GWAScatalog | Coagulation factor levels | 8 | 4 | 7.93E-14 | 1.44E-10 | OBP2B:ABO:SURF6:MED22 |
| xyl | GWAScatalog | Blood protein levels | 1776 | 6 | 4.42E-07 | 0.000401 | OBP2B:ABO:SURF6:MED22:SURF1:REXO4 |
| xyl | GWAScatalog | Epithelial ovarian cancer | 26 | 2 | 1.49E-05 | 0.009019 | ABO:SURF6 |
| xyl | GWAScatalog | Serum alkaline phosphatase levels | 35 | 2 | 2.73E-05 | 0.012371 | ABO:SURF6 |
| xyl | GWAScatalog | Tonsillectomy | 58 | 2 | 7.55E-05 | 0.027421 | ABO:SURF6 |
| xyl | GWAScatalog | LDL cholesterol levels in current drinkers | 71 | 2 | 0.000113 | 0.034302 | ABO:SURF6 |
| xyl | GWAScatalog | Triglyceride levels x alcohol consumption (drinkers vs non-drinkers) interaction (2df) | 90 | 2 | 0.000182 | 0.037892 | ABO:SURF6 |
| xyl | GWAScatalog | Triglyceride levels x alcohol consumption (regular vs non-regular drinkers) interaction (2df) | 91 | 2 | 0.000186 | 0.037892 | ABO:SURF6 |
| xyl | GWAScatalog | LDL cholesterol levels x alcohol consumption (regular vs non-regular drinkers) interaction (2df) | 93 | 2 | 0.000195 | 0.037892 | ABO:SURF6 |
| xyl | GWAScatalog | LDL cholesterol levels x alcohol consumption (drinkers vs non-drinkers) interaction (2df) | 97 | 2 | 0.000212 | 0.037892 | ABO:SURF6 |
| xyl | GWAScatalog | LDL cholesterol levels | 101 | 2 | 0.00023 | 0.037892 | ABO:SURF6 |
| xyl | GWAScatalog | Allergic rhinitis | 113 | 2 | 0.000287 | 0.043464 | OBP2B:ABO |
| xyl | Positional_gene_sets | chr9q34 | 285 | 8 | 1.79E-17 | 5.37E-15 | GBGT1:OBP2B:ABO:SURF6:MED22:SURF1:REXO4:DBH-AS1 |
| xyl | Wikipathways | Globo Sphingolipid Metabolism | 22 | 2 | 1.06E-05 | 0.005756 | GBGT1:ABO |

### Supplementary Table 13

**FinnDiane physicians and nurses participating in the collection of the FinnDiane study subjects.**

| **The Finnish Diabetic Nephropathy Study Centers** | |
| --- | --- |
| Anjalankoski Health Center | S.Koivula, T.Uggeldahl |
| Central Finland Central Hospital, Jyväskylä | T.Forslund, A.Halonen, A.Koistinen, P.Koskiaho, M.Laukkanen, J.Saltevo, M.Tiihonen |
| Central Hospital of Åland Islands, Mariehamn | M.Forsen, H.Granlund, A.-C.Jonsson, B.Nyroos |
| Central Hospital of Kanta-Häme, Hämeenlinna | P.Kinnunen, A.Orvola, T.Salonen, A.Vähänen |
| Central Hospital of Kymenlaakso, Kotka | R.Paldanius, M.Riihelä, L.Ryysy |
| Central Hospital of Länsi-Pohja, Kemi | H.Laukkanen, P.Nyländen, A.Sademies |
| Central Ostrobothnian Hospital District, Kokkola | S.Anderson, B.Asplund, U.Byskata, P.Liedes, M.Kuusela, T.Virkkala |
| City of Espoo Health Center: |  |
| Espoonlahti | A.Nikkola, E.Ritola |
| Tapiola | M.Niska, H.Saarinen |
| Samaria | E.Oukko-Ruponen, T.Virtanen |
| Viherlaakso | A.Lyytinen |
| City of Helsinki Health Center: |  |
| Puistola | H.Kari, T.Simonen |
| Suutarila | A.Kaprio, J.Kärkkäinen, B.Rantaeskola |
| Töölö | P.Kääriäinen, J.Haaga, A-L.Pietiläinen |
| City of Hyvinkää Health Center | S.Klemetti, T.Nyandoto, E.Rontu, S.Satuli-Autere |
| City of Vantaa Health Center: |  |
| Korso | R.Toivonen, H.Virtanen |
| Länsimäki | R.Ahonen, M.Ivaska-Suomela, A.Jauhiainen |
| Martinlaakso | M.Laine, T.Pellonpää, R.Puranen |
| Myyrmäki | A.Airas, J.Laakso, K.Rautavaara |
| Rekola | M.Erola, E.Jatkola |
| Tikkurila | R.Lönnblad, A.Malm, J.Mäkelä, E.Rautamo |
| Heinola Health Center | P.Hentunen, J.Lagerstam |
| Helsinki University Central Hospital, Department of Medicine, Division of Nephrology | A.Ahola, M.Feodoroff, D.Gordin, O.Heikkilä, K.Hietala, M.Korolainen J.Kytö, S.Lindh, K.Pettersson-Fernholm, A.Sandelin, L.Thorn, J.Tuomikangas, T.Vesisenaho, J.Wadén |
| Herttoniemi Hospital, Helsinki | V.Sipilä |
| Hospital of Lounais-Häme, Forssa | T.Kalliomäki, J.Koskelainen, R.Nikkanen, N.Savolainen, H.Sulonen, E.Valtonen |
| Hyvinkää Hospital | L. Norvio, A. Hämäläinen |
| Iisalmi Hospital | E.Toivanen |
| Jokilaakso Hospital, Jämsä | A.Parta, I.Pirttiniemi |
| Jorvi Hospital, Helsinki University Central Hospital | S.Aranko, S.Ervasti, R.Kauppinen-Mäkelin, A.Kuusisto, T.Leppälä, K.Nikkilä, L.Pekkonen |
| Jyväskylä Health Center, Kyllö | K.Nuorva, M.Tiihonen |
| Kainuu Central Hospital, Kajaani | S.Jokelainen, K.Kananen, M.Karjalainen, P.Kemppainen, A-M.Mankinen, A.Reponen, M.Sankari |
| Kerava Health Center | H.Stuckey, P.Suominen |
| Kirkkonummi Health Center | A.Lappalainen, M.Liimatainen, J.Santaholma |
| Kivelä Hospital, Helsinki | A.Aimolahti, E.Huovinen |
| Koskela Hospital, Helsinki | V.Ilkka, M.Lehtimäki |
| Kotka Health Center | E.Pälikkö-Kontinen, A.Vanhanen |
| Kouvola Health Center | E.Koskinen, T.Siitonen |
| Kuopio University Hospital | E.Huttunen, R.Ikäheimo, P.Karhapää, P.Kekäläinen, M.Laakso, T.Lakka, E.Lampainen, L.Moilanen, L.Niskanen, U.Tuovinen, I.Vauhkonen, E.Voutilainen |
| Kuusamo Health Center | T.Kääriäinen, E.Isopoussu |
| Kuusankoski Hospital | E.Kilkki, I.Koskinen, L.Riihelä |
| Laakso Hospital, Helsinki | T.Meriläinen, P.Poukka, R.Savolainen, N.Uhlenius |
| Lahti City Hospital | A.Mäkelä, M.Tanner |
| Lapland Central Hospital, Rovaniemi | L.Hyvärinen, K.Lampela, S.Pöykkö, T.Rompasaari, S.Severinkangas, T.Tulokas |
| Lappeenranta Health Center | P. Erola, L. Härkönen, P.Linkola, I.Pulli, E.Repo |
| Lohja Hospital | T.Granlund, K.Hietanen, M.Porrassalmi, M.Saari, T.Salonen, M.Tiikkainen, |
| Länsi-Uusimaa Hospital, Tammisaari | I.-M.Jousmaa, J.Rinne |
| Loimaa Health Center | A.Mäkelä, P.Eloranta |
| Malmi Hospital, Helsinki | H.Lanki, S.Moilanen, M.Tilly-Kiesi |
| Mikkeli Central Hospital | A.Gynther, R.Manninen, P.Nironen, M.Salminen, T.Vänttinen |
| Mänttä Regional Hospital | I.Pirttiniemi, A-M.Hänninen |
| North Karelian Hospital, Joensuu | U-M.Henttula, P.Kekäläinen, M.Pietarinen, A.Rissanen, M.Voutilainen |
| Nurmijärvi Health Center | A.Burgos, K.Urtamo |
| Oulaskangas Hospital, Oulainen | E.Jokelainen, P-L.Jylkkä, E.Kaarlela, J.Vuolaspuro |
| Oulu Health Center | L.Hiltunen, R.Häkkinen, S.Keinänen-Kiukaanniemi |
| Oulu University Hospital | R.Ikäheimo |
| Päijät-Häme Central Hospital | H.Haapamäki, A.Helanterä, S.Hämäläinen, V.Ilvesmäki, H.Miettinen |
| Palokka Health Center | P.Sopanen, L.Welling |
| Pieksämäki Hospital | V.Sevtsenko, M.Tamminen |
| Pietarsaari Hospital | M-L.Holmbäck, B.Isomaa, L.Sarelin |
| Pori City Hospital | P.Ahonen, P.Merisalo, E.Muurinen, K.Sävelä |
| Porvoo Hospital | M.Kallio, B.Rask, S.Rämö |
| Raahe Hospital | A.Holma, M.Honkala, A.Tuomivaara, R.Vainionpää |
| Rauma Hospital | K.Laine, K.Saarinen, T.Salminen |
| Riihimäki Hospital | P.Aalto, E.Immonen, L.Juurinen |
| Salo Hospital | A.Alanko, J.Lapinleimu, P.Rautio, M.Virtanen |
| Satakunta Central Hospital, Pori | M.Asola, M.Juhola, P.Kunelius, M.-L.Lahdenmäki, P.Pääkkönen, M.Rautavirta |
| Savonlinna Central Hospital | T.Pulli, P.Sallinen, M.Taskinen, E.Tolvanen, T.Tuominen, H.Valtonen, A.Vartia, S-L.Viitanen |
| Seinäjoki Central Hospital | O.Antila, E.Korpi-Hyövälti, T.Latvala, E.Leijala, T.Leikkari, M.Punkari N.Rantamäki, H.Vähävuori |
| South Karelia Central Hospital, Lappeenranta | T.Ensala, E.Hussi, R.Härkönen, U.Nyholm, J.Toivanen |
| Tampere Health Center | A.Vaden, P.Alarotu, E.Kujansuu, H.Kirkkopelto-Jokinen, M.Helin, S.Gummerus, L.Calonius, T.Niskanen, T.Kaitala, T.Vatanen |
| Tampere University Hospital | I.Ala-Houhala, R.Kannisto, T.Kuningas, P.Lampinen, M.Määttä, H.Oksala, T.Oksanen, A.Putila, H.Saha, K.Salonen, H.Tauriainen, S.Tulokas |
| Tiirismaa Health Center, Hollola | T.Kivelä, L.Petlin, L.Savolainen |
| Turku Health Center | A.Artukka, I.Hämäläinen, L.Lehtinen, E.Pyysalo, H.Virtamo, M.Viinikkala, M.Vähätalo |
| Turku University Central Hospital | K.Breitholz, R.Eskola, K.Metsärinne, U.Pietilä, P.Saarinen, R.Tuominen, S.Äyräpää |
| Vaajakoski Health Center | K.Mäkinen, P.Sopanen |
| Valkeakoski Regional Hospital | S.Ojanen, E.Valtonen, H.Ylönen, M.Rautiainen, T.Immonen |
| Vammala Regional Hospital | I.Isomäki, R.Kroneld, L.Mustaniemi, M.Tapiolinna-Mäkelä |
| Vasa Central Hospital | S.Bergkulla, U.Hautamäki, V-A.Myllyniemi, I.Rusk |

### Supplementary Table 14

**Instruments for the urinary metabolites in the two sample MR analysis.** Variants (n=82) associated with metabolites in the COJO analysis with p < 5.0×10^-08^.

| **Metabolite** | **CHR:POS:EA:NEA** | **Rsid** | **EAF** | **Beta (SE)** | **P** | **N** |
| --- | --- | --- | --- | --- | --- | --- |
| 1-Methylnicotinamide | 2:135598913:A:G | rs17322446 | 0.183 | -0.13 (0.02) | 6.6×10^-10^ | 7619.42 |
| 2-Hydroxyisobutyrate | 12:122344302:A:G | rs1795967 | 0.169 | -0.51 (0.02) | 6.5×10^-120^ | 7821.75 |
| 3-Aminoisobutyrate | 1:155641153:A:C | rs186125941 | 0.018 | 0.40 (0.07) | 6.1×10^-09^ | 5379.59 |
| 3-Aminoisobutyrate | 5:34584621:A:C | rs16903139 | 0.914 | 0.23 (0.03) | 5.0×10^-14^ | 6255.84 |
| 3-Aminoisobutyrate | 5:34853162:T:C | rs338296 | 0.428 | 0.12 (0.02) | 8.0×10^-11^ | 6367.54 |
| 3-Aminoisobutyrate | 5:34868497:A:T | rs72732827 | 0.987 | -0.60 (0.08) | 1.2×10^-13^ | 5462.8 |
| 3-Aminoisobutyrate | 5:34896132:A:G | rs138425947 | 0.978 | -0.51 (0.07) | 5.4×10^-14^ | 4681.79 |
| 3-Aminoisobutyrate | 5:34899723:T:C | rs56007938 | 0.015 | 0.66 (0.08) | 1.1×10^-17^ | 5419.33 |
| 3-Aminoisobutyrate | 5:34911884:T:C | rs2308957 | 0.015 | -0.78 (0.09) | 3.2×10^-18^ | 4406.88 |
| 3-Aminoisobutyrate | 5:34982167:T:C | rs116116288 | 0.012 | 0.88 (0.09) | 8.2×10^-25^ | 5376.75 |
| 3-Aminoisobutyrate | 5:34993215:A:G | rs11744796 | 0.433 | -0.18 (0.02) | 9.5×10^-24^ | 6496.13 |
| 3-Aminoisobutyrate | 5:35000653:T:C | rs7737763 | 0.434 | -0.44 (0.03) | 2.5×10^-67^ | 6311.81 |
| 3-Aminoisobutyrate | 5:35003112:A:G | rs468327 | 0.218 | 0.36 (0.03) | 1.3×10^-33^ | 5887.7 |
| 3-Aminoisobutyrate | 5:35037115:T:C | rs37369 | 0.100 | 0.84 (0.04) | 4.3×10^-99^ | 5317.32 |
| 3-Aminoisobutyrate | 5:35039437:A:G | rs2279651 | 0.431 | 0.45 (0.02) | 7.5×10^-82^ | 6419.22 |
| 3-Aminoisobutyrate | 5:35152241:T:C | rs954286 | 0.939 | 0.26 (0.04) | 2.1×10^-12^ | 6488.8 |
| 3-Aminoisobutyrate | 5:36219710:T:C | rs138373837 | 0.019 | -0.39 (0.07) | 2.0×10^-09^ | 5829.69 |
| 3-Aminoisobutyrate | 12:345369:C:G | rs2080403 | 0.454 | -0.16 (0.02) | 3.5×10^-22^ | 6424.66 |
| 3-hydroxyhippurate | 7:17287106:A:G | rs6968554 | 0.352 | -0.09 (0.02) | 5.0×10^-08^ | 7803.63 |
| 3-Hydroxyisobutyrate | 9:107525165:T:G | rs2472479 | 0.565 | -0.16 (0.02) | 5.0×10^-22^ | 8108.34 |
| 3-Hydroxyisobutyrate | 11:26700078:A:G | rs73436381 | 0.081 | -0.18 (0.03) | 4.9×10^-09^ | 7630.99 |
| 3-Hydroxyisobutyrate | 12:101551386:T:C | rs2625153 | 0.250 | 0.11 (0.02) | 9.1×10^-09^ | 8000.98 |
| 3-Hydroxyisovalerate | 1:6334301:A:G | rs114200864 | 0.041 | -0.28 (0.05) | 2.9×10^-10^ | 6861.94 |
| 3-Hydroxyisovalerate | 1:151904146:A:T | rs2999545 | 0.385 | -0.11 (0.02) | 4.7×10^-12^ | 8464.04 |
| 3-Hydroxyisovalerate | 3:182758040:T:C | rs4859267 | 0.294 | 0.13 (0.02) | 1.3×10^-12^ | 8275.56 |
| 3-Hydroxyisovalerate | 6:160578860:T:C | rs1564348 | 0.836 | -0.12 (0.02) | 3.0×10^-08^ | 8199.21 |
| 3-Hydroxyisovalerate | 12:122326812:T:C | rs2230681 | 0.172 | -0.13 (0.02) | 5.2×10^-09^ | 8202.26 |
| 3-Hydroxyisovalerate | 13:31579587:A:G | rs73167015 | 0.948 | -0.22 (0.04) | 4.6×10^-09^ | 7742.19 |
| 3-Hydroxyisovalerate | 16:20557634:A:T | rs7499358 | 0.982 | -0.62 (0.06) | 8.7×10^-22^ | 7387.28 |
| 3-Hydroxyisovalerate | 16:20570661:T:C | rs8056693 | 0.030 | 0.30 (0.05) | 2.0×10^-09^ | 7371.99 |
| 3-Hydroxyisovalerate | 16:20608891:C:G | rs540815683 | 0.986 | -0.81 (0.11) | 1.9×10^-13^ | 3281.8 |
| 4-Deoxyerythronic acid | 2:241793545:A:G | rs55649245 | 0.326 | -0.17 (0.02) | 3.2×10^-20^ | 8058.43 |
| 4-Deoxyerythronic acid | 2:241813788:T:C | rs10933641 | 0.277 | 0.19 (0.02) | 2.6×10^-20^ | 7714.35 |
| 4-Deoxyerythronic acid | 8:74868909:A:G | rs72661850 | 0.691 | -0.12 (0.02) | 1.5×10^-11^ | 7975.79 |
| 4-Deoxythreonate | 4:22817242:A:G | rs181558 | 0.149 | 0.13 (0.02) | 5.3×10^-09^ | 8093.96 |
| 4-Deoxythreonate | 12:4521511:A:T | rs78470967 | 0.043 | 0.34 (0.04) | 2.0×10^-14^ | 6376.66 |
| Alanine | 2:227463256:A:T | rs10153528 | 0.924 | 0.19 (0.03) | 3.6×10^-08^ | 6545.3 |
| Alanine | 5:1188285:A:G | rs11133665 | 0.258 | -0.11 (0.02) | 3.5×10^-09^ | 8414.9 |
| Arabinose | 16:9178672:C:G | rs149644414 | 0.973 | 0.37 (0.07) | 2.1×10^-08^ | 4208.52 |
| Citrate | 17:6599874:T:G | rs111817161 | 0.957 | -0.25 (0.04) | 9.5×10^-09^ | 5856.25 |
| Citrate | 17:26824156:A:G | rs11567842 | 0.642 | -0.11 (0.02) | 3.0×10^-13^ | 8208.65 |
| Creatine | 1:10698598:A:G | rs12752101 | 0.024 | -0.64 (0.12) | 5.8×10^-08^ | 1524.4 |
| Ethanolamine | 4:109716840:A:T | rs62313082 | 0.379 | 0.14 (0.02) | 7.7×10^-16^ | 7163.84 |
| Formate | 1:11940483:T:C | rs4846068 | 0.585 | 0.13 (0.02) | 2.2×10^-15^ | 8192.68 |
| Formate | 8:18272377:T:C | rs4921913 | 0.770 | -0.32 (0.02) | 9.3×10^-64^ | 8124.69 |
| Glycine | 2:211540507:A:C | rs1047891 | 0.317 | 0.23 (0.02) | 2.9×10^-37^ | 7547.62 |
| Glycine | 5:1188285:A:G | rs11133665 | 0.261 | -0.13 (0.02) | 1.6×10^-13^ | 8231.81 |
| Glycine | 5:150624099:T:C | rs72794144 | 0.028 | 0.38 (0.06) | 2.5×10^-11^ | 7281.83 |
| Glycine | 5:150702299:A:G | rs61067578 | 0.842 | 0.24 (0.02) | 1.1×10^-24^ | 8306.36 |
| Glycine | 5:150708711:C:G | rs147000073 | 0.987 | -1.08 (0.11) | 1.7×10^-24^ | 3644.86 |
| Glycine | 5:150709611:A:C | rs78736052 | 0.946 | -0.21 (0.04) | 1.1×10^-08^ | 7967.11 |
| Glycine | 9:6571402:A:T | rs191428709 | 0.989 | -0.55 (0.10) | 1.0×10^-08^ | 5327.54 |
| Glycine | 9:6649491:T:C | rs62565993 | 0.124 | 0.20 (0.03) | 3.4×10^-14^ | 7118.76 |
| Glycolic acid | 2:27336827:A:C | rs12617392 | 0.419 | -0.10 (0.02) | 3.9×10^-08^ | 6312.89 |
| Glycolic acid | 3:38395562:T:C | rs3132440 | 0.653 | -0.11 (0.02) | 8.2×10^-09^ | 5973.43 |
| Histidine | 5:950299:C:G | rs61208283 | 0.056 | 0.35 (0.06) | 1.0×10^-09^ | 3181.83 |
| Histidine | 5:1188285:A:G | rs11133665 | 0.255 | -0.31 (0.02) | 1.9×10^-39^ | 5256.52 |
| Histidine | 17:37636695:T:G | rs4795371 | 0.243 | -0.22 (0.02) | 5.1×10^-18^ | 4641.92 |
| Propylene Glycol | 4:88213884:T:C | rs6811902 | 0.601 | -0.16 (0.02) | 2.8×10^-20^ | 5357.01 |
| Quinic acid | 7:17287998:A:G | rs2106727 | 0.353 | -0.12 (0.02) | 3.1×10^-13^ | 8295.39 |
| Quinic acid | 11:60748390:T:C | rs183169338 | 0.012 | 0.45 (0.08) | 3.6×10^-08^ | 6549.03 |
| Taurine | 22:18514138:T:C | rs149659516 | 0.039 | -0.36 (0.07) | 3.3×10^-08^ | 3134.16 |
| Threonine | 5:1188285:A:G | rs11133665 | 0.259 | -0.15 (0.02) | 1.3×10^-16^ | 8417.15 |
| Threonine | 17:37631883:C:G | rs11078902 | 0.241 | -0.12 (0.02) | 3.7×10^-10^ | 7570.43 |
| trans-Aconitate | 5:14574155:C:G | rs12109981 | 0.865 | -0.14 (0.03) | 4.5×10^-08^ | 6694.03 |
| Trigonelline | 7:17287998:A:G | rs2106727 | 0.355 | -0.10 (0.02) | 6.5×10^-11^ | 8640.36 |
| Tryptophan | 5:1188285:A:G | rs11133665 | 0.258 | -0.17 (0.02) | 1.4×10^-19^ | 8133.39 |
| Tryptophan | 5:1225613:A:G | rs7704058 | 0.796 | -0.15 (0.02) | 5.1×10^-14^ | 8213.15 |
| Tryptophan | 17:37633970:A:C | rs12453397 | 0.756 | 0.16 (0.02) | 7.2×10^-16^ | 7357.99 |
| Tyrosine | 1:155178782:A:T | rs760077 | 0.401 | -0.10 (0.02) | 1.8×10^-09^ | 8028.71 |
| Tyrosine | 5:950299:C:G | rs61208283 | 0.057 | 0.28 (0.05) | 4.2×10^-09^ | 4746.89 |
| Tyrosine | 5:1188285:A:G | rs11133665 | 0.259 | -0.30 (0.02) | 7.5×10^-55^ | 8233.76 |
| Tyrosine | 5:1225434:T:C | rs7704882 | 0.794 | -0.20 (0.02) | 4.6×10^-24^ | 8344.34 |
| Tyrosine | 5:1237205:A:C | rs4532396 | 0.907 | 0.19 (0.03) | 2.9×10^-09^ | 6542.61 |
| Tyrosine | 6:111492119:T:C | rs241768 | 0.294 | 0.11 (0.02) | 2.6×10^-10^ | 8178.53 |
| Tyrosine | 17:37633970:A:C | rs12453397 | 0.758 | 0.25 (0.02) | 5.7×10^-36^ | 7351.3 |
| Uracil | 1:97915614:T:C | rs3918290 | 0.018 | 0.40 (0.07) | 1.2×10^-08^ | 5767.08 |
| Valine | 5:1225613:A:G | rs7704058 | 0.797 | -0.11 (0.02) | 1.2×10^-08^ | 8308.13 |
| Valine | 9:10907333:C:G | rs142108776 | 0.012 | -0.48 (0.09) | 2.1×10^-08^ | 6447.16 |
| Valine | 15:76246609:T:G | rs11634260 | 0.763 | 0.12 (0.02) | 2.7×10^-08^ | 6859.09 |
| Xylose | 1:48690229:A:C | rs10788884 | 0.674 | 0.11 (0.02) | 1.1×10^-10^ | 7696.23 |
| Xylose | 9:136146597:T:C | rs550057 | 0.244 | 0.20 (0.02) | 8.9×10^-26^ | 7449.87 |

CHR:POS:EA:NEA: Chromosome position, effect allele, and non-effect allele. Rsid: variant rs-identifier. Metabolite: the associated urinary metabolite. EAF: effect allele frequency. Beta (SE): effect estimate for the effect allele (effect estimate standard deviation). P: p-value of the association. N: number of individuals in the analysis.

### Supplementary Figure 1

**Heritability estimates for the urinary metabolites.** The metabolites are order by the meta-analysis estimates of heritability. Full circle indicates p-value < 0.05 and * heterogeneity test p-value < 0.05.

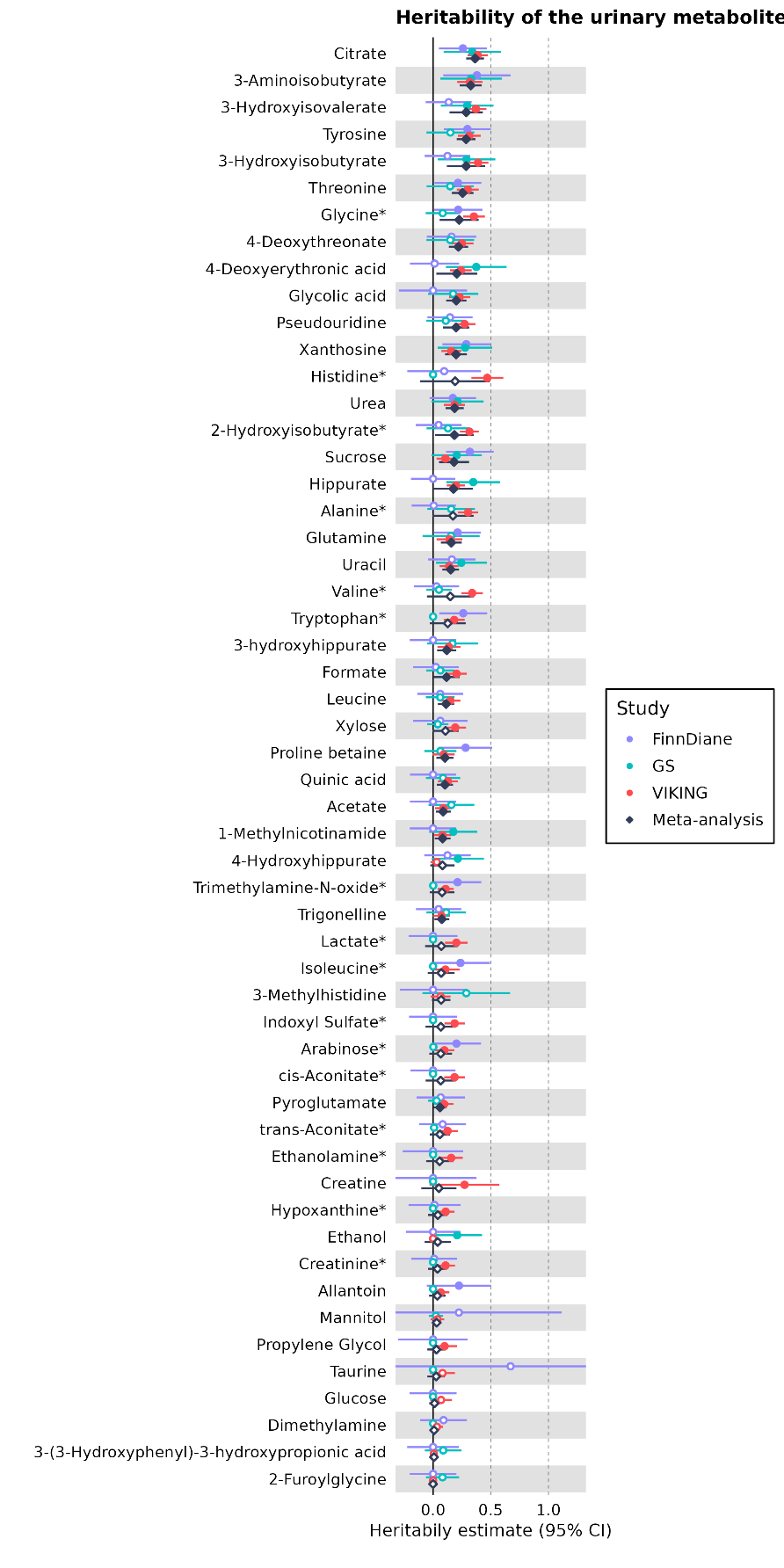

### Supplementary Figure 2

**LocusZoom plots of the COJO lead signals**. Association with urinary metabolite 250kb around the COJO lead variant for each of the 52 genome-wide significant associations. Plot title includes the metabolite name and the lead variant rsid (chromosome:position:effect allele:non-effect allele). LD information was calculated using the 1000 Genomes Phase 3 European population.

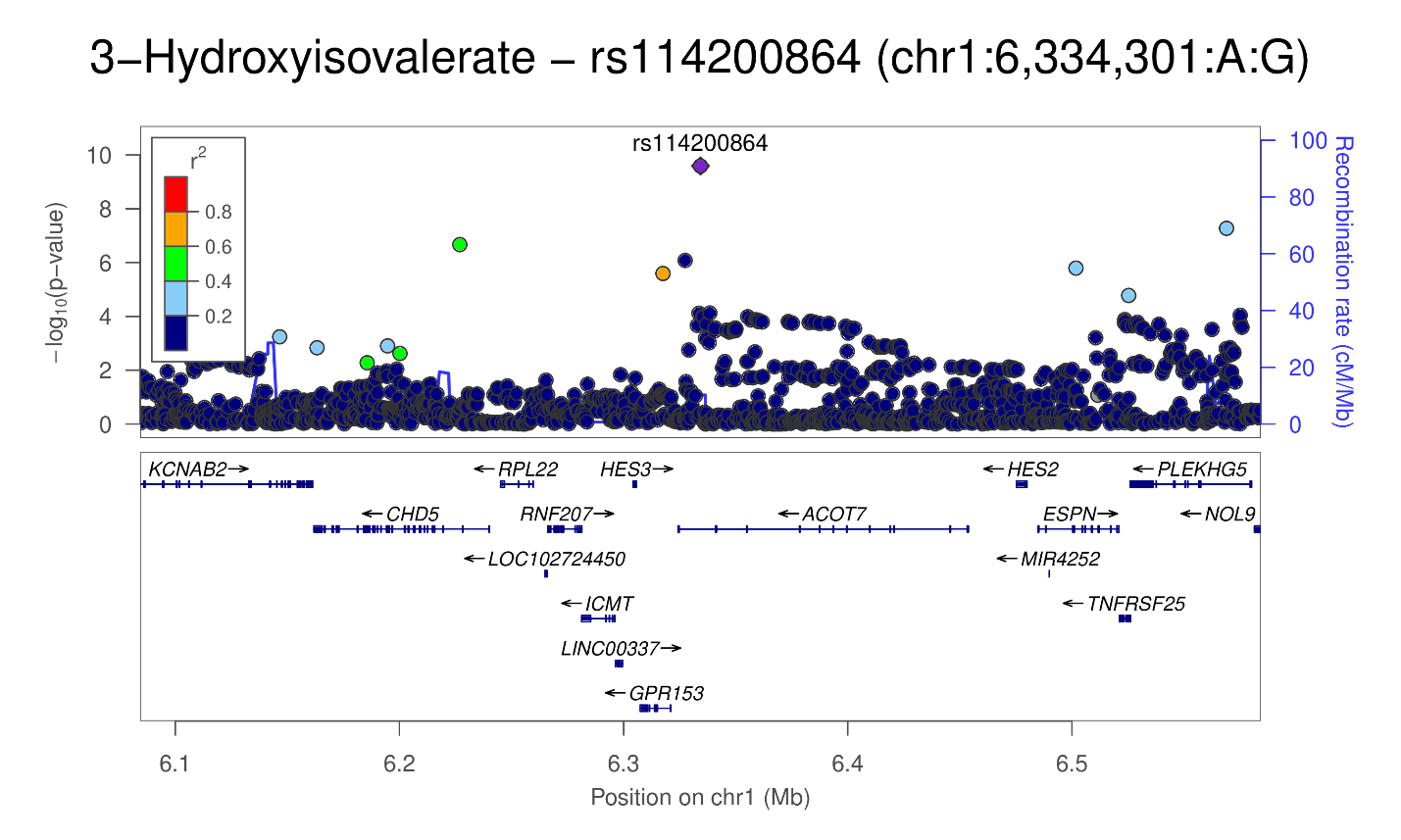

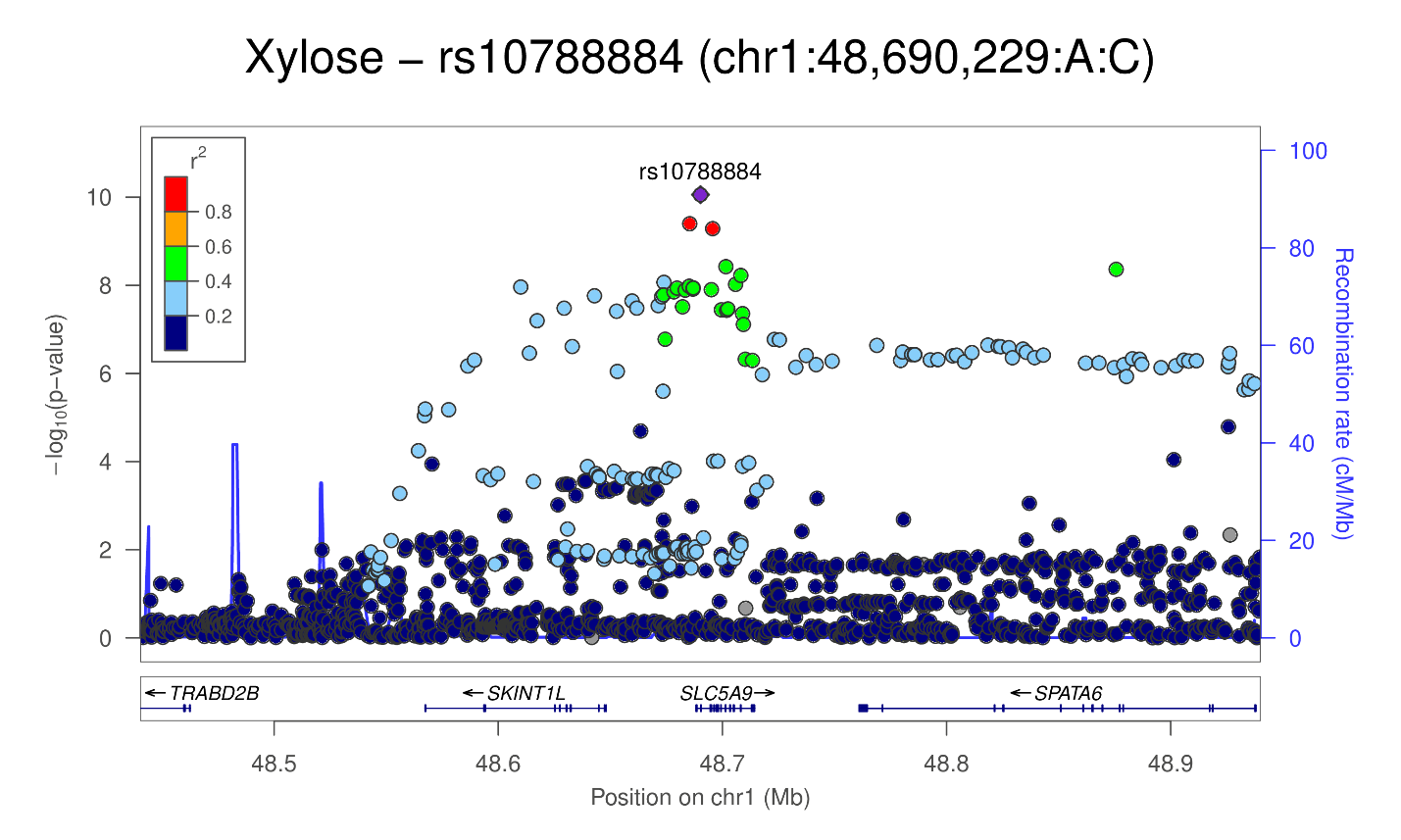

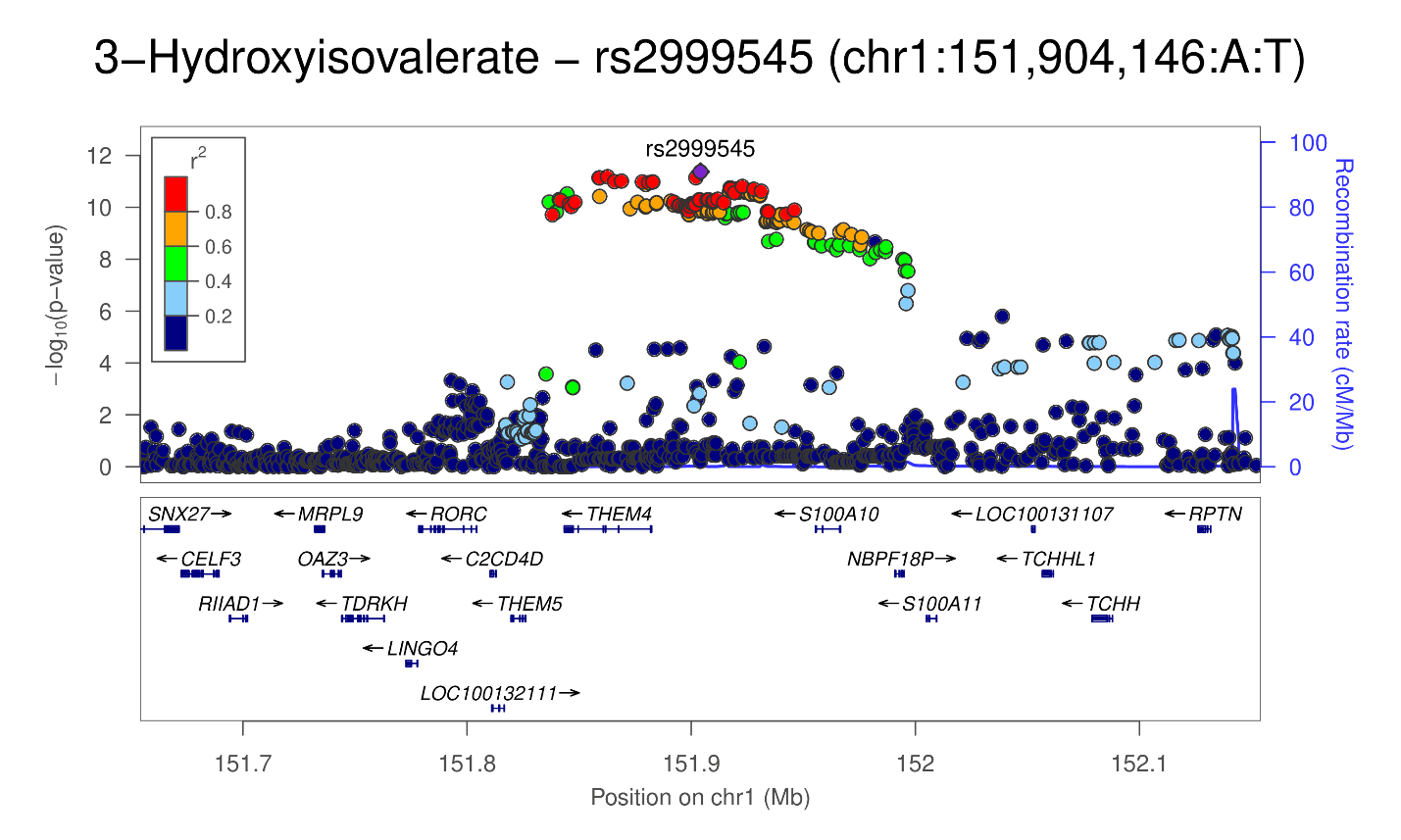

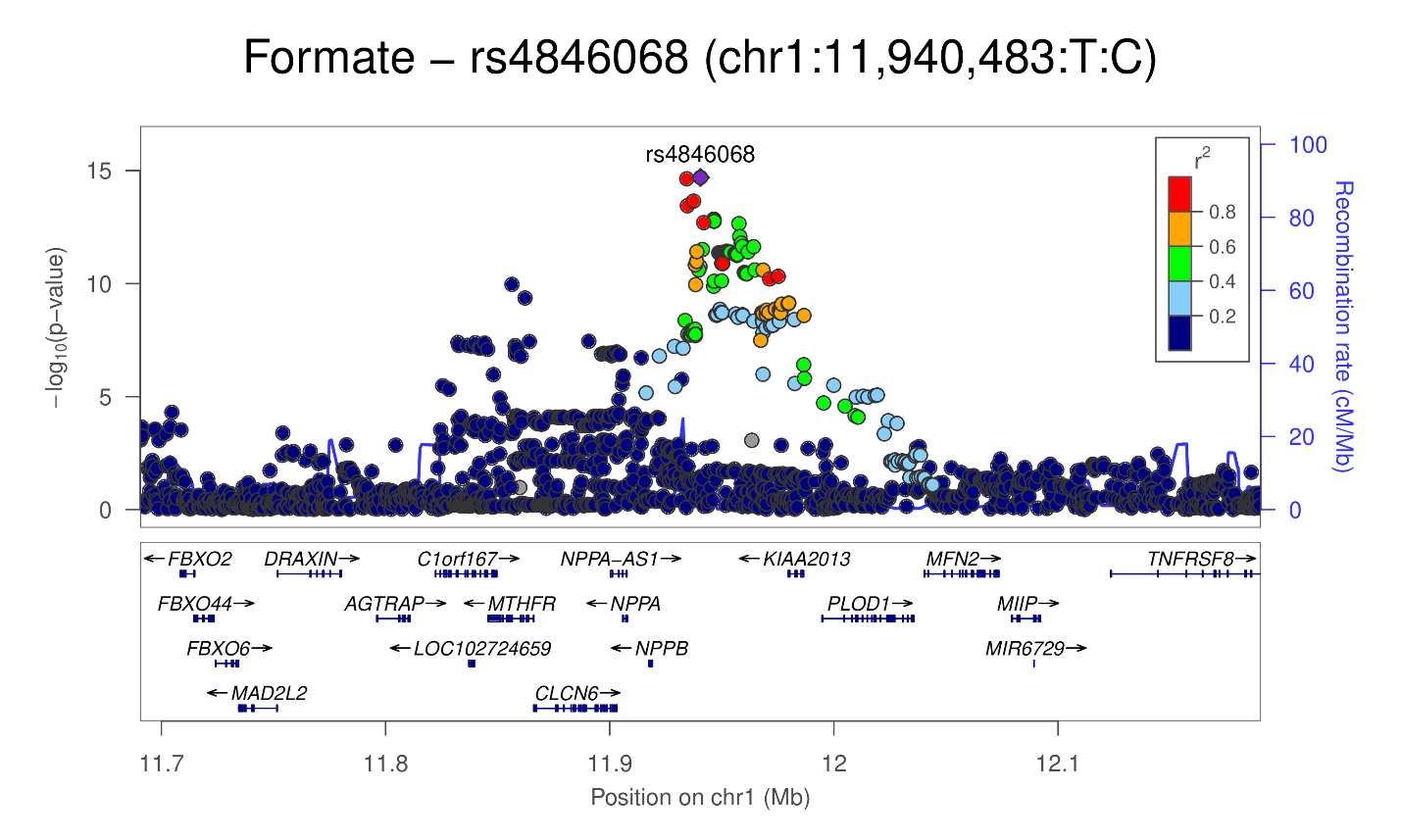

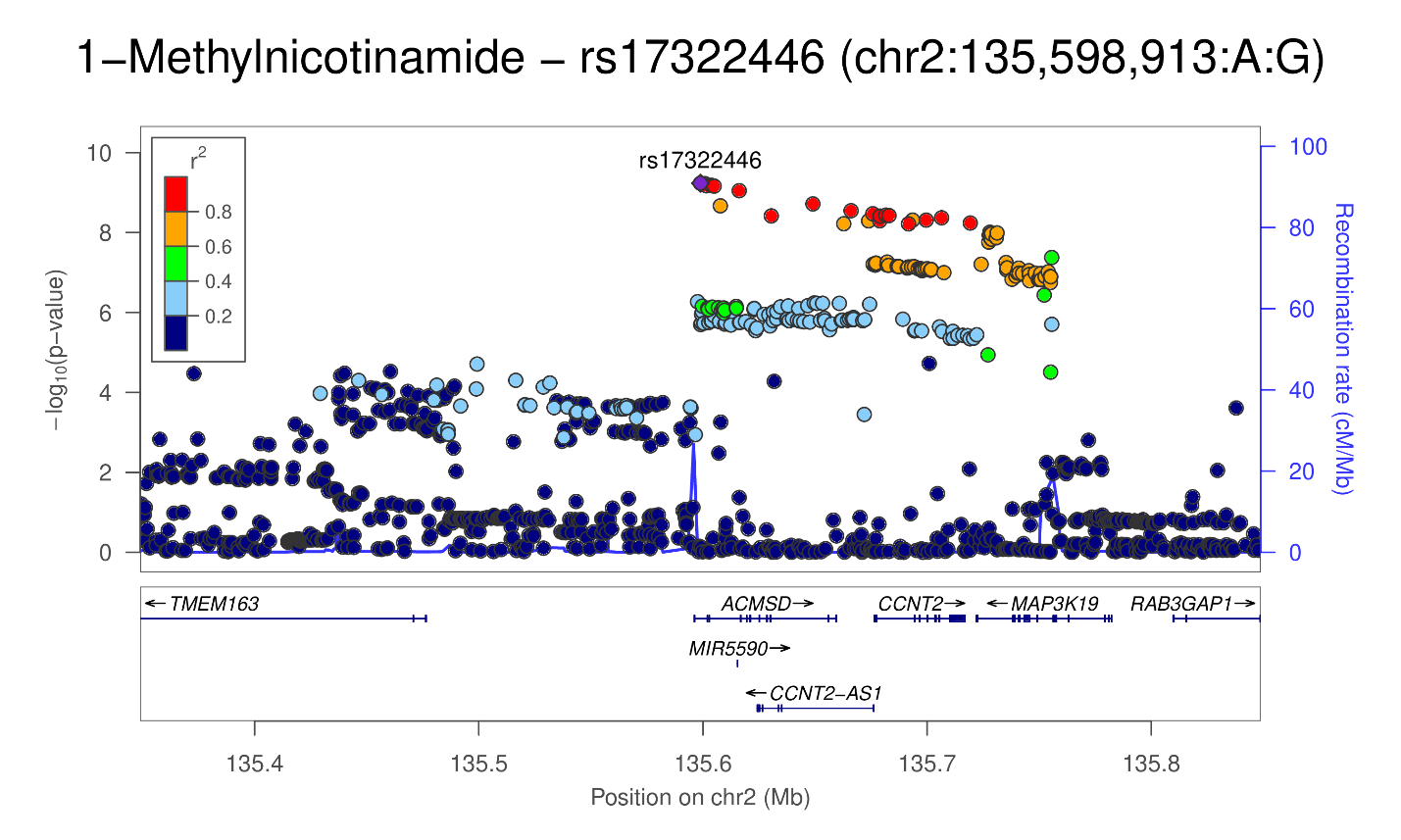

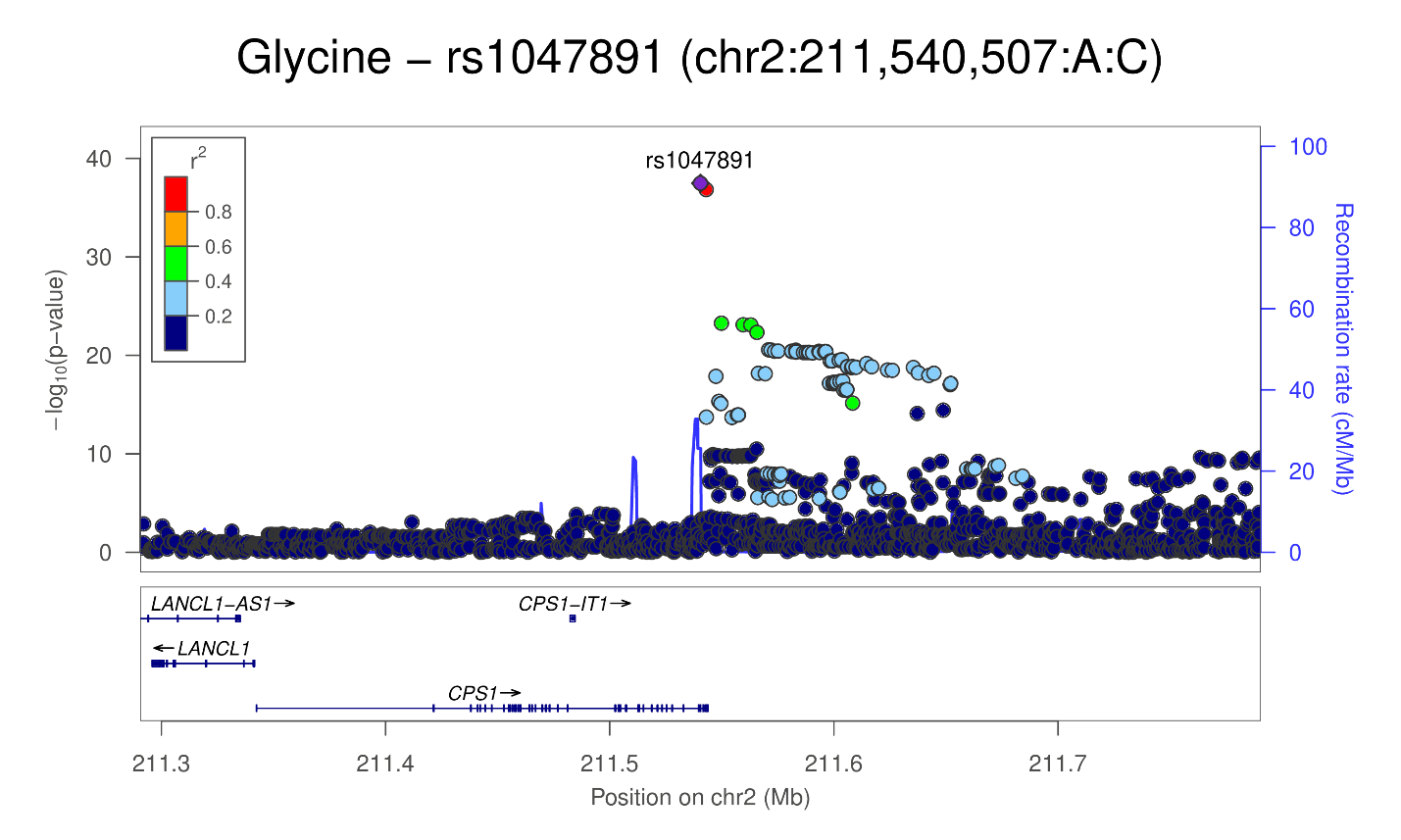

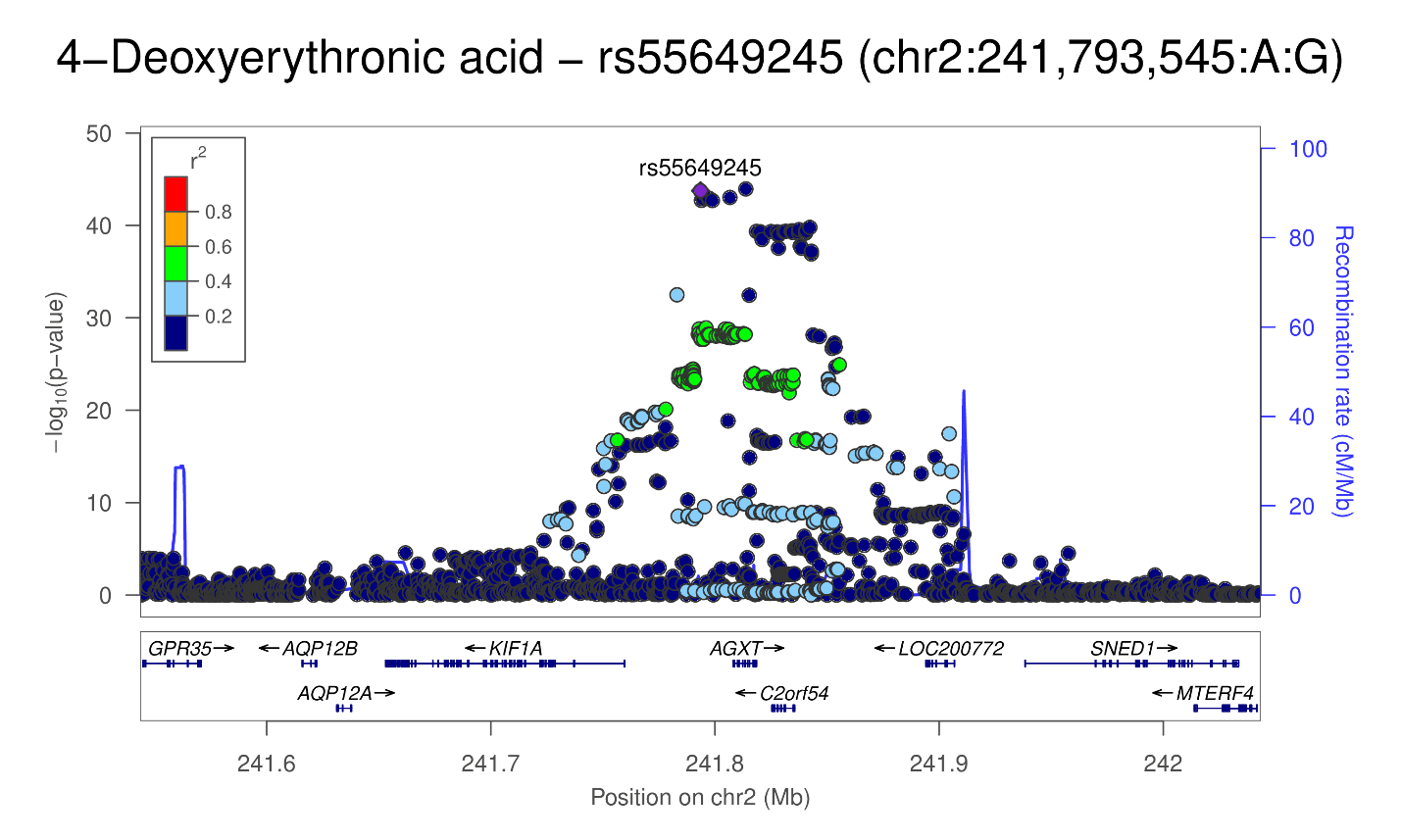

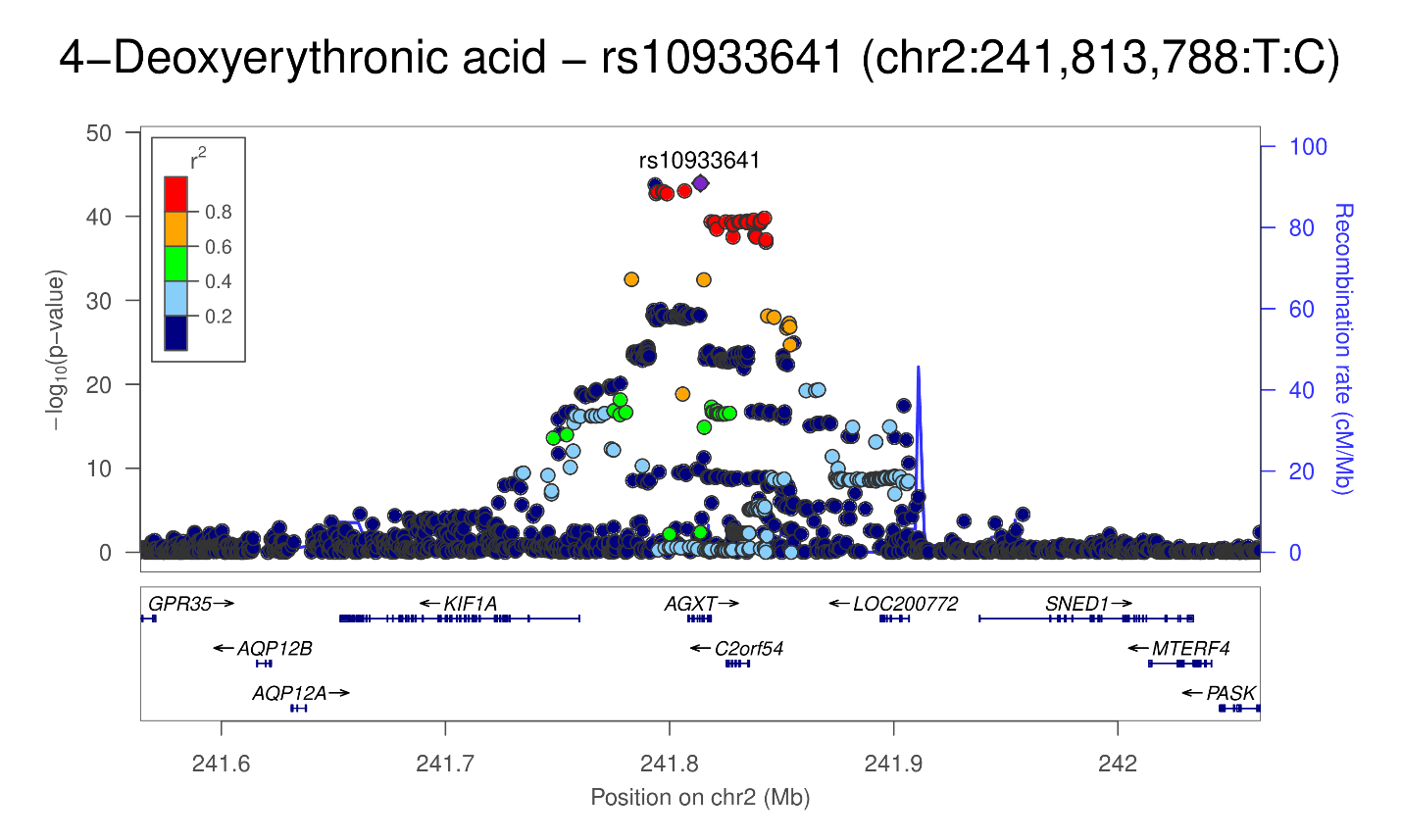

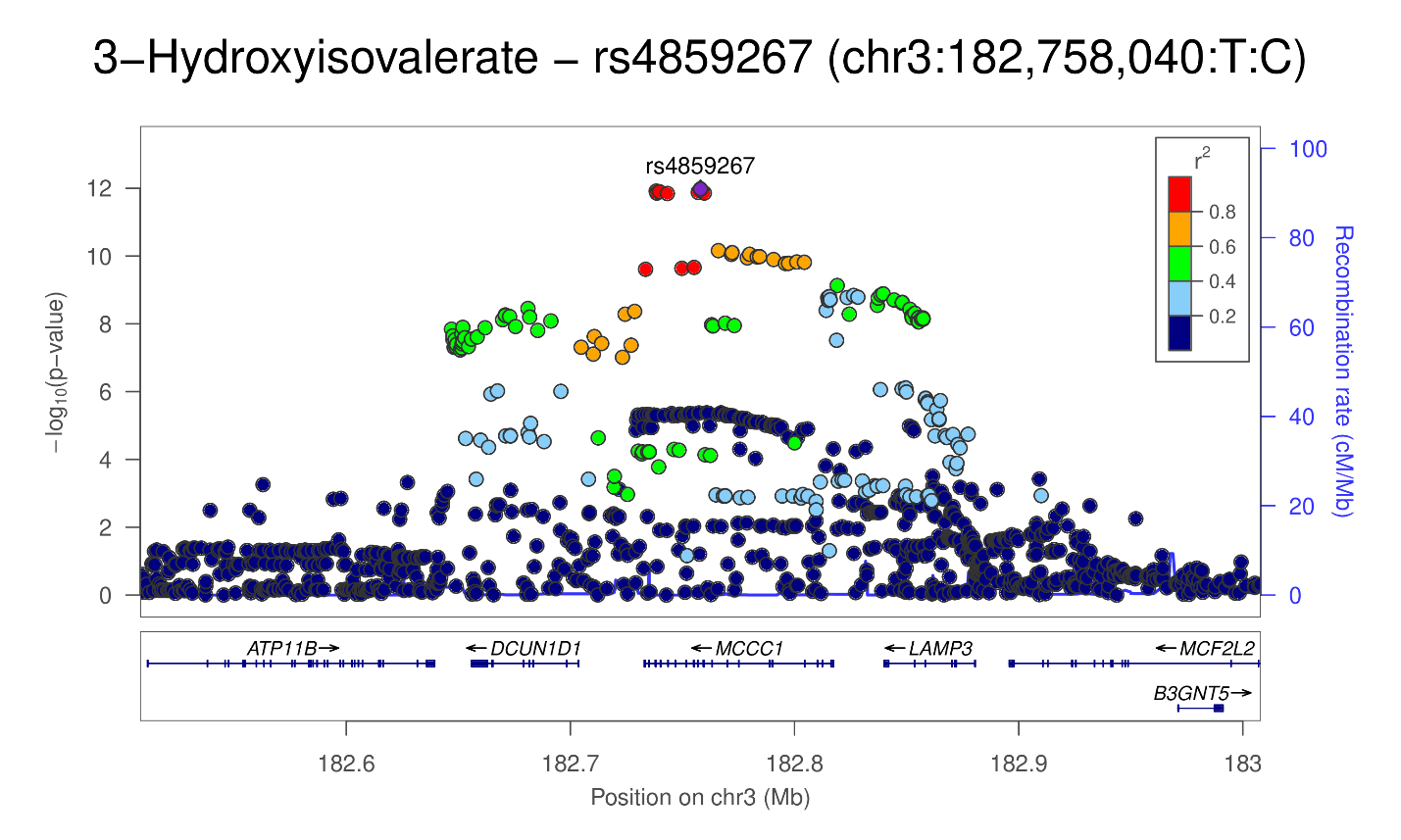

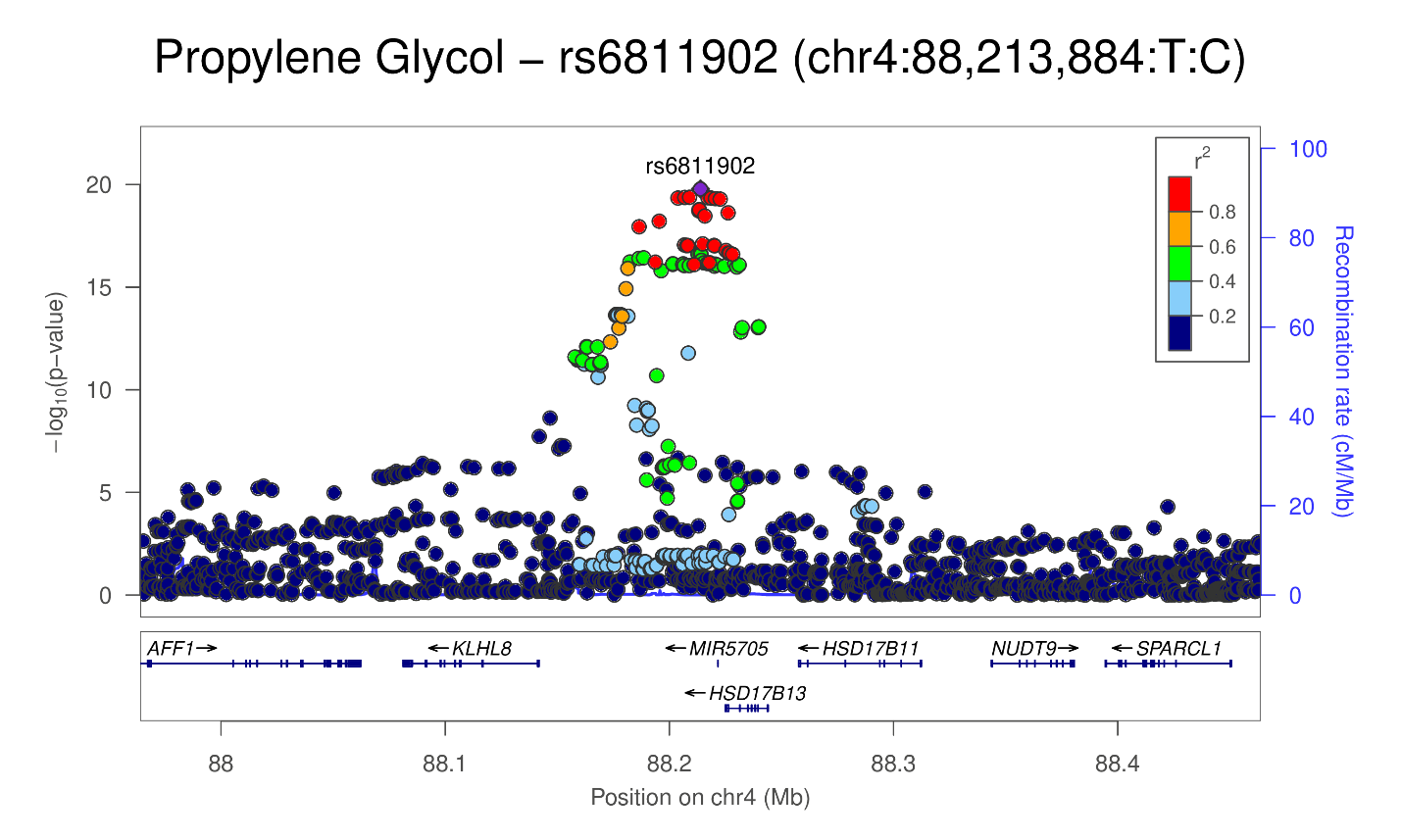

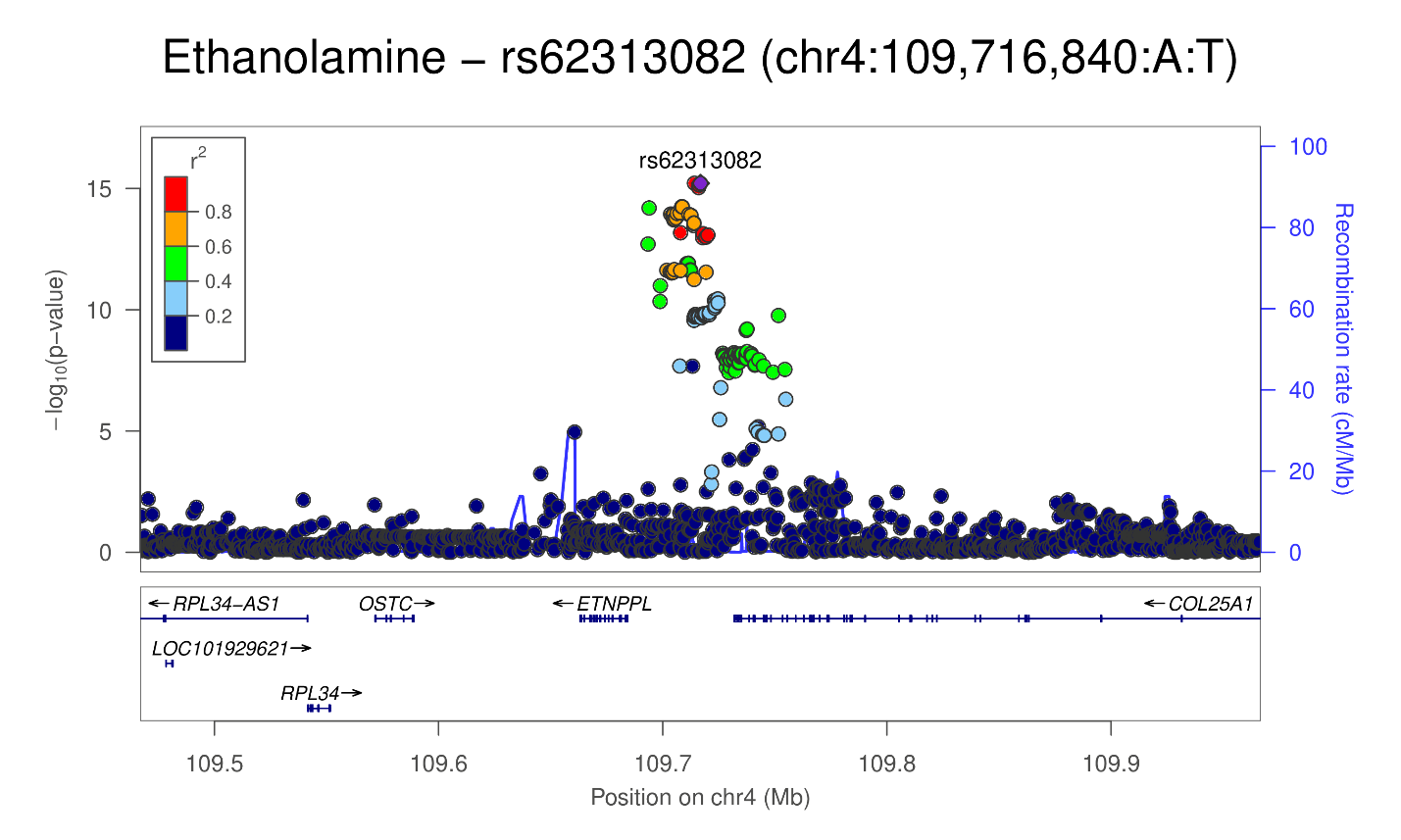

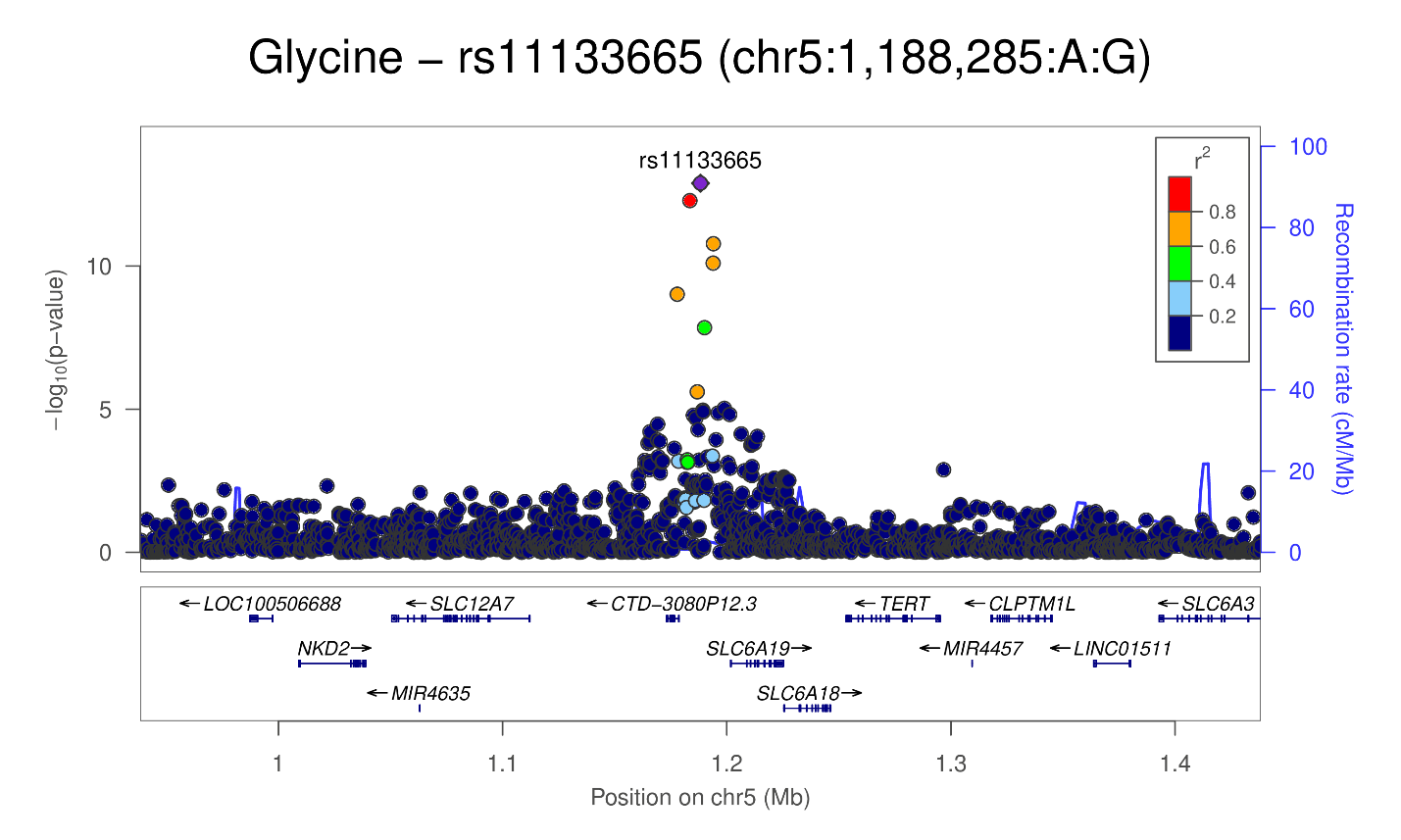

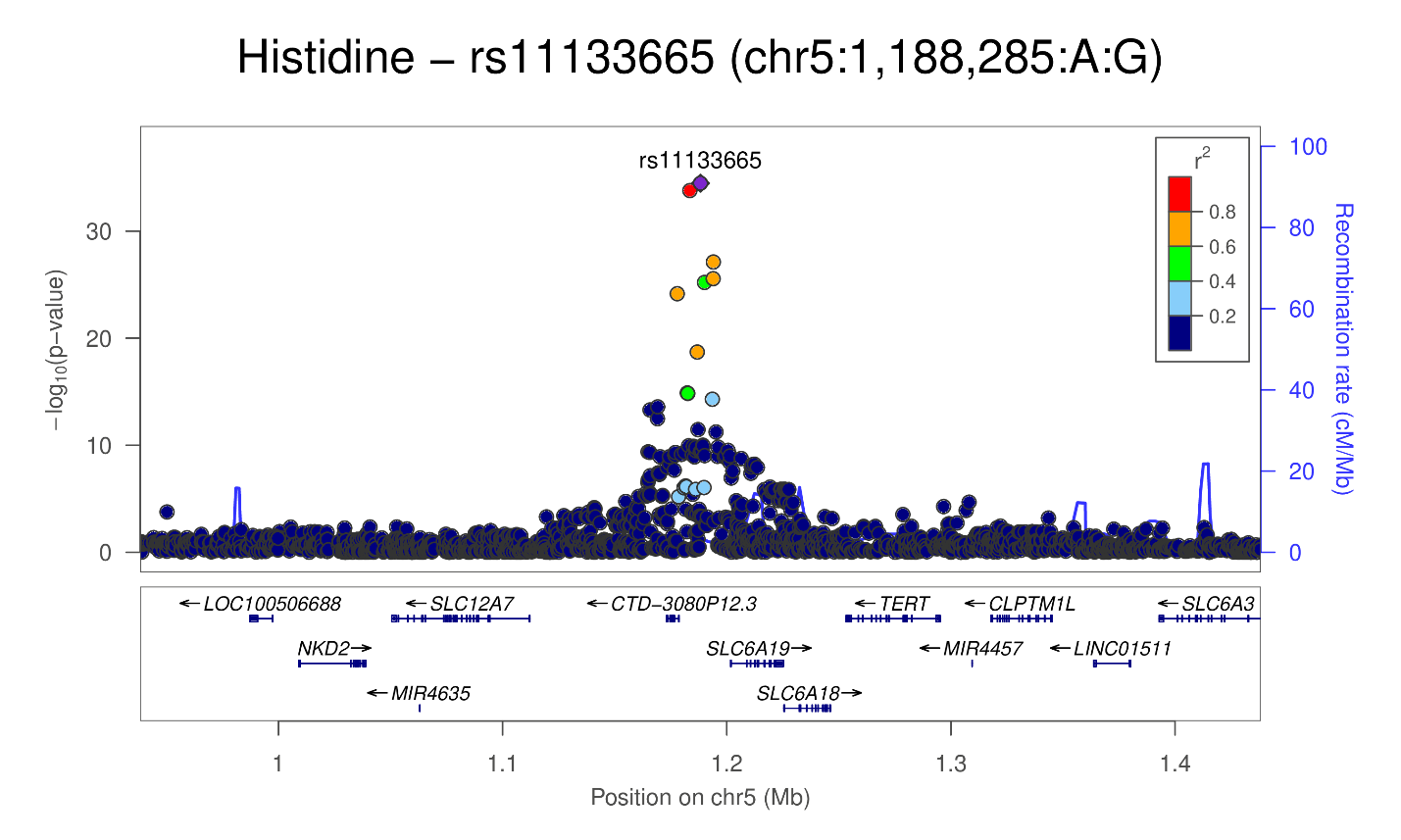

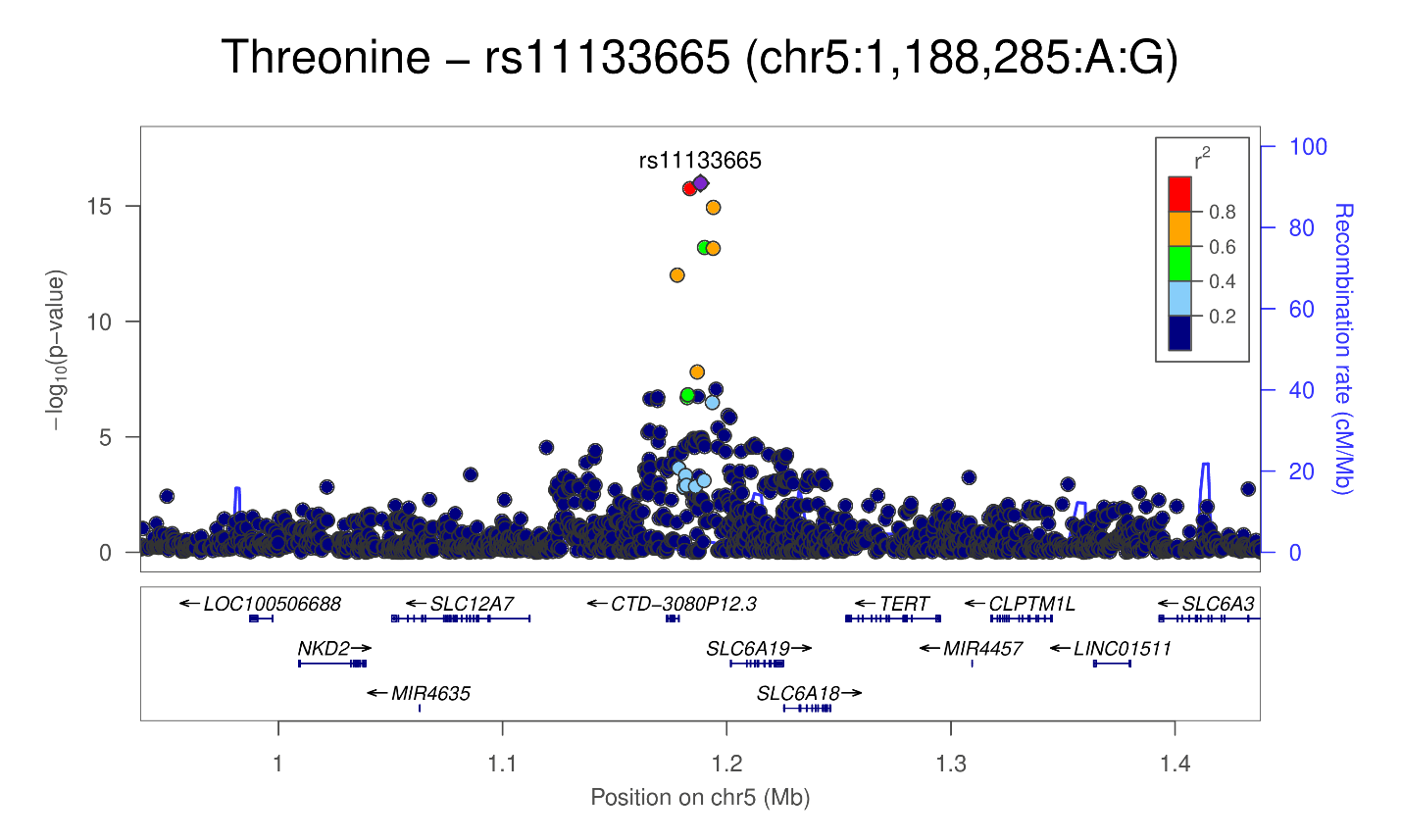

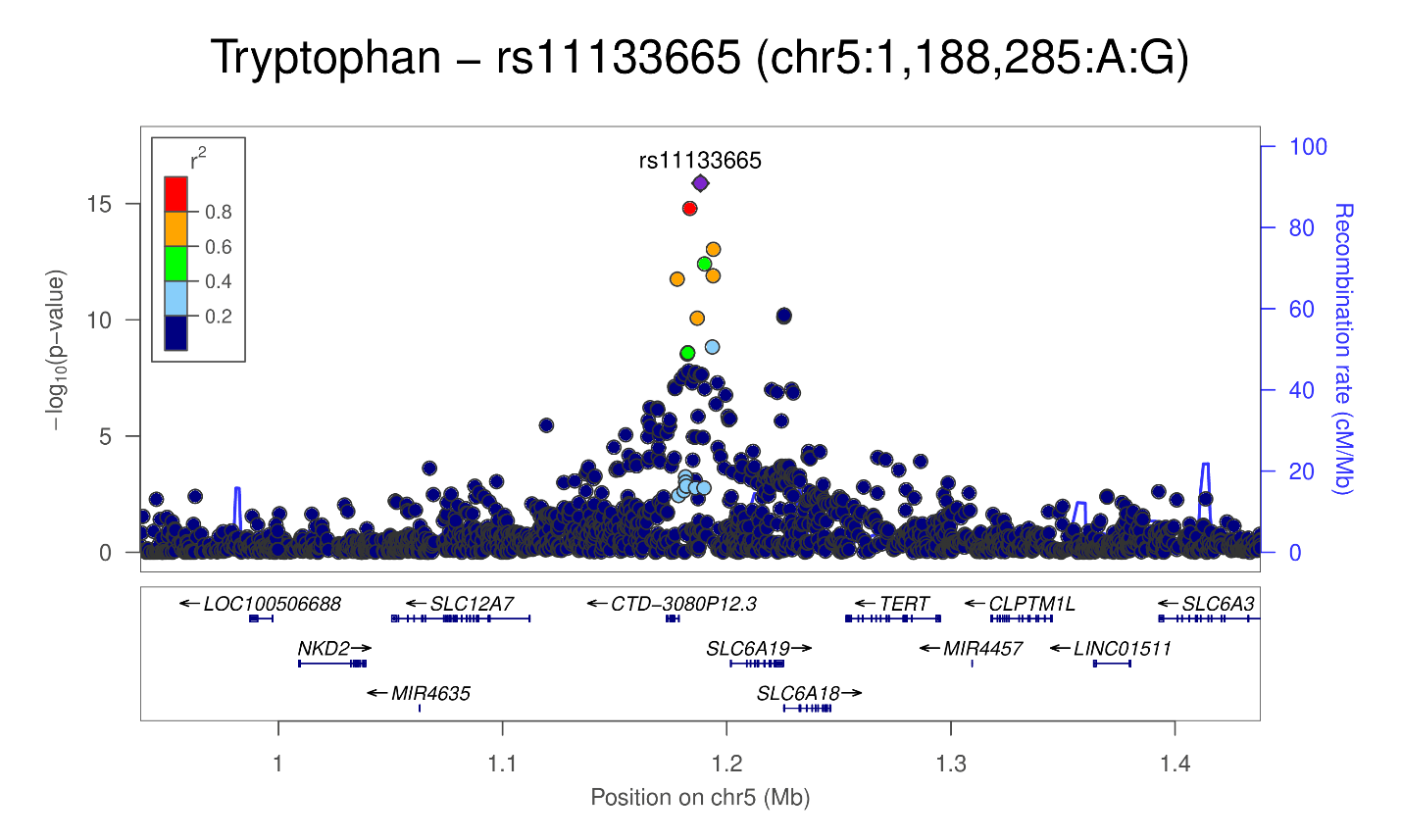

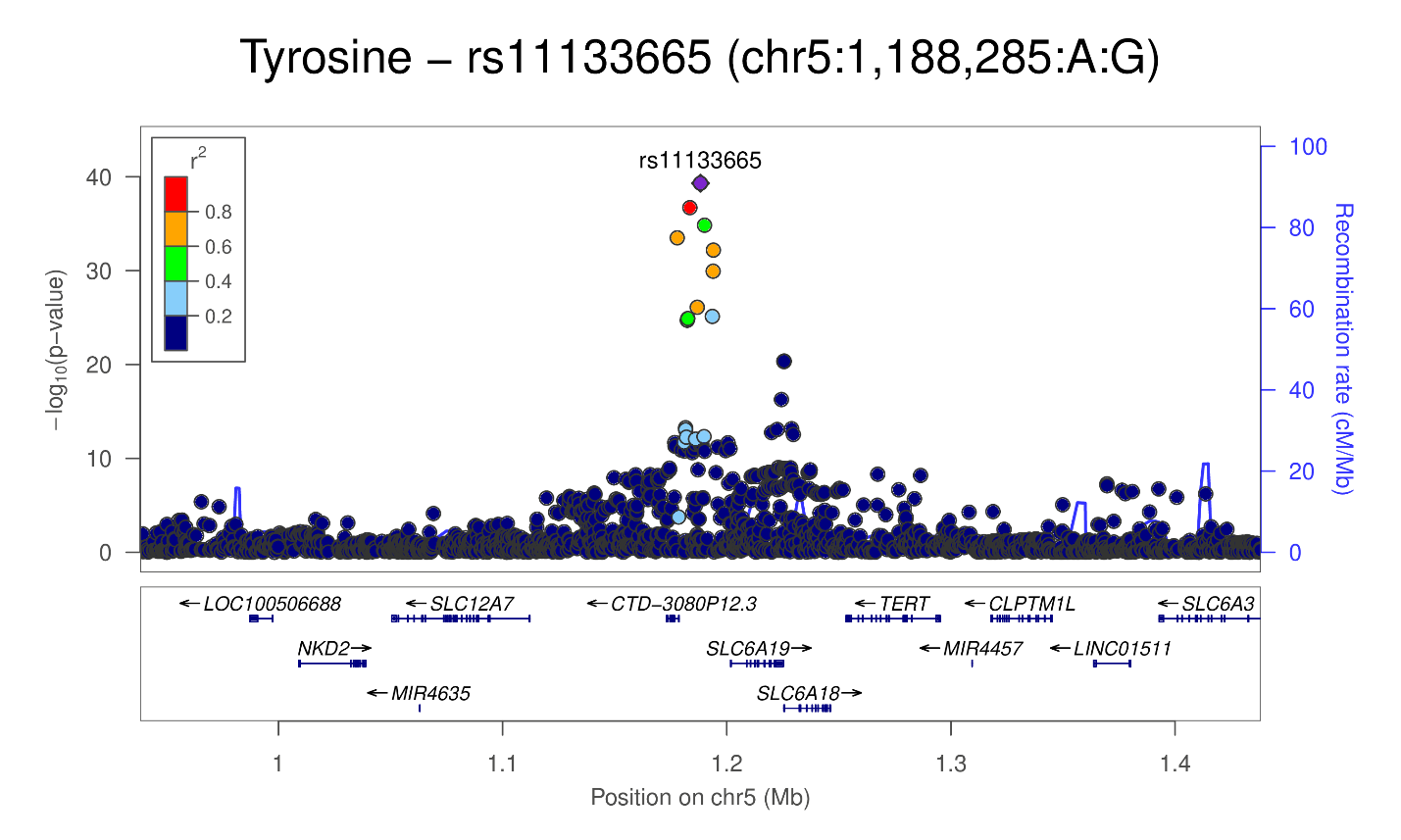

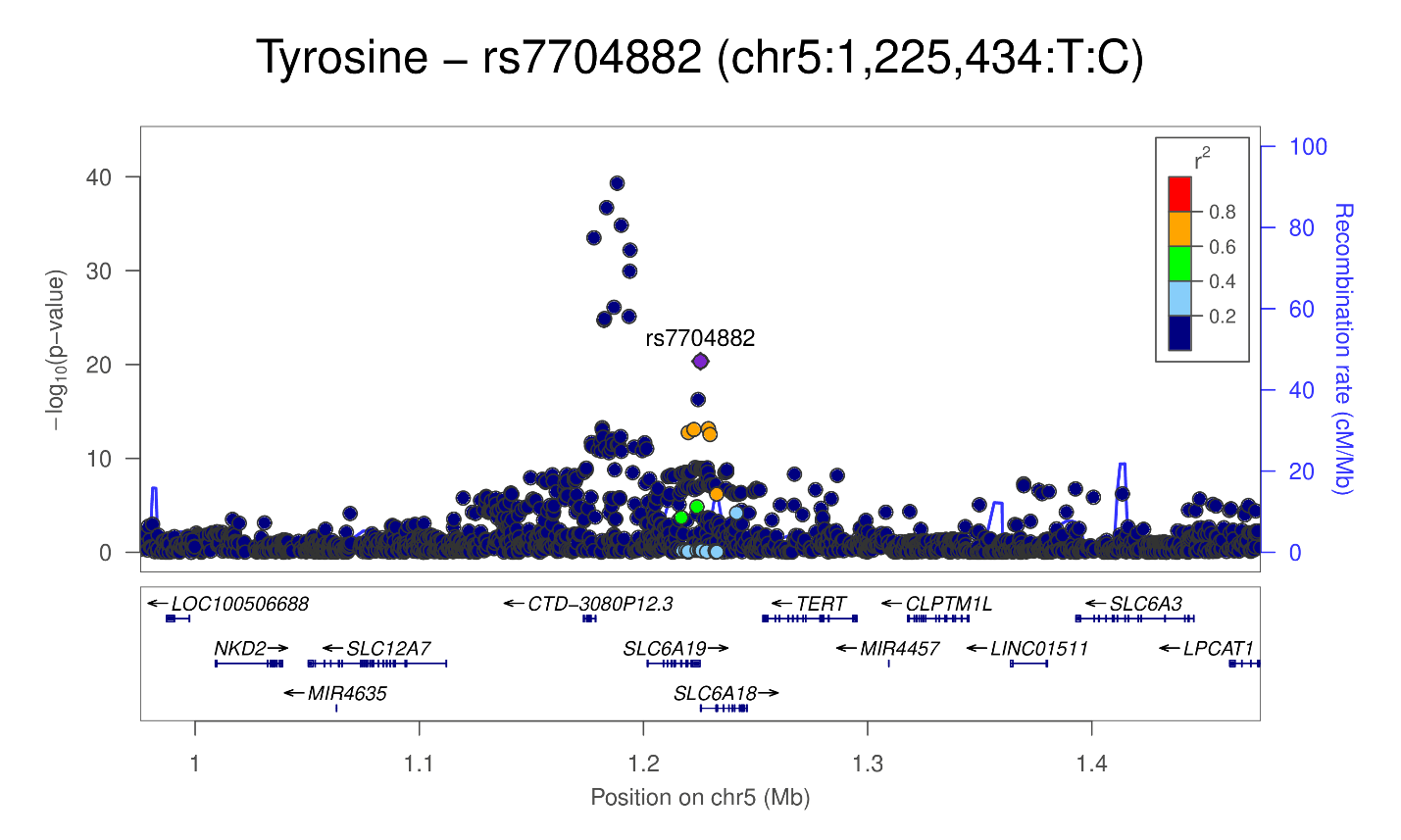

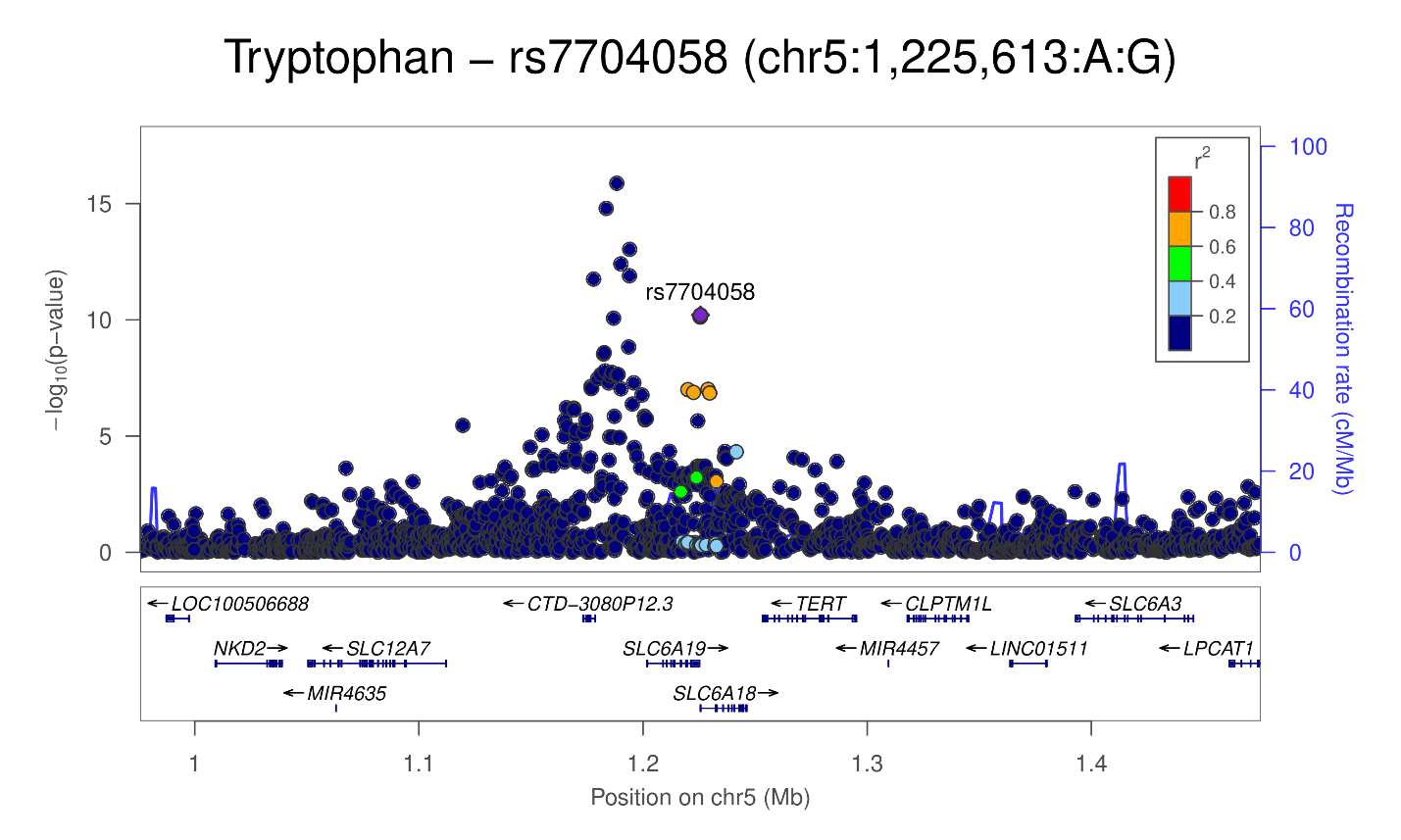

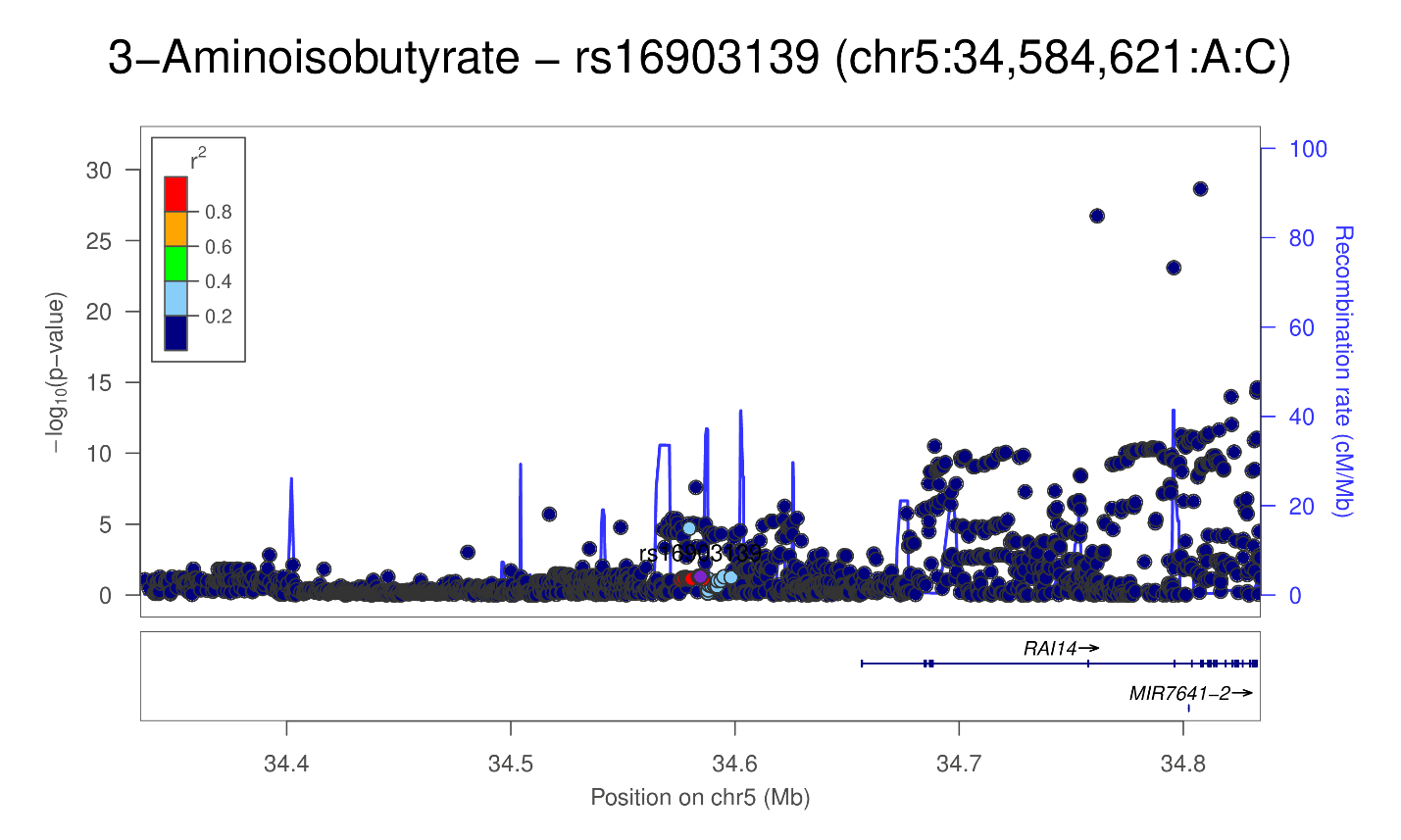

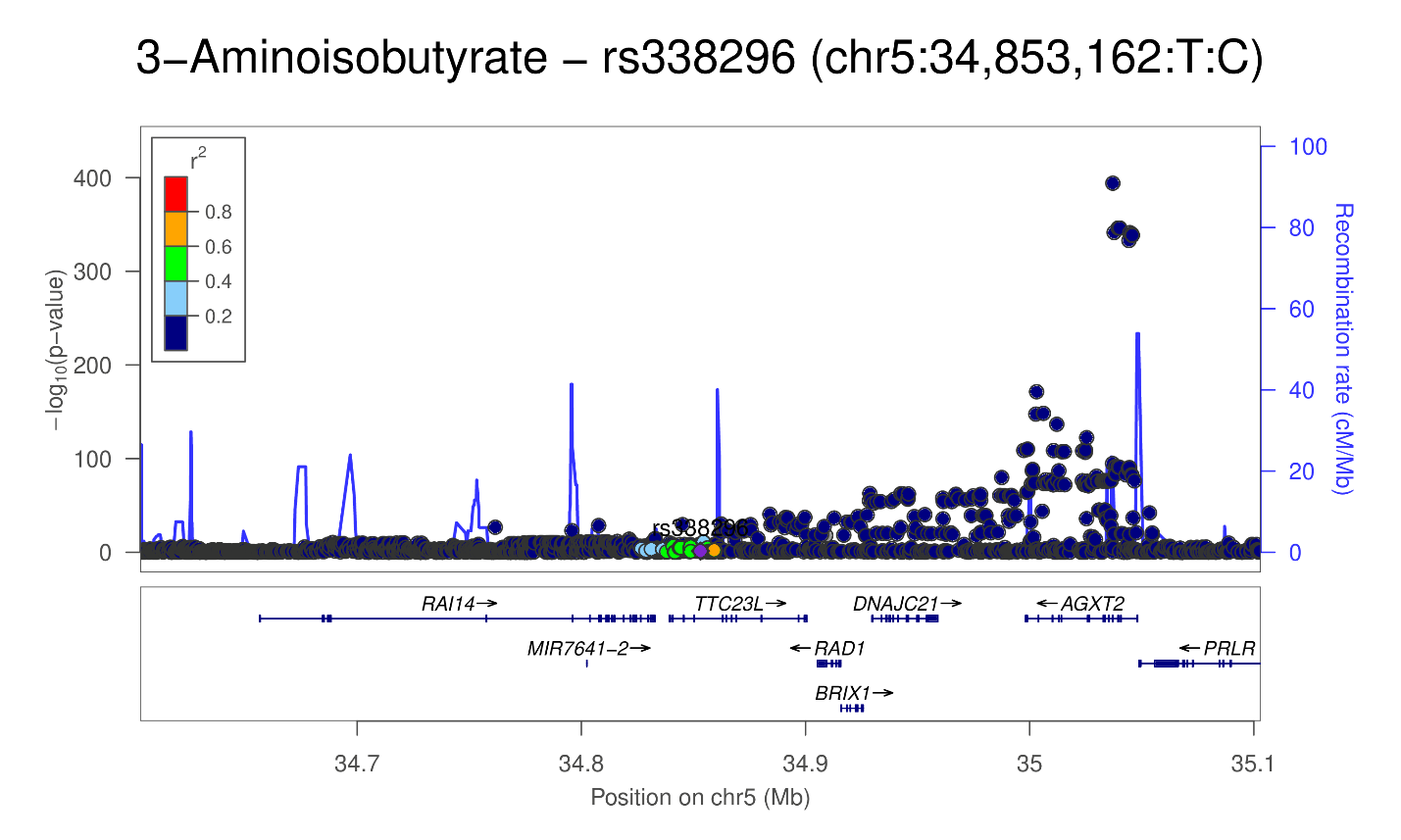

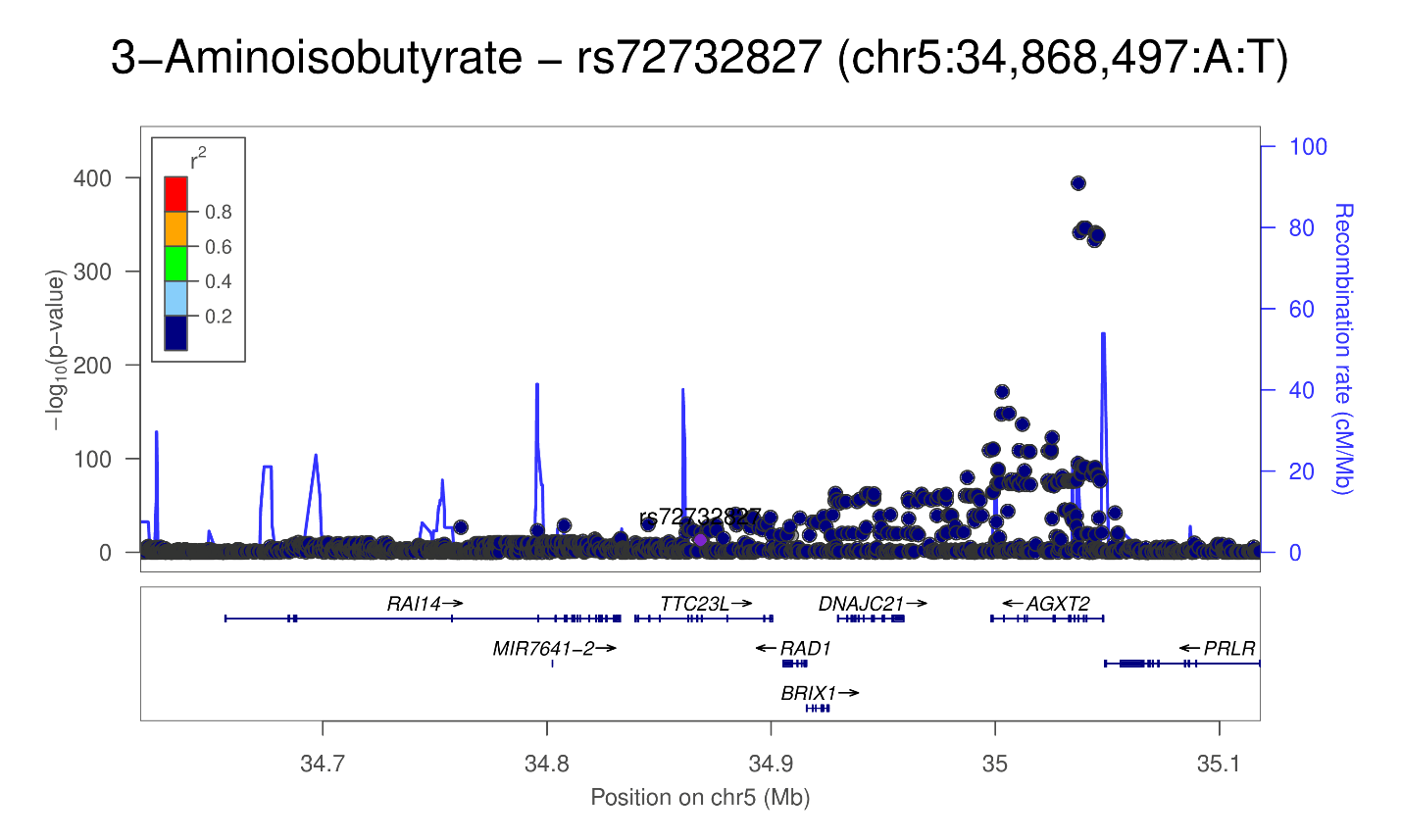

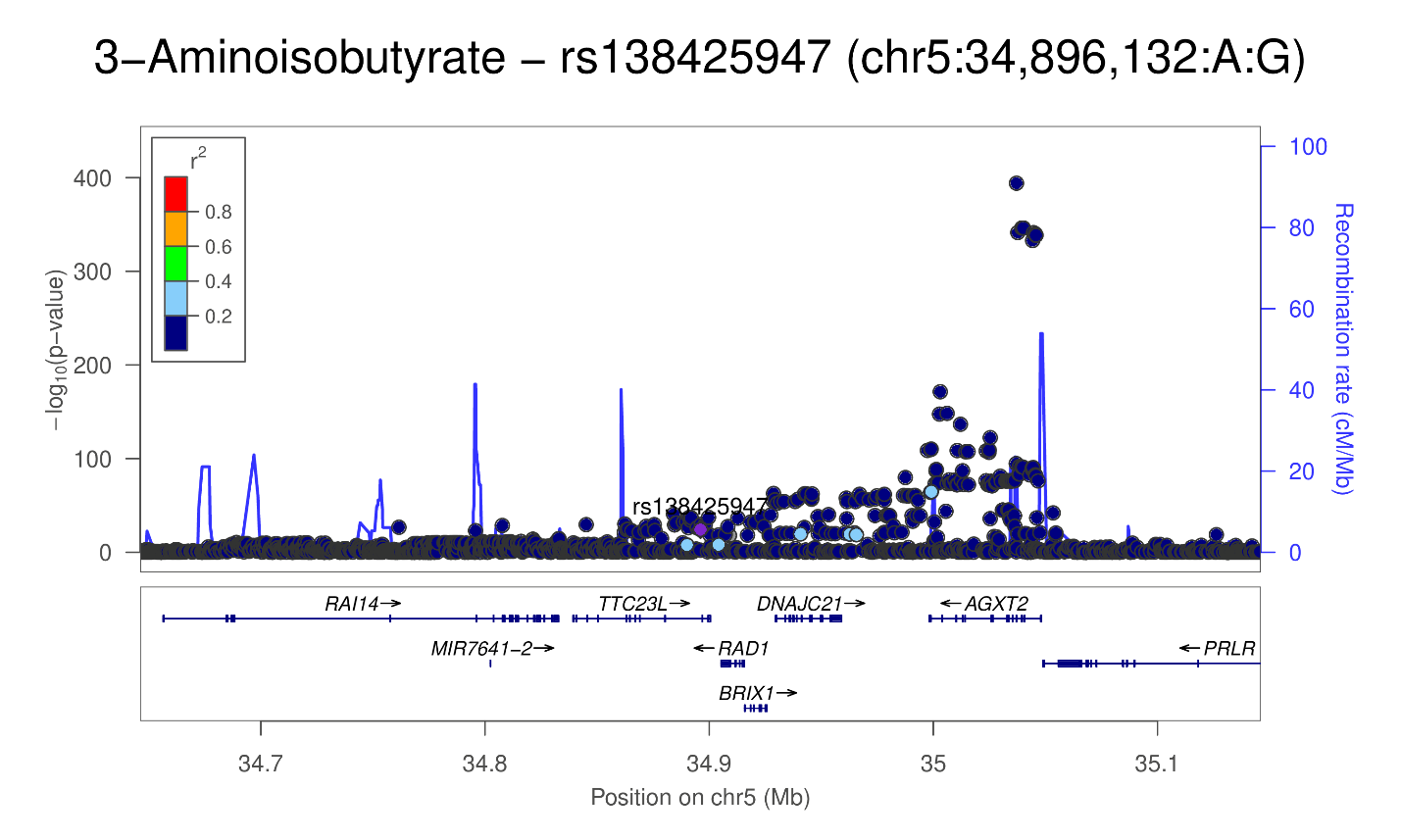

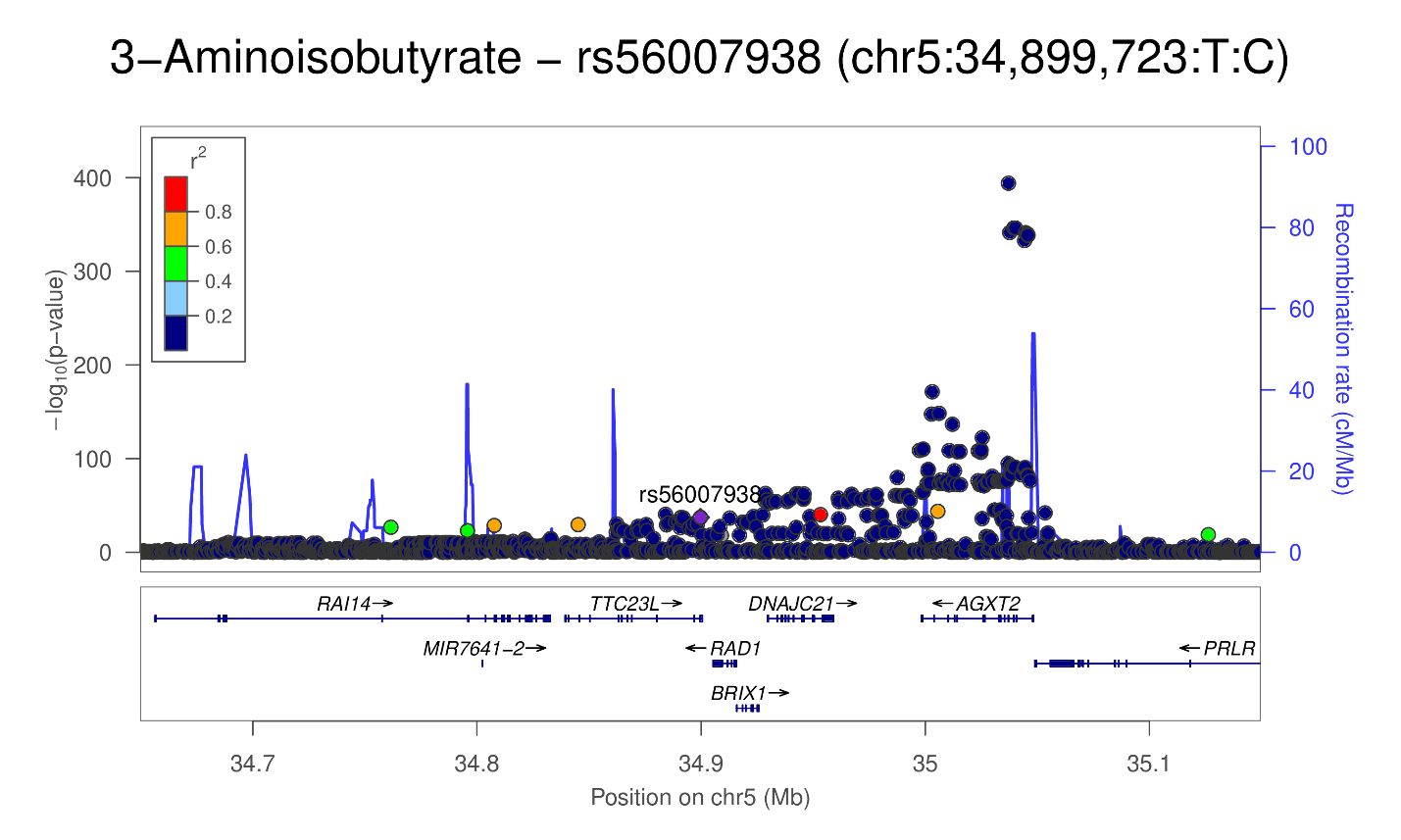

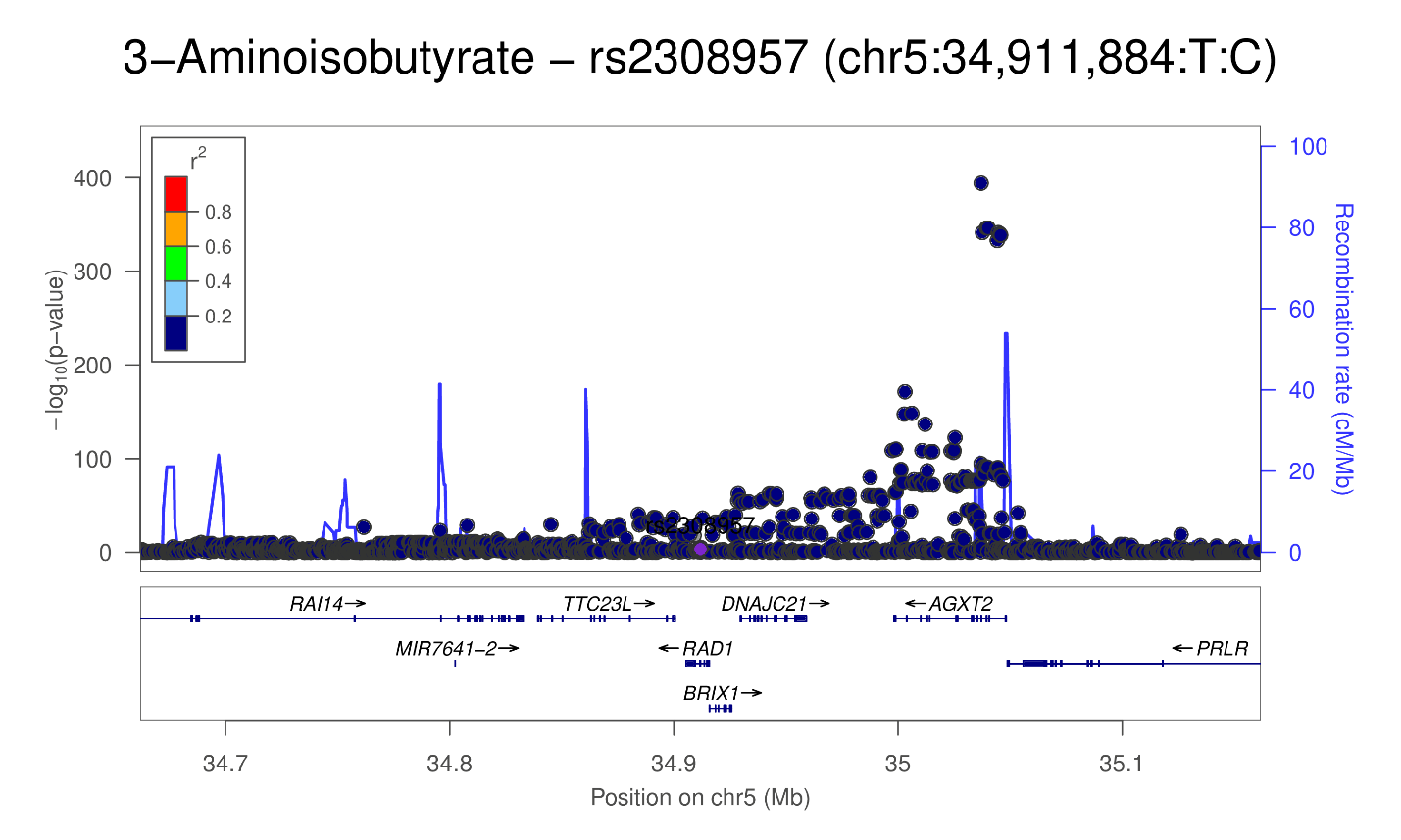

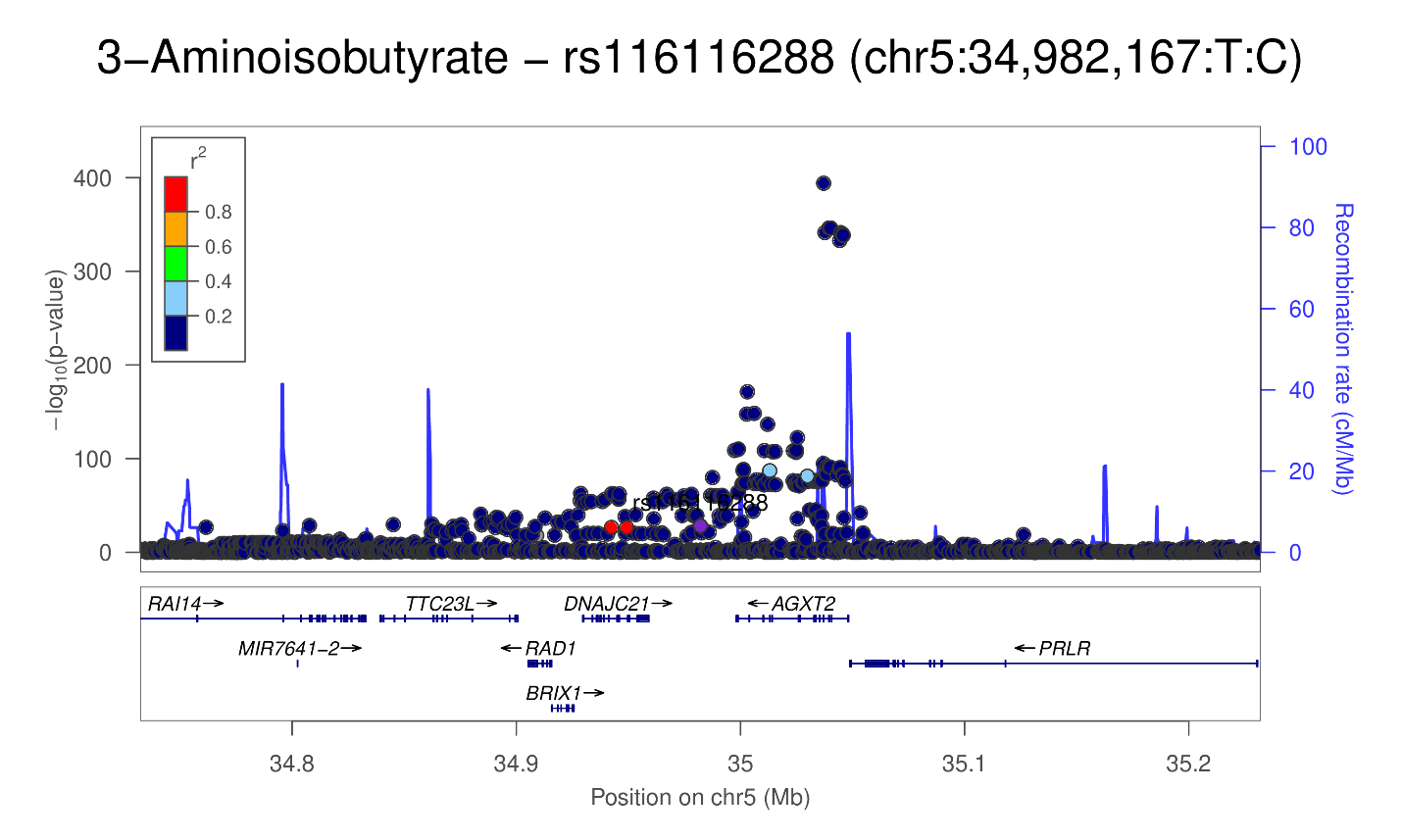

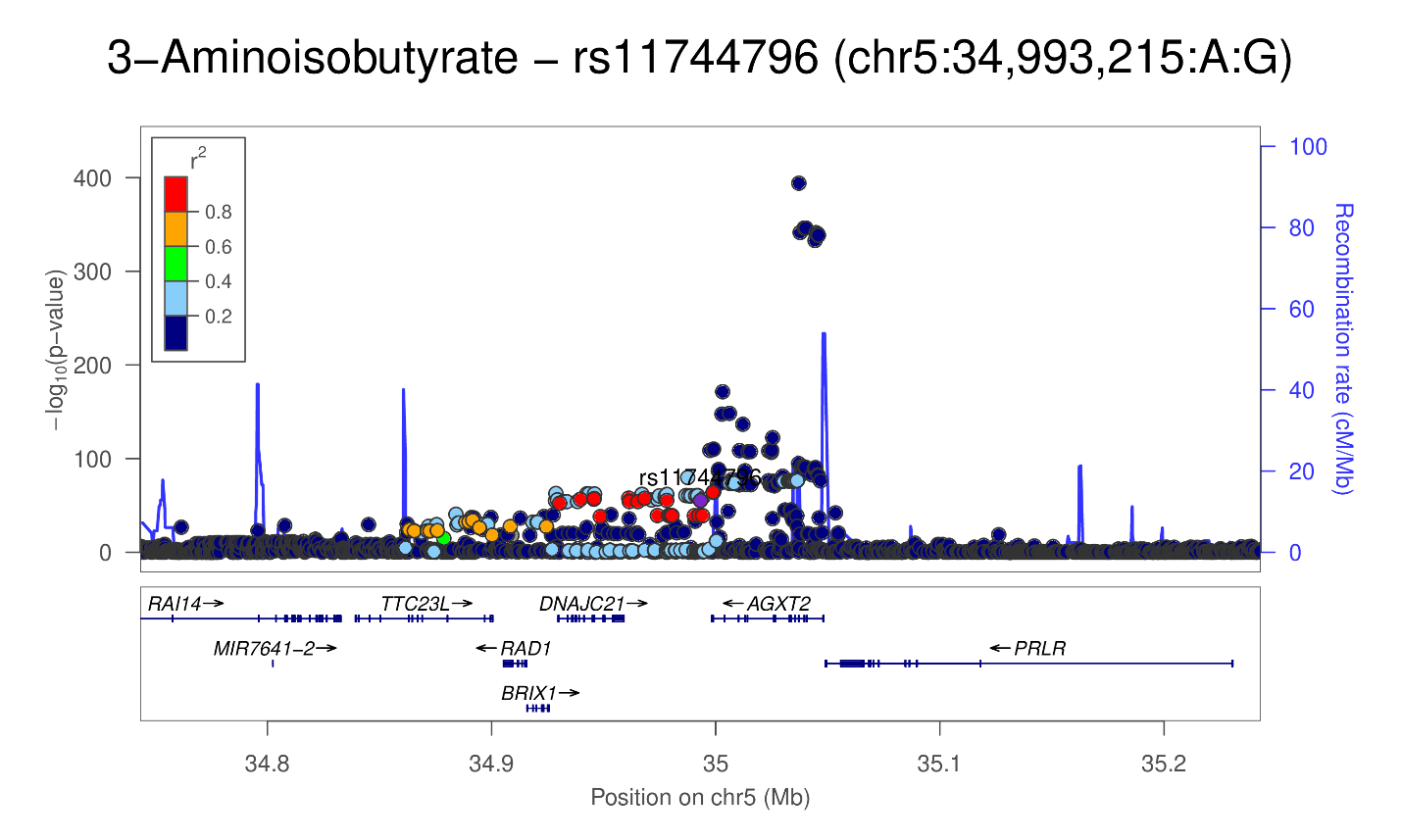

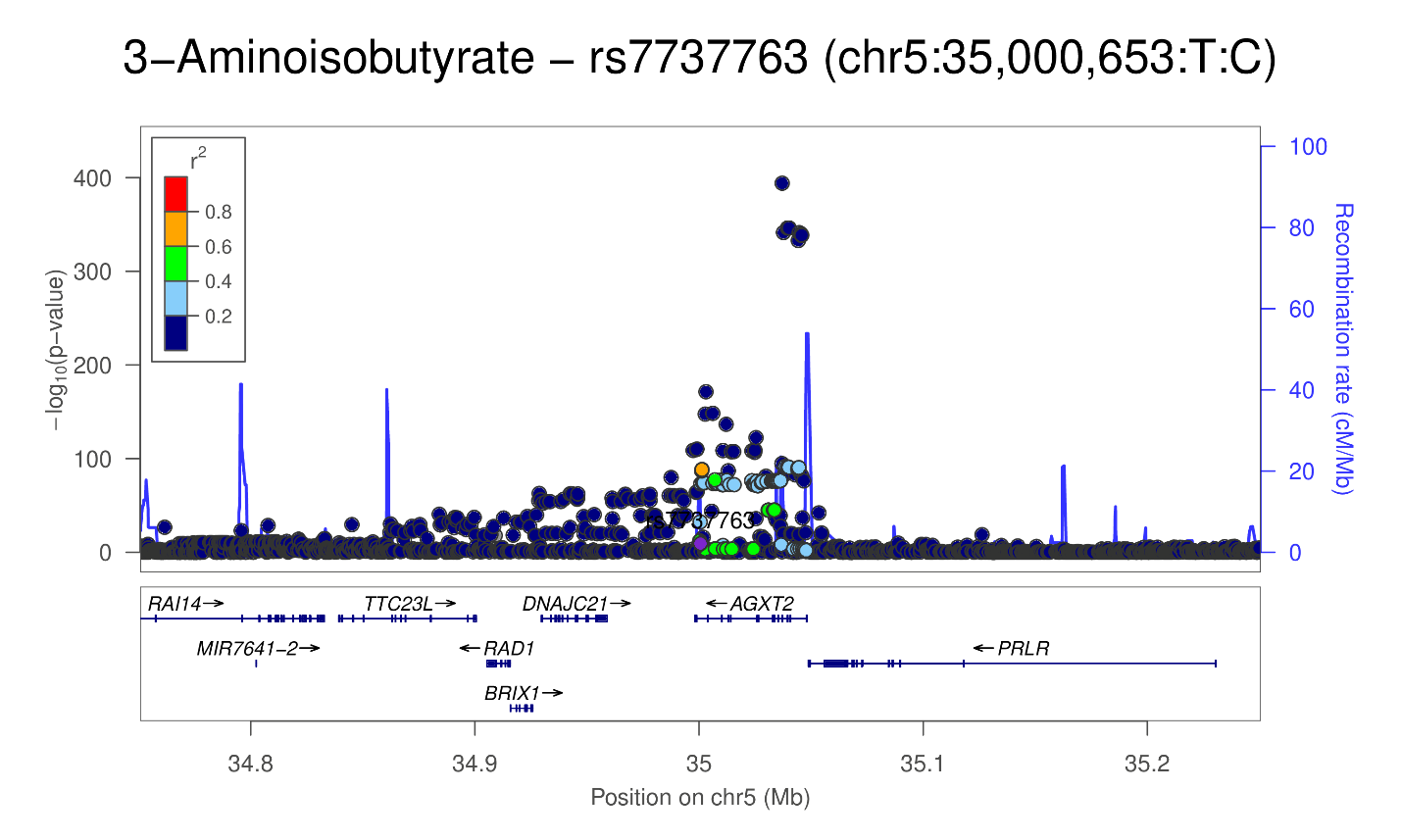

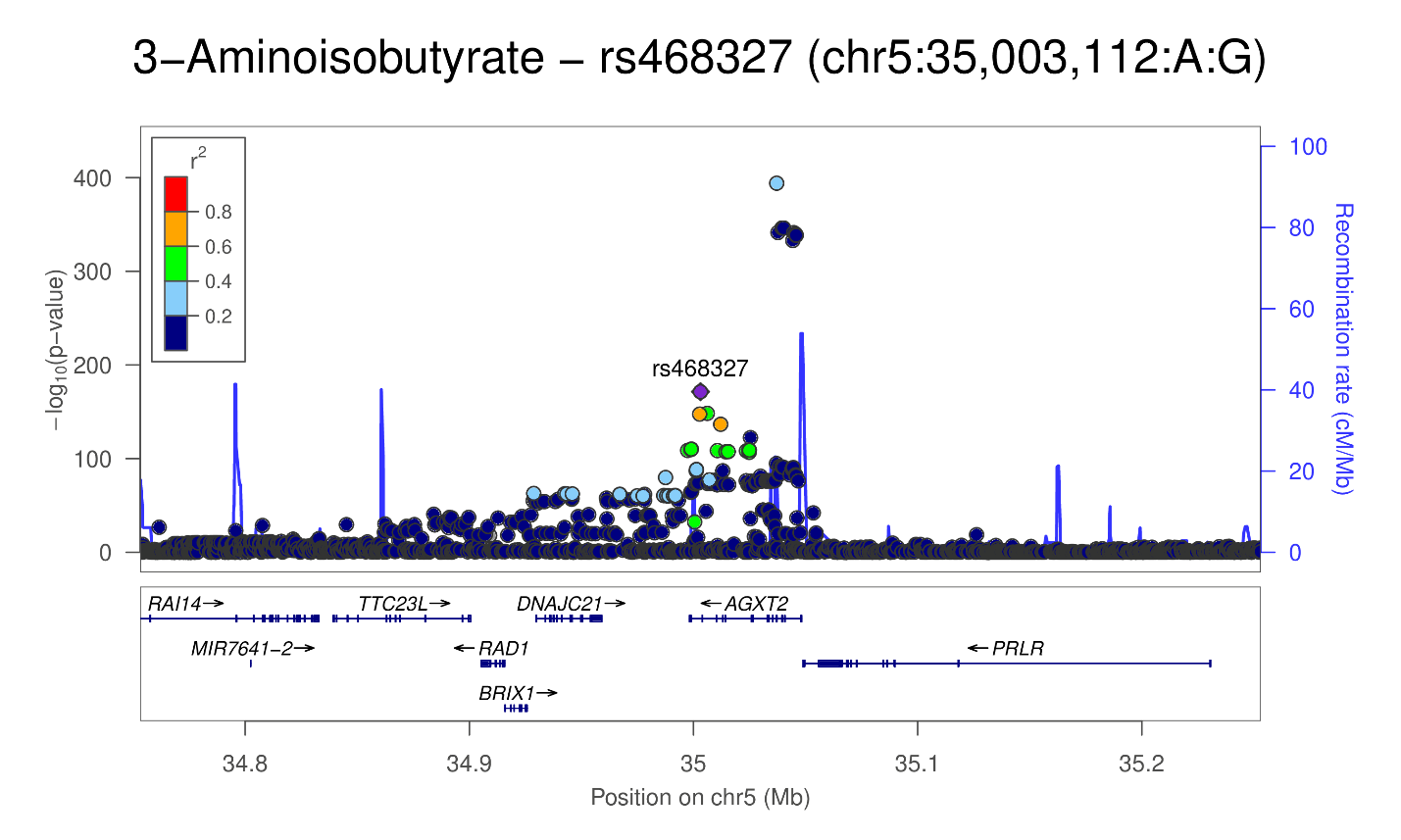

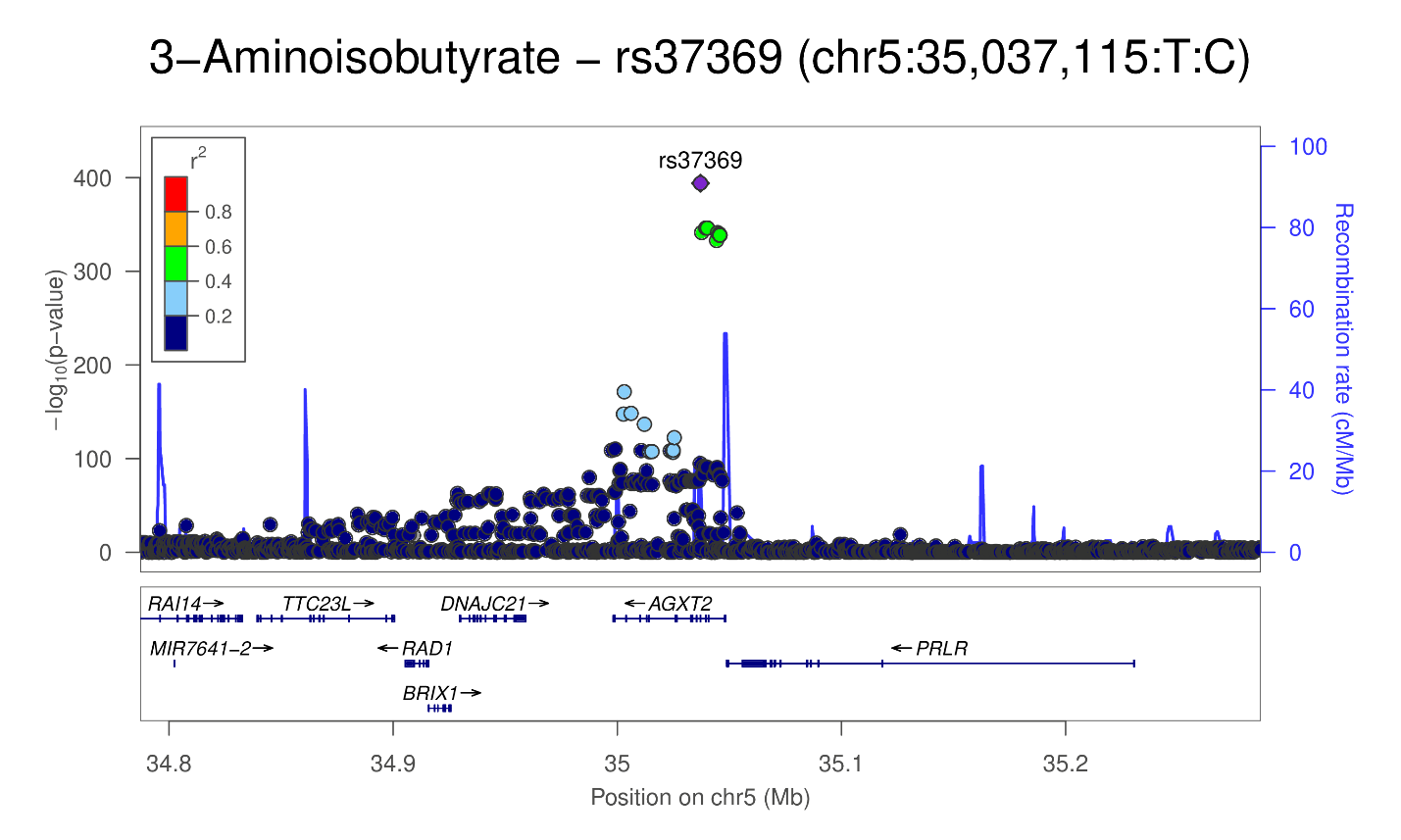

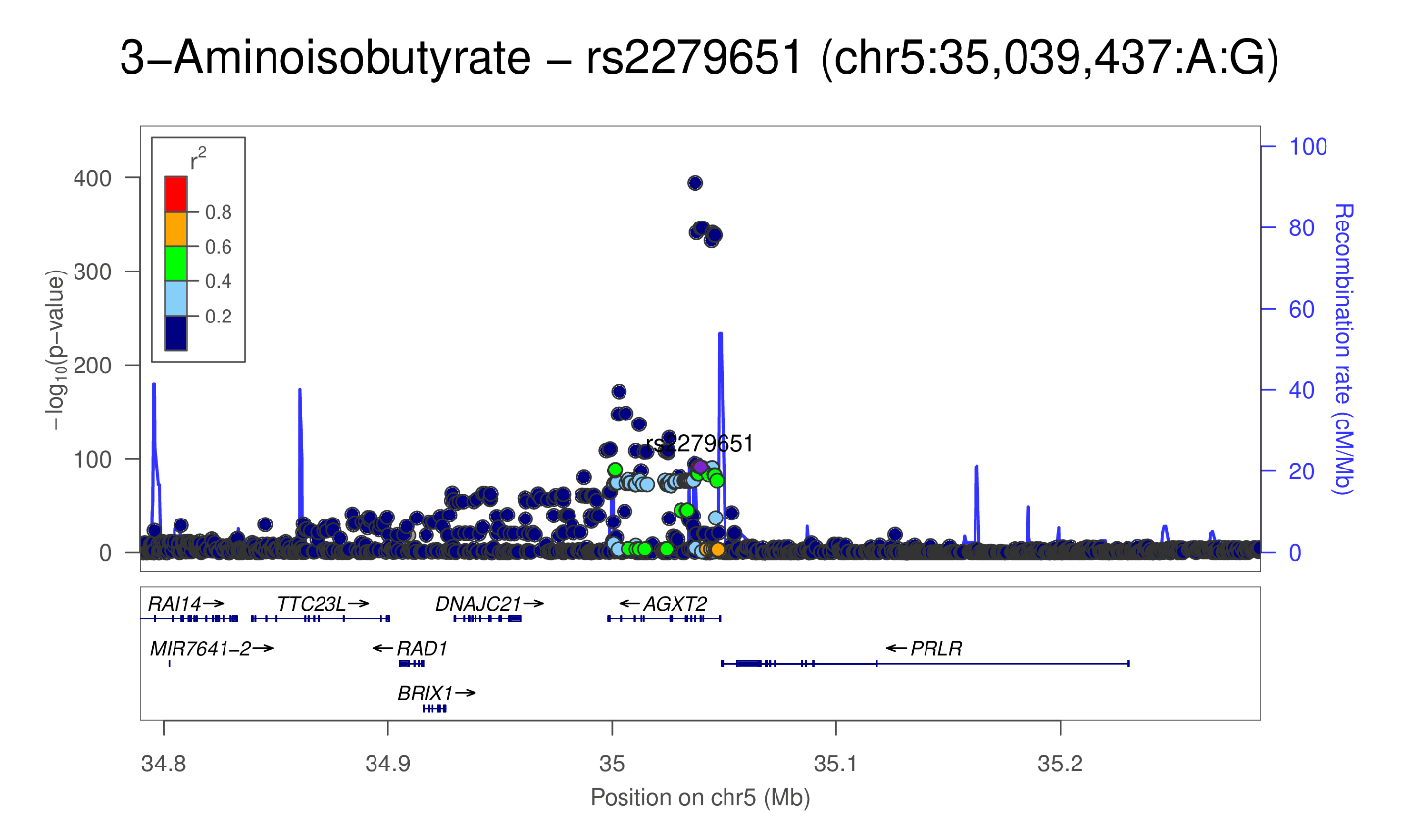

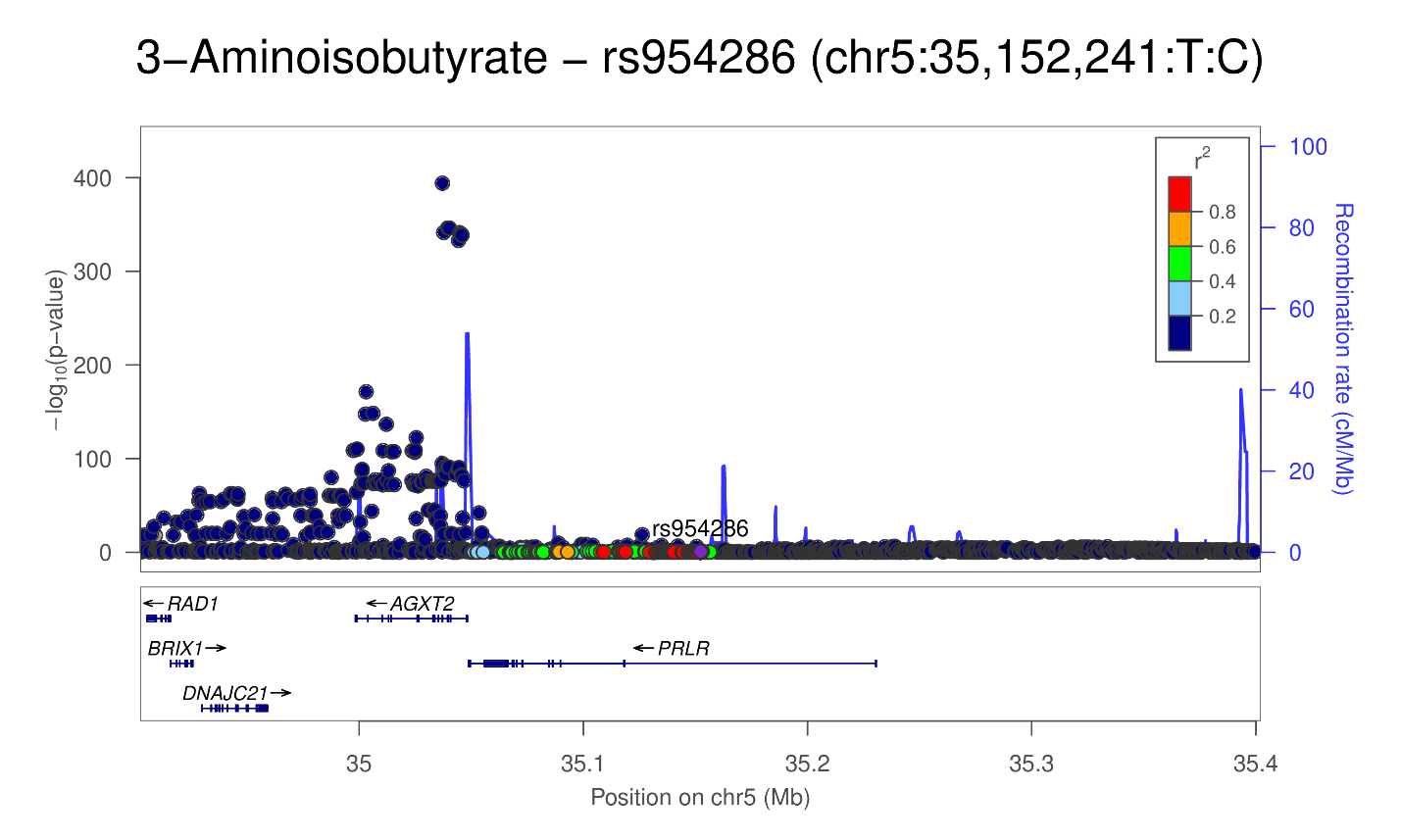

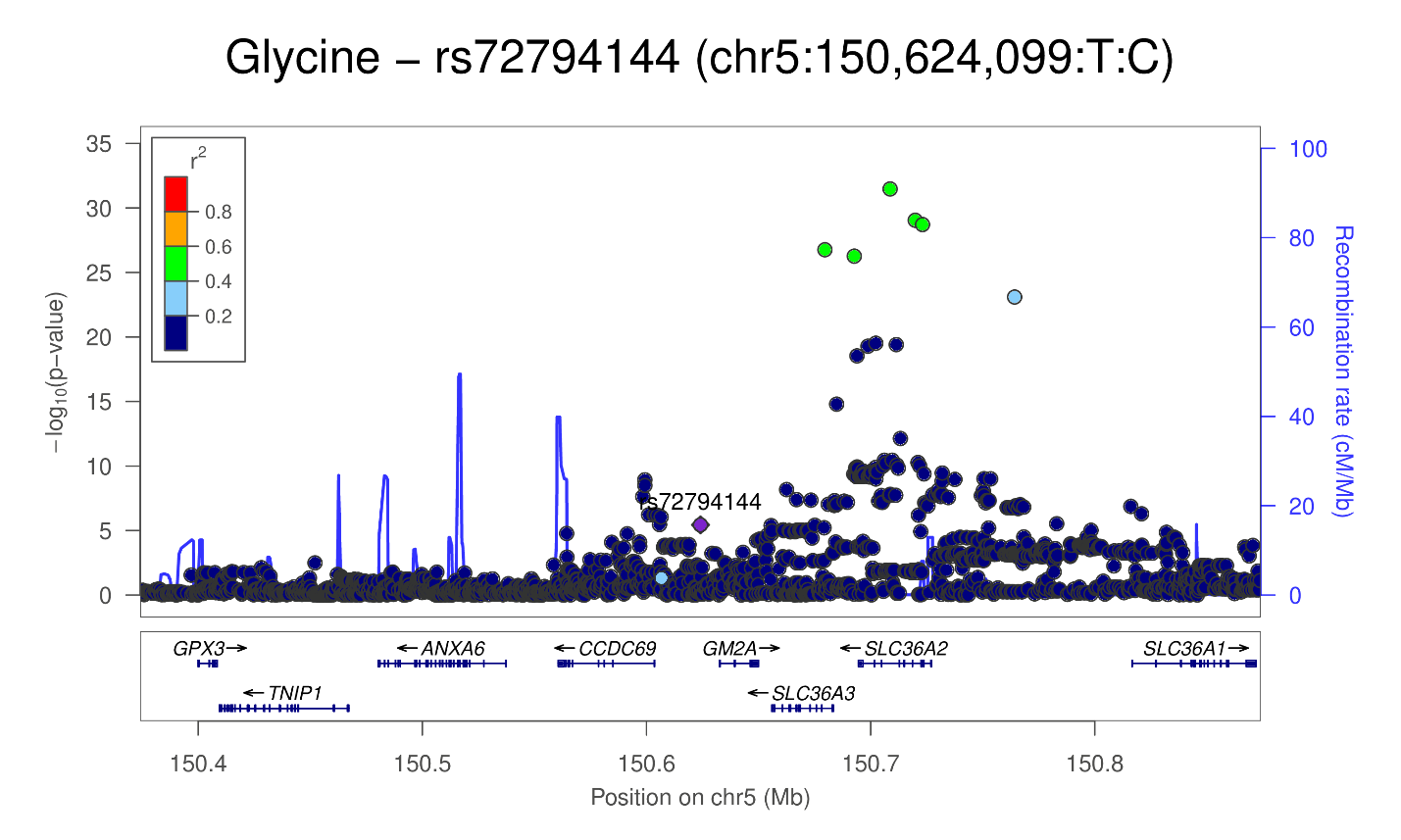

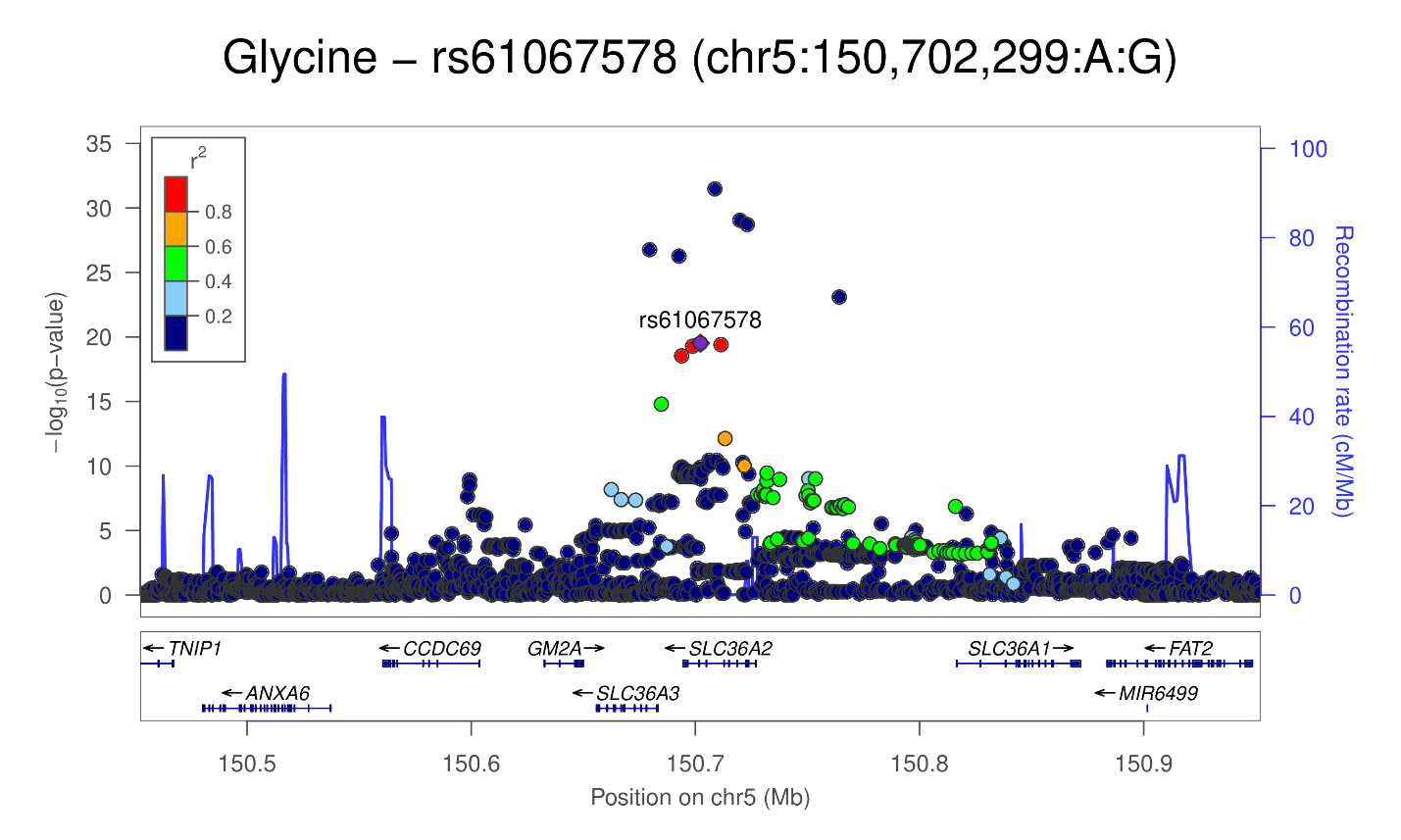

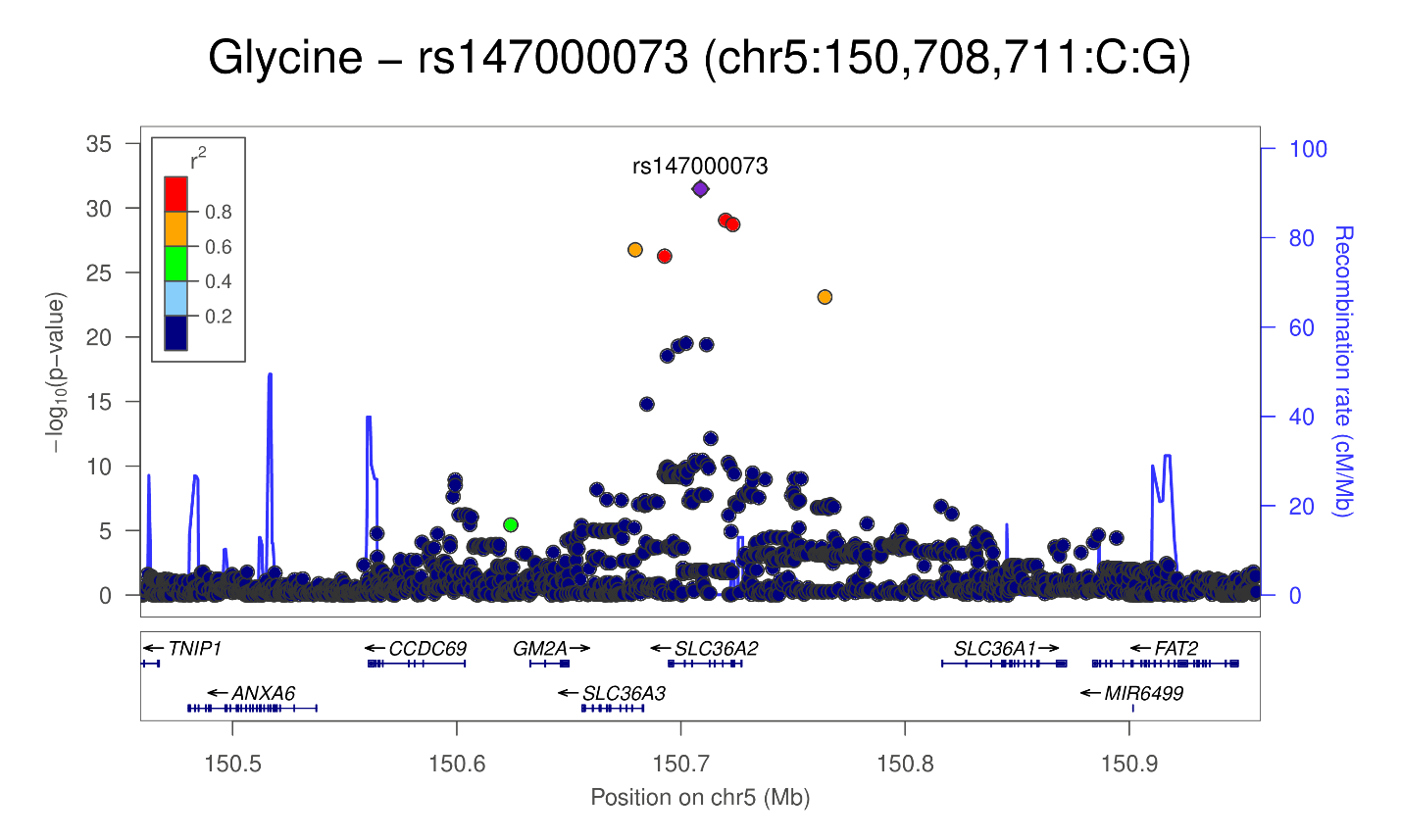

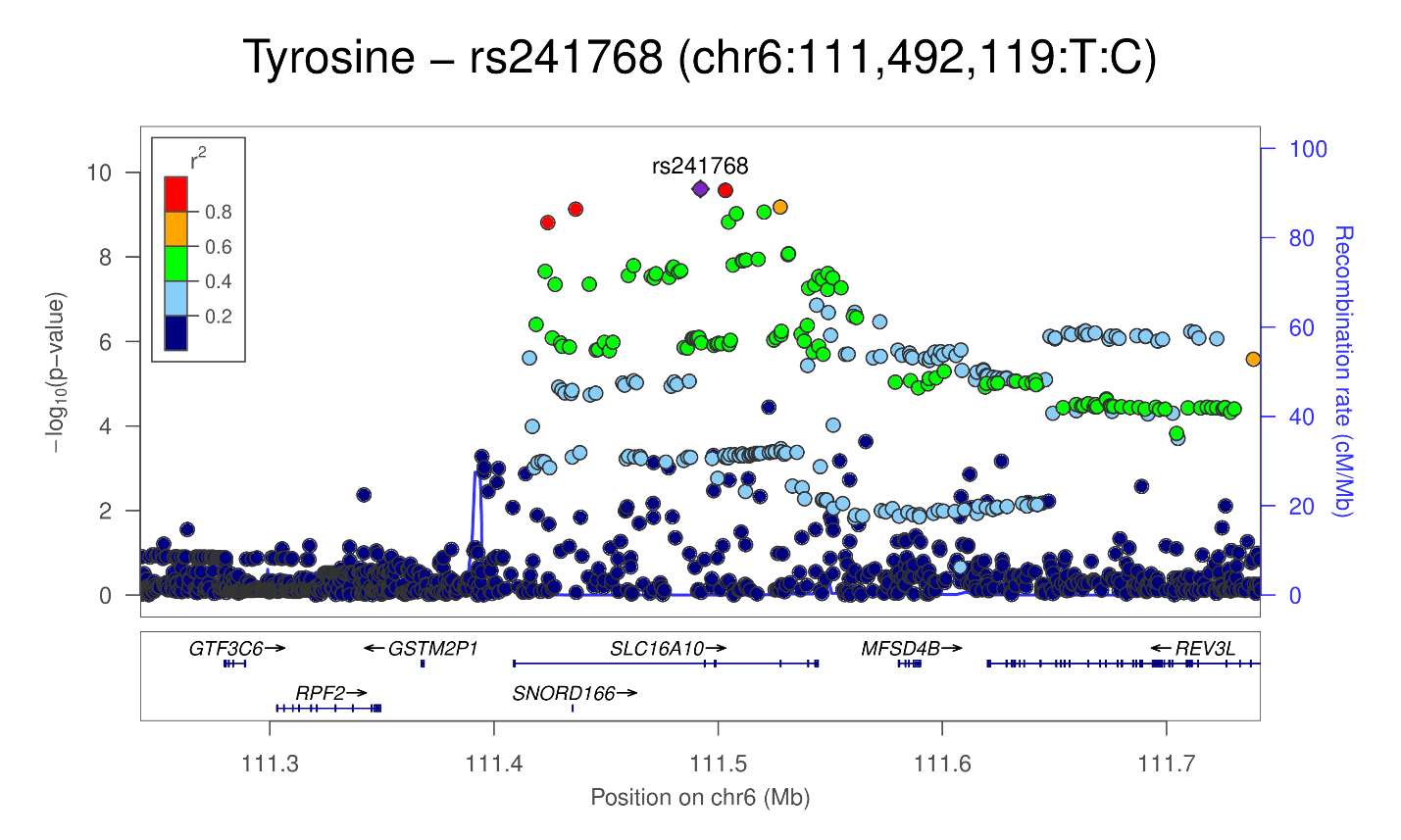

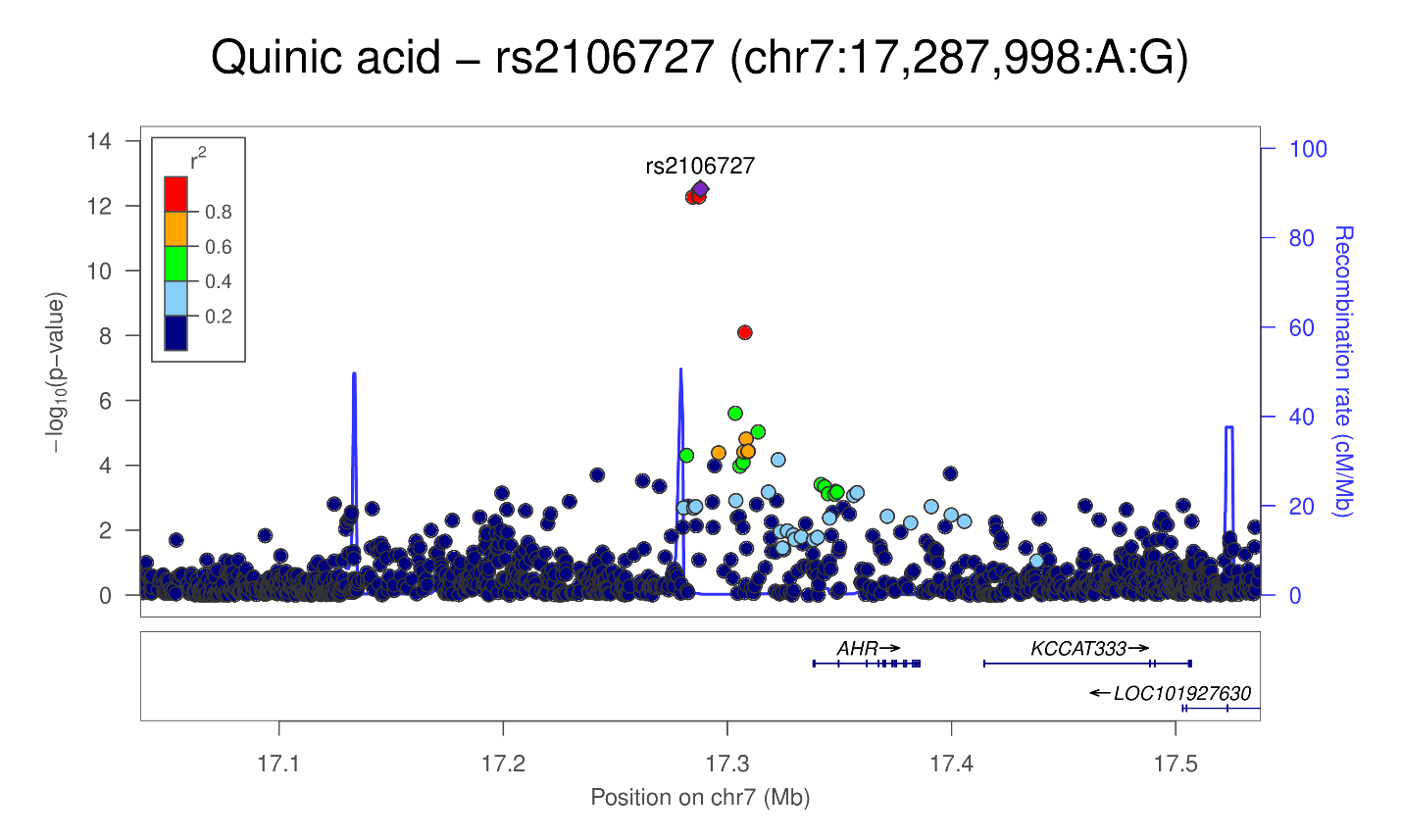

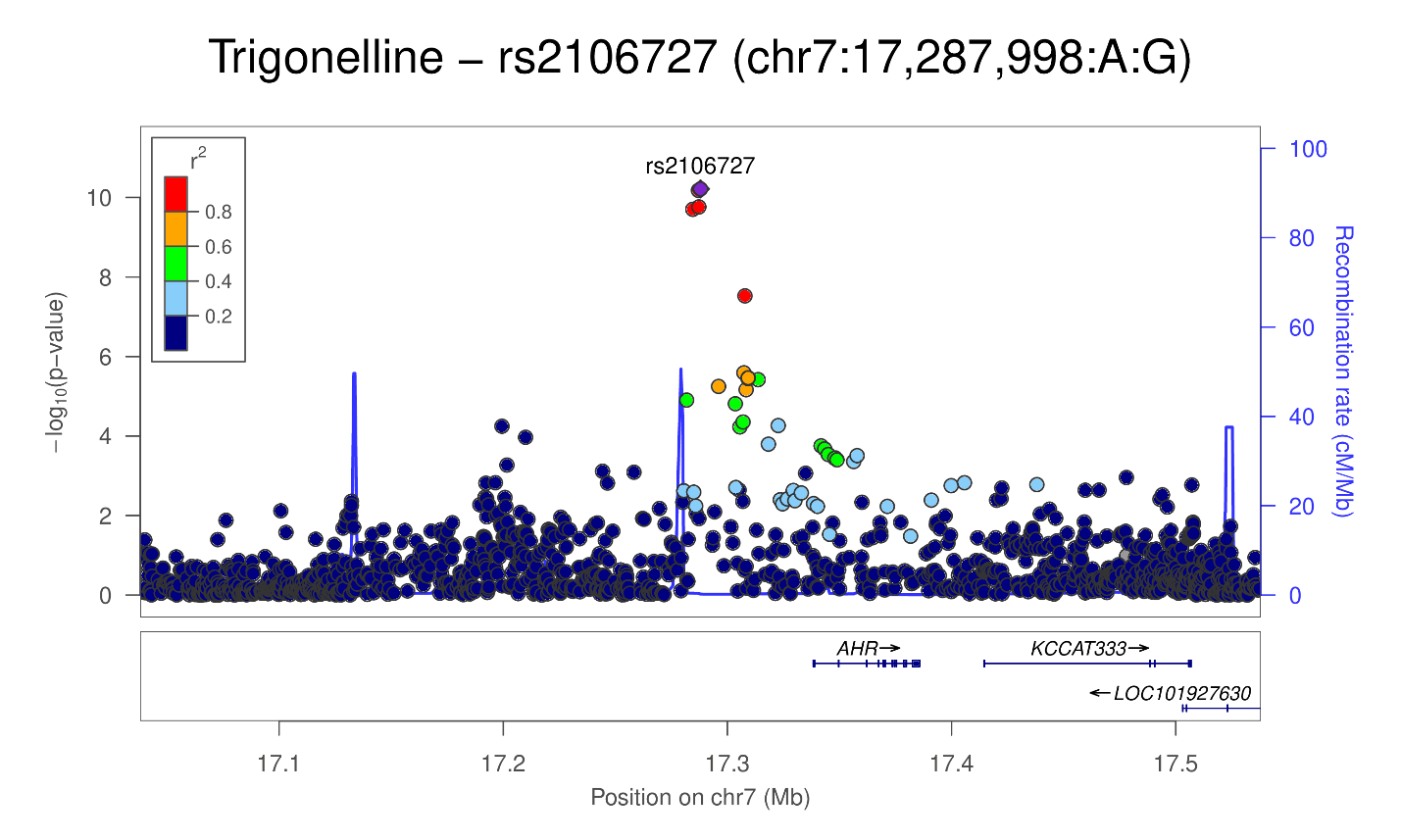

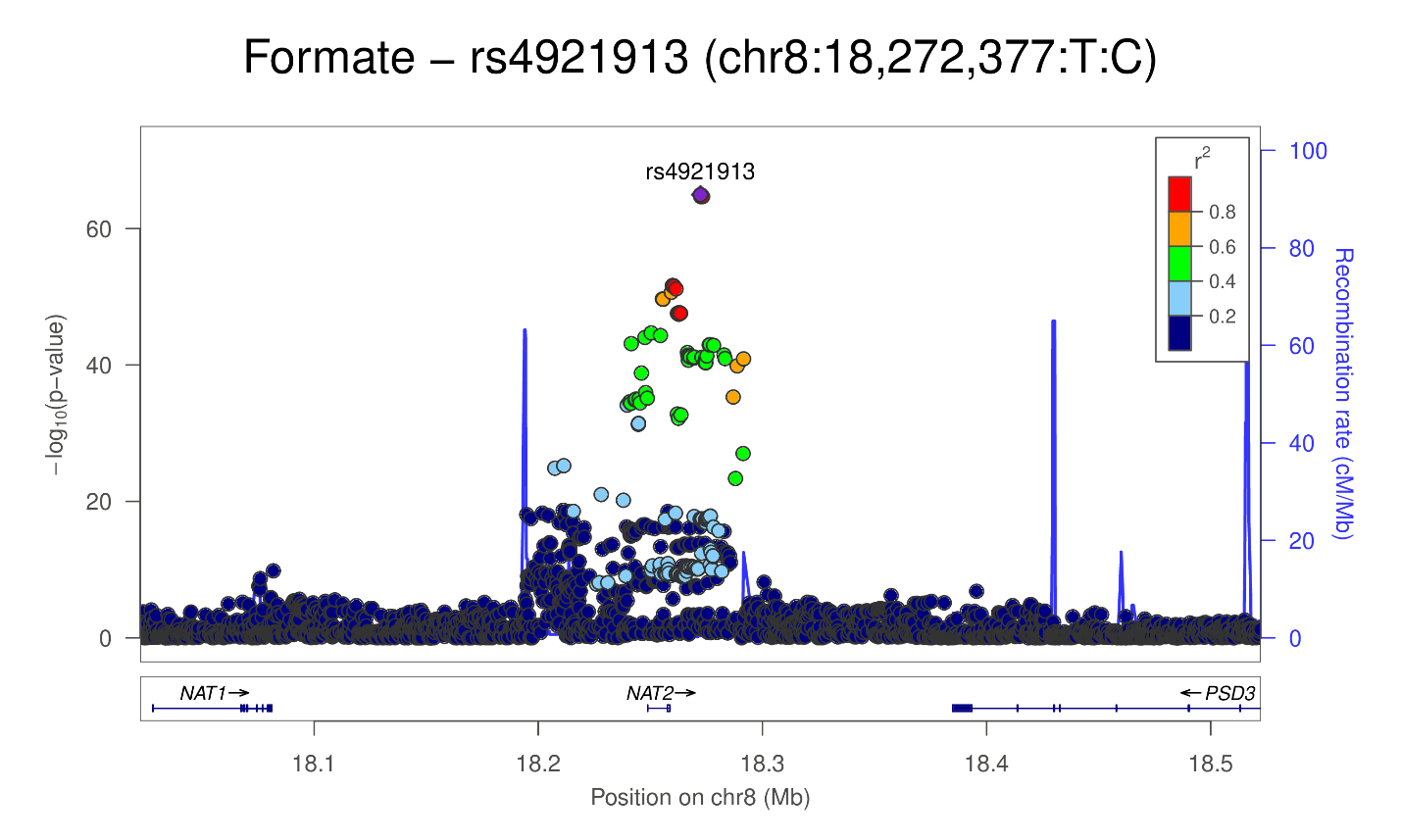

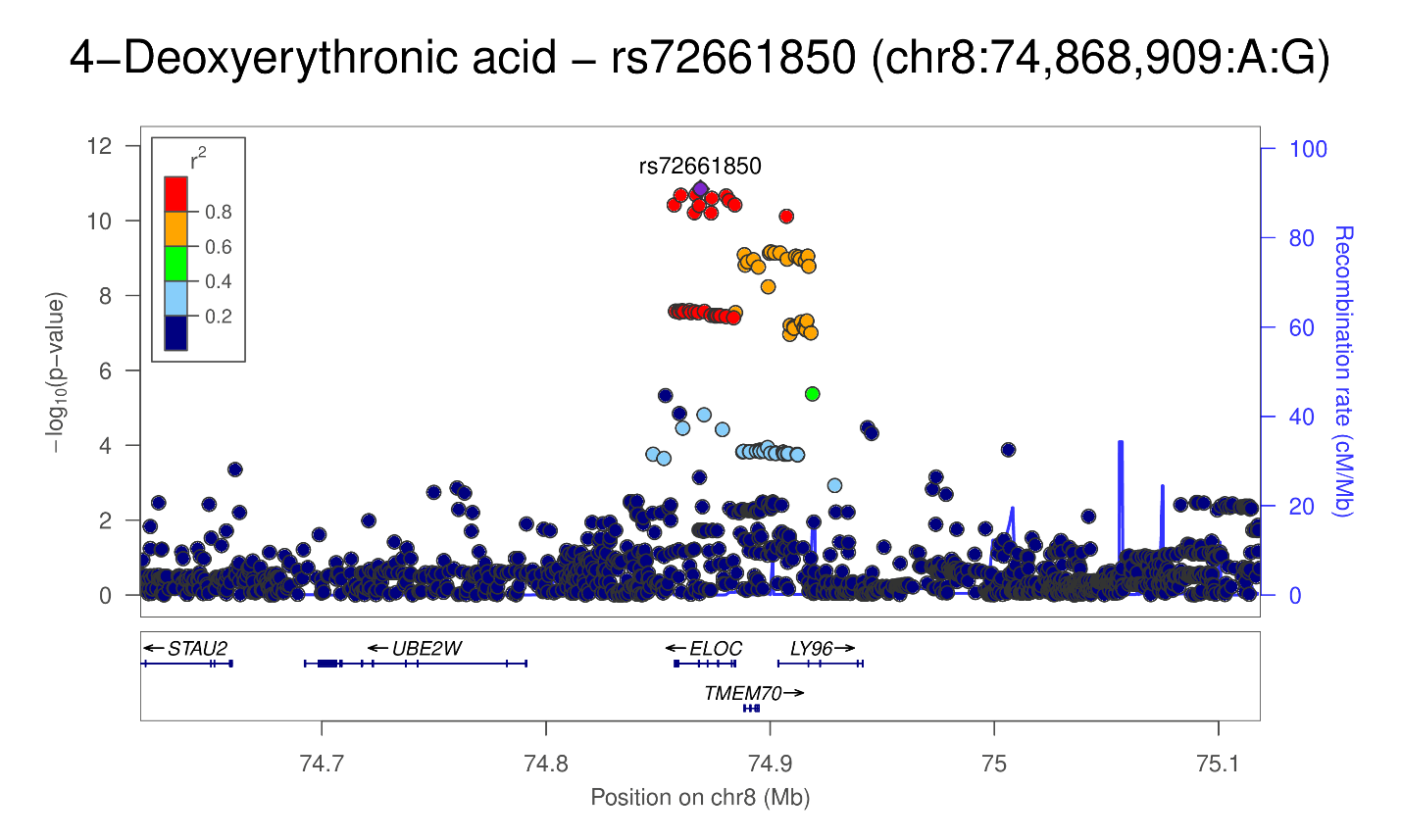

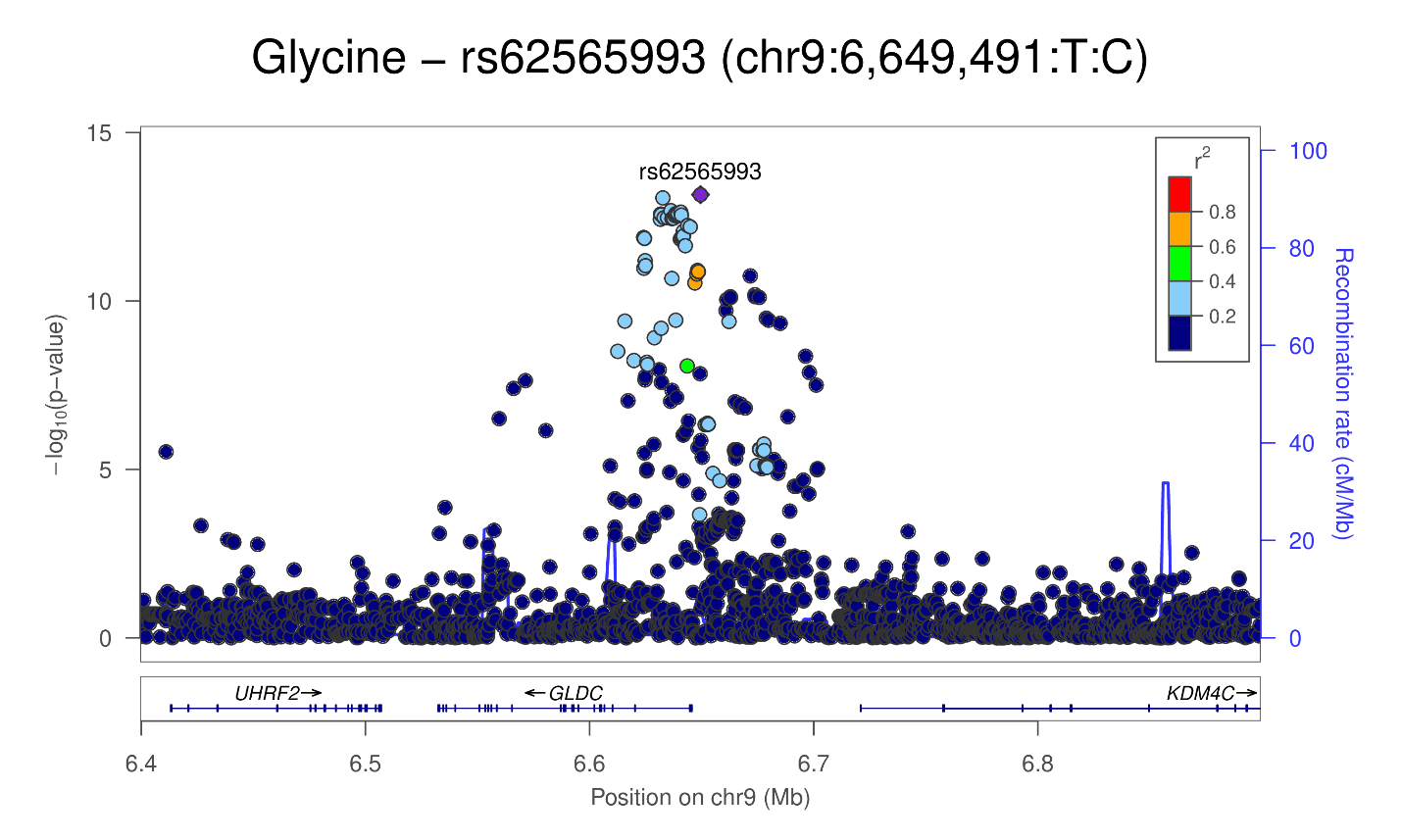

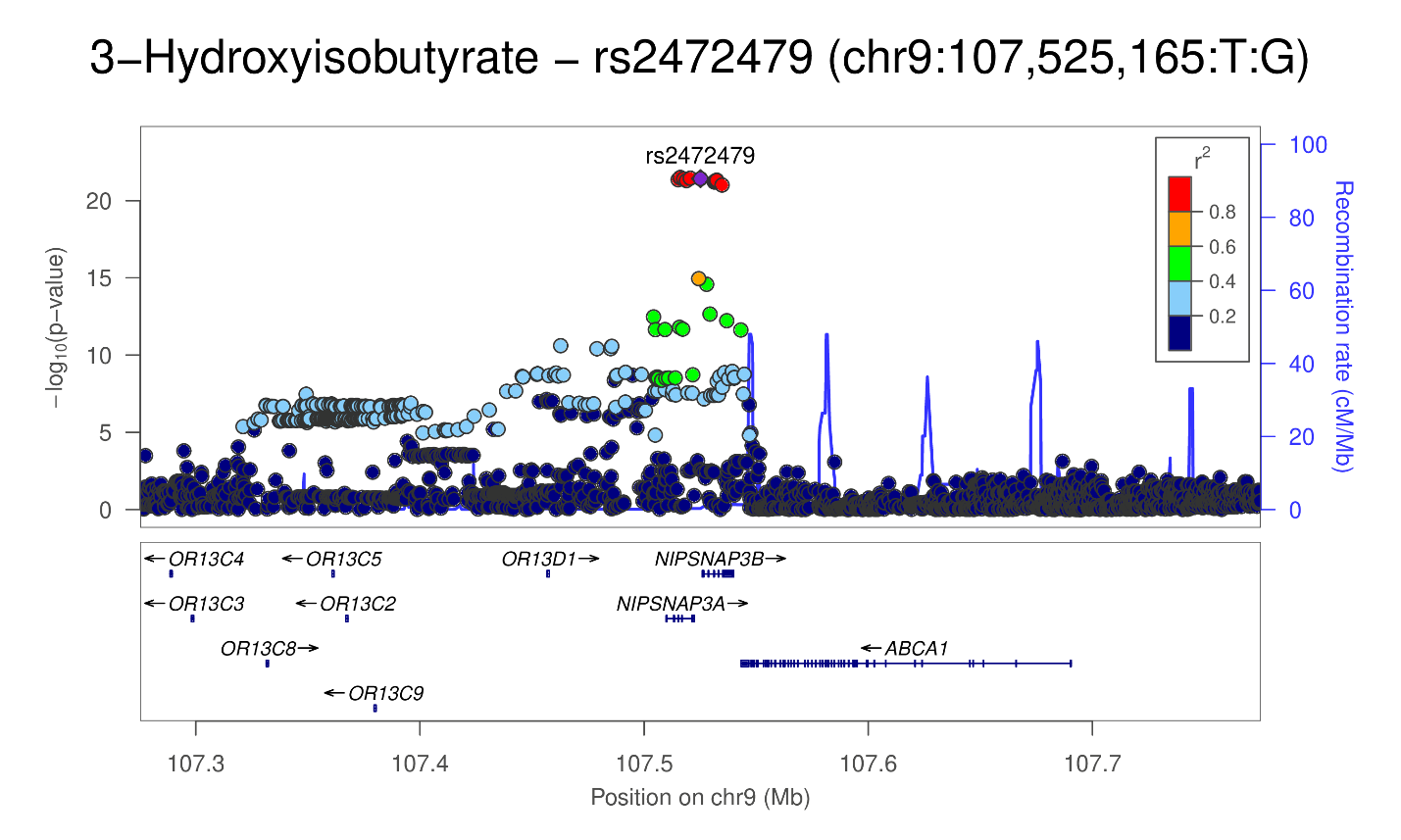

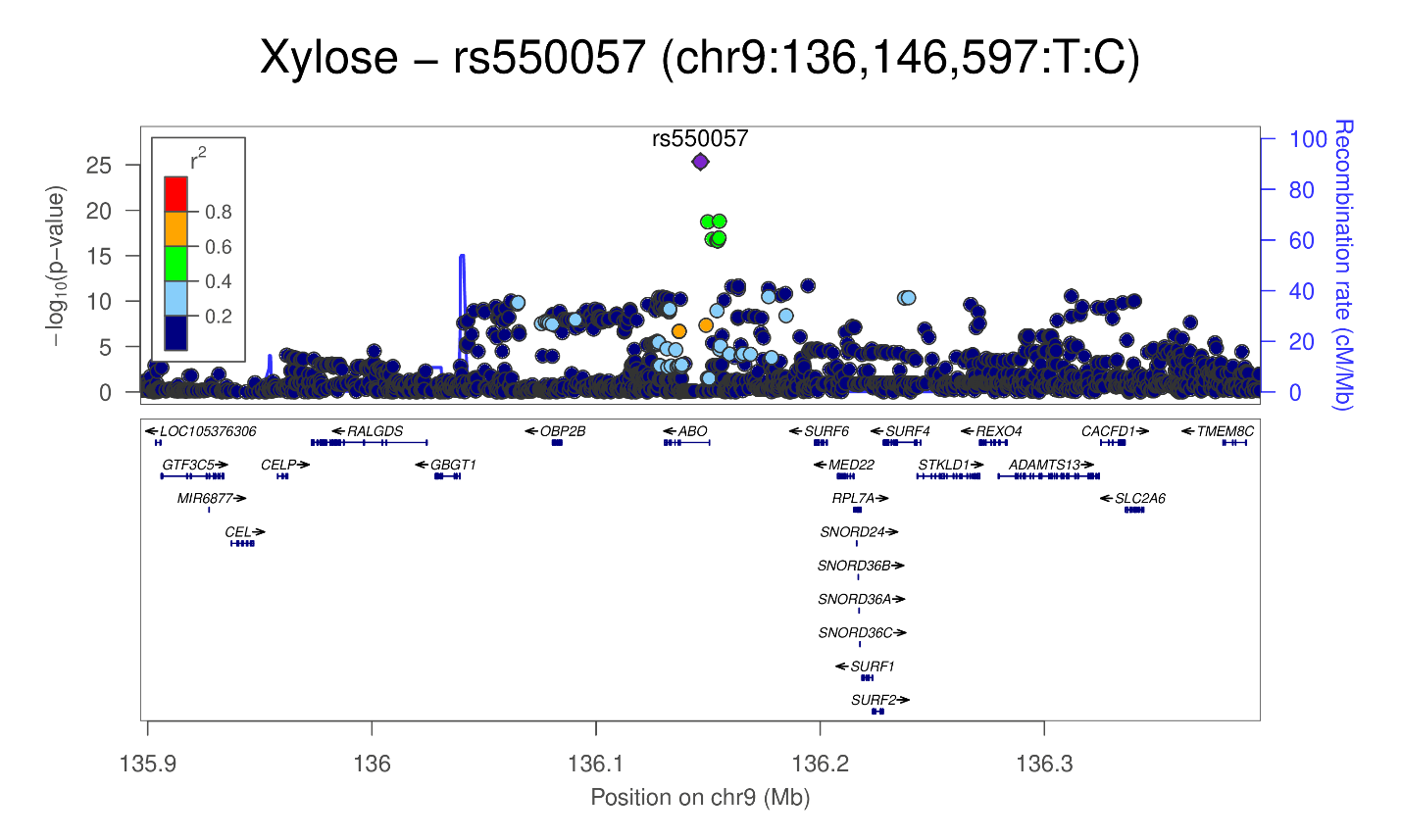

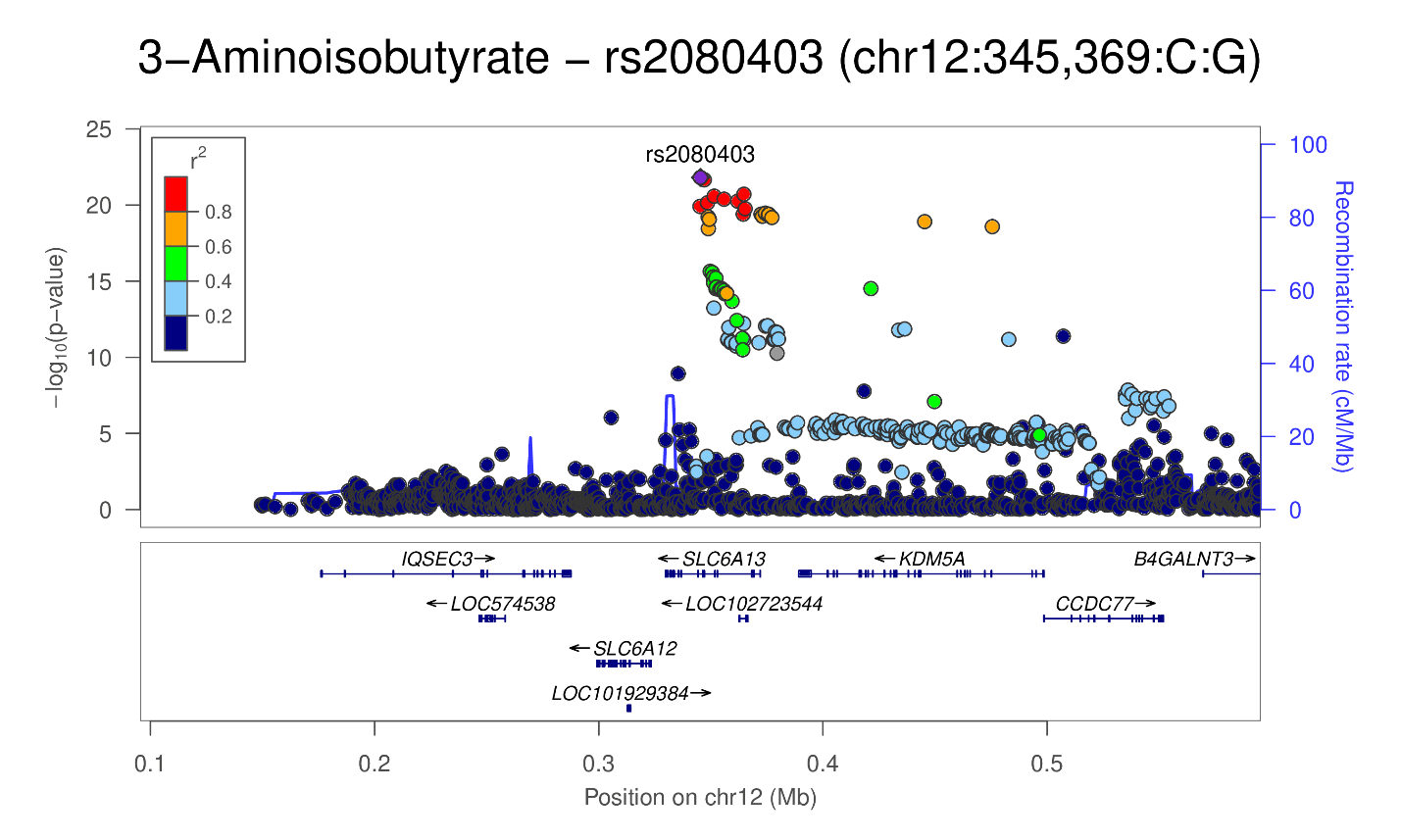

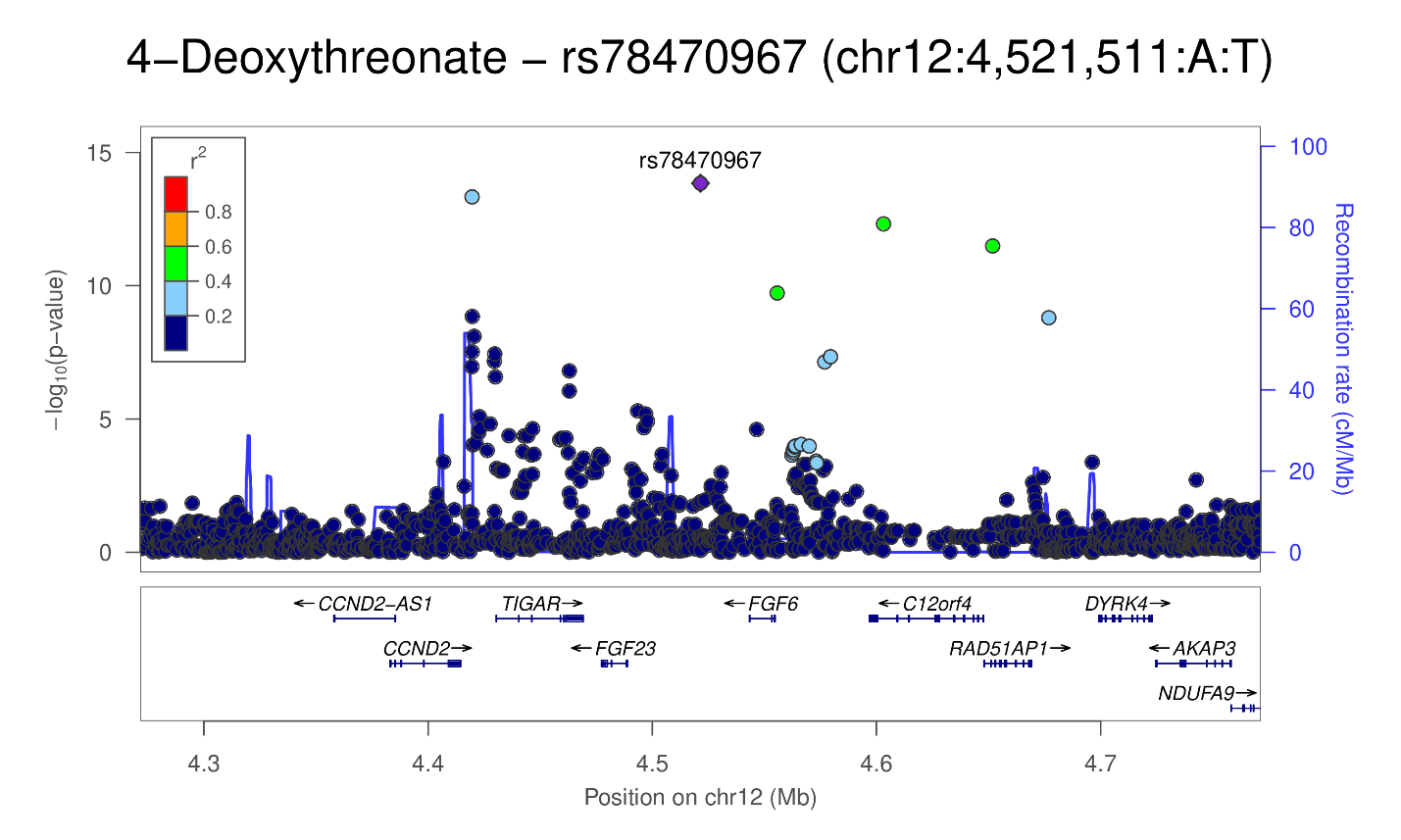

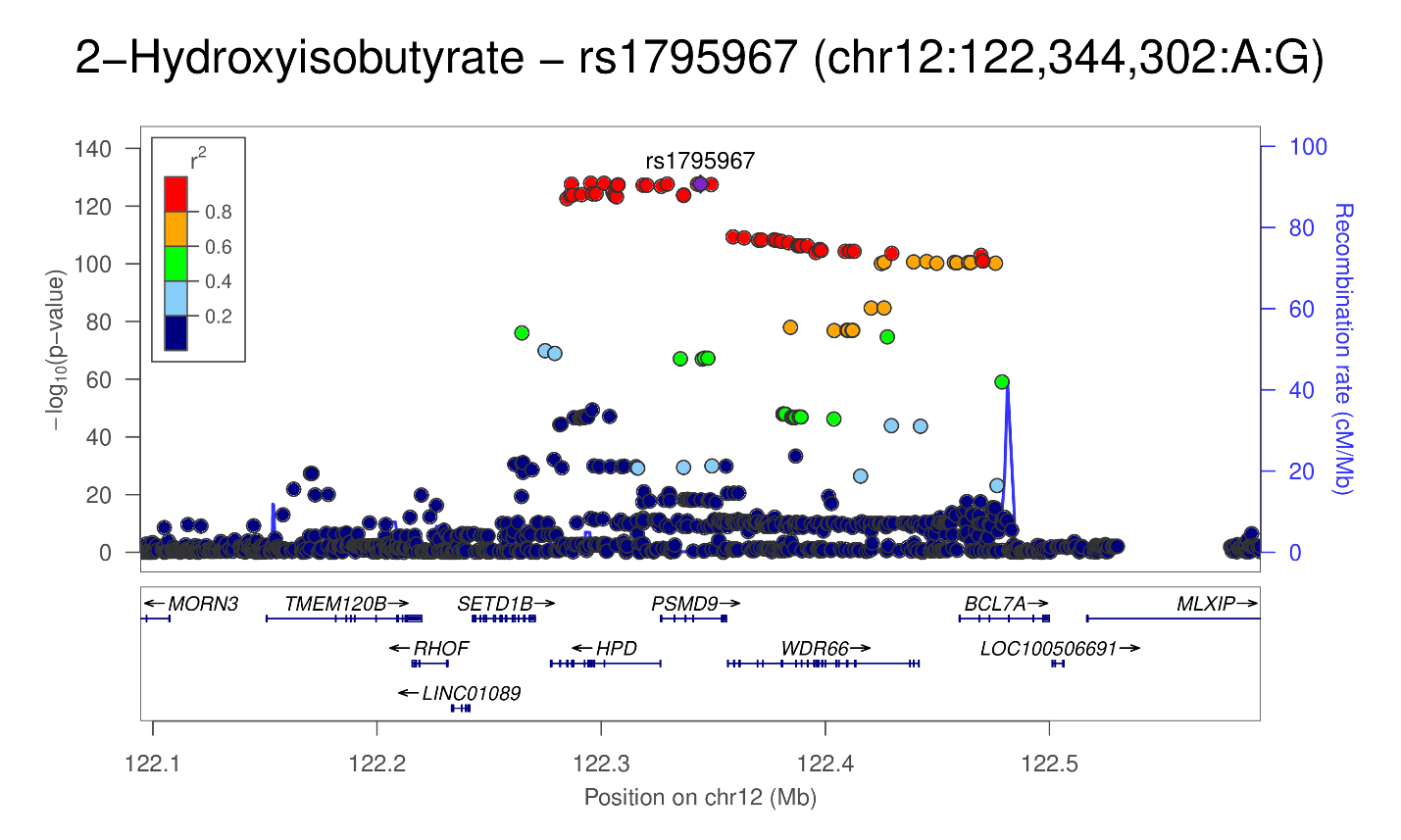

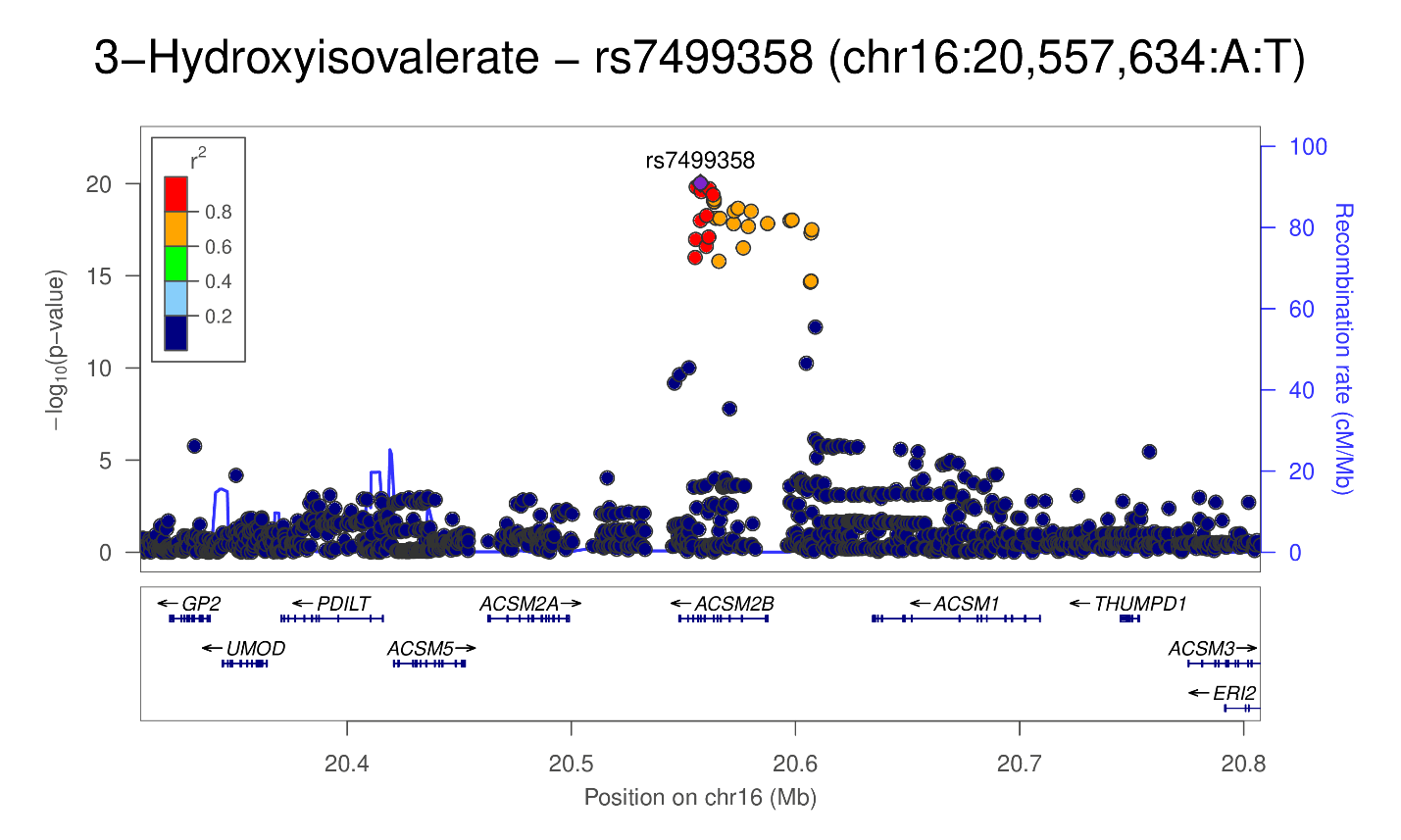

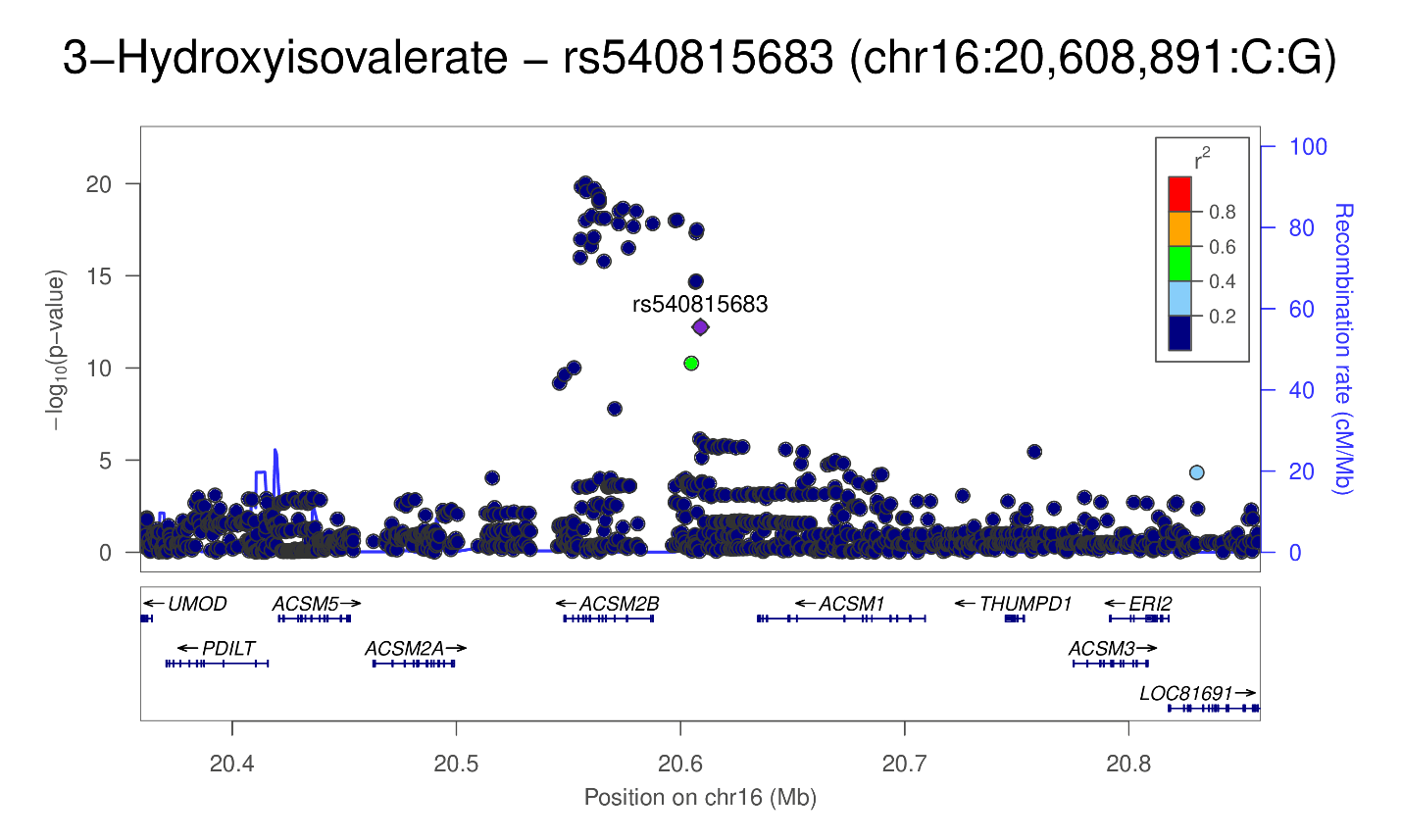

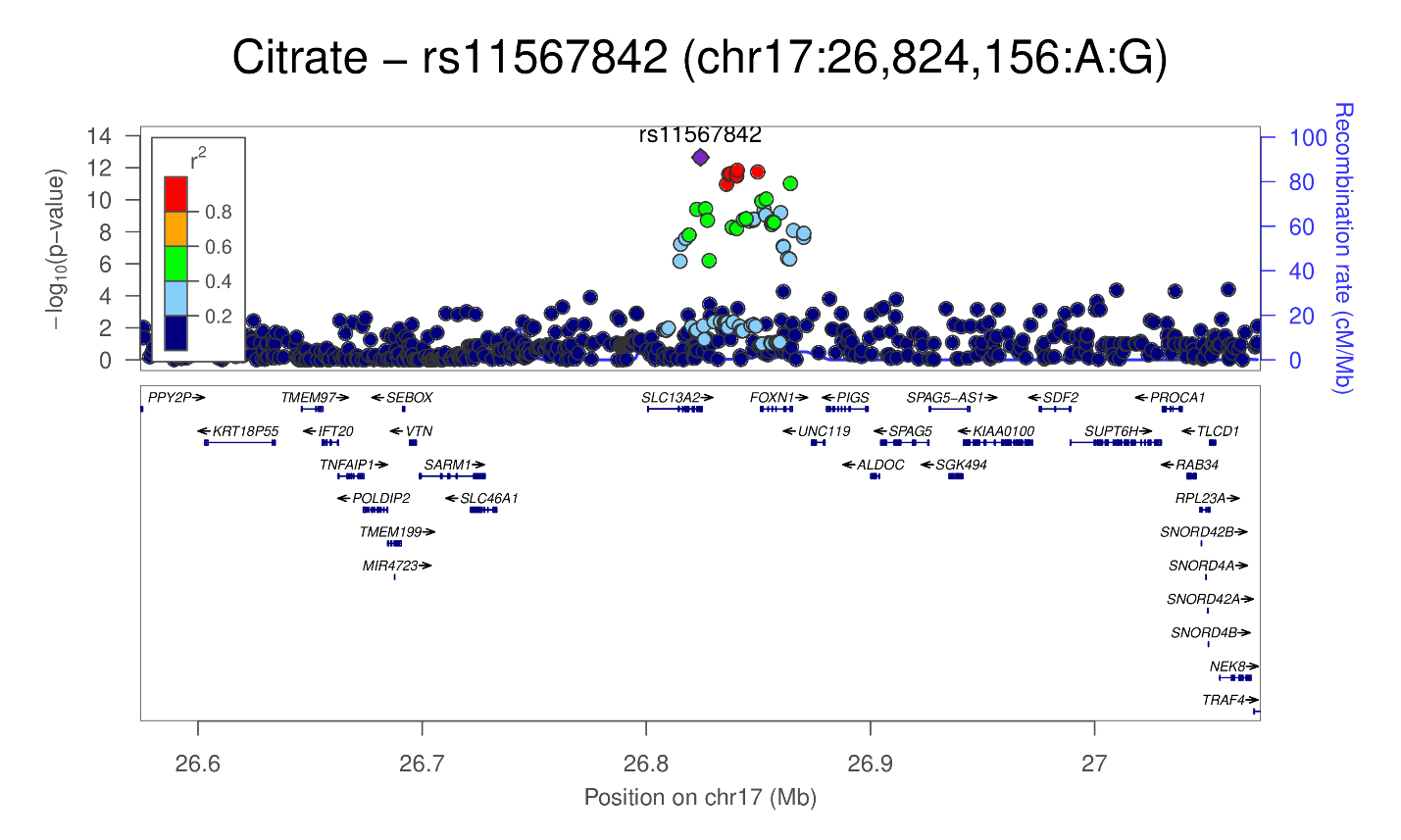

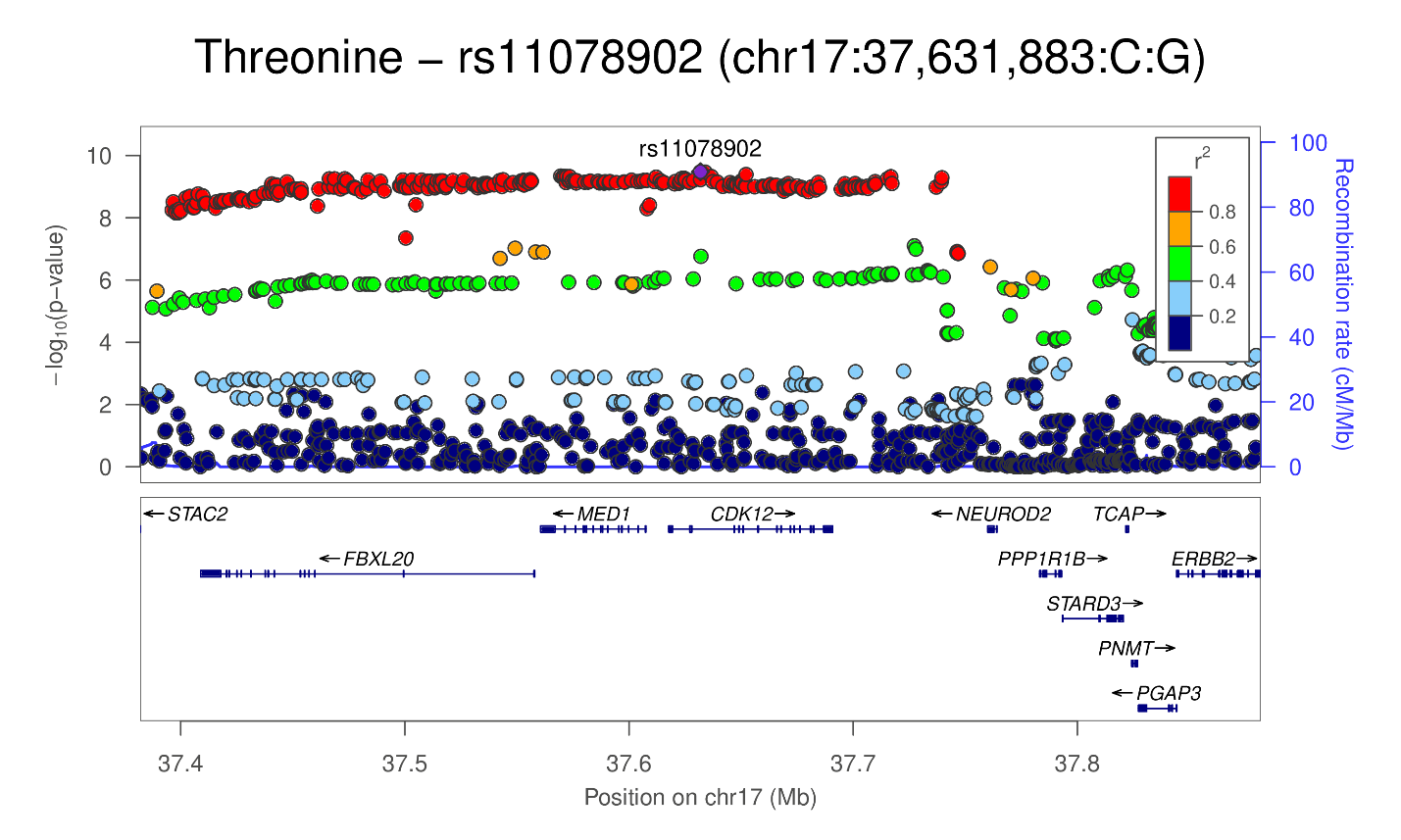

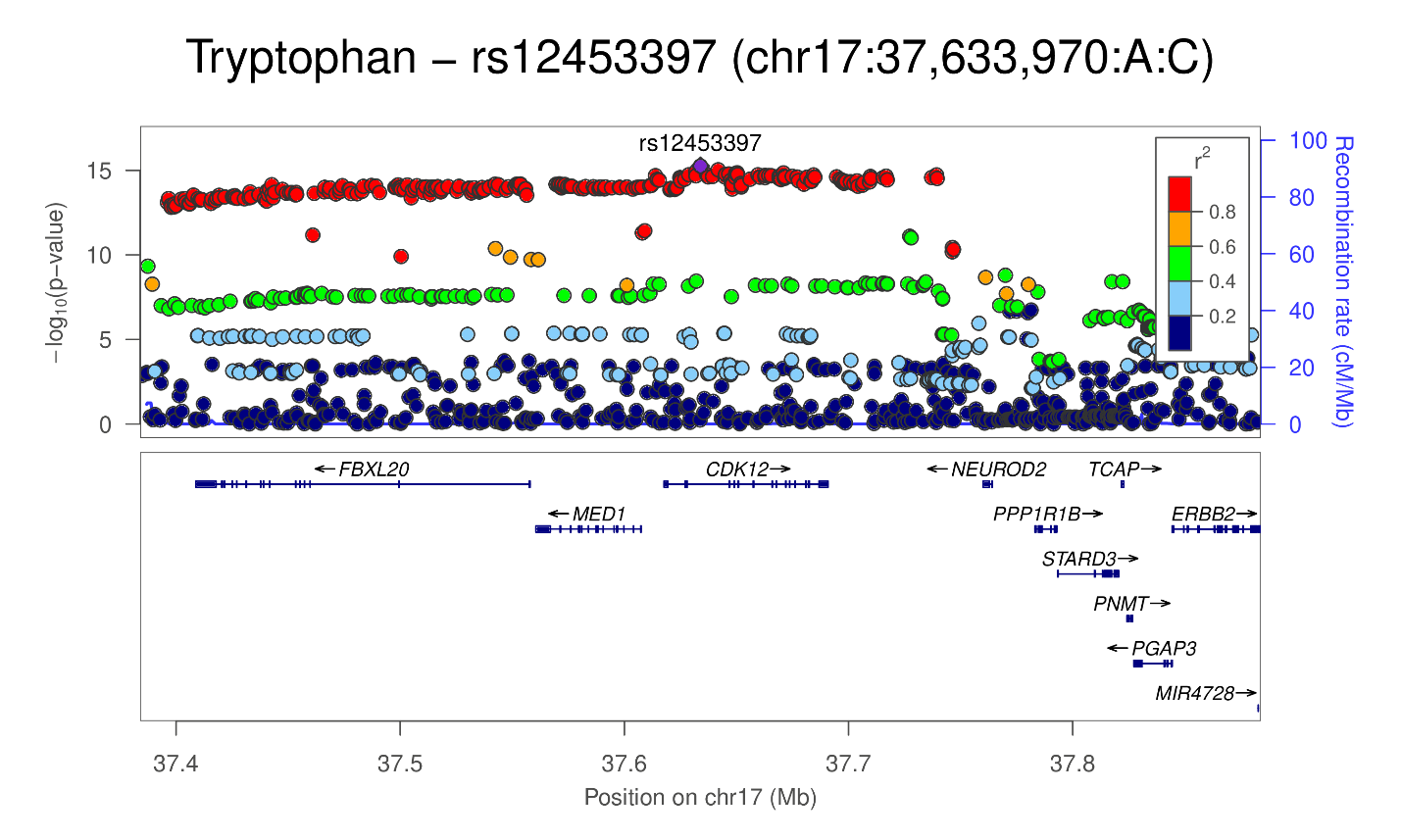

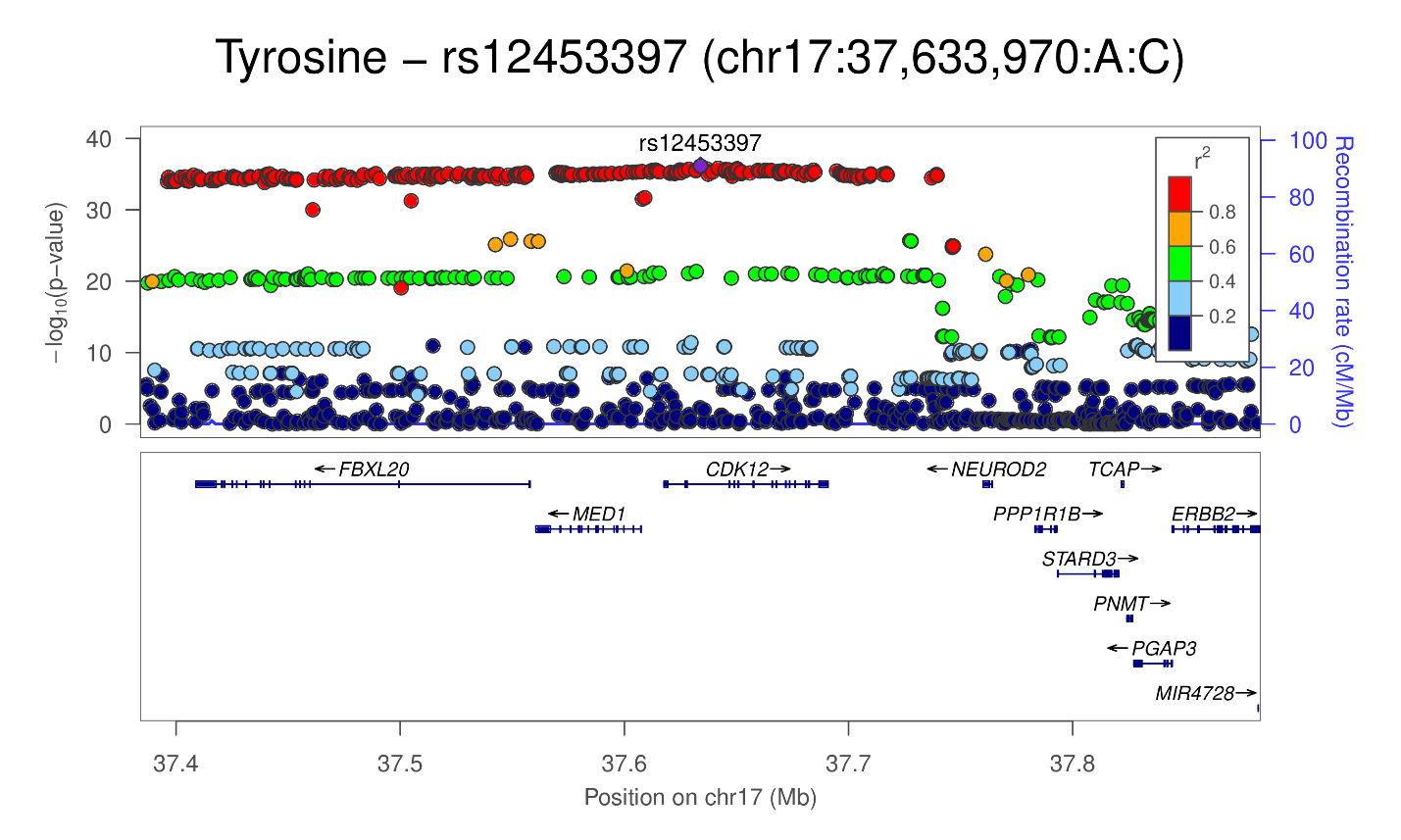

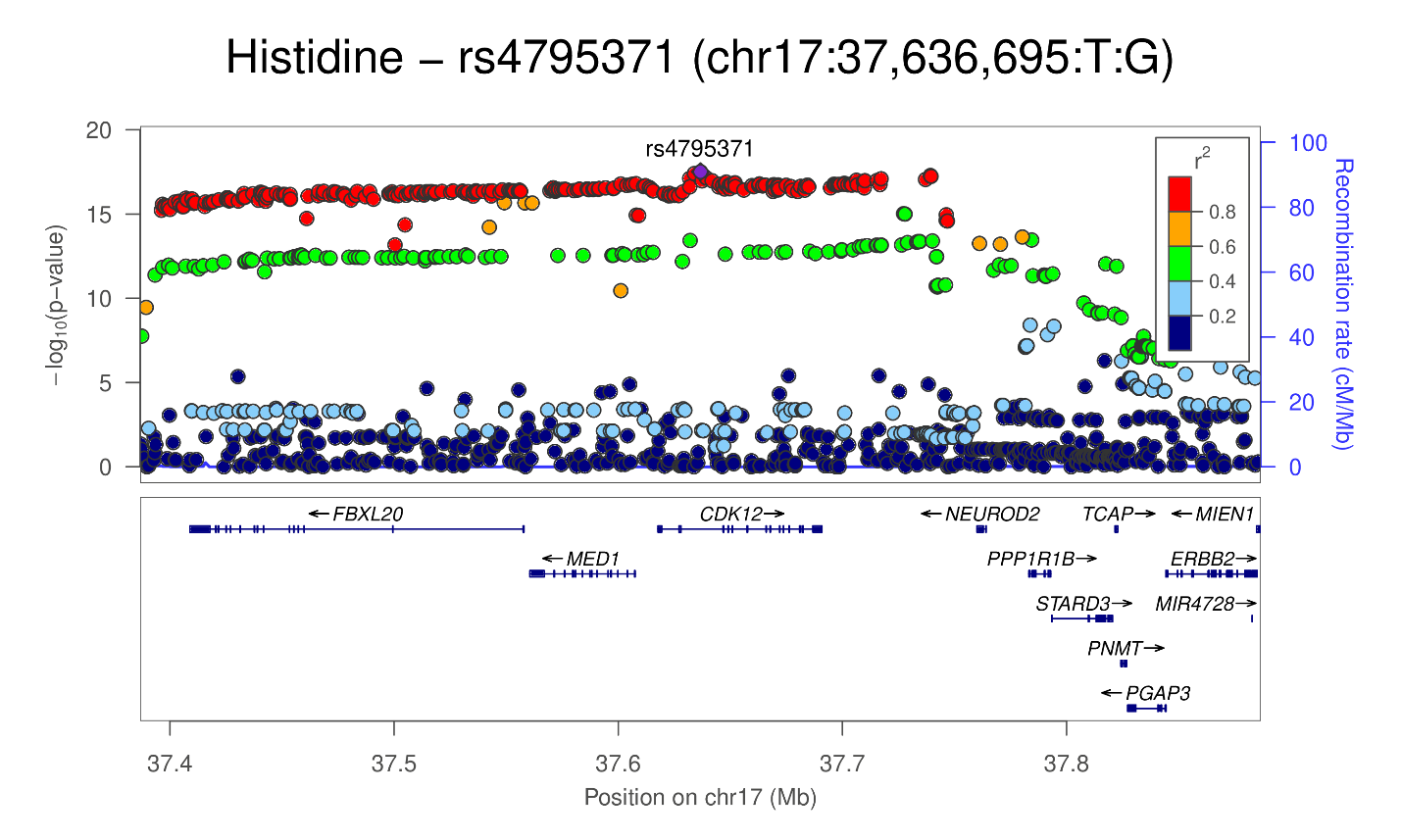

### Supplementary Figure 3

***AGTX2* expression in the kidneys.** In single-cell RNA-Seq of human kidneys, *AGXT2* is expressed in the proximal convoluted tubules in special. *AGXT2* encodes the alanine--glyoxylate aminotransferase 2. Data from Wilson et al.^10^, figure obtained from <http://humphreyslab.com/SingleCell/comparison.php>.

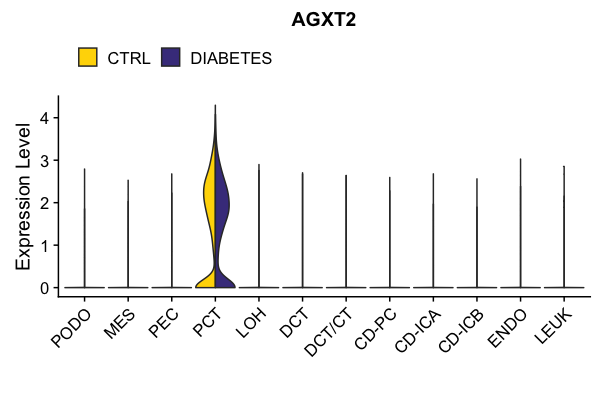

**Supplementary Figure 4**

**Tyrosine GWAS signal vs. *SLC6A18* and *SLC6A19* kidney eQTL signals.** Tyrosine signal p-values plotted against *SLC6A18* and SLC6A19 eQTL p-values and coloured by r^2^ values with either of the two independent tyrosine lead variants in the locus rs11133665 or rs7704882: A) *SLC6A18* rs11133665, B) *SLC6A18* rs7704882, C) *SLC6A19* rs11133665, and D) *SLC6A19* rs7704882.

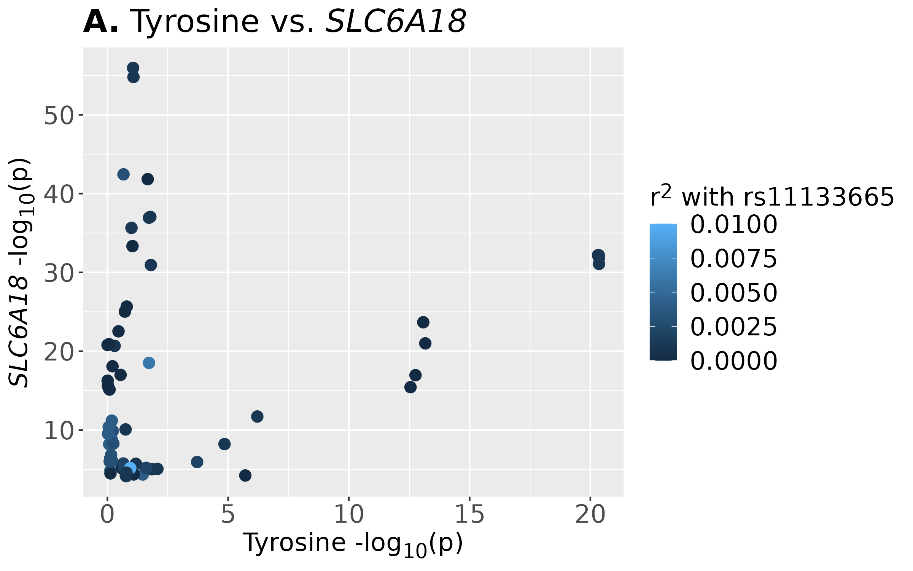

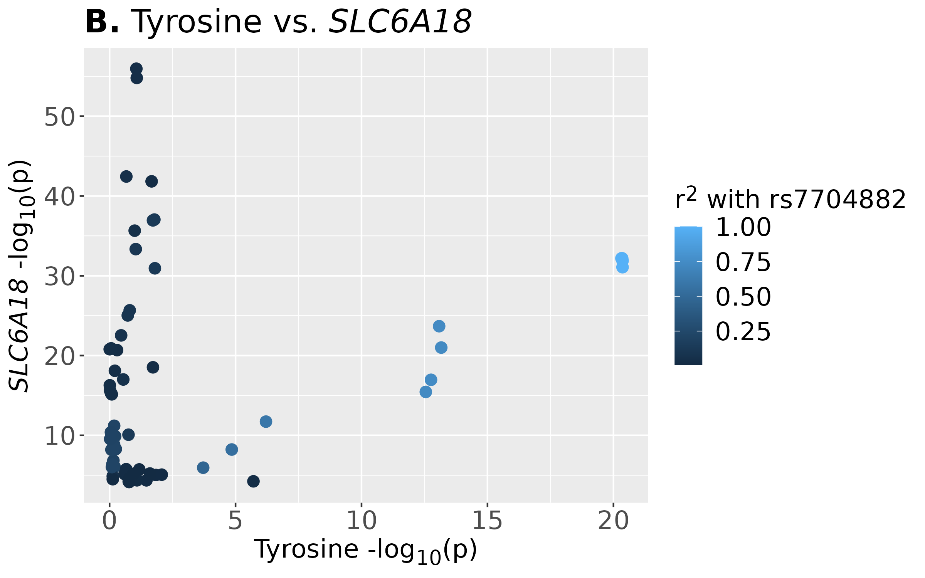

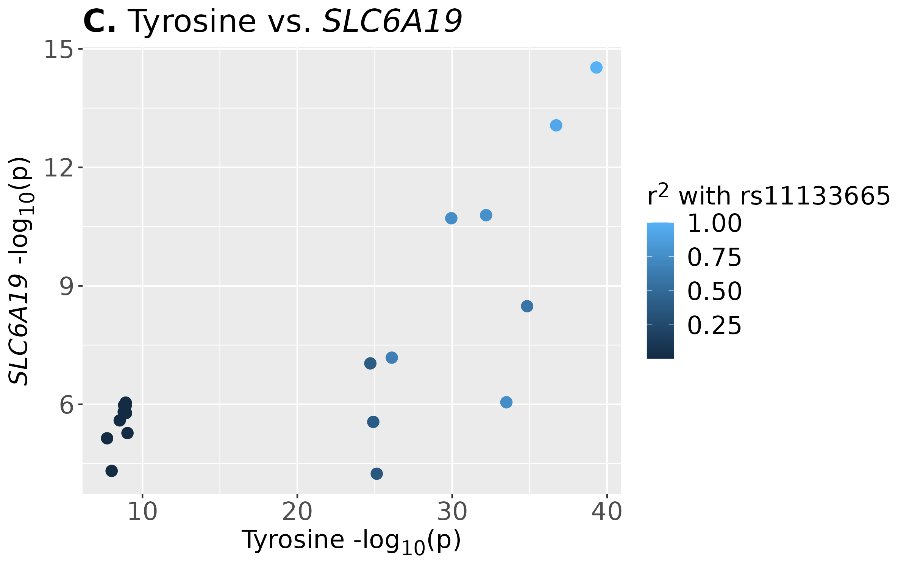

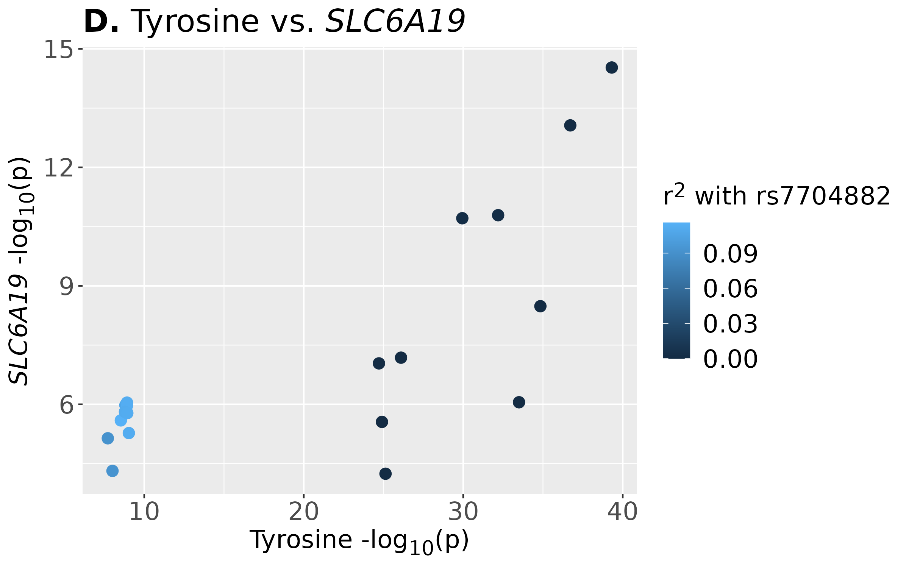

### Supplementary Figure 5

**Regional association with 3-aminoisobutyrate and blood and kidney eQTL target genes**. A. LocusZoom plot of the 3-aminoisobutyrate signal centred around lead variant rs2080403. B. Blood eQTL association for *SLC6A13* and *NINJ2* overlaid on top of the 3-aminoisobutyrate association signal. C. Kidney eQTL associations for *SLC6A13*, *AC007406.2,* and *CCDC77* overlaid on top of the 3-aminoisobutyrate association signal. The R2 values with rs11133665 are calculated based on 1000 Genomes phase 3 European population. Variants with no R2 information are not shown.

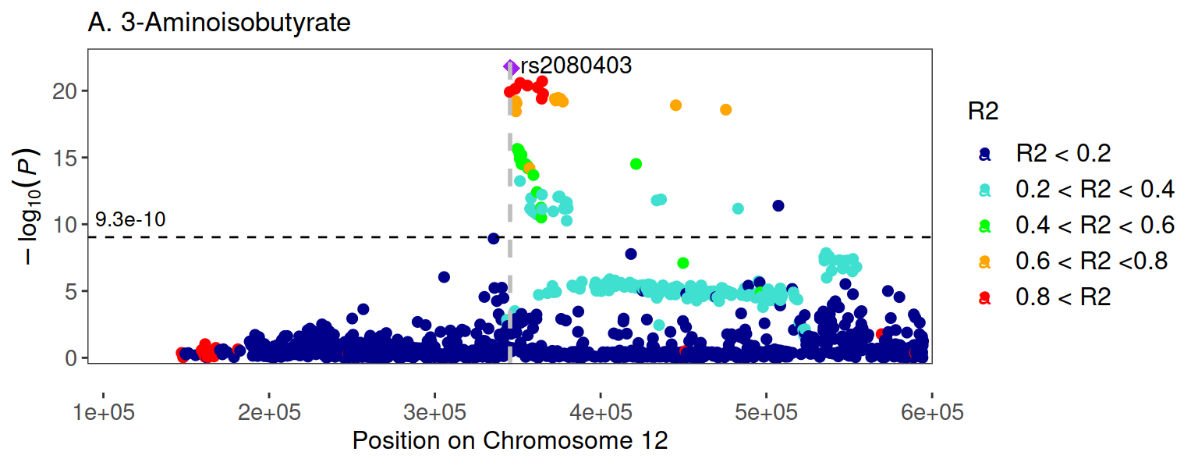

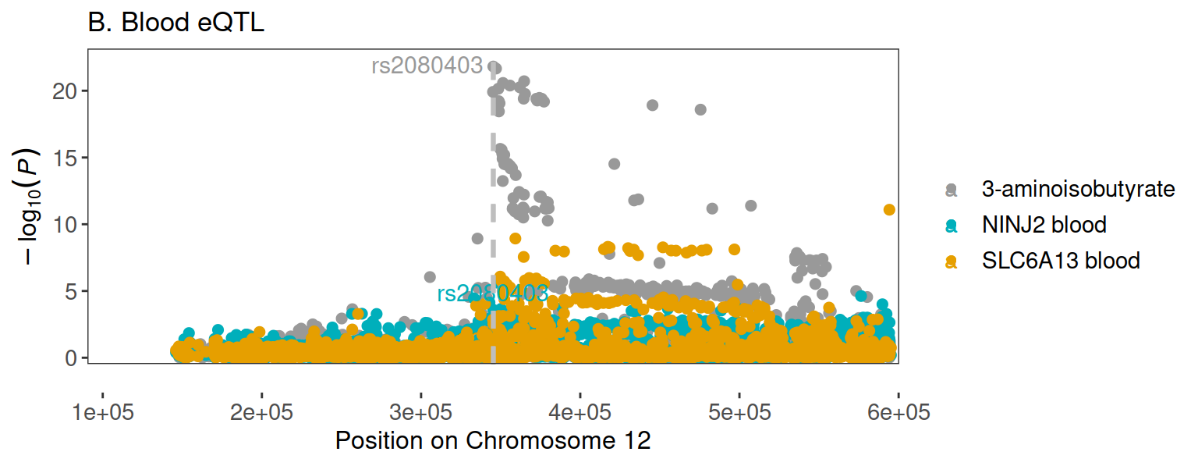

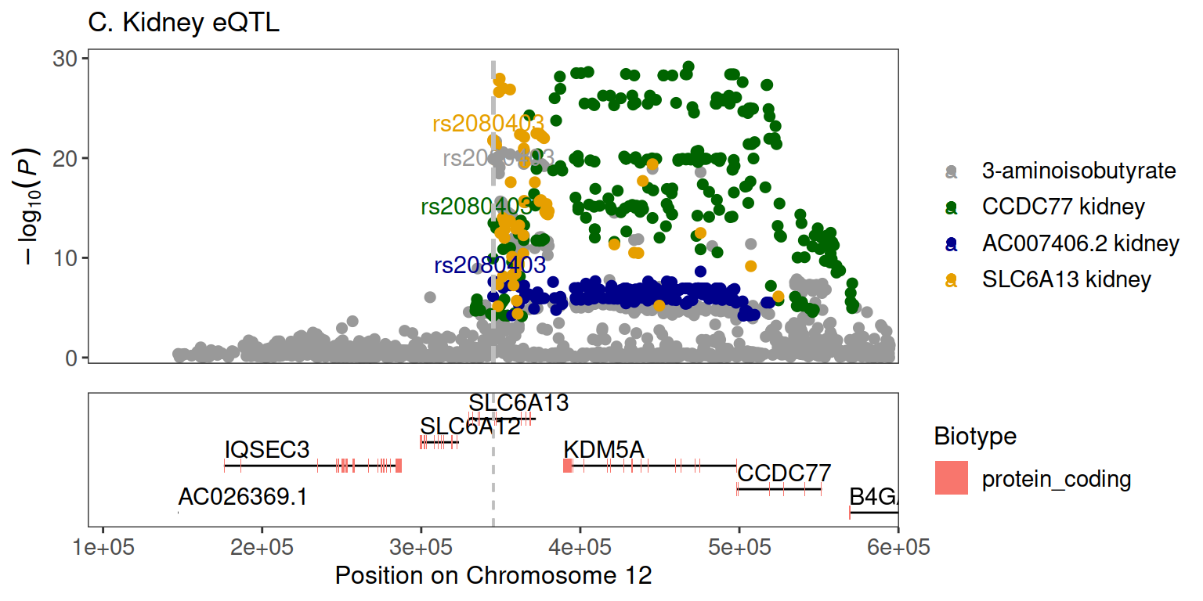

### Supplementary Figure 6

**Two-sample Mendelian Randomization results scatter plots for association of eGFR with urinary metabolites.** Plots are shown for analysis in which the genetic instrument for eGFR was significantly associated (*p*<4.6×10^-4^) with a urinary metabolite. The plots include the eGFR effect estimate on the x-axis, the metabolite effect estimate on the y-axis, and lines showing the causal effect estimates for the different methods applied.

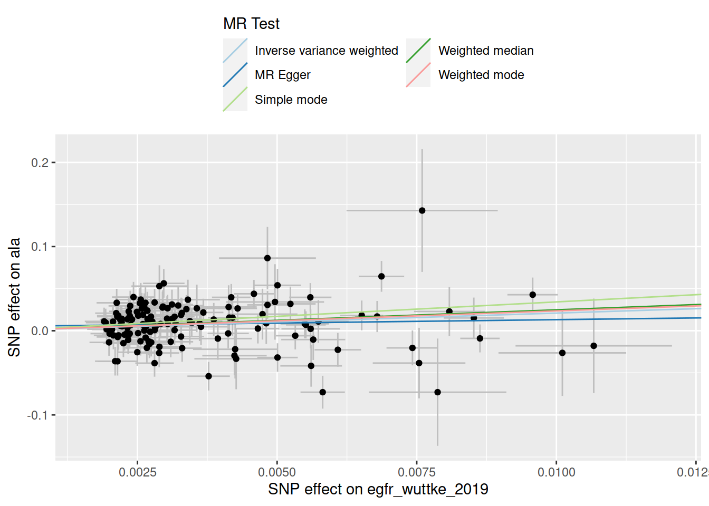

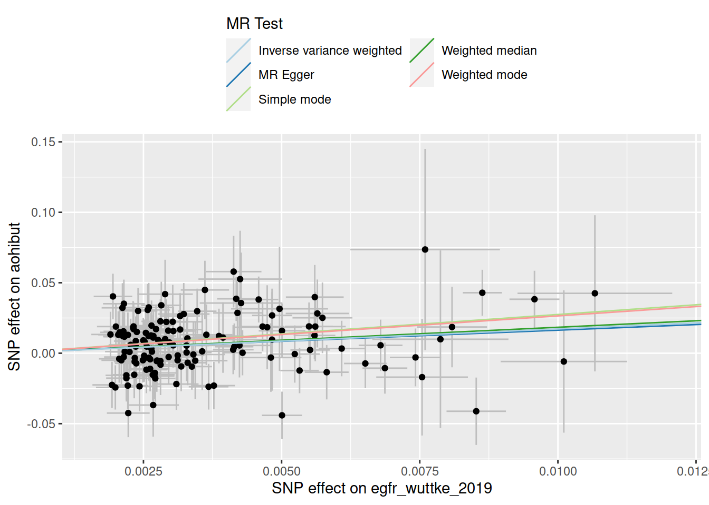

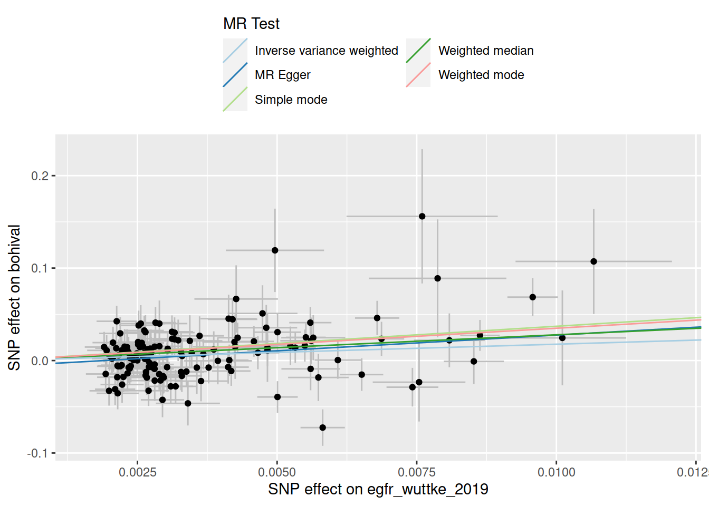

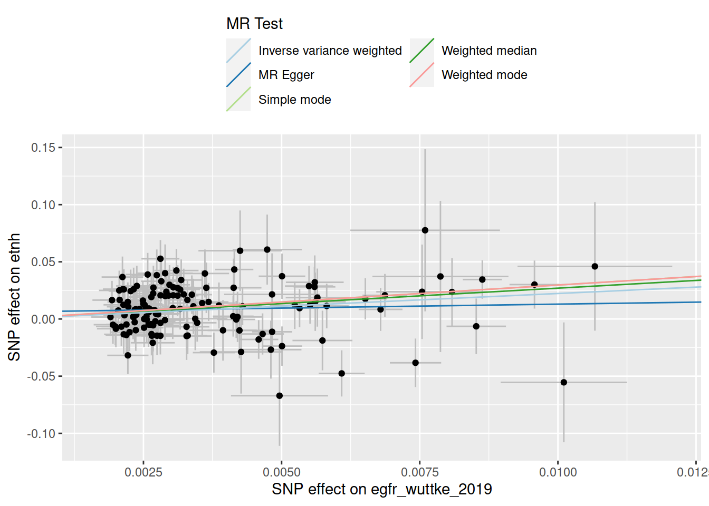

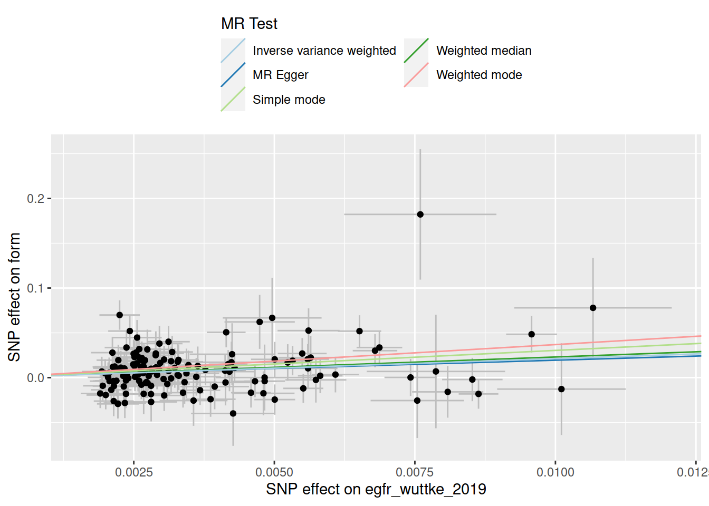

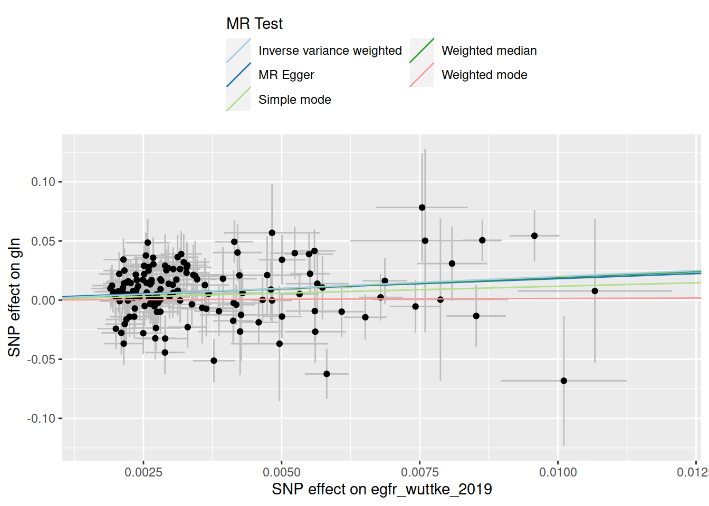

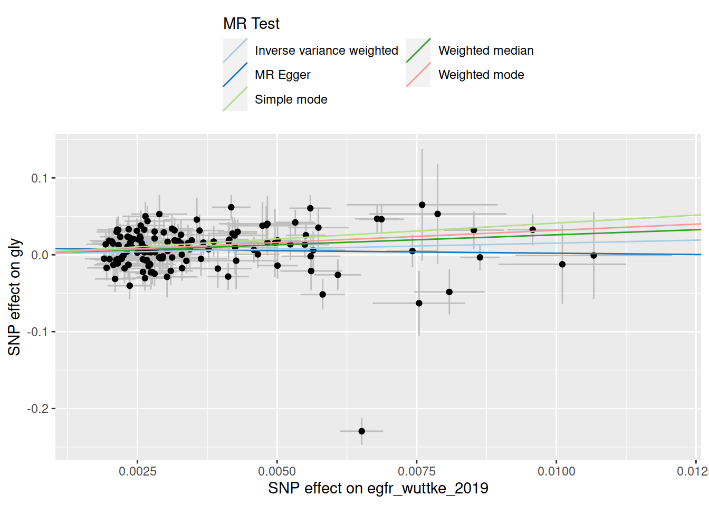

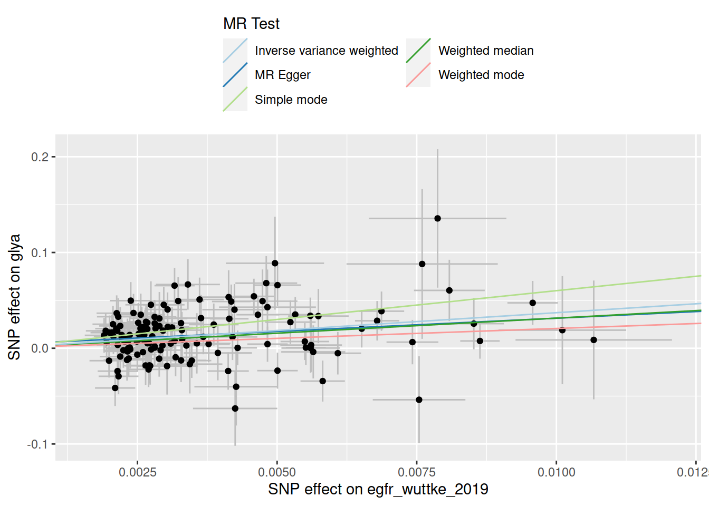

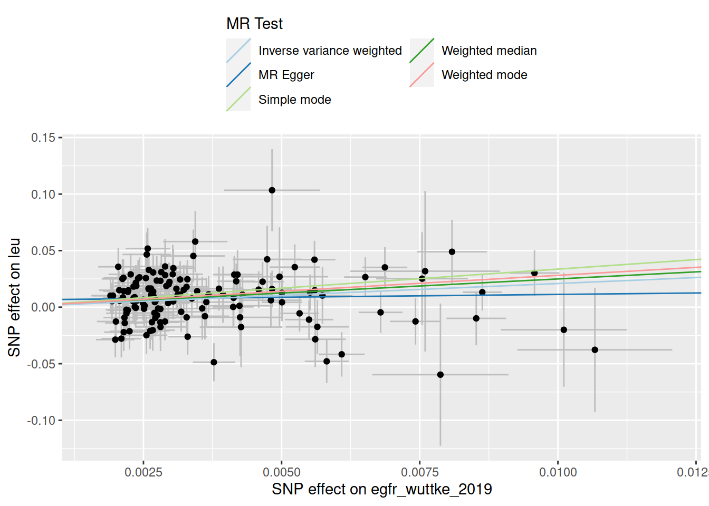

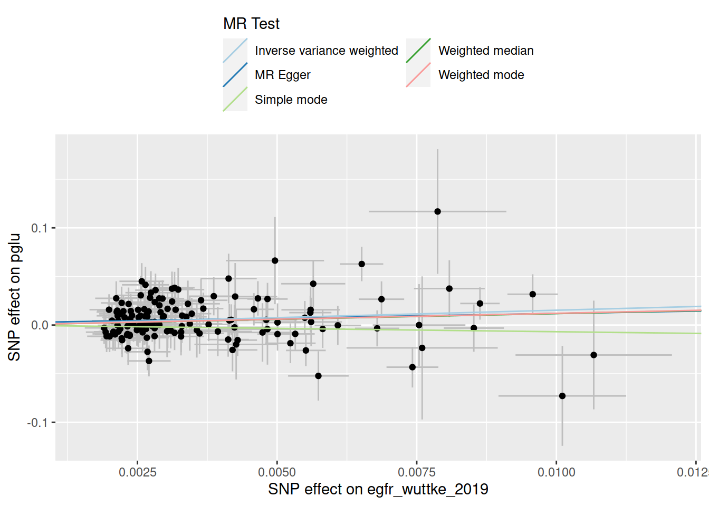

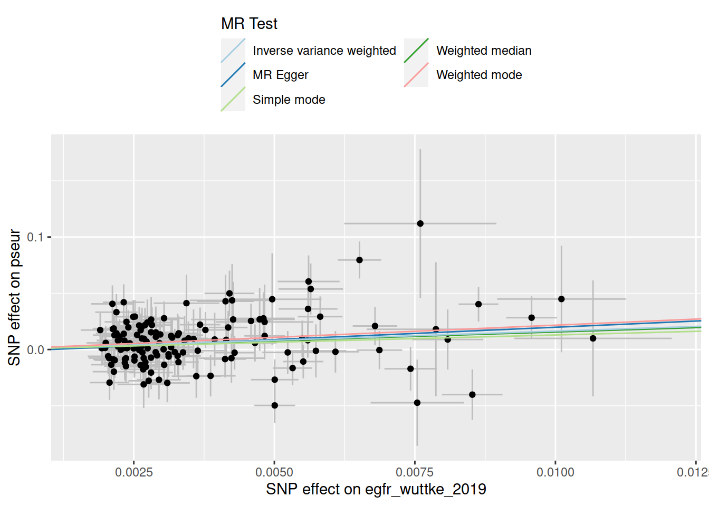

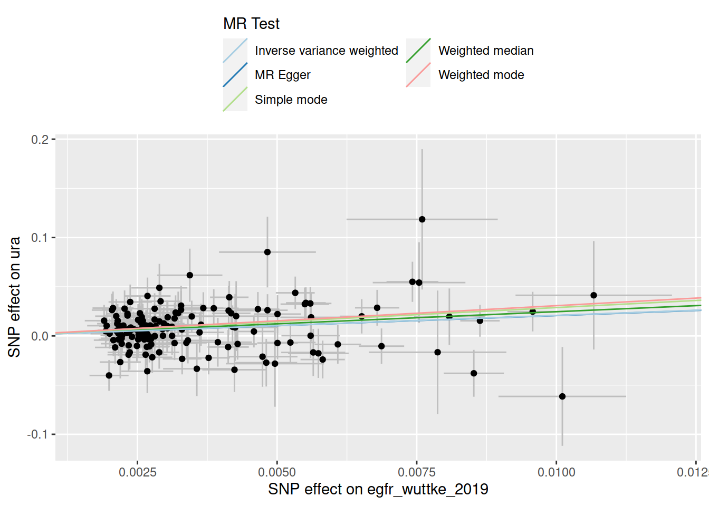

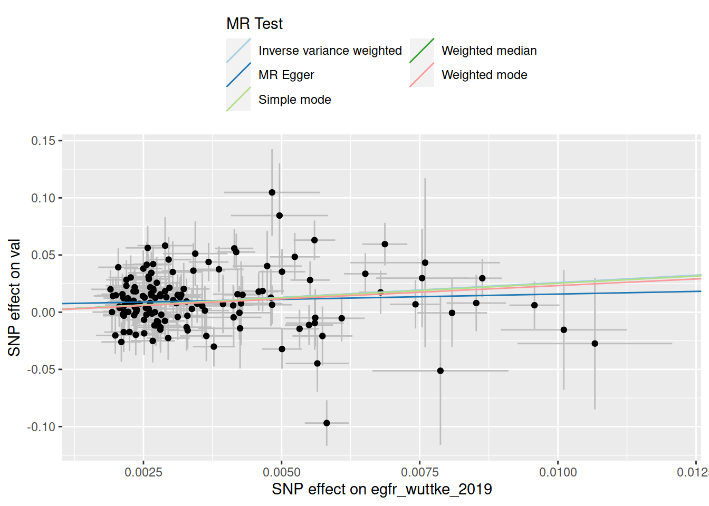

### Supplementary Figure 7

**Effect of eGFR adjustment on the metabolite associations.** Effect estimates in FinnDiane for the COJO lead variants included in the FinnDaine GWAS data set (n=41) not adjusted with eGFR (minimal model: age + sex + genotyping batch + genetic principal components) vs. adjusted for eGFR (minimal model + eGFR).

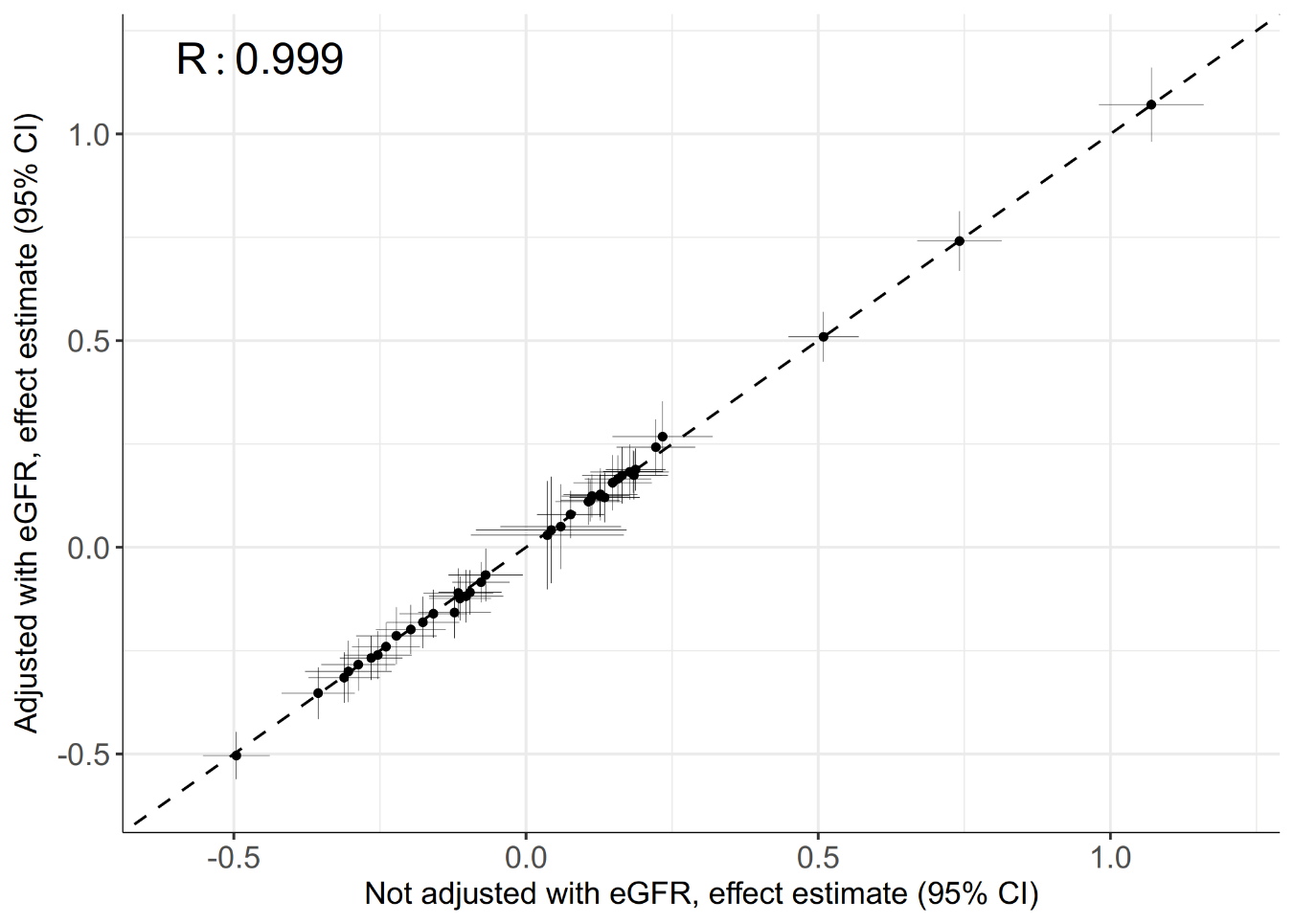
